# Enhancing genotype-phenotype association with optimized machine learning and biological enrichment methods

**DOI:** 10.1101/2024.06.14.24308920

**Authors:** Vaishnavi Jangale, Jyoti Sharma, Rajveer Singh Shekhawat, Pankaj Yadav

## Abstract

Genome-wide association studies (GWAS) are surging again owing to newer high-quality T2T-CHM13 and human pangenome references. Conventional GWAS methods have several limitations, including high false negatives. Non-conventional machine learning-based methods are warranted for analyzing newly sequenced, albeit complex, genomic regions.

We present a robust machine learning-based framework for feature selection and association analysis, incorporating functional enrichment analysis to avoid false negatives. We benchmarked four popular single nucleotide polymorphism (SNP) feature selection methods: least absolute shrinkage and selection operator, ridge regression, elastic-net, and mutual information. Furthermore, we evaluated four association methods: linear regression, random forest, support vector regression (SVR), and XGBoost. We assessed proposed framework on diverse datasets, including subsets of publicly available PennCATH datasets as well as imputed, rare-variants, and simulated datasets. Low-density lipoprotein (LDL) cholesterol level was used as a phenotype for illustration. Our analysis revealed elastic-net combined with SVR consistently outperformed other methods across various datasets. Functional annotation of top 100 SNPs from PennCATH-real dataset revealed their expression in LDL cholesterol-related tissues. Our analysis validated three previously known genes (APOB, TRAPPC9, and EEPD1) implicated in cholesterol-regulated pathways. Also, rare-variant dataset analysis confirmed 37 known genes associated with LDL cholesterol. We identified several important genes, including APOB (familial-hypercholesterolemia), PTK2B (Alzheimer’s disease), and PTPN12 (myocardial ischemia/reperfusion injuries) as potential drug targets for cholesterol-related diseases.

Our comprehensive analyses highlight elastic-net combined with SVR for association analysis could overcome limitations of conventional GWAS approaches. Our framework effectively detects common and rare variants associated with complex traits, enhancing the understanding of complex diseases.

## I. INTRODUCTION

Genome-wide association studies (GWAS) have transformed the field of genetics by enabling researchers to identify genetic variants associated with complex traits and diseases on a genome-wide scale. After the completion of the Human Genome Project (HGP) [1], GWAS has been recognised as the most effective approach for identifying variants associated with phenotypes of interest. The popularity of GWAS is expected to surge in the near future owing to the availability of newer high-quality gap-less reference genomes like T2T-CHM13 and human pangenome [2, 3]. Over the past two decades, GWAS has identified genetic risk loci such as FTO2 for obesity [4] and PTPN22 for autoimmune diseases [5]. Additionally, GWAS also uncovered pathways such as the IL-12/IL-23 pathway linked to Crohn’s disease [6], which encouraged clinical studies for medicines targeting the pathways.

Despite the widespread use and success of GWAS in identifying numerous disease-associated variants, conventional GWAS methods suffer from major limitations. The stringent p-value criteria used in GWAS may overlook variants with low or moderate effect sizes, potentially leading to false negatives [7] and hindering the detection of variants with modest yet biologically significant associations [8, 9]. Furthermore, conventional linear and logistic regression models used in GWAS [10] often fail to consider epistatic interactions between genetic variants [8]. Consequently, the estimated heritability derived from such GWAS analysis may not accurately reflect the true genetic component underlying complex traits/diseases, giving rise to “missing heritability” [11]. Missing heritability poses a significant challenge in GWAS analysis, underscoring the need for alternative approaches to enhance the detection of genetic associations. Moreover, the reliability of conventional GWAS results is often questioned due to the lack of functional annotations, making it challenging to interpret the biological significance of identified variants. Without this contextual information, it becomes challenging to discern whether a detected association is causative or merely a marker in linkage disequilibrium (LD) with the true functional variant.

To address the limitations of GWAS, researchers have employed machine learning (ML) methods, such as decision tree-based and penalized regression-based approaches, for association analyses. Extreme Gradient Boosting (XGBoost), based on gradient-boosted decision trees, effectively incorporates pairwise epistatic interactions of single nucleotide polymorphisms (SNPs) within one tree. A notable feature of XGBoost is its capacity to restrict interactions between SNPs within single trees, facilitating the study of SNP interactions and enabling prediction models to include complex non-linear interactions in a non-additive form [12]. In contrast to decision tree approaches, penalized regression-based methods, such as ridge regression and least absolute shrinkage and selection operator (LASSO), are comparatively less complex ML methods. These methods simultaneously select variants and estimate their effects on phenotype by imposing constraints on model coefficients [13]. Ridge regularization stabilizes parameter estimation by shrinking predictors, while LASSO regularization facilitates variant selection by driving many regression coefficients to zero [14]. Elastic-net regularization, another ML approach, combines ridge and LASSO penalties to provide shrinkage and automated variant selection. These ML methods are particularly beneficial for GWAS analysis, where epistatic interactions and multicollinearity among nearby SNPs are prevalent due to LD.

However, ML methods encounter challenges due to the high dimensionality of GWAS data, characterized by a large number of SNPs relative to sample size, also known as the “curse of dimensionality” [15]. To address this issue, pre-selection of SNPs using various feature selection methods has been explored [16], albeit with limited success, particularly for complex continuous traits like low-density lipoprotein (LDL) cholesterol levels. To overcome these challenges and enhance our understanding of complex traits and diseases, innovative approaches integrating advanced statistical methodologies, association methods, functional annotations, and biological insights are essential.

Here, we employ elastic-net regularization as a feature selection approach, leveraging its ability to address computational challenges associated with high-dimensional data and mitigate spurious associations [17]. Other widely used feature selection methods such as LASSO, ridge, and mutual information are also implemented to compare the performance of the elastic-net method. Additionally, we utilize linear regression (LR), random forest (RF), XGBoost, and support vector regression (SVR) ML methods for association testing on the selected features/SNPs. Kernel-based ML methods like SVR for association testing can identify interactions by exploring all possible combinations of SNPs within a GWAS dataset. The effectiveness of our approach is validated on real (common and rare variant), imputed and simulated datasets. We perform functional enrichment analysis of identified associated SNPs across diverse biological processes to confirm their biological relevance, ensuring the validity of our findings. The step-wise illustration of the proposed framework is displayed in Figure 1.

**FIG. 1.**
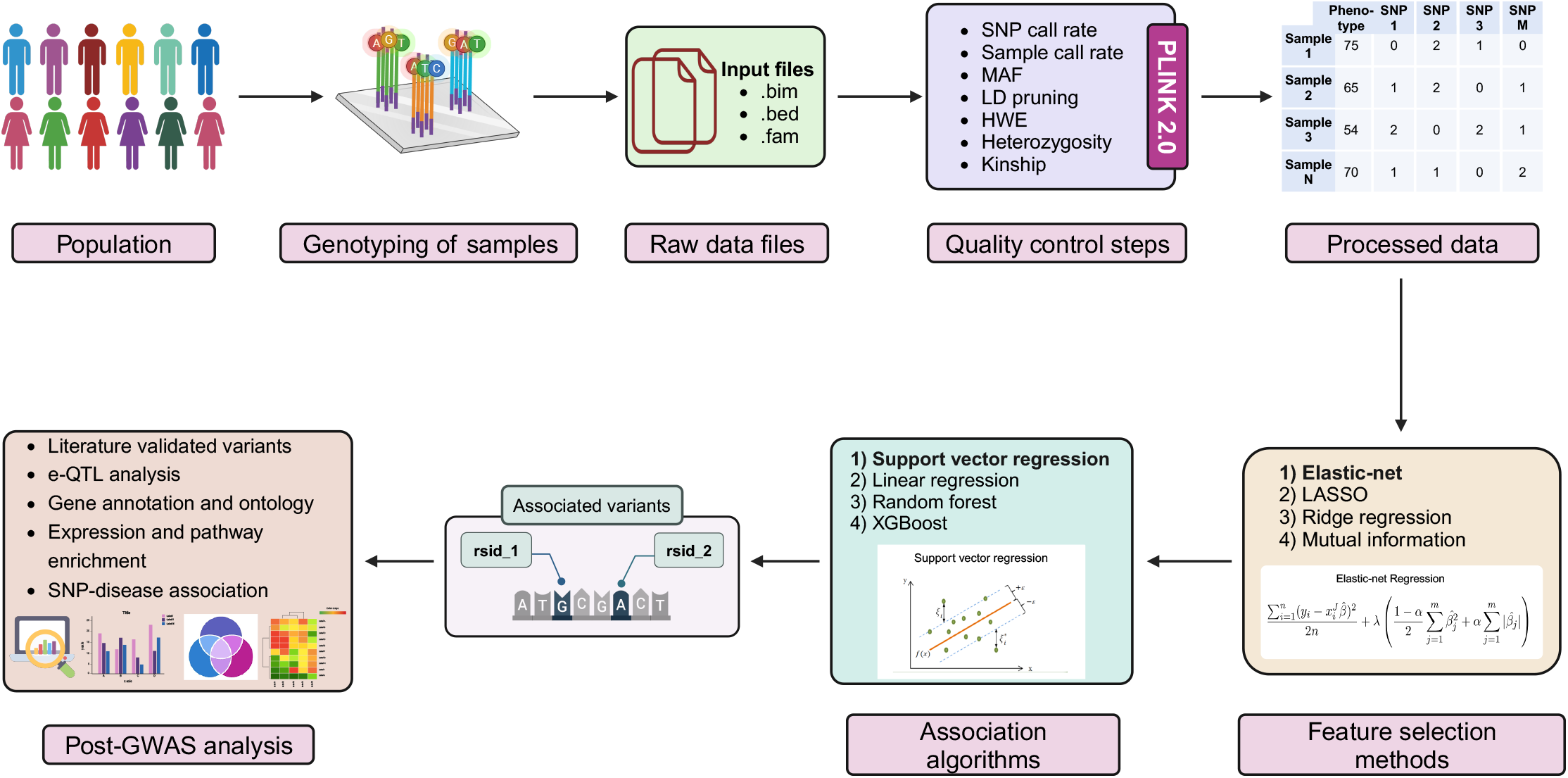
The stepwise workflow of the proposed framework, highlighting the optimal combination of ML methods (shown in **bold**) for the identification of trait-associated variants, followed by validation through post-GWAS analysis Abbreviations- GWAS:genome-wide association studies; SNP:single nucleotide polymorphism; MAF:minor allele frequency; LD:linkage disequilibrium; HWE:Hardy-Weinberg equilibrium; ML:machine learning; LASSO:least absolute shrinkage and selection operator; SVR:support vector regression

## II. METHODS

Our proposed framework includes the quality control (QC) of the data, four feature selection methods such as LASSO [14], ridge [18], elastic-net [19], and mutual information [20], followed by three association methods (i.e. linear regression (LR) [21], random forest (RF) [22], and support vector regression (SVR) [23]). Let *X, Y* = (*X, Y*) *∈* ℝ^*a*×*b*^ is a GWAS dataset of a complex trait for *a* individual and *b* number of SNPs. Let 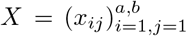, where *x*_*ij*_ is the allele of the *i*^*th*^ individual at *j*^*th*^ SNP and 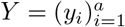 indicates the *a* × 1 quantitative phenotype vector, where *y*_*i*_ is the phenotype value of the *i*^*th*^ individual.

### A. Dataset

#### 1. Real PennCATH dataset

A subset of 1401 patients out of 3850 participated in an angiographic CAD case-control GWA investigation from the PennCATH study. For the subset, 861,473 SNPs were genotyped using the Affymetrix 6.0 GeneChip platform. The Institutional Review Board of the University of Pennsylvania approved the study, and all participants provided informed consent along with details regarding their ethnicity [24]. The details of PennCATH data are explained in the supplementary.

#### 2. Imputed and rare-variant PennCATH dataset

Before imputation process, the PennCATH dataset was phased using SHAPEIT4 method [25]. Afterwards, the phased dataset was imputed using two widely used tools, namely, IMPUTE5 [26] and Beagle5.4 [27]. In order to generate high-quality imputed datasets, stringent thresholds are applied, including imputation scores *>*= 90% in IMPUTE5-generated datasets and probability scores *>*= 70% in Beagle5.4-generated datasets. These stringent criteria are essential for ensuring the reliability and accuracy of the imputed datasets. The total number of SNPs generated after the implementation of IMPUTE5 and Beagle5.4 on the PennCATH dataset was 14,848,075 and 8,803,043, respectively, while maintaining a consistent sample size of 1401 as in the original PennCATH study.

Next, we evaluated the efficiency of the proposed framework using a rare-variants dataset from the imputed PennCATH data. We selected SNPs with MAF between 1% and 5% during the QC step to focus on rare SNPs. The restriction on MAF range was done to prioritize the analysis of rare SNPs, which are often masked by common SNPs in the real PennCATH dataset. This approach aimed to assess the ability of proposed framework to identify and analyze rare genetic associations that conventional GWAS might overlook.

#### 3. Simulated dataset

For the simulated dataset, we utilized genotype data from HapMap3 (http://ccr.coriell.org/Sections/Collections/NHGRI/?SsId=11) and generated phenotype data using the G2P (Genotype-to-Phenotype) simulation tool [28]. The HapMap3 dataset provided a comprehensive genotypic information set comprising 1,397 samples and 1,457,897 SNPs. In the G2P simulation, we simulate a single quantitative trait and set the number of quantitative trait nucleotides (QTNs) to 5,000, which is consistent with our objective of selecting 5,000 SNPs after feature selection methods. The effects of these QTNs were modelled using a normal distribution, and the heritability parameter was set to 0.45. This approach ensured the simulated phenotype data reflected realistic genetic architectures.

### B. QC of dataset

The efficiency of GWAS to identify true genetic connections is dependent on the overall quality of the dataset. Even simple statistical tests of association are hampered by unprocessed genome-wide SNP data, potentially leading to false-negative and false-positive associations. Furthermore, concerns with general data quality will most likely affect subsequent analyses and studies beyond the initial GWAS. [29]. The QC of PennCATH-real dataset was performed by Plink2.0 [30], a free, open-source, cross-platform software, which is widely used for QC and analysis of GWAS data. SNP call rate serves as the initial step in QC. This process entails filtering out SNPs that exhibit high rates of missing data across individuals, with a threshold set at 100%. Subsequently, the sample call rate filter involves the exclusion of samples with substantial amounts of missing data across SNPs, employing a threshold of 95%. The MAF is then assessed, representing the frequency of the least common allele within a particular population, with a threshold of 0.01. LD analysis examines the random association of different genetic loci within the same population, employing a threshold of 0.3. HWE evaluates the relation between allele and genotype frequencies, with SNPs having a p-value less than 1×10^*−*5^ considered as outliers and subsequently excluded from the dataset. Given the population-based nature of the PennCATH study, individuals such as twins, first cousins, and other family members are excluded based on a kinship threshold of 0.0884 [31]. We implemented a relatively relaxed threshold for LD and HWE to facilitate the feature selection approaches.

### C. Feature selection methods

Only a few thousand samples and up to a million SNPs can be found in a genotyping dataset utilized in a GWAS [32]. Feature selection is necessary to reduce the size of the data and the effect of the curse of dimensionality [33]. Moreover, irrelevant and insignificant variants affect ML techniques from uncovering true gene-gene relationships within the dataset [34]. By directly using the dataset to train the ML model, it may learn all noise and random fluctuations in the dataset, which may cause overfitting of the model. The advantages of feature selection methods are that only significant variants are selected, which lowers the dimensionality of the data, accelerates learning, optimizes the learned model and enhances prediction performance [33, 35, 36]. For this purpose, we have used ML models such as LASSO, ridge, elastic-net, and mutual information to select a set of 5000 SNPs. This number was determined based on the Bonferroni correction method, which adjusts the significance threshold (0.05) to reduce false positives by accounting for the number of tests performed [31]. To adhere to this method, we set a suggestive p-value threshold of 1 × 10^*−*5^ for the association test, as referenced from the NHGRI GWAS Catalog [37]. Consequently, to achieve this specified p-value threshold, 5000 SNPs were identified using the feature selection techniques, following the formula: 0.05*/*number of independent variants.

#### 1. LASSO

Equation 1 is linear regression in which *ŷ*_*i*_ is a dependent variable/predicted phenotype and has a linear relationship with allele *x*_*ij*_. *β*_*j*_ is the regression coefficient, i.e. effect sizes of alleles/features, *ϵ*_*i*_ is the normal errors with mean 0 and known variance *σ*^2^.

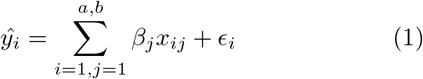

LASSO uses L1 penalty 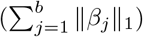 on residual sum of squares (RSS) [14]. The main objective of LASSO is to regularize and select features by minimizing the RSS between actual and predicted values of phenotype (*y*_*i*_ *− ŷ*_*i*_)^2^ with L1 added penalty.

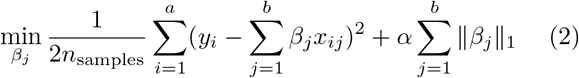

Depending on the penalization parameter or the strength of penalty *α* in equation 2, some coefficients may be precisely zero, which causes only relevant features in the model. It provides a sparse solution and minimizes the model’s variance and bias. If the value of *α* equals zero, then it will become an ordinary least square regression.

#### 2. Ridge regression

Unlike LASSO, ridge regression uses L2 penalty 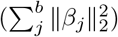 on RSS [18]. The ridge regression objective is to update effect sizes by minimizing the residual sum of squares.

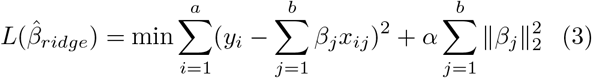

The larger the value of penalization parameter *α* in the above equation 3, the smaller the coefficients become, but never reduce to absolute zero. So, ridge regression does not perform feature selection and cannot reduce model complexity. However, GWAS data have multicollinearity among their features; ridge regression is instrumental in avoiding it [38].

#### 3. Elastic-net

Elastic-net combines both LASSO (L1 penalty) and ridge regression (L2 penalty) [19]. Elastic net may create a greater number of accurately connected features than LASSO. It also has a substantially lower false positive rate than ridge regression [39].

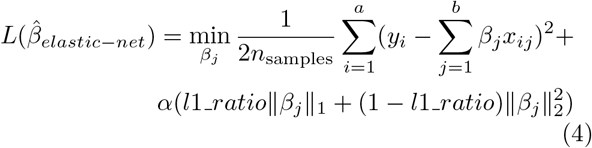

Here, in the above equation 4 *α* value is a strength of penalty, and *l*1 *ratio* is elastic-net mixing parameter with 0 *<*= *l*1 *ratio <*= 1. If *l*1 *ratio* is set to zero, resulting in ridge regression, and if its value is set to one, it is equivalent to a LASSO penalty. Even if there is collinearity among features, elastic-net handles it effectively and keeps mean square error at a minimal [40].

#### 4. Mutual information

Mutual information was first p roposed b y Shannon [20]. This method is widely used for feature selection. Mutual information shows how much dependence between two random variables.

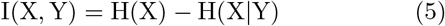

In the above equation 5, I(X, Y) is the mutual information of the X and Y, where H(X) is the entropy of the X, and H(X — Y) is the conditional entropy of the X given Y. Mutual information values can be equal to zero (independent variables) or larger than zero (dependent variables). The larger the mutual information value is, the stronger the correlation between the two random variables. In GWAS, the phenotypic vector is considered one random variable, and the genotype vector is another random variable [41].

### D. Association analysis methods

Among selected SNPs through feature selection, an association of SNPs was performed to determine the most significant SNPs. For that, conventional GWAS has some limitations, such as not taking SNP-SNP interactions into account. Due to the advancement of ML models, such as RF [22] and SVR [23] with statistical method LR [21] are used.

#### 1. Linear regression

LR analysis predicts the value of one variable depending on another. The variable that we predict is known as the dependent variable, i.e., phenotype. The variable used to predict the value of another variable is known as the independent variable, i.e. genotype. This type of analysis determines the coefficients of a linear equation using one or more independent variables that best predict the value of the dependent variable. LR finds a straight line or surface that minimizes the difference between expected and actual output values. Simple LR employs the “least squares” method to determine the best-fit line for a set of paired data points. As explained in equation 1, *x*_*ij*_ is the allele that will predict the value of *ŷ*_*i*_. The regression coefficient is defined as the slope of regression line *β*_*j*_ and measures the effect sizes of the allele *x*_*ij*_ [42].

#### 2. Random forest

The RF method is an extension of the bagging method, as it uses both bagging and feature randomness to generate an uncorrelated forest of decision trees. Feature randomization, or feature bagging, generates a random collection of features, resulting in low correlation among decision trees. The RF algorithm is composed of a collection of decision trees, with each tree in the ensemble consisting of data points selected from a training set with replacement, known as the bootstrap sample. One-third of the training sample is set aside as test data, also known as the out-of-bag (OOB) sample. Another instance of randomization is then introduced by feature bagging, which increases dataset variety while decreasing correlation among decision trees. In a regression job, the individual decision trees will be averaged. Finally, the OOB sample is used for cross-validation to finalize the prediction [22]. The parameters in RF are *n estimators*, which is the number of trees, and *max depth*, which is the maximum depth of the tree [43].

#### 3. Support vector regression

SVR is a potent ML technique for regression applications. It works by maximizing the margin between the hyperplane and the nearest data points, or support vectors, and determining which hyperplane best fits the training set by minimizing the cost function as described in equation 6. SVR uses different kinds of kernels or functions such as sigmoid, polynomial, radial basis function, and linear. SVR uses a number of hyper-parameters that can be changed to regulate the model behaviour [23].

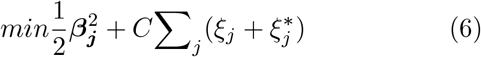

In the above equation 6, the C is the inverse regularization parameter that controls the strength of penalty, and the *ξ* parameter defines the epsilon tube of width |*ξ −ξ*^*∗*^| that the training loss function does not penalize within, with points expected to be close to the actual value of phenotype [43].

#### 4. XGBoost

XGBoost is a proficient machine learning method with great efficiency and predicted accuracy. It is a tree-boosting methodology, which sequentially creates an ensemble of decision trees to correct errors from previous trees, making it highly successful for a wide range of predictive modelling problems. XGBoost’s regularization prevents overfitting, which is critical for increasing model generalization. XGBoost’s objective function comprises a convex loss function that measures the difference between predicted and actual values, as well as a regularization term that penalizes model complexity. This can be expressed as in equation 7:

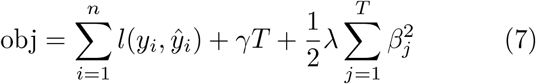

where *l* is the loss function, *T* is the number of leaves, *γ* is the regularization parameter for the number of leaves, and *λ* is the regularization parameter for the leaf weights (*β*_*j*_). XGBoost also enables parallel processing and handles missing information, which improves its adaptability and performance. This method is well known for its capacity to handle large-scale data and generate robust models with excellent predictive power [44].

### E. Evaluation metrics

The performance of each combination of feature selection methods and association methods was evaluated using the coefficient of determination (*R*^2^) [45]. *R*^2^ is a statistical measure that provides information about how well the model fits the data, i.e., goodness of fit in the context of regression.

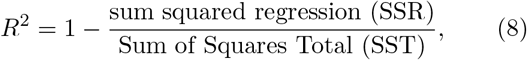

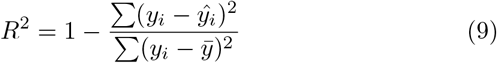

In the above equation 9, *y*_*i*_ is the actual phenotype, *ŷ*_*i*_ is the predicted phenotype, and 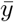 is the mean of the actual phenotype. *R*^2^ elucidates the extent to which the variance in the phenotype is explained by the genotype data of samples. A *R*^2^ = 1 represents that the given genotype dataset accounts for all variations in the phenotype observed in the sample data. Conversely, a *R*^2^ = 0 indicates that the genotype dataset does not explain any of the variations observed in the phenotype. *R*^2^ serves as a measure of how well the genotype predicts the phenotype, with higher values indicating stronger predictive power.

Furthermore, another evaluation metric, MAE, is used, which serves as a measure of model accuracy. MAE calculates the average of the absolute difference between values predicted by the model 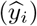 and the actual value (*y*_*i*_), as shown in the equation 10.

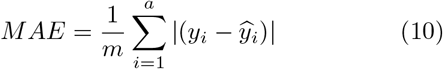

MAE is less influenced by outliers due to its linear weighting, offering a more balanced assessment of model performance. A lower MAE indicates higher accuracy in predicting the phenotype based on genotype data.

## III. RESULT

We performed a comprehensive assessment of four different feature selection methods and four association analysis methods using real PennCATH (common and rare variants), imputed and simulated datasets. The results of our analyses are described below in detail.

### A. Performance evaluation on PennCATH-real dataset

The PennCATH-real dataset comprised 1401 samples and 861,473 SNPs. Several stringent standards were used during data preprocessing to ensure the quality. Some 688,840 SNPs with a call rate *<* 100% were removed for further analysis. Next, SNPs were filtered using two criteria: 1) divergence from Hardy-Weinberg equilibrium (HWE) (*P <* 1 × 10^*−*5^) removed 31 SNPs; 2) minor allele frequency (MAF) *>* 0.01 removed 41,118 SNPs. Some 61,582 SNPs were eliminated by LD pruning (r2 *<* 0.3). All 1401 samples had the call rate *>* 95%. Kinship analysis was performed using a threshold of 0.09, which did not remove any samples. After implementing all quality control (QC) measures and eliminating missing values from the phenotypic data, the final dataset consisted of 1282 samples and 69,902 SNPs.

Feature selection was performed to remove irrelevant or ambiguous SNPs with little or no predictive importance for the phenotype. We applied four different feature selection approaches on the above preprocessed PennCATH dataset. For LASSO method, we used an *α* = 4.5 × 10^*−*4^, which resulted in 5003 SNPs. The ridge regression identified 5000 SNPs at an *α* value of 5 × 10^*−*3^. Likewise, elastic-net approach resulted in 5037 SNPs with an *α* value of 3.3 × 10^*−*3^ and an l1 ratio of 0.5. The mutual-information method selected the top 5000 SNPs based on their mutual information scores. The Venn diagram shows the number of overlapping SNPs selected by four different feature selection methods (see Figure 2). Some 80 SNPs were shared by all four methods. The maximum overlap of 1572 SNPs was observed between elastic-net and ridge regression methods. LASSO and mutual information methods shared 1142 SNPs.

**FIG. 2.**
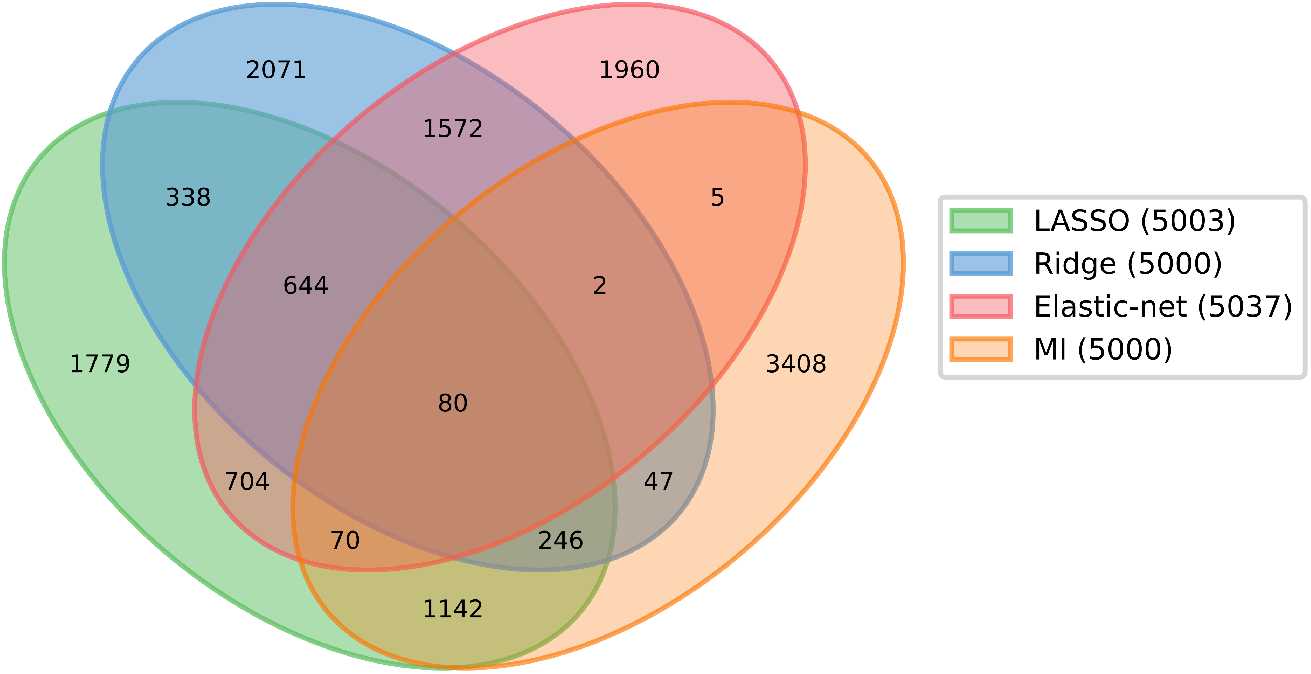
Venn diagram illustrating the distribution of shared and distinct SNPs among four feature selection methods. 80 SNPs were common across all four methods. The legend shows the four feature selection methods and the number of SNPs selected by each method in parenthesis Abbreviations- LASSO:least absolute shrinkage and selection operator; MI:mutual information

After the feature selection step, the selected SNPs were tested for association with LDL level as phenotype using four different methods viz. LR, RF, SVR, and XGBoost. The performance of each method was evaluated using the *R*^2^ and mean absolute error (MAE). When different feature selection methods were paired with various association methods, majority of the combinations displayed lower *R*^2^ values and higher MAE values. For instance, combinations of elastic-net with RF under-performed with *R*^2^ = -13.75 and MAE = 26.02 (see Table I). Notably, elastic-net combined with SVR outperformed other combinations, achieving an *R*^2^ of 0.84 and an MAE of 8.67.

**TABLE I.** Performance evaluation of various combinations of feature selection and association methods for all five datasets. **Bold values** denote the best performance value.

| Association methods →<br>Feature selection ↓ | Linear Regression<br>(LR) |  | Random Forest<br>(RF) |  | Support Vector<br>Regression (SVR) |  | XGBoost |  |
| --- | --- | --- | --- | --- | --- | --- | --- | --- |
|  | PennCATH dataset |  |  |  |  |  |  |  |
| | $R^2$ | MAE | $R^2$ | MAE | $R^2$ | MAE | $R^2$ | MAE |
| LASSO | 0.013 | 16.056 | -41.106 | 26.329 | 0.380 | 15.416 | -7.27 | 26.94 |
| Ridge | 0.603 | 11.576 | -16.344 | 25.972 | 0.693 | 11.597 | -2.42 | 28.73 |
| Elastic net | 0.735 | 9.961 | -13.749 | 26.018 | **0.844** | **8.666** | -5.26 | 27.82 |
| Mutual Information | -2.706 | 31.257 | -75.594 | 26.451 | -2.169 | 32.073 | -5.65 | 28.32 |
| Imputed dataset with IMPUTE5 |  |  |  |  |  |  |  |  |
| LASSO | 0.23 | 14.69 | -41.12 | 25.91 | 0.50 | 14.31 | -10.44 | 26.65 |
| Ridge | 0.75 | 10.27 | -12.19 | 25.31 | 0.78 | 10.64 | -6.26 | 27.09 |
| Elastic net | 0.88 | 7.29 | -13.44 | 26.02 | **0.93** | **6.00** | -5.87 | 26.06 |
| Mutual Information | -2.58 | 30.88 | -260.36 | 26.74 | -2.02 | 31.41 | -12.01 | 27.32 |
| Imputed dataset with Beagle5.4 |  |  |  |  |  |  |  |  |
| LASSO | 0.36 | 14.07 | -59.45 | 25.98 | 0.58 | 13.68 | -6.06 | 26.44 |
| Ridge | 0.77 | 9.82 | -61.72 | 25.81 | 0.80 | 10.33 | -5.58 | 27.26 |
| Elastic net | 0.89 | 6.86 | -55.05 | 25.80 | **0.93** | **5.88** | -6.18 | 24.98 |
| Mutual Information | -2.42 | 32.26 | -59.56 | 27.03 | -1.94 | 32.92 | -8.49 | 28.17 |
| Rare-variants PennCATH dataset |  |  |  |  |  |  |  |  |
| LASSO | -7.06 | 24.62 | -123.18 | 28.38 | -3.46 | 24.24 | -12.14 | 28.69 |
| Ridge | 0.23 | 14.90 | -69.57 | 29.07 | 0.45 | 13.91 | -14.00 | 30.17 |
| Elastic net | 0.58 | 10.75 | -83.35 | 28.26 | **0.78** | **8.65** | -16.67 | 28.05 |
| Mutual Information | -8.96 | 29.35 | -106.02 | 29.06 | -5.11 | 30.62 | -12.25 | 30.78 |
| Simulated dataset |  |  |  |  |  |  |  |  |
| LASSO | 0.57 | 12.46 | -1.10 | 20.64 | 0.72 | 12.16 | -0.75 | 21.47 |
| Ridge | 0.74 | 10.57 | -25.45 | 24.13 | 0.74 | 12.86 | -6.51 | 25.53 |
| Elastic net | 0.89 | 7.36 | -0.95 | 20.18 | **0.90** | **7.78** | -0.87 | 21.23 |
| Mutual Information | -1.67 | 32.10 | -27.23 | 25.11 | -1.94 | 31.80 | -6.63 | 26.31 |

The hyperparameter tuning for RF and XGBoost models was done, as described in the supplementary material, to optimize their performance. For the RF model, we obtained the following optimal parameters: *max depth* = None, *min samples leaf* = 4, *min samples split* = 2, and *n estimators* = 400. Similarly, optimal parameters for XGBoost model were as follows: *learning rate* = 0.1, *max depth* = 3, and *n estimators* = 300. Despite using above-optimized parameters, both RF and XGBoost could not perform well, as shown in Table I. This is probably due to the fact that tree-based methods like RF and XGBoost face challenges in predicting unseen values. Their predictions tend to be constrained between the maximum and minimum values observed in the training data. This results in an average prediction of observed values. This limitation is particularly evident when predicting patterns for quantitative data such as LDL cholesterol levels [46].

Furthermore, we computed the permutation importance scores as per details provided in [47] to assess the relative importance of each selected SNP in explaining the variance of LDL-cholesterol level. These scores indicate how much the predictive accuracy of the model decreases when a particular SNP is randomly shuffled. Higher permutation importance scores suggest the greater influence of the SNP on the phenotype. We chose top 100 SNPs based on the permutation importance scores having the most impact on LDL-cholesterol levels. A comparative graph of the permutation importance scores for both the selected 5,000 SNPs and the top 100 SNPs is provided in supplementary material (see Figure S1).

### B. Performance evaluation on other datasets

The performance of feature selection and association analysis methods was further assessed on two imputed, one rare-variant, and one simulated dataset. The basic QC steps were applied on all four datasets, as described in the Methods section. Next, we separately passed the preprocessed datasets to four feature selection methods to select approximately 5000 SNPs from each method. Afterwards, we performed association analysis on these selected SNPs using four ML-based methods, adhering to the same hyperparameters as outlined for the PennCATH-real dataset. The results obtained from different combinations of feature selection and association analysis methods using imputed, rare-variant, and simulated datasets are summarized in Table I. Again, elastic-net combined with SVR outperformed other combinations of feature selection and association analysis methods, consistent with the results obtained from PennCATH-real dataset.

## IV. POST-GWAS ANALYSES

We performed further post-GWAS analyses of top 100 SNPs identified from the permutation importance score of PennCATH-real dataset to elucidate their functional and biological role in LDL-cholesterol regulation. These analyses provided deeper insights into the genetic mechanisms underlying LDL cholesterol levels and identified potential targets for therapeutic intervention.

### A. Comparison with previous findings

At first, we conducted a comprehensive literature search to ascertain the experimentally validated functions of SNPs identified in our analysis. One SNP rs4591370 obtained from PennCATH-real dataset has been previously reported to be significantly (*P* = 8.2 × 10^*−*9^) associated with circulating LDL-cholesterol concentrations [48]. Further, rs7232775 showed association with blood urea nitrogen (*P* =8.0 × 10^*−*14^) [49]; and rs12438724 has been associated with fibrotic idiopathic interstitial pneumonias (*P* =4.10 × 10^*−*8^) [50]. Both these SNPs have been related to cholesterol levels in blood serum. Next, we also searched the genome-wide repository of associations between SNPs and phenotypes (GRASP) [51] database to get relevant literature associated with our resultant SNPs. The query results revealed that the majority of the identified SNPs displayed causal relationships with LDL-cholesterol levels. Notably, among 100 identified SNPs, 23 were experimentally validated to be associated with LDL cholesterol and/or LDL cholesterol-related diseases such as coronary artery disease (CAD), blood pressure, and Alzheimer’s disease. Table S5 provides detailed information on these 23 SNPs with their corresponding p-values.

### B. Rare variants analysis

We used the RAVAR database [52] to elucidate the functional role of rare variants identified in our study.

In total, we identified 113 genes that are reported in the RAVAR database as having functional roles in cholesterol-related traits and diseases, such as very low-density lipoprotein cholesterol measurement, LDL cholesterol, hypertrophic cardiomyopathy, blood pressure, subcutaneous adipose tissue measurement, HbA1c measurement, anxiety disorder, and free cholesterol:total lipids ratio. These 113 genes were identified using SKAT [53], burden test [54], and collapsing analysis [54]. A detailed description of these genes is provided in the supplementary Table S3 (PennCATH rare-variant), S4 (PennCATH-imputed from Beagle5.4), and S5 (PennCATH-imputed from IMPUTE5).

### C. eQTL analysis

The expression quantitative trait locus (eQTL) analysis serves as a fundamental tool for elucidating the regulation of gene expression. The genotype-tissue expression (GTEx) [55] and eQTLGen consortium [56] were utilized to investigate the SNP expression across specific tissues and whole blood. Additionally, we assessed the expression of the genes using functional mapping and annotation of genome-wide association studies (FUMAGWAS) [57]. The GENE2FUNC tool of FUMAGWAS was used to assess tissue specificity based on the genes annotated from identified SNPs, that are more (or less) expressed in a specific tissue compared to all other tissues. Among the top 100 SNPs identified through our framework, eQTL analysis resulted in 64 SNPs with significantly higher expression levels (*P <* 0.05) in various tissues, including whole blood, adipose (both visceral and subcutaneous), fibroblast, oesophagus (gastroesophageal junction and mucosa), heart, artery, pancreas, brain, and thyroid. These organs are also identified in the expression analysis of genes annotated from identified SNPs from FUMAGWAS. The differentially expressed genes (DEG) sets were significantly enriched for heart, liver, pancreas, blood, etc. These tissues play a key role in regulating cholesterol levels. Figure 3A highlights the affected tissues/organs resultant from both eQTL and expression analyses. All eQTLs and non-eQTLs identified from our framework are shown in Figure 3B. Furthermore, differential expression analysis of annotated genes highlights the presence of state-of-the-art genes implicated in LDL-cholesterol regulation. For instance, APOB gene, a well-known carrier of LDL cholesterol, exhibited differential expression in the liver, small intestine, and heart [58, 59]. Also, RAB2 gene is involved in cholesterol efflux mechanisms [60] and TRAPPC9 is associated with serum LDL cholesterol levels [61]. Both these genes showed differential expression patterns, further emphasizing their importance in cholesterol homeostasis. A heatmap of the corresponding genes and their expression is visualised in Figure 3D. The associated p-values of these SNPs can be found in supplementary Table S4. These findings describe genetic variations impacting various tissues/organs, leading to different traits and diseases due to fluctuating cholesterol levels.

**FIG. 3.**
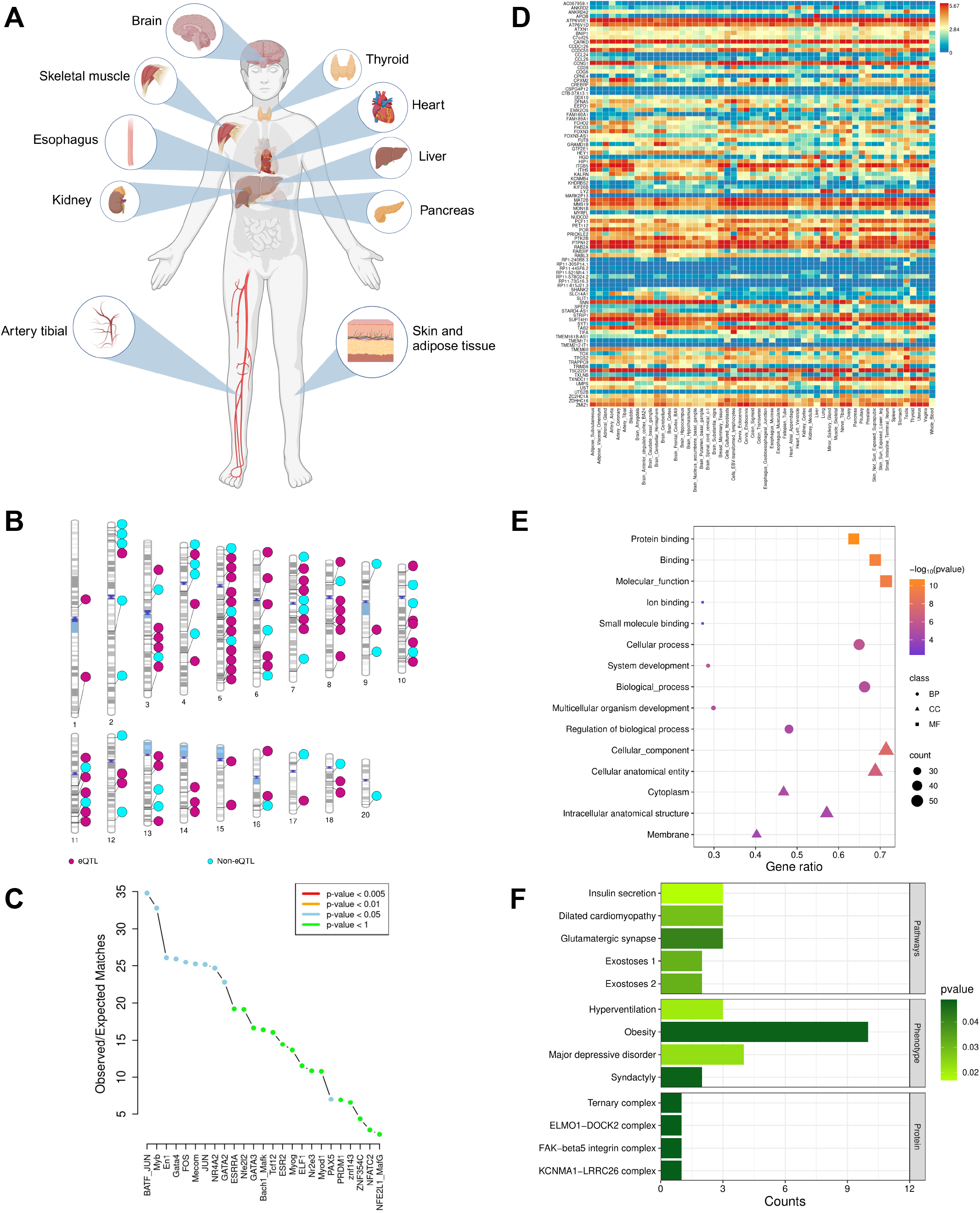
Post-GWAS analysis of top 100 SNPs identified using PennCATH-real dataset through our proposed framework. **(A)** Visualization of tissues/organs exhibiting enrichment of identified SNPs and annotated genes, affected by fluctuations in LDL cholesterol levels, **(B)** Chromosomal mapping of eQTLs and non-eQTLs in identified SNPs, **(C)** Enrichment-plot of TFs detected from identified SNPs, **(D)** heatmap of DEG annotated from identified SNPs in corresponding tissues, **(E)** GO analysis of annotated genes, **(F)** Top pathways, phenotypes, and proteins enriched from annotated genes. Abbreviations- GWAS:genome-wide association studies; SNP:single nucleotide polymorphism; LDL:low-density lipoprotein; eQTL:expression quantitative trait locus; TF:transcription factor; DEG:differentially expressed genes; GO:gene ontology; BP:biological process; CC:cellular component; MF:molecular function

### D. Variants involved in transcriptional regulation

In order to interpret the effects of SNPs, it is crucial to study their impact on gene regulation mechanisms. SNPs often alter gene regulation through changes to transcription factor binding sites (TFBS) [62]. Since intronic and intergenic regions have the highest concentration in identified SNPs, searching for TFBS could be beneficial. This can help to understand the molecular pathways through which SNPs impact the phenotype of interest. SNP2TFBS database [63, 64] was used to identify and visualize the SNPs affecting the TFBSs. Transcription factor (TF) enrichment plot generated from SNPs identified in our proposed pipeline highlights significant associations (*P <* 0.05) with cholesterol-related TF (Figure 3C). For instance, the absence of GATA4 TF leads to increased plasma cholesterol levels [65] and has an association with LDL-cholesterol [66]. Overexpression of GATA2 leads to increased cholesterol efflux from macrophages [67]. GATA2 also play a role in regulating cholesterol storage [68]. Inhibition of FOS TF impacts cholesterol biosynthesis, and JUN family TFs, acting as heterodimeric partners of FOS, influence cell membrane composition in the presence of cholesterol [69]. The absence of MECOM TF reduces LDL uptake by human umbilical vein endothelial cells by four-fold [70]. Additionally, NR4A2 TF inhibits oxidized LDL uptake by macrophages, diminishes pro-inflammatory cytokine and chemokine expression, and is associated with blood pressure regulation [71, 72]. Moreover, we identified important TF binding sites such as BATF JUN, Myb, En1, and PAX5, enriching our understanding of cholesterol-related transcriptional regulation.

### E. Functional enrichment analysis

Since most of the resultant SNPs are located in non-coding regions of the genome, we conducted regulatory region enrichment analysis using HaploReg v4.2 [73], based on data from the Roadmap Epigenomics Consortium. Majority of the identified SNPs, along with their proxy SNPs within the LD range (*r*^2^ *>* 0.8), show significant enrichment for enhancer regions (including promoter and enhancer histone modifications) DNase sensitivity, and motif alterations, particularly in whole blood, heart (left ventricle), and pancreas tissues. Detailed results are provided in the supplementary Table S6.

The enrichment analysis was performed to reveal significantly enriched biological processes, pathways, regulatory motifs and protein complexes. We used g:Profiler, a web server for functional enrichment analysis, to perform the gene ontology (GO) analysis [74]. The significantly enriched molecular functions, biological processes, and cellular components involved majority of binding functions like protein, ion and small molecule, cellular and developmental processes, and cytoplasm and membrane, respectively. The top GO terms are shown in Figure 3E. The pathway enrichment analysis revealed pathways related to insulin secretion, synapse and heparan sulphate.

### F. Functional gene annotation

The annotated genes from identified SNPs were also involved in various cholesterol-modulated phenotypes. For instance, TRAPPC9 gene [61] is responsible for obesity-related traits (a condition characterized by elevated LDL cholesterol) [75]. Additionally, higher LDL cholesterol has been linked to major depressive disorders [76], and genes associated with these conditions, such as CCL24 [77], have been identified as well. Conversely, genes responsible for syndactyly, a condition characterized by fused digits, were identified, which correlated with decreased serum cholesterol levels [78, 79]. Notably, proteins such as fructose-bisphosphatase-1 (FBPase) and synaptotagmin-1 were identified as regulators of abnormal cardiac atrium morphology, such as atrial fibrillation (AF), where both low cholesterol levels and high cholesterol variability were linked to increased AF risk [80]. Furthermore, these two proteins are also implicated in hyperventilation conditions. Liver FBPase is known to be upregulated by obesity and dietary fat intake, suggesting a potential link between LDL and FBPase activity [81]. Cholesterol depletion disrupts synaptotagmin-1-induced membrane bending, which impairs synaptic transmission and neuronal function [82]. Additionally, glypican6 regulates cardiomyocyte growth via the ERK1/2 signalling pathway [83], which is also involved in cholesterol trafficking [84]. Gustafsen, Camilla, *et al*. suggested in their study to investigate the specificity of the interaction between glypicans and PCSK9 [85], which may give a biological understanding of increasing circulating LDL cholesterol levels [86]. Moreover, EEPD1 gene (endonuclease-exonuclease-phosphatase family domain containing 1) functions as part of the Liver X Receptors (LXRs)-regulated program, promoting ABCA1-dependent (ATP-binding cassette transporter A1) cholesterol efflux from macrophages [87].

The identified SNPs were annotated to proteins using the CORUM database within g:Profiler, a resource that manually annotates protein complexes from mammalian organisms. Among the annotated protein complexes, we identified several noteworthy associations with cholesterol. Firstly, the Ternary complex containing GATA4, SRF, and MYOCD was observed, wherein cholesterol loading suppressed the expression of MYOCD [88]. Additionally, the ELMO1-DOCK2 complex, annotated from identified SNPs, revealed cholesterol sulfate as a potent inhibitor of DOCK2 [89]. The FAK-beta5 integrin complex (VEGF-induced complex) demonstrated cholesterol’s regulatory role in VEGF:VEGFR-1 signalling, influencing cell migration and viability, potentially impacting disease progression [90] and also regulating the innate immune response [91]. Furthermore, the KCNMA1-LRRC26 complex highlighted the importance of KCNMA1 in cholesterol transport, as evidenced by significant cholesterol accumulation in the absence of KCNMA1 in mouse embryonic fibroblasts. This finding suggests a conserved role for KCNMA1 in the efficient cholesterol transport, implicating its importance in human physiology as well [92]. The top enriched pathways, phenotypes, and proteins are displayed in Figure 3E.

### G. Drug targets analysis

In this study, we analyzed potential drug targets among resultant genes associated with diseases influenced by cholesterol level fluctuations. For instance, Mipomersen, an antisense oligonucleotide drug targeting the APOB gene, is used to treat homozygous familial hypercholesterolemia by inhibiting APOB synthesis [93]. Similarly, Baricitinib and Leflunomide, anti-rheumatic drugs targeting the PTK2B gene, are used to treat Alzheimer’s disease by inhibiting Janus kinases [94]. Auranofin, acting through the PTPN12-ErbB-2 signalling axis, reduces damage from myocardial ischemia/reperfusion injuries by targeting the PTPN12 gene [95].

### H. Gene targets for personalized medicine treatments

Our pipeline also identifies genes that can be utilized in personalized medicine treatments. For instance, deficiency in the conserved oligomeric Golgi complex subunit 6 (COG6) gene causes congenital disorders of glycosylation (CDG), a rare autosomal recessive disease. Li G. *et al*. conducted a targeted NGS study on COG6, uncovering compound heterozygous variants that broaden the mutation spectrum and extend the genotype-phenotype relationship in CDG [96]. Moreover, ATXN1, a dosage-sensitive gene, is involved in neurodegenerative disorders like spinocerebellar ataxia type 1 and Alzheimer’s disease. Nitschke L. *et al*. reported that mutations in miR760’s binding site within the 5’ UTR or in the 3’ UTR binding sites of miRNAs and RNA-binding proteins could increase ATXN1 expression, causing ataxia symptoms. These findings underscore the importance of identifying ATXN1 regulatory regions and performing whole-genome sequencing in ataxia patients to identify potential disease-causing mutations in non-coding regions [97].

## V. DISCUSSION

The emergence of GWAS has significantly enriched our understanding of human disease genetics in the past two decades, transitioning from analyzing common variants to exploring rare variants. GWAS findings provide valuable insights into disease biology, aiding in clinical application and revealing population-level risk stratification. However, conventional GWAS methods have several limitations in identifying causal variants accurately. For instance, in our analysis of the PennCATH dataset using a conventional GWAS pipeline [30] at a significance threshold of 5×10^*−*5^, only five SNPs were selected, which shows insufficient statistical power of conventional methods.

In this work, we integrated ML-based feature selection and association analysis to mitigate false positives, evaluate epistasis, and account for missing heritability. Our proposed approach is capable of managing the high dimensionality inherent in GWAS data and provides alternatives to stringent p-value thresholds. We used various ML techniques for feature selection, including LASSO, ridge, elastic-net, and mutual information. Each of these techniques selected approximately 5000 SNPs on the PennCATH-real dataset. Further, we evaluated the associations between these selected SNPs and LDL cholesterol levels using LR, RF, SVR, and XGBoost methods. Notably, employing the elastic net for feature selection in combination with SVR yielded promising results, as evidenced by an *R*^2^ value of 0.84 and an MAE of 8.66. However, other feature selection and association algorithm combinations did not perform well. We also considered a popular non-linear ML method, XGBoost, for association tests, which is known to have a comparative performance in case-control GWAS [12]. However, in our analysis, it appeared that this method did not perform well for continuous complex trait studies. Next, based on permutation feature importance scores, we ranked the top 100 most effective SNPs out of 5000 selected SNPs from the combination of elastic-net and SVR methods.

In post-GWAS analyses, we conducted functional enrichment to gain insights into the biological significance of 100 SNPs identified from our proposed framework. Initially, we studied the expression patterns of these SNPs across different tissues and observed significant expression levels in tissues/organs known to play crucial roles in regulating cholesterol levels, including whole blood, adipose tissue, heart, and pancreas. Additionally, our study demonstrates the significant enrichment of down-regulated and both-sided DEG sets in the same tissues/organs. Notably, genes such as APOB, RAB2, and TRAPPC9 exhibit differential expression patterns across various tissues, highlighting their potential role in cholesterol homeostasis. Although most of the identified SNPs are located in non-coding regions, they are enriched for regions that activate transcription through TFBS. Key findings of such SNPs include the role of GATA4 TF in plasma cholesterol regulation, the impact of GATA2 on cholesterol efflux and storage, and the influence of FOS and JUN TFs on cholesterol biosynthesis and cell membrane composition. Additionally, MECOM deficiency affects LDL uptake, while NR4A2 inhibits oxidized LDL uptake. Moreover, the identified SNPs play significant roles in enhancer regions, exhibit DNase activities, and show changes in motif regions in whole blood, heart, and pancreas tissues. Furthermore, GO analysis highlighted the involvement of annotated genes in binding functions, cellular and developmental processes, as well as cytoplasmic and membrane-related activities. Pathway enrichment analysis unveiled significant enrichment of pathways associated with insulin secretion, synapse function, and heparan sulfate metabolism, indicating potential biological mechanisms underlying the observed associations with LDL-cholesterol. Additionally, the annotated genes exhibit the intricate role of LDL cholesterol in regulating obesity, major depressive disorders, syndactyly, abnormal cardiac morphology, and neuronal function. Furthermore, the involvement of key proteins such as FBPase, synaptotagmin-1, and glypican-6 suggests potential pathways through which LDL-cholesterol modulates various physiological processes. The potential of synaptotagmin-1 as a therapeutic target for neurodegenerative and neurodevelopmental diseases is noteworthy due to its involvement in synaptic transmission and neuronal function. Addressing the specificity of interactions between glypicans and PCSK9 may offer insights into mechanisms influencing circulating LDL-cholesterol levels. Moreover, the annotation of identified SNPs to protein complexes further reveals the importance of cholesterol in modulating cellular functions and signalling pathways, highlighting its potential implications in innate immunity, disease progression and cellular homeostasis. In addition, our proposed framework identifies potential drug targets among genes associated with cholesterol-related diseases. For example, Mipomersen targets APOB for treating familial hypercholesterolemia, Auranofin targets PTPN12 to reduce myocardial ischemia, while Baricitinib and Leflunomide target PTK2B for Alzheimer’s. Further, genes like COG6 and ATXN1 hold promise for personalized medicine in conditions like CDG and neurodegenerative disorders, respectively. In summary, the identified SNPs, genes, and proteins provide valuable insights into potential therapeutic targets for managing cholesterol-related diseases. In the future, GWAS datasets from diverse populations should be included to enhance the generalizability of findings. By incorporating samples from various populations would gain more statistical power and provide a comprehensive exploration of genetic and environmental factors. The current challenge in our study is the inability to use other large GWAS datasets in our framework due to their unavailability. Future studies should focus on incorporating these large datasets to improve the robustness and reliability of our proposed framework for detecting genetic associations related to complex traits. This would enable a more thorough evaluation of our methodology and its effectiveness in identifying meaningful genetic associations across diverse populations, thereby advancing our understanding of complex diseases. Our findings on LDL cholesterol indicate that integrating diverse datasets can reveal crucial insights into the genetic architecture and identify potential therapeutic targets for LDL cholesterol-related traits and diseases. By broadening the scope of genetic data and considering population-specific genetic variations, we can better understand the complexities of disease mechanisms and develop more effective, personalized therapeutic strategies.

In conclusion, our research demonstrates the efficacy of incorporating ML techniques, particularly elastic-net for feature selection and SVR for association analysis, on both real and simulated datasets. Our approach effectively addresses the limitations of conventional GWAS methods. Additionally, our results yield significant biological insights. This novel framework not only improves the detection of genetic associations with complex traits but also holds promise for the development of targeted medications and personalized treatments.

## Data Availability

This study utilizes publicly available data, and the access identifiers for these data are provided inside the manuscript. The PennCATH cohort study is available at \url{https://pbreheny.github.io/adv-gwas-tutorial/quality_control.html}.

https://pbreheny.github.io/adv-gwas-tutorial/quality_control.html

## VI. DECLARATIONS

### A. Ethical Approval

Patient consent and ethical approval for publication are not applicable.

## VII. DATA AVAILABILITY

This study utilizes publicly available data, and the access identifiers for these data are provided inside the manuscript. The PennCATH cohort study is available at https://pbreheny.github.io/adv-gwas-tutorial/quality_control.html.

### A. Consent for publication

“Not applicable”.

### C. Author’s Contributions

V.J. and J.S. contributed equally to this work. V.J. performed the data analysis. J.S. and R.S.S. contributed to the biological enrichment and interpretation of these enrichment results. J.S. and P.Y. contributed to the concept and design of the paper. All the authors contributed to the writing and revision of the manuscript.

## References

[1] R. J. Loos, 15 years of genome-wide association studies and no signs of slowing down, Nature Communications 11, 5900 (2020).

[2] S. Nurk, S. Koren, A. Rhie, M. Rautiainen, A. V. Bzikadze, A. Mikheenko, M. R. Vollger, N. Altemose, L. Uralsky, A. Gershman, et al., The complete sequence of a human genome, Science 376, 44 (2022).

[3] T. Wang, L. Antonacci-Fulton, K. Howe, H. A. Lawson, J. K. Lucas, A. M. Phillippy, A. B. Popejoy, M. Asri, C. Carson, M. J. Chaisson, et al., The human pangenome project: a global resource to map genomic diversity, Nature 604, 437 (2022).

[4] T. M. Frayling, N. J. Timpson, M. N. Weedon, E. Zeggini, R. M. Freathy, C. M. Lindgren, J. R. Perry, K. S. Elliott, H. Lango, N. W. Rayner, et al., A common variant in the fto gene is associated with body mass index and predisposes to childhood and adult obesity, Science 316, 889 (2007).

[5] K. A. Siminovitch, Ptpn22 and autoimmune disease, Nature genetics 36, 1248 (2004).

[6] K. Wang, H. Zhang, S. Kugathasan, V. Annese, J. P. Bradfield, R. K. Russell, P. M. Sleiman, M. Imielinski, J. Glessner, C. Hou, et al., Diverse genome-wide association studies associate the il12/il23 pathway with crohn disease, The American Journal of Human Genetics 84, 399 (2009).

[7] G. Peng, L. Luo, H. Siu, Y. Zhu, P. Hu, S. Hong, J. Zhao, X. Zhou, J. D. Reveille, L. Jin, et al., Gene and pathway-based second-wave analysis of genome-wide association studies, European Journal of Human Genetics 18, 111 (2010).

[8] R. M. Cantor, K. Lange, and J. S. Sinsheimer, Prioritizing gwas results: a review of statistical methods and recommendations for their application, The American Journal of Human Genetics 86, 6 (2010).

[9] Q. Zhang, Q. Long, and J. Ott, Apriorigwas, a new pattern mining strategy for detecting genetic variants associated with disease through interaction effects, PLoS computational biology 10, e1003627 (2014).

[10] B. Han, X.-w. Chen, Z. Talebizadeh, and H. Xu, Genetic studies of complex human diseases: characterizing snp-disease associations using bayesian networks, BMC systems biology 6, 1 (2012).

[11] E. E. Eichler, J. Flint, G. Gibson, A. Kong, S. M. Leal, J. H. Moore, and J. H. Nadeau, Missing heritability and strategies for finding the underlying causes of complex disease, Nature reviews genetics 11, 446 (2010).

[12] A. Medvedev, S. Mishra Sharma, E. Tsatsorin, E. Nabieva, and D. Yarotsky, Human genotype-to-phenotype predictions: Boosting accuracy with nonlinear models, PloS one 17, e0273293 (2022).

[13] T. T. Wu, Y. F. Chen, T. Hastie, E. Sobel, and K. Lange, Genome-wide association analysis by lasso penalized logistic regression, Bioinformatics 25, 714 (2009).

[14] R. Tibshirani, Regression shrinkage and selection via the lasso, Journal of the Royal Statistical Society Series B: Statistical Methodology 58, 267 (1996).

[15] Z. Chen, M. Boehnke, X. Wen, and B. Mukherjee, Revisiting the genome-wide significance threshold for common variant gwas, G3 11, jkaa056 (2021).

[16] N. Pudjihartono, T. Fadason, A. W. Kempa-Liehr, and J. M. O’Sullivan, A review of feature selection methods for machine learning-based disease risk prediction, Frontiers in Bioinformatics 2, 927312 (2022).

[17] S. Cho, K. Kim, Y. J. Kim, J.-K. Lee, Y. S. Cho, J.-Y. Lee, B.-G. Han, H. Kim, J. Ott, and T. Park, Joint identification of multiple genetic variants via elastic-net variable selection in a genome-wide association analysis, Annals of Human Genetics 74, 416 (2010).

[18] A. E. Hoerl and R. W. Kennard, Ridge regression: Biased estimation for nonorthogonal problems, Technometrics 12, 55 (1970).

[19] H. Zou and T. Hastie, Regularization and variable selection via the elastic net, Journal of the Royal Statistical Society Series B: Statistical Methodology 67, 301 (2005).

[20] C. E. Shannon, A mathematical theory of communication, The Bell system technical journal 27, 379 (1948).

[21] R. R. Hocking, Developments in linear regression methodology: 1959–l982, Technometrics 25, 219 (1983).

[22] L. Breiman, Random forests, Machine learning 45, 5 (2001).

[23] C. Cortes and V. Vapnik, Support-vector networks, Machine learning 20, 273 (1995).

[24] A. e. a. Helgadottir, A variant of the gene encoding leukotriene a4 hydrolase confers ethnicity-specific risk of myocardial infarction, Nature genetics 38, 68 (2006).

[25] O. Delaneau, J.-F. Zagury, M. R. Robinson, J. L. Marchini, and E. T. Dermitzakis, Accurate, scalable and integrative haplotype estimation, Nature communications 10, 5436 (2019).

[26] S. Rubinacci, O. Delaneau, and J. Marchini, Genotype imputation using the positional burrows wheeler transform, PLoS genetics 16, e1009049 (2020).

[27] B. L. Browning, Y. Zhou, and S. R. Browning, A one-penny imputed genome from next-generation reference panels, The American Journal of Human Genetics 103, 338 (2018).

[28] Y. Tang and X. Liu, G2p: a genome-wide-association-study simulation tool for genotype simulation, phenotype simulation and power evaluation, Bioinformatics 35, 3852 (2019).

[29] S. Turner, L. L. Armstrong, Y. Bradford, C. S. Carlson, D. C. Crawford, A. T. Crenshaw, M. de Andrade, K. F. Doheny, J. L. Haines, G. Hayes, et al., Quality control procedures for genome-wide association studies, Current protocols in human genetics 68, 1 (2011).

[30] C. C. Chang, C. C. Chow, L. C. Tellier, S. Vattikuti, S. M. Purcell, and J. J. Lee, Second-generation plink: rising to the challenge of larger and richer datasets, Gigascience 4, s13742 (2015).

[31] A. T. Marees, H. de Kluiver, S. Stringer, F. Vorspan, E. Curis, C. Marie-Claire, and E. M. Derks, A tutorial on conducting genome-wide association studies: Quality control and statistical analysis, International Journal of Methods in Psychiatric Research 27, e1608 (2018).

[32] S. Szymczak, J. M. Biernacka, H. J. Cordell, O. González-Recio, I. R. König, H. Zhang, and Y. V. Sun, Machine learning in genome-wide association studies, Genetic epidemiology 33, S51 (2009).

[33] I. Guyon and A. Elisseeff, An introduction to variable and feature selection, Journal of machine learning research 3, 1157 (2003).

[34] J. H. Moore, F. W. Asselbergs, and S. M. Williams, Bioinformatics challenges for genome-wide association studies, Bioinformatics 26, 445 (2010).

[35] L. Yu and H. Liu, Efficient feature selection via analysis of relevance and redundancy, The Journal of Machine Learning Research 5, 1205 (2004).

[36] N. Pudjihartono, T. Fadason, A. W. Kempa-Liehr, and J. M. O’Sullivan, A review of feature selection methods for machine learning-based disease risk prediction, Frontiers in Bioinformatics 2, 927312 (2022).

[37] D. Welter, J. MacArthur, J. Morales, T. Burdett, P. Hall, H. Junkins, A. Klemm, P. Flicek, T. Manolio, L. Hindorff, and H. Parkinson, The NHGRI GWAS Catalog, a curated resource of SNP-trait associations, Nucleic Acids Research 42, D1001 (2013).

[38] N. Malo, O. Libiger, and N. J. Schork, Accommodating linkage disequilibrium in genetic-association analyses via ridge regression, The American Journal of Human Genetics 82, 375 (2008).

[39] G. Tutz and J. Ulbricht, Penalized regression with correlation-based penalty, Statistics and Computing 19, 239 (2009).

[40] P. Bühlmann and S. Van De Geer, Statistics for high-dimensional data: methods, theory and applications (Springer Science & Business Media, 2011).

[41] H. Guo, Z. Yu, J. An, G. Han, Y. Ma, and R. Tang, A two-stage mutual information based bayesian lasso algorithm for multi-locus genome-wide association studies, Entropy 22, 329 (2020).

[42] A. Schneider, G. Hommel, and M. Blettner, Linear regression analysis: part 14 of a series on evaluation of scientific publications, Deutsches Ärzteblatt International 107, 776 (2010).

[43] F. Pedregosa, G. Varoquaux, A. Gramfort, V. Michel Thirion, O. Grisel, M. Blondel, P. Prettenhofer, R. Weiss, V. Dubourg, J. Vanderplas, A. Passos, D. Cournapeau, M. Brucher, M. Perrot, and E. Duchesnay, Scikit-learn: Machine learning in Python, Journal of Machine Learning Research 12, 2825 (2011).

[44] T. Chen and C. Guestrin, Xgboost: A scalable tree boosting system, in Proceedings of the 22nd acm sigkdd international conference on knowledge discovery and data mining (2016) pp. 785–794.

[45] N. J. Nagelkerke et al., A note on a general definition of the coefficient of determination, biometrika 78, 691 (1991).

[46] H. Zhang, D. Nettleton, and Z. Zhu, Regression-enhanced random forests, arXiv preprint arXiv:1904.10416 (2019).

[47] A. Altmann, L. Toloşi, O. Sander, and T. Lengauer, Permutation importance: a corrected feature importance measure, Bioinformatics 26, 1340 (2010).

[48] M. S. Sandhu, D. M. Waterworth, S. L. Debenham, E. Wheeler, K. Papadakis, J. H. Zhao, K. Song, X. Yuan, T. Johnson, S. Ashford, M. Inouye, R. Luben, M. Sims, D. Hadley, W. McArdle, P. Barter, Y. A. Kesaniemi, R. W. Mahley, R. McPherson, S. M. Grundy, S. A. Bingham, K.-T. Khaw, R. J. Loos, G. Waeber, I. Barroso, D. P. Strachan, P. Deloukas, P. Vollenweider, N. J. Wareham, and V. Mooser, Ldl-cholesterol concentrations: a genome-wide association study, The Lancet 371, 483 (2008).

[49] Y. J. Kim, M. J. Go, C. Hu, C. B. Hong, Y. K. Kim, J. Y. Lee, J.-Y. Hwang, J. H. Oh, D.-J. Kim, N. H. Kim, et al., Large-scale genome-wide association studies in east asians identify new genetic loci influencing metabolic traits, Nature genetics 43, 990 (2011).

[50] T. E. Fingerlin, E. Murphy, W. Zhang, A. L. Peljto, K. K. Brown, M. P. Steele, J. E. Loyd, G. P. Cosgrove, D. Lynch, S. Groshong, et al., Genome-wide association study identifies multiple susceptibility loci for pulmonary fibrosis, Nature genetics 45, 613 (2013).

[51] R. Leslie, C. J. O’Donnell, and A. D. Johnson, GRASP: analysis of genotype–phenotype results from 1390 genome-wide association studies and corresponding open access database, Bioinformatics 30, i185 (2014).

[52] C. Cao, M. Shao, C. Zuo, D. Kwok, L. Liu, Y. Ge, Z. Zhang, F. Cui, M. Chen, R. Fan, et al., Ravar: a curated repository for rare variant–trait associations, Nucleic Acids Research 52, D990 (2024).

[53] M. C. Wu, S. Lee, T. Cai, Y. Li, M. Boehnke, and X. Lin, Rare-variant association testing for sequencing data with the sequence kernel association test, The American Journal of Human Genetics 89, 82 (2011).

[54] B. Li and S. M. Leal, Methods for detecting associations with rare variants for common diseases: application to analysis of sequence data, The American Journal of Human Genetics 83, 311 (2008).

[55] G. Consortium, K. G. Ardlie, D. S. Deluca, A. V. Segrè, T. J. Sullivan, T. R. Young, E. T. Gelfand, C. A. Trowbridge, J. B. Maller, T. Tukiainen, et al., The genotype-tissue expression (gtex) pilot analysis: multitissue gene regulation in humans, Science 348, 648 (2015).

[56] U. Võsa, A. Claringbould, H.-J. Westra, M. J. Bonder, P. Deelen, B. Zeng, H. Kirsten, A. Saha, R. Kreuzhuber, S. Yazar, et al., Large-scale cis-and trans-eqtl analyses identify thousands of genetic loci and polygenic scores that regulate blood gene expression, Nature genetics 53, 1300 (2021).

[57] K. Watanabe, E. Taskesen, A. Bochoven, and D. Posthuma, Functional mapping and annotation of genetic associations with fuma, Nature Communications 8 (2017).

[58] S. Devaraj, J. R. Semaan, and I. Jialal, enBiochemistry, apolipoprotein B, in enStatPearls (StatPearls Publishing, Treasure Island (FL), 2024).

[59] J. Behbodikhah, S. Ahmed, A. Elyasi, L. J. Kasselman, J. De Leon, A. D. Glass, and A. B. Reiss, Apolipoprotein b and cardiovascular disease: biomarker and potential therapeutic target, Metabolites 11, 690 (2021).

[60] S. Robichaud, G. Fairman, V. Vijithakumar, E. Mak, D. P. Cook, A. R. Pelletier, S. Huard, B. C. Vanderhyden, D. Figeys, M. Lavallée-Adam, et al., Identification of novel lipid droplet factors that regulate lipophagy and cholesterol efflux in macrophage foam cells, Autophagy 17, 3671 (2021).

[61] Z. S. Liang, I. Cimino, B. Yalcin, N. Raghupathy, V. E. Vancollie, X. Ibarra-Soria, H. V. Firth, D. Rimmington, I. S. Farooqi, C. J. Lelliott, S. C. Munger, S. O’Rahilly, A. C. Ferguson-Smith, A. P. Coll, and D. W. Logan, Trappc9 deficiency causes parent-of-origin dependent microcephaly and obesity, PLOS Genetics 16, 1 (2020).

[62] L. O. Bryzgalov, E. V. Antontseva, M. Y. Matveeva, A. G. Shilov, E. V. Kashina, V. A. Mordvinov, and T. I. Merkulova, Detection of regulatory snps in human genome using chip-seq encode data, PLoS one 8, e78833 (2013).

[63] A. Mathelier, X. Zhao, A. W. Zhang, F. Parcy, R. Worsley-Hunt, D. J. Arenillas, S. Buchman, C.-y. Chen, A. Chou, H. Ienasescu, J. Lim, C. Shyr, G. Tan, M. Zhou, B. Lenhard, A. Sandelin, and W. W. Wasserman, JASPAR 2014: an extensively expanded and updated open-access database of transcription factor binding profiles, Nucleic Acids Research 42, D142 (2013).

[64] S. Kumar, G. Ambrosini, and P. Bucher, SNP2TFBS – a database of regulatory SNPs affecting predicted transcription factor binding site affinity, Nucleic Acids Research 45, D139 (2016).

[65] J. V. Patankar, P. G. Chandak, S. Obrowsky, T. Pfeifer, C. Diwoky, A. Uellen, W. Sattler, R. Stollberger, G. Hoefler, A. Heinemann, et al., Loss of intestinal gata4 prevents diet-induced obesity and promotes insulin sensitivity in mice, American Journal of Physiology-Endocrinology and Metabolism 300, E478 (2011).

[66] L. Bideyan, M. L. Rodríguez, C. Priest, J. P. Kennelly, Y. Gao, A. Ferrari, P. Rajbhandari, A.-C. Feng, S. G. Tevosian, S. T. Smale, et al., Hepatic gata4 regulates cholesterol and triglyceride homeostasis in collaboration with lxrs, Genes & Development 36, 1129 (2022).

[67] C. Yin, A. M. Vrieze, M. Rosoga, J. Akingbasote, E. N. Pawlak, R. A. Jacob, J. Hu, N. Sharma, J. D. Dikeakos, L. Barra, et al., Efferocytic defects in early atherosclerosis are driven by gata2 overexpression in macrophages, Frontiers in immunology 11, 594136 (2020).

[68] A. Kohlmeier, C. A. M. Sison, B. D. Yilmaz, J. S. Coon V M. T. Dyson, and S. E. Bulun, Gata2 and progesterone receptor interaction in endometrial stromal cells undergoing decidualization, Endocrinology 161, bqaa070 (2020).

[69] Y. Choi, H. Jeon, J. W. Akin, T. E. Curry Jr, and M. Jo, The fos/ap-1 regulates metabolic changes and cholesterol synthesis in human periovulatory granulosa cells, Endocrinology 162, bqab127 (2021).

[70] J. Lv, S. Meng, Q. Gu, R. Zheng, X. Gao, J.-d. Kim, M. Chen, B. Xia, Y. Zuo, S. Zhu, et al., Epigenetic landscape reveals mecom as an endothelial lineage regulator, Nature communications 14, 2390 (2023).

[71] S. Safe, U.-H. Jin, B. Morpurgo, A. Abudayyeh, M. Singh, and R. B. Tjalkens, Nuclear receptor 4a (nr4a) family–orphans no more, The Journal of steroid biochemistry and molecular biology 157, 48 (2016).

[72] I. Kardys, C. M. van Tiel, C. J. de Vries, H. Pannekoek, A. G. Uitterlinden, A. Hofman, J. C. Witteman, and M. P. de Maat, Haplotypes of the nr4a2/nurr1 gene and cardiovascular disease: the rotterdam study, Human mutation 30, 417 (2009).

[73] L. D. Ward and M. Kellis, HaploReg: a resource for exploring chromatin states, conservation, and regulatory motif alterations within sets of genetically linked variants, Nucleic Acids Research 40, D930 (2011).

[74] U. Raudvere, L. Kolberg, I. Kuzmin, T. Arak, P. Adler, H. Peterson, and J. Vilo, g:Profiler: a web server for functional enrichment analysis and conversions of gene lists (2019 update), Nucleic Acids Research 47, W191 (2019).

[75] B. Klop, J. W. F. Elte, and M. Castro Cabezas, Dyslipidemia in obesity: mechanisms and potential targets, Nutrients 5, 1218 (2013).

[76] E. J. Kim, J. Hong, and J.-W. Hwang, The association between depressive mood and cholesterol levels in korean adolescents, Psychiatry investigation 16, 737 (2019).

[77] E. Trojan, J. Chwastek, A. Basta-Kaim, et al., A potential contribution of chemokine network dysfunction to the depressive disorders, Current neuropharmacology 14, 705 (2016).

[78] J. M. Chinsky and R. D. Steiner, Chapter 30 - inborn errors of metabolism, in Developmental-Behavioral Pediatrics (Fourth Edition), edited by W. B. Carey, A. C. Crocker, W. L. Coleman, E. R. Elias, and H. M. Feldman (W.B. Saunders, Philadelphia, 2009) fourth edition ed., pp. 287–313.

[79] F. D. Porter et al., Malformation syndromes due to inborn errors of cholesterol synthesis, The Journal of clinical investigation 110, 715 (2002).

[80] H.-J. Lee, S.-R. Lee, E.-K. Choi, K.-D. Han, and S. Oh, Low lipid levels and high variability are associated with the risk of new-onset atrial fibrillation, Journal of the American Heart Association 8, e012771 (2019).

[81] S. Visinoni, N. F. I. Khalid, C. N. Joannides, A. Shulkes, M. Yim, J. Whitehead, T. Tiganis, B. J. Lamont, J. M. Favaloro, J. Proietto, et al., The role of liver fructose-1, 6-bisphosphatase in regulating appetite and adiposity, Diabetes 61, 1122 (2012).

[82] H. Y. Ali Moussa, K. C. Shin, J. Ponraj, S. J. Kim, J.-K. Ryu, S. Mansour, and Y. Park, Requirement of cholesterol for calcium-dependent vesicle fusion by strengthening synaptotagmin-1-induced membrane bending, Advanced Science 10, 2206823 (2023).

[83] L. N. R. Thota and A. Z. Chignalia, The role of the glypican and syndecan families of heparan sulfate proteoglycans in cardiovascular function and disease, American Journal of Physiology-Cell Physiology 323, C1052 (2022).

[84] X. Zhou, Z. Yin, X. Guo, D. P. Hajjar, and J. Han, Inhibition of erk1/2 and activation of liver x receptor synergistically induce macrophage abca1 expression and cholesterol efflux, Journal of biological chemistry 285, 6316 (2010).

[85] C. Gustafsen, D. Olsen, J. Vilstrup, S. Lund, A. Reinhardt, N. Wellner, T. Larsen, C. B. Andersen, K. Weyer, J.-p. Li, et al., Heparan sulfate proteoglycans present pcsk9 to the ldl receptor, Nature communications 8, 503 (2017).

[86] M. Canuel, X. Sun, M.-C. Asselin, E. Paramithiotis, A. Prat, and N. G. Seidah, Proprotein convertase subtilisin/kexin type 9 (pcsk9) can mediate degradation of the low density lipoprotein receptor-related protein 1 (lrp-1), PloS one 8, e64145 (2013).

[87] J. K. Nelson, D. S. Koenis, S. Scheij, E. C. L. Cook, M. Moeton, A. Santos, J.-M. A. Lobaccaro, S. Baron, and N. Zelcer, Eepd1 is a novel lxr target gene in macrophages which regulates abca1 abundance and cholesterol efflux, Arteriosclerosis, Thrombosis, and Vascular Biology 37, 423 (2017).

[88] X.-D. Xia, X.-H. Yu, L.-Y. Chen, S.-l. Xie, Y.-G. Feng, R.-Z. Yang, Z.-W. Zhao, H. Li, G. Wang, and C.-K. Tang, Myocardin suppression increases lipid retention and atherosclerosis via downregulation of abca1 in vascular smooth muscle cells, Biochimica et Biophysica Acta (BBA)-Molecular and Cell Biology of Lipids 1866, 158824 (2021).

[89] K. Kunimura, T. Uruno, and Y. Fukui, DOCK family proteins: key players in immune surveillance mechanisms, International Immunology 32, 5 (2019).

[90] C. Casalou, A. Costa, T. Carvalho, A. L. Gomes, Z. Zhu, Y. Wu, and S. Dias, Cholesterol regulates vegfr-1 (flt-1) expression and signaling in acute leukemia cells, Molecular Cancer Research 9, 215 (2011).

[91] S. M. Pokharel, N. K. Shil, J. B. Gc, Z. T. Colburn, S.-Y. Tsai, J. A. Segovia, T.-H. Chang, S. Bandyopadhyay, S. Natesan, J. C. Jones, et al., Integrin activation by the lipid molecule 25-hydroxycholesterol induces a proinflammatory response, Nature communications 10, 1482 (2019).

[92] W. Wang, X. Zhang, Q. Gao, M. Lawas, L. Yu, X. Cheng, M. Gu, N. Sahoo, X. Li, P. Li, et al., A voltage-dependent k+ channel in the lysosome is required for refilling lysosomal ca2+ stores, Journal of Cell Biology 216, 1715 (2017).

[93] Z. Li, B. Zhang, Q. Liu, Z. Tao, L. Ding, B. Guo, E. Zhang, H. Zhang, Z. Meng, S. Guo, Y. Chen, J. Peng, J. Li, C. Wang, Y. Huang, H. Xu, and Y. Wu, Genetic association of lipids and lipid-lowering drug target genes with non-alcoholic fatty liver disease, eBioMedicine 90, 104543 (2023).

[94] M. K. Kwok, S. L. Lin, and C. M. Schooling, Re-thinking alzheimer’s disease therapeutic targets using gene-based tests, EBioMedicine 37, 461 (2018).

[95] C.-F. Yang, Y.-Y. Chen, J. P. Singh, S.-F. Hsu, Y.-W. Liu, C.-Y. Yang, C.-W. Chang, S.-N. Chen, R.-H. Shih, S.-T. D. Hsu, Y.-S. Jou, C.-F. Cheng, and T.-C. Meng, Targeting protein tyrosine phosphatase PTP-PEST (PTPN12) for therapeutic intervention in acute myocardial infarction, Cardiovascular Research 116, 1032 (2019).

[96] G. Li, Y. Xu, X. Hu, N. Li, R. Yao, T. Yu, X. Wang, W. Guo, and J. Wang, Compound heterozygous variants of the cog6 gene in a chinese patient with deficiency of subunit 6 of the conserved oligomeric golgi complex (cog6-cdg), European Journal of Medical Genetics 62, 44 (2019).

[97] L. Nitschke, A. Tewari, S. L. Coffin, E. Xhako, K. Pang, V. A. Gennarino, J. L. Johnson, F. A. Blanco, Z. Liu, and H. Y. Zoghbi, mir760 regulates atxn1 levels via interaction with its 5’ untranslated region, Genes & development 34, 1147 (2020).

