## Supplementary Document for "Enhancing genotype-phenotype association with optimized machine learning and biological enrichment methods"

**Details of PennCATH dataset:**
The phenotypic and SNP data used in this study came from the PennCATH cohort, a Genome-wide association study (GWAS) led by the University of Pennsylvania Medical Center that examined cardiovascular risk factors and coronary artery disease (CAD). A total of 3850 individuals having cardiac catheterization were recruited for the PennCATH trial. Individuals who were fasting provided blood samples, from which plasma and DNA were extracted. The blood samples were analyzed further, and the amounts of lipoprotein and glucose were determined. Clinical information was recorded, including gender, age, triglyceride levels, cholesterol levels of high-density lipoprotein (HDL), low-density lipoprotein (LDL), and the presence of CAD.

**Random forest with multiple parameters:**
For our study, we utilized random forest (RF) as an association technique and elastic-net as a method for feature selection. The hyperparameters of the RF model were meticulously fine-tuned to identify the optimal configuration for our dataset. The tuning process included using a parameter grid that contained the following values: $n\_estimators$ [100, 200, 300, 400, 500], $max\_depth$ [None, 2, 5, 10], $min\_samples\_split$ [2, 5, 10], and $min\_samples\_leaf$ [1, 2, 4]. The hyperparameter optimization process resulted in the identification of the optimal parameters as follows: $max\_depth$: None, $min\_samples\_leaf$: 4, $min\_samples\_split$: 2, $n\_estimators$: 400. The model's performance was assessed on the test set for other parameters, resulting in the following outcomes: The MSE is 999.62, RMSE is 31.61, MAE is 25.73, $R^2$ score is -70.26.

**XGBoost results on PennCATH dataset:**
We utilized XGBoost as an approach for identifying associations and elastic-net as a tool for selecting features. The hyperparameters of XGBoost were meticulously tuned to get the optimal configuration for our dataset. The evaluated parameters were $n\_estimators$ with the values [100, 200, 300], $learning\_rate$ with the values [0.01, 0.1, 0.2], and $max\_depth$ with the values [3, 5, 7]. After doing hyperparameter optimization, the model that had the highest performance was selected. It had the following parameter values: $learning\_rate$ = 0.1, $max\_depth$ = 3, $n\_estimators$= 300. The model produced a $R^2$ score of -6.79, a root mean squared error (RMSE) of 31.90, a mean absolute error (MAE) of 25.71, and a mean squared error (MSE) of 1018.23 on another parameter.

**Permutation importance score:**
We evaluated the significance of 5000 SNPs from the elastic-net feature selection and the top 100 SNPs, highlighting their impact on model performance (Figure: S1 (A) 5000 SNPs, (B) Top 100 SNPs).

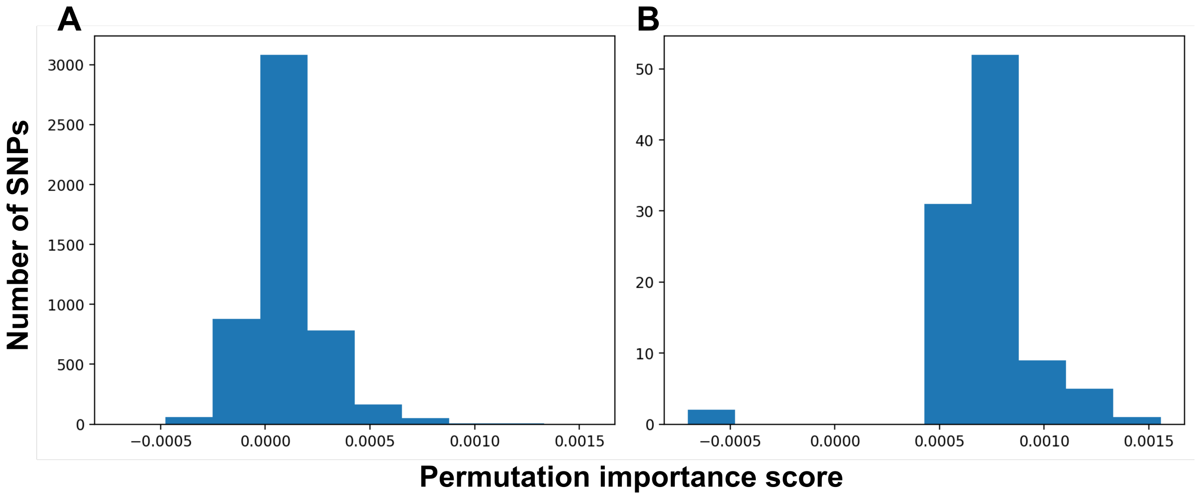

Figure S1: Permutation importance score of (A) 5000 SNPs selected from elastic-net feature selection method and (B) top 100 SNPs.

**eQTL analysis:**

Detailed results of eQTL analysis by GTEx are given in the below Table S1. The comprehensive results of associated SNPs documented in the literature are given in Table S2.

Table S1: Expression of identified top 100 SNPs in various tissues using GTEx

| SNPs | p-value | Gene | GTEx tissue |
|---|---|---|---|
| rs6778643 | $1.60 \times 10^{-44}$ | ITGB5 | Artery - Tibial |
| rs188384 | $9.30 \times 10^{-29}$ | CCDC50 | Artery - Tibial |
| rs2020009 | $1.50 \times 10^{-25}$ | RP11-73G16.3 | Testis |
| rs11642880 | $2.60 \times 10^{-22}$ | SNN | Esophagus - Muscularis |
| rs12438724 | $8.60 \times 10^{-21}$ | CSPG4P12 | Muscle - Skeletal |
| rs9567431 | $4.90 \times 10^{-20}$ | TUSC8 | Prostate |
| rs11784756 | $1.90 \times 10^{-19}$ | SUPT4H1 | Whole Blood |
| rs4516649 | $5.30 \times 10^{-19}$ | CPNE4 | Cells - Cultured fibroblasts |
| rs1240259 | $3.40 \times 10^{-18}$ | RAB3IP | Whole Blood |
| rs1453012 | $1.26 \times 10^{-16}$ | TRIM36 | Whole Blood |
| rs660586 | $5.90 \times 10^{-16}$ | ATXN1 | Whole Blood |
| rs41365345 | $3.50 \times 10^{-14}$ | GTF2E1 | Muscle - Skeletal |
| rs17559708 | $2.16 \times 10^{-13}$ | CTB-37A13.1 | Whole Blood |
| rs29795 | $4.30 \times 10^{-13}$ | BNIP1 | Adipose - Subcutaneous |
| rs10464982 | $2.85 \times 10^{-12}$ | TRAPPC9 | Whole Blood |
| rs9301479 | $3.39 \times 10^{-12}$ | CARKD | Whole Blood |

| | | | |
|---|---|---|---|
| rs10890879 | $8.20 \times 10^{-12}$ | DDX10 | Testis |
| rs4591370 | $1.80 \times 10^{-11}$ | APOB | Heart - Left Ventricle |
| rs3017499 | $7.70 \times 10^{-11}$ | SHANK2 | Adipose - Visceral (Omentum) |
| rs2029818 | $1.21 \times 10^{-10}$ | HEY1 | Whole Blood |
| rs6903827 | $2.20 \times 10^{-10}$ | TAB2 | Cells - Cultured fibroblasts |
| rs3768480 | $4.40 \times 10^{-10}$ | STRIP1 | Whole Blood |
| rs17127435 | $5.42 \times 10^{-10}$ | GRAMD1B | Whole Blood |
| rs4917774 | $2.20 \times 10^{-9}$ | SLIT1 | Heart - Atrial Appendage |
| rs2726050 | $2.60 \times 10^{-9}$ | MARK2P13 | Skin - Not Sun Exposed (Suprapubic) |
| rs17046334 | $5.57 \times 10^{-9}$ | SYT1 | Whole Blood |
| rs1896384 | $4.40 \times 10^{-8}$ | CPXM2 | Artery - Aorta |
| rs1952183 | $6.30 \times 10^{-8}$ | FOXN3 | Heart - Left Ventricle |
| rs10802219 | $8.73 \times 10^{-8}$ | KIF26B | Whole Blood |
| rs272709 | $9.10 \times 10^{-8}$ | DFNA5 | Whole Blood |
| rs9319501 | $1.03 \times 10^{-7}$ | MON1B | Whole Blood |
| rs6912831 | $1.90 \times 10^{-7}$ | TXLNB | Adipose - Subcutaneous |
| rs454266 | $2.20 \times 10^{-7}$ | EMX2OS | Artery - Tibial |
| rs324884 | $4.90 \times 10^{-7}$ | TMEM161B-AS1 | Whole Blood |
| rs299289 | $5.20 \times 10^{-7}$ | NUDCD2 | Artery - Tibial |
| rs1013192 | $6.50 \times 10^{-7}$ | CTB-37A13.1 | Whole Blood |
| rs7180563 | $6.60 \times 10^{-7}$ | FAM189A1 | Minor Salivary Gland |
| rs11255291 | $6.80 \times 10^{-7}$ | ITIH5 | Cells - EBV-transformed lymphocytes |
| rs636926 | $7.90 \times 10^{-7}$ | FCHO2 | Artery - Aorta |
| rs4732439 | $9.80 \times 10^{-7}$ | CCL26 | Heart - Atrial Appendage |
| rs10058089 | $1.13 \times 10^{-6}$ | STARD4-AS1 | Whole Blood |
| rs7232775 | $1.43 \times 10^{-6}$ | SLC14A1 | Whole Blood |
| rs10940058 | $1.70 \times 10^{-6}$ | RP11-305P14.1 | Testis |
| rs12680681 | $1.80 \times 10^{-6}$ | RAB2A | Artery - Tibial |
| rs17776811 | $4.30 \times 10^{-6}$ | FOXN3 | Pancreas |
| rs2059843 | $8.30 \times 10^{-6}$ | LINC02142 | Testis |
| rs2201409 | $1.40 \times 10^{-5}$ | SPEF2 | Artery - Tibial |
| rs17366040 | $1.40 \times 10^{-5}$ | RP11-521M14.1 | Adipose - Subcutaneous |
| rs810083 | $2.80 \times 10^{-5}$ | ZMIZ1 | Thyroid |

| | | | |
|---|---|---|---|
| rs9298320 | $3.20 \times 10^{-5}$ | RP11-578O24.2 | Esophagus - Gastroesophageal Junction |
| rs848481 | $4.40 \times 10^{-5}$ | PTPN12 | Artery - Tibial |
| rs17655474 | $4.40 \times 10^{-5}$ | DNAH17-AS1 | Testis |
| rs1192439 | $4.80 \times 10^{-5}$ | RP1-240B8.3 | Brain - Cortex |
| rs12499915 | $5.20 \times 10^{-5}$ | TIFA | Heart - Left Ventricle |
| rs10892984 | $5.30 \times 10^{-5}$ | GRAMD1B | Brain - Cerebellar Hemisphere |
| rs1501142 | $7.30 \times 10^{-5}$ | CD38 | Esophagus - Mucosa |
| rs9695 | $7.80 \times 10^{-5}$ | MFSD14B | Whole Blood |
| rs9576907 | $8.90 \times 10^{-5}$ | COG6 | Muscle - Skeletal |
| rs10230715 | $9.30 \times 10^{-5}$ | C7orf25 | Muscle - Skeletal |
| rs3924222 | $1.20 \times 10^{-4}$ | FUT8 | Nerve - Tibial |
| rs1105460 | $1.40 \times 10^{-4}$ | FHOD3 | Skin - Not Sun Exposed (Suprapubic) |
| rs11530859 | $1.70 \times 10^{-4}$ | ANKRD42 | Thyroid |
| rs12494729 | $1.80 \times 10^{-4}$ | PRICKLE2 | Thyroid |
| rs6916224 | $2.50 \times 10^{-4}$ | TBX18-AS1 | Artery - Tibial |

Table S2: GRASP table for associated SNPs documented in the literature

| Snp Id | p-value | Phenotype | chr | Functional Class |
|---|---|---|---|---|
| rs4591370 | $7.80 \times 10^{-82}$ | LDL cholesterol | 2 | downstream, upstream |
| rs4591370 | $2.00 \times 10^{-68}$ | Total cholesterol | 2 | downstream, upstream |
| rs4591370 | $1.30 \times 10^{-19}$ | LDL cholesterol | 2 | downstream, upstream |
| rs7232775 | $8.00 \times 10^{-14}$ | Blood urea nitrogen (BUN) | 18 | intron |
| rs4591370 | $8.20 \times 10^{-9}$ | LDL cholesterol in serum | 2 | downstream, upstream |
| rs4591370 | $2.20 \times 10^{-8}$ | Coronary artery disease (CAD) | 2 | downstream, upstream |
| rs4591370 | $2.90 \times 10^{-8}$ | Total cholesterol | 2 | downstream, upstream |
| rs12438724 | $4.10 \times 10^{-8}$ | Pulmonary fibrosis | 15 | downstream, upstream |
| rs11255291 | $1.50 \times 10^{-6}$ | Follicle stimulating hormone (FSH) | 10 | upstream, downstream |
| rs17776811 | $7.40 \times 10^{-6}$ | Circulating sex hormone-binding globulin levels | 14 | intron, intron |
| rs4591370 | $8.70 \times 10^{-6}$ | LDL cholesterol | 2 | downstream, upstream |
| rs11675322 | $1.90 \times 10^{-5}$ | Percent mammographic density | 2 | downstream, upstream |
| rs4591370 | $2.00 \times 10^{-5}$ | LDL cholesterol change with statins | 2 | downstream, upstream |
| rs4591370 | $2.00 \times 10^{-5}$ | LDL cholesterol | 2 | downstream, upstream |
| rs272709 | $2.20 \times 10^{-5}$ | Extraversion | 7 | downstream, upstream |
| rs848481 | $2.50 \times 10^{-5}$ | Serum creatinine | 7 | intron |
| rs1896384 | $3.60 \times 10^{-5}$ | Blood pressure response to candesartan treatment | 10 | downstream, upstream |
| rs7180563 | $4.00 \times 10^{-5}$ | Alzheimers disease (APOE4/E3 heterozygotes) | 15 | downstream, upstream |
| rs10733392 | $4.20 \times 10^{-5}$ | Parkinsons disease (PD) (age of onset) | 9 | intron |
| rs10230715 | $4.40 \times 10^{-5}$ | Coronary artery disease (CAD) | 7 | intron |
| rs4591370 | $6.90 \times 10^{-5}$ | Lp-PLA2 mass | 2 | downstream, upstream |
| rs928641 | $8.30 \times 10^{-5}$ | Spatial span | 9 | intron |
| rs2238090 | $8.60 \times 10^{-5}$ | Schizophrenia | 12 | intron |

Table S3: Beagle5.4 imputed rare variants identified in RAVAR database

| Gene Symbol | Trait Label | Method Software |
|---|---|---|
| ABCC3 | bilirubin measurement | BOLT-LMM |
| ARHGAP26 | anterior horn disorder | collapsing analyse |
| ATP9B | chronic cystitis | collapsing analyse |
| BDH1 | acalculous cholecystitis | collapsing analyse |
| CCDC50 | cesarean section | collapsing analyse |
| CDH13 | spondyloarthropathy | collapsing analyse |
| CLDN10 | urate measurement | BOLT-LMM |
| DGKB | urate measurement | Burden test |
| EEPD1 | Low back pain | collapsing analyse |
| FADS2 | QT interval | SKAT |
| FGF14 | encephalomyelitis | collapsing analyse |
| FYB1 | phenyl alanyl tryptophan measurement | SKAT |
| GPR161 | cardiomyopathy | collapsing analyse |
| HHIPL2 | HbA1c measurement | Burden test |
| IL27RA | Iron deficiency anemia | collapsing analyse |
| KCNN3 | Cholecystitis | collapsing analyse |
| LITAF | QT interval | SKAT |
| NRXN3 | 3-methoxytyrosine measurement | SKAT |
| NTN1 | systolic blood pressure | SKAT |
| NXN | fecal incontinence | collapsing analyse |
| PCDH15 | osteoarthritis | collapsing analyse |
| PDE1C | dental caries | collapsing analyse |
| PIK3C2G | otitis externa | collapsing analyse |
| PPFIA4 | systolic blood pressure | SKAT |
| RBMS2 | isoleucine | collapsing analyse |
| RELN | vitamin D measurement | SKAT |
| SARDH | hearing loss | collapsing analyse |
| SHANK2 | autism spectrum disorder | TADA model |
| SLC12A8 | streptococcal pneumonia | collapsing analyse |
| TMEM156 | gastritis | collapsing analyse |
| TMEM171 | serum albumin | collapsing analyse |
| TOX | Inguinal hernia | collapsing analyse |
| UGT1A8 | bilirubin measurement | Burden test |
| UNC13C | endometriosis | collapsing analyse |

Table S4: PennCATH rare variants identified in RAVAR database

| Gene Symbol | Trait Label | Method Software |
|---|---|---|
| ACO2 | systolic blood pressure | SKAT |
| ANK2 | spermatocele | collapsing analyse |
| ARMC2 | Joint laxity | collapsing analyse |
| BCL2 | systolic blood pressure | SKAT |
| BMPER | abnormal result of function studies | collapsing analyse |
| CACNA1S | creatinine | collapsing analyse |
| CACNA2D1 | block | collapsing analyse |
| CAPN14 | Appendix Adenocarcinoma | collapsing analyse |
| CCDC141 | hypertrophic cardiomyopathy | collapsing analyse |
| CEACAM7 | Umbilical hernia | collapsing analyse |
| CSMD1 | gastritis | collapsing analyse |
| DDX1 | gout | collapsing analyse |
| DLG5 | protein-glutamine gamma-glutamyltransferase e measurement | collapsing analyse |
| EXTL3 | body height | Burden test |
| FBXO39 | citrate measurement | collapsing analyse |
| FOXN3 | aneurysm | collapsing analyse |
| FUT9 | Maternal diabetes | collapsing analyse |
| GRIN2B | schizophrenia | Burden test |
| HIPK2 | gangrene | collapsing analyse |
| KCNQ5 | anxiety disorder | collapsing analyse |
| LRP4 | heel bone mineral density | BOLT-LMM |
| MGST2 | femoral hernia | collapsing analyse |
| NCOA1 | Premature rupture of membranes | collapsing analyse |
| NFIB | 3-methylglutaconate measurement | SKAT |
| NRG3 | skin disease | collapsing analyse |
| PIP4K2A | Sleep Disorder | collapsing analyse |
| PTPRD | cirrhosis of liver | collapsing analyse |
| RARB | respiratory system disease | collapsing analyse |
| SLC14A2 | urea | collapsing analyse |
| SLC44A1 | Strabismus | collapsing analyse |
| SNTB1 | malignant renal pelvis neoplasm | collapsing analyse |
| SNX29 | obsessive-compulsive disorder | collapsing analyse |
| SVOP | Monteggia’s fracture | Burden test |
| TP63 | respiratory system disease | collapsing analyse |
| TSPAN12 | fibromyalgia | collapsing analyse |
| XYLT1 | medical procedure | collapsing analyse |
| ZFPM2 | fasting blood glucose measurement | collapsing analyse |

Table S5: Impute5 rare variants identified in RAVAR database

| Gene Symbol | Trait Label | Method Software |
|---|---|---|
| ACO2 | systolic blood pressure | SKAT |
| ANK2 | spermatocele | collapsing analyse |
| ARMC2 | Joint laxity | collapsing analyse |
| BCL2 | systolic blood pressure | SKAT |
| BMPER | abnormal result of function studies | collapsing analyse |
| CACNA1S | creatinine | collapsing analyse |
| CACNA2D1 | block | collapsing analyse |
| CAPN14 | Appendix Adenocarcinoma | collapsing analyse |
| CCDC141 | hypertrophic cardiomyopathy | collapsing analyse |
| CD28 | skin and soft tissue Staphylococcus aureus infection | collapsing analyse |
| CEACAM7 | Umbilical hernia | collapsing analyse |
| CSMD1 | gastritis | collapsing analyse |
| DDX1 | gout | collapsing analyse |
| DLG5 | protein-glutamine gamma-glutamyltransferase e measurement | collapsing analyse |
| EHHADH | very low density lipoprotein cholesterol measurement | collapsing analyse |
| EXTL3 | body height | Burden test |
| FBXO39 | citrate measurement | collapsing analyse |
| FOXN3 | aneurysm | collapsing analyse |
| GRIN2B | schizophrenia | Burden test |
| HIP1 | skin neoplasm | collapsing analyse |
| HIPK2 | gangrene | collapsing analyse |
| KALRN | mean platelet volume | BOLT-LMM |
| KCNQ5 | anxiety disorder | collapsing analyse |
| LRP4 | heel bone mineral density | BOLT-LMM |
| MGST2 | femoral hernia | collapsing analyse |
| MTUS1 | 2-aminoadipate measurement | Burden test |
| NCOA1 | Premature rupture of membranes | collapsing analyse |
| NFIB | 3-methylglutaconate measurement | SKAT |
| NRG3 | skin disease | collapsing analyse |
| PIP4K2A | Sleep Disorder | collapsing analyse |
| PTPRD | cirrhosis of liver | collapsing analyse |

| | | |
|---|---|---|
| RARB | respiratory system disease | collapsing analyse |
| SLC14A2 | urea | collapsing analyse |
| SLC44A1 | Strabismus | collapsing analyse |
| SNTB1 | malignant renal pelvis neoplasm | collapsing analyse |
| SNX29 | obsessive-compulsive disorder | collapsing analyse |
| SVOP | Monteggia's fracture | Burden test |
| TP63 | respiratory system disease | collapsing analyse |
| TSPAN12 | fibromyalgia | collapsing analyse |
| UST | abnormal result of diagnostic imaging | collapsing analyse |
| XYLT1 | medical procedure | collapsing analyse |
| ZFPM2 | fasting blood glucose measurement | collapsing analyse |

The detailed results of regulatory region enrichment analysis using HaploReg v4.2 are displayed in Table S6.

**Table S6**: Regulatory region enrichment analysis of top 100 SNPs identified from PennCATH-real dataset using our proposed pipeline

### HaploReg v4.2

HaploReg is a tool for exploring annotations of the noncoding genome at variants on haplotype blocks, such as candidate regulatory SNPs at disease-associated loci. Using LD information from the 1000 Genomes Project, linked SNPs and small indels can be visualized along with chromatin state and protein binding annotation from the Roadmap Epigenomics and ENCODE projects, sequence conservation across mammals, the effect of SNPs on regulatory motifs, and the effect of SNPs on expression from eQTL studies. HaploReg is designed for researchers developing mechanistic hypotheses of the impact of non-coding variants on clinical phenotypes and normal variation.

**Update 2015.11.05: Version 4.1** GWAS and eQTL have been updated; a simpler pruning strategy is applied when combining GWAS; and links out to other NHGRI/EBI GWAS hits and GRASP QTL hits are provided.

**Update 2015.09.15:** Version 4.0 now includes many recent eQTL results including the GTEx pilot, four different options for defining enhancers using Roadmap Epigenomics data, and a complete set of source files for download and local analysis. Older versions available: v3, v2, v1.

**Update 2023.04.29:** Version 4.2 released. Parts of the site may still not work correctly - please email haploreg (at) mit.edu for support.

- Build Query
- Set Options
- Documentation

#### Build Query

Use one of the three methods below to enter a set of variants. If an r² threshold is specified (see the Set Options tab), results for each variant will be shown in a separate table along with other variants in LD. If r² is set to NA, only queried variants will be shown, together in one table.

Query (comma-delimited list of rsIDs OR a single region as chrN:start-end):

or, upload a text file (one refSNP ID per line): Browse… No file selected.

or, select a GWAS:

#### Set Options

LD threshold, r² (select NA to only show query variants): 0.8

1000G Phase 1 population for LD calculation: ○ AFR ◉ AMR ○ ASN ○ EUR

Source for epigenomes: ChromHMM (Core 15-state model)

Mammalian conservation algorithm: ○ GERP ◉ SiPhy-omega ○ both

Show position relative to: ◉ GENCODE genes ○ RefSeq genes ○ both

Condense lists in table longer than: 3

Condense indel oligos longer than: 6

Output mode: ◉ HTML ○ Text

#### Documentation

For details on data sources and methods along with usage examples, see the full documentation (opens in a pop-up window.)

The HaploReg database and web interface were produced by Luke Ward in the Kellis Lab at MIT. HaploReg is hosted by the Broad Institute.

To cite HaploReg, please refer to our publication in Nucleic Acids Research: HaploReg: a resource for exploring chromatin states, conservation, and regulatory motif alterations within sets of genetically linked variants. (PMID:22064851).

The underlying data are available in the following directory.

Contact:

Submit Query

Query SNP: rs3017499 and variants with $r^2$ >= 0.8

| chr | pos (hg38) | LD (r²) | LD (D') | variant | Ref | Alt | AFR freq | AMR freq | ASN freq | EUR freq | SiPhy cons | Promoter histone marks | Enhancer histone marks | DNAse | Proteins bound | Motifs changed | NHGRI/EBI GWAS hits | GRASP QTL hits | Selected eQTL hits | GENCODE genes | dbSNP func annot |
|---|---|---|---|---|---|---|---|---|---|---|---|---|---|---|---|---|---|---|---|---|---|
| 11 | 70832092 | 1 | 1 | rs2921332 | A | G | 0.55 | 0.70 | 0.77 | 0.69 | | | | | | | | | | SHANK2 | intronic |
| 11 | 70832103 | 1 | 1 | rs2921333 | A | G | 0.55 | 0.70 | 0.77 | 0.69 | | | | | | Myc | | | | SHANK2 | intronic |
| 11 | 70832335 | 1 | 1 | rs3017499 | C | T | 0.53 | 0.70 | 0.77 | 0.69 | | | | THYM | | Pax-5,Sox | | 1 hit | | SHANK2 | intronic |

Query SNP: rs950968 and variants with $r^2$ >= 0.8

| chr | pos (hg38) | LD (r²) | LD (D') | variant | Ref | Alt | AFR freq | AMR freq | ASN freq | EUR freq | SiPhy cons | Promoter histone marks | Enhancer histone marks | DNAse | Proteins bound | Motifs changed | NHGRI/EBI GWAS hits | GRASP QTL hits | Selected eQTL hits | GENCODE genes | dbSNP func annot |
|---|---|---|---|---|---|---|---|---|---|---|---|---|---|---|---|---|---|---|---|---|---|
| 2 | 8423196 | 1 | 1 | rs950968 | T | C | 0.79 | 0.61 | 0.46 | 0.56 | | | BLD | | | HDAC2,Znf143 | | | | 40kb 5' of LINC00299 | |
| 2 | 8423294 | 0.98 | 1 | rs950969 | C | G | 0.77 | 0.60 | 0.46 | 0.53 | | | BLD | | | 4 altered motifs | | | | 40kb 5' of LINC00299 | |

Query SNP: rs2136844 and variants with $r^2$ >= 0.8

| chr | pos (hg38) | LD (r²) | LD (D') | variant | Ref | Alt | AFR freq | AMR freq | ASN freq | EUR freq | SiPhy cons | Promoter histone marks | Enhancer histone marks | DNAse | Proteins bound | Motifs changed | NHGRI/EBI GWAS hits | GRASP QTL hits | Selected eQTL hits | GENCODE genes | dbSNP func annot |
|---|---|---|---|---|---|---|---|---|---|---|---|---|---|---|---|---|---|---|---|---|---|
| 4 | 4682915 | 1 | 1 | rs7694247 | G | A | 0.64 | 0.54 | 0.70 | 0.58 | | | ESDR, GI, LIV | | | | | | | 45kb 3' of RP11-323F5.2 | intronic |
| 4 | 4682922 | 1 | 1 | rs7670935 | C | G | 0.64 | 0.54 | 0.70 | 0.58 | | | ESDR, GI, LIV | | | 6 altered motifs | | | | 45kb 3' of RP11-323F5.2 | intronic |
| 4 | 4682988 | 1 | 1 | rs7685944 | T | G | 0.64 | 0.54 | 0.70 | 0.59 | | | ESDR, GI, LIV | | | p300 | | | 1 hit | 45kb 3' of RP11-323F5.2 | intronic |
| 4 | 4683163 | 1 | 1 | rs11726317 | A | G | 0.63 | 0.54 | 0.70 | 0.59 | | | ESDR, GI | | | BCL,Zfx | | | | 45kb 3' of RP11-323F5.2 | intronic |
| 4 | 4683218 | 0.98 | 1 | rs2887444 | G | A | 0.64 | 0.54 | 0.70 | 0.59 | | | ESDR, GI | | | HNF4,Nanog | | | | 45kb 3' of RP11-323F5.2 | intronic |

| | | | | | | | | | | | | | | | | | | | | | |
|---|---|---|---|---|---|---|---|---|---|---|---|---|---|---|---|---|---|---|---|---|---|
| 4 | 4683276 | 0.98 | 1 | rs201031512 | A | AC | 0.62 | 0.53 | 0.69 | 0.57 | | | GI | | | 22 altered motifs | | | | 45kb 3' of RP11-323F5.2 | intronic |
| 4 | 4683278 | 0.99 | 1 | rs113493162 | TA | T | 0.63 | 0.53 | 0.70 | 0.58 | | | GI | | | 20 altered motifs | | | | 45kb 3' of RP11-323F5.2 | intronic |
| 4 | 4683415 | 1 | 1 | rs2136844 | A | T | 0.64 | 0.54 | 0.70 | 0.59 | | | GI | | | HNF1 | | 1 hit | | 46kb 3' of RP11-323F5.2 | intronic |
| 4 | 4683468 | 1 | 1 | rs2136843 | A | G | 0.64 | 0.54 | 0.70 | 0.59 | | | GI | | | Irf | | 1 hit | | 46kb 3' of RP11-323F5.2 | intronic |
| 4 | 4683677 | 1 | 1 | rs2887445 | C | T | 0.64 | 0.54 | 0.70 | 0.59 | | | GI | | | BDP1,CTCF,RXRA | | | | 46kb 3' of RP11-323F5.2 | intronic |
| 4 | 4683730 | 1 | 1 | rs2887446 | A | G | 0.64 | 0.54 | 0.70 | 0.59 | | | GI | | | LBP-1 | | | | 46kb 3' of RP11-323F5.2 | intronic |
| 4 | 4683816 | 1 | 1 | rs2369288 | G | A | 0.64 | 0.54 | 0.70 | 0.58 | | | GI | | | Arid5a,RORalpha1 | | | | 46kb 3' of RP11-323F5.2 | intronic |
| 4 | 4683860 | 1 | 1 | rs2369289 | G | A | 0.64 | 0.54 | 0.70 | 0.58 | | | GI | | | Pou3f2,Pou6f1,TEF-1 | | 1 hit | | 46kb 3' of RP11-323F5.2 | intronic |
| 4 | 4684009 | 1 | 1 | rs10009731 | T | C | 0.64 | 0.54 | 0.70 | 0.58 | | | | | | 6 altered motifs | | 1 hit | | 46kb 3' of RP11-323F5.2 | intronic |
| 4 | 4684111 | 1 | 1 | rs9993619 | G | A | 0.64 | 0.54 | 0.70 | 0.58 | | | GI | | | MZF1::1-4,RREB-1 | | | | 46kb 3' of RP11-323F5.2 | intronic |
| 4 | 4684175 | 1 | 1 | rs9993700 | G | A | 0.64 | 0.54 | 0.70 | 0.59 | | | GI | | | 18 altered motifs | | | | 46kb 3' of RP11-323F5.2 | intronic |
| 4 | 4684178 | 1 | 1 | rs10007043 | A | G | 0.64 | 0.54 | 0.70 | 0.59 | | | | | | 7 altered motifs | | | | 46kb 3' of RP11-323F5.2 | intronic |
| 4 | 4684203 | 1 | 1 | rs9993711 | G | T | 0.63 | 0.54 | 0.70 | 0.59 | | | GI | | | 13 altered motifs | | | | 46kb 3' of RP11-323F5.2 | intronic |
| 4 | 4684367 | 1 | 1 | rs9993910 | G | A | 0.64 | 0.54 | 0.70 | 0.59 | | | | | | TCF12 | | | | 46kb 3' of RP11-323F5.2 | intronic |
| 4 | 4684386 | 0.99 | 1 | rs10010093 | T | A | 0.63 | 0.53 | 0.70 | 0.58 | | | | | | Ik-3 | | | | 46kb 3' of RP11-323F5.2 | intronic |
| 4 | 4684498 | 0.97 | 0.99 | rs9994026 | G | A | 0.60 | 0.54 | 0.69 | 0.58 | | | | | | CACD,NRSF | | | | 47kb 3' of RP11-323F5.2 | intronic |
| 4 | 4684800 | 1 | 1 | rs10010480 | T | C | 0.63 | 0.54 | 0.70 | 0.59 | | | | | | Gm397 | | | | 47kb 3' of RP11-323F5.2 | intronic |
| 4 | 4684931 | 1 | 1 | rs6822533 | G | C | 0.64 | 0.54 | 0.70 | 0.59 | | | ESDR | | | | | | | 47kb 3' of RP11-323F5.2 | intronic |
| 4 | 4684988 | 1 | 1 | rs9968282 | G | C | 0.64 | 0.54 | 0.70 | 0.59 | | | ESDR | | | 9 altered motifs | | | | 47kb 3' of RP11-323F5.2 | intronic |
| 4 | 4685122 | 0.81 | 0.94 | rs59988523 | TGGC | T | 0.85 | 0.51 | 0.68 | 0.51 | | | ESDR | | | 5 altered motifs | | | | 47kb 3' of RP11-323F5.2 | intronic |
| 4 | 4685551 | 0.84 | 1 | rs4594662 | C | T | 0.61 | 0.49 | 0.51 | 0.54 | | | 10 tissues | | | GATA,Hand1,SRF | | | | 48kb 3' of RP11-323F5.2 | intronic |
| 4 | 4685612 | 1 | 1 | rs4458410 | A | G | 0.64 | 0.54 | 0.70 | 0.59 | | | 10 tissues | ESDR,OVRY,MUS | | BRCA1,CEBPA,RFX5 | | 1 hit | | 48kb 3' of RP11-323F5.2 | intronic |
| 4 | 4685910 | 0.84 | 1 | rs7694974 | A | G | 0.61 | 0.49 | 0.51 | 0.54 | | | 10 tissues | 4 tissues | | | | 1 hit | | 48kb 3' of RP11-323F5.2 | intronic |
| 4 | 4686054 | 0.84 | 1 | rs7699823 | A | G | 0.61 | 0.49 | 0.51 | 0.54 | | | 10 tissues | 5 tissues | | 4 altered motifs | | | | 48kb 3' of RP11-323F5.2 | intronic |
| 4 | 4686171 | 0.84 | 1 | rs10000106 | G | C | 0.61 | 0.49 | 0.51 | 0.54 | | | | GI | | 4 altered motifs | | | | 48kb 3' of RP11-323F5.2 | intronic |
| 4 | 4686222 | 0.84 | 1 | rs10009010 | C | T | 0.61 | 0.49 | 0.51 | 0.54 | | | | GI,MUS | | 4 altered motifs | | | | 48kb 3' of RP11-323F5.2 | intronic |
| 4 | 4686401 | 0.84 | 1 | rs10013666 | A | G | 0.61 | 0.49 | 0.51 | 0.54 | | | | | | AP-2,ERalpha-a,RXRA | | | | 48kb 3' of RP11-323F5.2 | intronic |
| 4 | 4686448 | 0.81 | 1 | rs10009304 | C | A | 0.61 | 0.48 | 0.51 | 0.52 | | | | | | 7 altered motifs | | | | 49kb 3' of RP11-323F5.2 | intronic |
| 4 | 4686456 | 0.81 | 1 | rs10013757 | A | G | 0.61 | 0.48 | 0.51 | 0.52 | | | | | | PLAG1,TCF12 | | | | 49kb 3' of RP11-323F5.2 | intronic |
| 4 | 4686458 | 0.8 | 1 | rs10000410 | G | T | 0.60 | 0.48 | 0.51 | 0.52 | | | | | | TCF12 | | | | 49kb 3' of RP11-323F5.2 | intronic |
| 4 | 4686519 | 0.84 | 1 | rs7673523 | G | C | 0.61 | 0.49 | 0.51 | 0.54 | | | GI | | | | | | | 49kb 3' of RP11-323F5.2 | intronic |
| 4 | 4687470 | 0.83 | 1 | rs10016929 | A | G | 0.57 | 0.49 | 0.51 | 0.53 | | | | | | | | | | 50kb 3' of RP11-323F5.2 | intronic |
| 4 | 4687645 | 0.83 | 1 | rs6446677 | T | C | 0.61 | 0.49 | 0.51 | 0.54 | | | | | | Ik-1 | | | | 50kb 3' of RP11-323F5.2 | intronic |
| 4 | 4687835 | 0.83 | 1 | rs6818421 | A | C | 0.61 | 0.49 | 0.51 | 0.54 | | | | | | AIRE | | 1 hit | | 50kb 3' of RP11-323F5.2 | intronic |
| 4 | 4687859 | 0.83 | 1 | rs34934205 | TCA | T | 0.61 | 0.49 | 0.51 | 0.54 | | | | | | 5 altered motifs | | | | 50kb 3' of RP11-323F5.2 | intronic |
| 4 | 4688649 | 0.98 | 0.99 | rs10015850 | C | T | 0.63 | 0.54 | 0.70 | 0.60 | | | | ESDR,KID,OVRY | | Myf | | | | 51kb 3' of RP11-323F5.2 | intronic |
| 4 | 4688727 | 0.98 | 0.99 | rs7668054 | A | G | 0.64 | 0.54 | 0.70 | 0.60 | | | ESDR, LNG, OVRY | ESDR | | BCL,NRSF | | | | 51kb 3' of RP11-323F5.2 | intronic |
| 4 | 4689207 | 0.95 | 0.99 | rs7673538 | A | G | 0.87 | 0.54 | 0.70 | 0.60 | | | | | | CDP | | | | 51kb 3' of RP11-323F5.2 | intronic |
| 4 | 4692243 | 0.91 | 0.96 | rs11731637 | T | C | 0.83 | 0.54 | 0.75 | 0.61 | | | 4 tissues | GI | | 5 altered motifs | | | | 54kb 3' of RP11-323F5.2 | intronic |
| 4 | 4692627 | 0.91 | 0.97 | rs13149919 | C | T | 0.84 | 0.54 | 0.75 | 0.61 | | | GI, LIV | | | 5 altered motifs | | | | 55kb 3' of RP11-323F5.2 | intronic |
| 4 | 4695944 | 0.91 | 0.97 | rs7656684 | G | A | 0.84 | 0.54 | 0.75 | 0.61 | | | GI, LIV | | | 9 altered motifs | | | | 58kb 3' of RP11-323F5.2 | intronic |
| 4 | 4698546 | 0.89 | 0.96 | rs4340754 | T | C | 0.84 | 0.54 | 0.75 | 0.61 | | | GI, PLCNT | | | Foxa | | | | 61kb 3' of RP11-323F5.2 | intronic |
| 4 | 4701225 | 0.87 | 0.94 | rs9291150 | G | C | 0.82 | 0.54 | 0.73 | 0.61 | | | | | | 6 altered motifs | | | | 63kb 3' of RP11-323F5.2 | intronic |
| 4 | 4708476 | 0.85 | 0.94 | rs2280040 | T | C | 0.86 | 0.55 | 0.72 | 0.61 | | | | GI | | Elf3 | | 1 hit | | 71kb 3' of RP11-323F5.2 | intronic |

Query SNP: rs1013192 and variants with $r^2$ >= 0.8

| chr | pos (hg38) | LD ($r^2$) | LD (D') | variant | Ref | Alt | AFR freq | AMR freq | ASN freq | EUR freq | SiPhy cons | Promoter histone marks | Enhancer histone marks | DNAse | Proteins bound | Motifs changed | NHGRI/EBI GWAS hits | GRASP QTL hits | Selected eQTL hits | GENCODE genes | dbSNP func annot |
|---|---|---|---|---|---|---|---|---|---|---|---|---|---|---|---|---|---|---|---|---|---|
| 5 | 169750355 | 0.8 | 0.95 | rs35787483 | C | T | 0.08 | 0.35 | 0.61 | 0.43 | | | | | | E2F | | | | DOCK2 | intronic |
| 5 | 169772099 | 1 | 1 | rs1013192 | C | T | 0.09 | 0.32 | 0.61 | 0.43 | | | | | | NRSF,Sin3Ak-20 | | 1 hit | | DOCK2 | intronic |
| 5 | 169774995 | 0.85 | 0.97 | rs11134589 | G | A | 0.07 | 0.30 | 0.46 | 0.40 | | | | | | 6 altered motifs | | | | CTB-37A13.1 | intronic |

Query SNP: rs29795 and variants with $r^2$ >= 0.8

| chr | pos (hg38) | LD ($r^2$) | LD (D') | variant | Ref | Alt | AFR freq | AMR freq | ASN freq | EUR freq | SiPhy cons | Promoter histone marks | Enhancer histone marks | DNAse | Proteins bound | Motifs changed | NHGRI/EBI GWAS hits | GRASP QTL hits | Selected eQTL hits | GENCODE genes | dbSNP func annot |
|---|---|---|---|---|---|---|---|---|---|---|---|---|---|---|---|---|---|---|---|---|---|
| 5 | 173100615 | 0.84 | 0.93 | rs793359 | T | G | 0.34 | 0.46 | 0.16 | 0.63 | | | BLD | | | PLZF,SZF1-1,Spz1 | | | 6 hits | CREBRF | intronic |
| 5 | 173101548 | 0.85 | 0.93 | rs412682 | G | A | 0.35 | 0.47 | 0.16 | 0.63 | | | | | KAP1,SETDB1 | GR,HMG-IY,Pax-4 | | | 6 hits | CREBRF | intronic |
| 5 | 173101696 | 0.85 | 0.93 | rs416043 | A | G | 0.34 | 0.47 | 0.16 | 0.63 | | | | | | Crx | | | 6 hits | CREBRF | intronic |
| 5 | 173102788 | 0.85 | 0.93 | rs384717 | T | C | 0.35 | 0.47 | 0.16 | 0.63 | | | 11 tissues | | | | | | 6 hits | CREBRF | intronic |
| 5 | 173103474 | 0.85 | 0.93 | rs807428 | A | G | 0.35 | 0.47 | 0.16 | 0.63 | | | 5 tissues | SKIN | | DMRT3 | | | 9 hits | CREBRF | intronic |
| 5 | 173103558 | 0.85 | 0.93 | rs793356 | T | C | 0.35 | 0.47 | 0.16 | 0.63 | | | 5 tissues | SKIN | | Rad21,SP1 | | | 9 hits | CREBRF | intronic |
| 5 | 173104604 | 0.84 | 0.93 | rs251233 | G | C | 0.35 | 0.46 | 0.16 | 0.63 | | | BRN | | | Zic | | | 6 hits | CREBRF | intronic |

| | | | | | | | | | | | | | | | | | | | | | |
|---|---|---|---|---|---|---|---|---|---|---|---|---|---|---|---|---|---|---|---|---|---|
| 5 | 173104860 | 0.85 | 0.93 | rs251234 | T | C | 0.35 | 0.47 | 0.16 | 0.63 | | | | | | EWSR1-FLI1,SP1,Smad4 | | | 6 hits | CREBRF | intronic |
| 5 | 173105562 | 0.8 | 0.93 | rs173692 | A | G | 0.33 | 0.45 | 0.16 | 0.60 | | | | | | 6 altered motifs | | | 5 hits | CREBRF | intronic |
| 5 | 173107677 | 0.83 | 0.91 | rs251236 | G | C | 0.35 | 0.47 | 0.16 | 0.63 | | | | | | | | | 9 hits | CREBRF | intronic |
| 5 | 173116814 | 0.84 | 0.93 | rs251237 | C | A | 0.35 | 0.46 | 0.16 | 0.63 | | | | | | Ik-2,NF-AT1 | | | 7 hits | CREBRF | intronic |
| 5 | 173118018 | 0.82 | 0.94 | rs1091969 | A | C | 0.34 | 0.45 | 0.16 | 0.62 | | | | | | AP-1 | | | 6 hits | CREBRF | intronic |
| 5 | 173121706 | 0.89 | 0.96 | rs166122 | G | A | 0.35 | 0.47 | 0.16 | 0.63 | | | | | | Gfi1,Gfi1b | | | 9 hits | CREBRF | intronic |
| 5 | 173125399 | 0.89 | 0.96 | rs251232 | T | A | 0.35 | 0.47 | 0.16 | 0.63 | | | | | | 7 altered motifs | | | 6 hits | CREBRF | intronic |
| 5 | 173131193 | 0.89 | 0.96 | rs3131913 | G | A | 0.35 | 0.47 | 0.16 | 0.63 | | | | | | 4 altered motifs | | | 9 hits | CREBRF | intronic |
| 5 | 173142841 | 0.98 | 1 | rs2544579 | C | G | 0.35 | 0.47 | 0.16 | 0.63 | | | | | | 7 altered motifs | | | 6 hits | 1.3kb 3' of CTC-209H22.3 | |
| 5 | 173143217 | 0.97 | 0.99 | rs171725 | T | C | 0.35 | 0.47 | 0.16 | 0.63 | | | | | | 5 altered motifs | | | 6 hits | 944bp 3' of CTC-209H22.3 | |
| 5 | 173147173 | 0.99 | 1 | rs29797 | G | C | 0.35 | 0.47 | 0.17 | 0.63 | | | | | | 7 altered motifs | | | 4 hits | BNIP1 | intronic |
| 5 | 173150400 | 1 | 1 | rs29795 | G | A | 0.35 | 0.47 | 0.17 | 0.64 | | | | | | CDP,Sox | | 1 hit | 9 hits | BNIP1 | intronic |
| 5 | 173153863 | 1 | 1 | rs255292 | C | A | 0.35 | 0.47 | 0.17 | 0.64 | | | | | | 5 altered motifs | | | 9 hits | BNIP1 | intronic |
| 5 | 173158076 | 0.98 | 0.99 | rs255298 | G | A | 0.36 | 0.47 | 0.16 | 0.64 | | | | | | 4 altered motifs | | | 5 hits | BNIP1 | intronic |
| 5 | 173158924 | 0.97 | 0.99 | rs5745160 | TC | T | 0.35 | 0.48 | 0.16 | 0.64 | | | | | | 22 altered motifs | | | 6 hits | BNIP1 | intronic |
| 5 | 173159269 | 0.97 | 0.99 | rs255299 | C | T | 0.34 | 0.47 | 0.16 | 0.63 | | | | | | CTCF,Nr2f2,SMC3 | | | 6 hits | BNIP1 | intronic |
| 5 | 173161024 | 0.82 | 0.99 | rs255303 | G | T | 0.35 | 0.43 | 0.07 | 0.64 | | | | | | Hdx | | | 6 hits | BNIP1 | intronic |

Query SNP: rs7800398 and variants with $r^2$ >= 0.8

| chr | pos (hg38) | LD (r²) | LD (D') | variant | Ref | Alt | AFR freq | AMR freq | ASN freq | EUR freq | SiPhy cons | Promoter histone marks | Enhancer histone marks | DNAse | Proteins bound | Motifs changed | NHGRI/EBI GWAS hits | GRASP QTL hits | Selected eQTL hits | GENCODE genes | dbSNP func annot |
|---|---|---|---|---|---|---|---|---|---|---|---|---|---|---|---|---|---|---|---|---|---|
| 7 | 21047458 | 1 | 1 | rs7800398 | A | C | 0.50 | 0.50 | 0.46 | 0.51 | | | | | | HNF1,Pou2f2 | | | | 25kb 3' of AC006481.1 | |

Query SNP: rs9301479 and variants with $r^2$ >= 0.8

| chr | pos (hg38) | LD (r²) | LD (D') | variant | Ref | Alt | AFR freq | AMR freq | ASN freq | EUR freq | SiPhy cons | Promoter histone marks | Enhancer histone marks | DNAse | Proteins bound | Motifs changed | NHGRI/EBI GWAS hits | GRASP QTL hits | Selected eQTL hits | GENCODE genes | dbSNP func annot |
|---|---|---|---|---|---|---|---|---|---|---|---|---|---|---|---|---|---|---|---|---|---|
| 13 | 110788604 | 1 | 1 | rs9301479 | A | G | 0.50 | 0.59 | 0.83 | 0.42 | | | | | | 6 altered motifs | | 2 hits | | 21kb 3' of RP11-120J20.4 | |

Query SNP: rs9695 and variants with $r^2$ >= 0.8

| chr | pos (hg38) | LD (r²) | LD (D') | variant | Ref | Alt | AFR freq | AMR freq | ASN freq | EUR freq | SiPhy cons | Promoter histone marks | Enhancer histone marks | DNAse | Proteins bound | Motifs changed | NHGRI/EBI GWAS hits | GRASP QTL hits | Selected eQTL hits | GENCODE genes | dbSNP func annot |
|---|---|---|---|---|---|---|---|---|---|---|---|---|---|---|---|---|---|---|---|---|---|
| 9 | 94592811 | 0.9 | -1 | rs2772023 | A | G | 0.54 | 0.30 | 0.59 | 0.45 | | MUS | | | | | | | 1 hit | FBP2 | intronic |
| 9 | 94592918 | 1 | 1 | rs10114599 | C | G | 0.46 | 0.68 | 0.41 | 0.50 | | | | MUS | | | | | 1 hit | FBP2 | intronic |
| 9 | 94594154 | 0.95 | 1 | rs10115489 | C | T | 0.67 | 0.69 | 0.41 | 0.50 | | | | 4 tissues | | 8 altered motifs | | | 1 hit | 360bp 5' of FBP2 | |
| 9 | 94595616 | 1 | 1 | rs2017136 | C | A | 0.47 | 0.68 | 0.41 | 0.50 | | | GI, PANC, MUS | THYM | | GR,NRSF,Sin3Ak-20 | | | 1 hit | 1.8kb 5' of FBP2 | |
| 9 | 94596696 | 0.98 | 1 | rs11347826 | TA | T | 0.47 | 0.67 | 0.40 | 0.49 | | | | | | 9 altered motifs | | | | 2.9kb 5' of FBP2 | |
| 9 | 94599142 | 0.99 | 1 | rs6479555 | G | A | 0.46 | 0.68 | 0.41 | 0.50 | | | MUS, LIV | IPSC | | AP-4,LBP-9,Rad21 | | | 1 hit | 4kb 3' of FBP1 | |
| 9 | 94599928 | 0.95 | 1 | rs7872412 | A | C | 0.47 | 0.67 | 0.41 | 0.50 | | MUS | LIV, MUS | | | 6 altered motifs | | | 1 hit | 3.2kb 3' of FBP1 | |
| 9 | 94599982 | 1 | 1 | rs7856456 | G | A | 0.46 | 0.68 | 0.41 | 0.50 | | MUS | LIV, MUS | GI | | Mtf1,Pou6f1 | | | 1 hit | 3.2kb 3' of FBP1 | |
| 9 | 94600071 | 1 | 1 | rs7856557 | G | A | 0.46 | 0.68 | 0.41 | 0.50 | | MUS | LIV, MUS | MUS,GI | | KAP1,Pax-1 | | | 1 hit | 3.1kb 3' of FBP1 | |
| 9 | 94600895 | 1 | 1 | rs4743960 | G | A | 0.46 | 0.68 | 0.41 | 0.50 | | MUS, LIV | LIV, GI, MUS | IPSC,MUS,MUS | | EBF,NF-kappaB | | | 3 hits | 2.2kb 3' of FBP1 | |
| 9 | 94601338 | 1 | 1 | rs10761346 | G | A | 0.46 | 0.68 | 0.41 | 0.50 | | | | | | Pax-8,SIX5 | | | 3 hits | 1.8kb 3' of FBP1 | |
| 9 | 94603360 | 1 | 1 | rs9695 | G | A | 0.47 | 0.68 | 0.42 | 0.50 | | 6 tissues | 15 tissues | 25 tissues | 8 bound proteins | Ets,Spdef | | | 3 hits | FBP1 | 3'-UTR |
| 9 | 94606801 | 1 | 1 | rs2297084 | G | A | 0.46 | 0.68 | 0.41 | 0.50 | | GI, LIV | LIV, GI | 4 tissues | | 5 altered motifs | | | 1 hit | FBP1 | intronic |
| 9 | 94608820 | 1 | 1 | rs4744359 | G | A | 0.46 | 0.68 | 0.41 | 0.50 | | | 9 tissues | 7 tissues | 4 bound proteins | 4 altered motifs | | | 1 hit | FBP1 | intronic |
| 9 | 94609780 | 1 | 1 | rs3824484 | C | T | 0.44 | 0.68 | 0.41 | 0.51 | | | 11 tissues | | | 5 altered motifs | | | 3 hits | FBP1 | intronic |
| 9 | 94610995 | 0.84 | 0.99 | rs1977483 | C | G | 0.44 | 0.64 | 0.40 | 0.48 | | | LIV, GI, LNG | | | LUN-1 | | | 1 hit | FBP1 | intronic |
| 9 | 94611370 | 1 | 1 | rs1999122 | C | A | 0.44 | 0.68 | 0.41 | 0.50 | | | BLD, LIV, GI | | | Foxd3,STAT | | | 3 hits | FBP1 | intronic |
| 9 | 94611661 | 1 | 1 | rs1999123 | G | A | 0.44 | 0.68 | 0.41 | 0.50 | | | 4 tissues | | | NRSF,Rad21 | | | 1 hit | FBP1 | intronic |
| 9 | 94612453 | 1 | 1 | rs7044434 | A | G | 0.44 | 0.68 | 0.41 | 0.50 | | LIV | 5 tissues | PANC | | 5 altered motifs | | | 1 hit | FBP1 | intronic |

Query SNP: rs9298320 and variants with $r^2$ >= 0.8

| chr | pos (hg38) | LD (r²) | LD (D') | variant | Ref | Alt | AFR freq | AMR freq | ASN freq | EUR freq | SiPhy cons | Promoter histone marks | Enhancer histone marks | DNAse | Proteins bound | Motifs changed | NHGRI/EBI GWAS hits | GRASP QTL hits | Selected eQTL hits | GENCODE genes | dbSNP func annot |
|---|---|---|---|---|---|---|---|---|---|---|---|---|---|---|---|---|---|---|---|---|---|
| 8 | 78870852 | 0.93 | 1 | rs10110533 | G | A | 0.36 | 0.19 | 0.16 | 0.19 | | | | | | 13 altered motifs | | | | 30kb 3' of RP11-79H23.3 | |
| 8 | 78876087 | 1 | 1 | rs72669435 | G | T | 0.19 | 0.18 | 0.16 | 0.19 | | | | | | | | | | 36kb 3' of RP11-79H23.3 | |
| 8 | 78876884 | 1 | 1 | rs72669438 | C | T | 0.19 | 0.18 | 0.16 | 0.19 | | | | | | LUN-1 | | | | 36kb 3' of RP11-79H23.3 | |
| 8 | 78878090 | 0.93 | 1 | rs7007241 | A | T | 0.35 | 0.19 | 0.16 | 0.19 | | | | | | | | | | 38kb 3' of RP11-79H23.3 | |
| 8 | 78879036 | 1 | 1 | rs11993442 | A | G | 0.20 | 0.18 | 0.16 | 0.19 | | | | | | CDP,Nkx2,Sox | | 1 hit | | 39kb 3' of RP11-79H23.3 | |
| 8 | 78880704 | 0.9 | 1 | rs28415328 | C | T | 0.36 | 0.19 | 0.16 | 0.19 | | | | | | | | | | 40kb 3' of RP11-79H23.3 | |
| 8 | 78880811 | 1 | 1 | rs28529281 | G | A | 0.19 | 0.18 | 0.16 | 0.19 | | | | | | 4 altered motifs | | | | 40kb 3' of RP11-79H23.3 | |
| 8 | 78882825 | 0.88 | 0.98 | rs78281603 | T | C | 0.30 | 0.19 | 0.16 | 0.19 | | | | | | Mef2,Pou2f2,Pou3f3 | | | | 42kb 3' of RP11-79H23.3 | |
| 8 | 78885306 | 0.95 | 1 | rs6982562 | C | G | 0.30 | 0.19 | 0.16 | 0.19 | | | | | | | | | | 45kb 3' of RP11-79H23.3 | |
| 8 | 78888747 | 1 | 1 | rs11991031 | G | A | 0.19 | 0.18 | 0.16 | 0.19 | | | | | | Ets | | | | 48kb 3' of RP11-79H23.3 | |
| 8 | 78889730 | 1 | 1 | rs1822554 | C | T | 0.19 | 0.18 | 0.16 | 0.19 | | | | | | Egr-1,Pdx1,SETDB1 | | | | 49kb 3' of RP11-79H23.3 | |
| 8 | 78890603 | 1 | 1 | rs1848587 | T | C | 0.19 | 0.18 | 0.16 | 0.19 | | | | | | TCF4 | | | | 50kb 3' of RP11-79H23.3 | |
| 8 | 78894064 | 0.94 | 1 | rs72678354 | A | G | 0.17 | 0.17 | 0.16 | 0.19 | | | | | | DMRT5,GATA,Mef2 | | | | 54kb 3' of RP11-79H23.3 | |

| | | | | | | | | | | | | | | | | | | | | |
|---|---|---|---|---|---|---|---|---|---|---|---|---|---|---|---|---|---|---|---|---|
| 8 | 78894069 | 0.94 | 1 | rs150187101 | A | AAAAT | 0.17 | 0.17 | 0.16 | 0.20 | | | | | | 18 altered motifs | | | | 54kb 3' of RP11-79H23.3 |
| 8 | 78894222 | 0.98 | 1 | rs9987170 | T | C | 0.19 | 0.18 | 0.16 | 0.19 | | | | | | | | | | 54kb 3' of RP11-79H23.3 |
| 8 | 78894333 | 0.82 | 1 | rs9987337 | G | T | 0.45 | 0.21 | 0.16 | 0.22 | | | | | | Gfi1,Pbx-1 | | | | 54kb 3' of RP11-79H23.3 |
| 8 | 78897033 | 1 | 1 | rs10957901 | G | A | 0.20 | 0.18 | 0.16 | 0.19 | | | | | | HNF1 | | | | 57kb 3' of RP11-79H23.3 |
| 8 | 78897375 | 1 | 1 | rs10097803 | C | T | 0.21 | 0.18 | 0.16 | 0.19 | | | | | | GR,Hbp1,Sox | | | | 57kb 3' of RP11-79H23.3 |
| 8 | 78897791 | 0.95 | 1 | rs10098173 | A | G | 0.30 | 0.19 | 0.16 | 0.19 | | | | | | 6 altered motifs | | 1 hit | | 57kb 3' of RP11-79H23.3 |
| 8 | 78897923 | 1 | 1 | rs9298320 | G | C | 0.20 | 0.18 | 0.16 | 0.19 | | | | | | Duxl,Pbx-1 | | 1 hit | | 57kb 3' of RP11-79H23.3 |
| 8 | 78898294 | 1 | 1 | rs17508243 | C | T | 0.19 | 0.18 | 0.16 | 0.19 | | | | | | Hand1,STAT,YY1 | | | | 58kb 3' of RP11-79H23.3 |
| 8 | 78899904 | 1 | 1 | rs10106354 | C | T | 0.19 | 0.18 | 0.16 | 0.19 | | | | BRN | | | | | | 59kb 3' of RP11-79H23.3 |
| 8 | 78900119 | 0.95 | 1 | rs10106730 | C | T | 0.30 | 0.19 | 0.16 | 0.19 | | | | | | 8 altered motifs | | 1 hit | | 60kb 3' of RP11-79H23.3 |
| 8 | 78901383 | 1 | 1 | rs10504684 | C | G | 0.19 | 0.18 | 0.16 | 0.19 | | | | | | | | 1 hit | | 61kb 3' of RP11-79H23.3 |
| 8 | 78902873 | 1 | 1 | rs11989307 | T | C | 0.19 | 0.18 | 0.16 | 0.19 | | | | | | Nrf1,Pou2f2,TATA | | 1 hit | | 62kb 3' of RP11-79H23.3 |
| 8 | 78903344 | 1 | 1 | rs35751168 | TTCTC | T | 0.19 | 0.18 | 0.16 | 0.19 | | | | | | 10 altered motifs | | | | 63kb 3' of RP11-79H23.3 |
| 8 | 78904614 | 0.98 | 1 | rs11991235 | T | G | 0.20 | 0.18 | 0.16 | 0.19 | | | BRN | | | 4 altered motifs | | 1 hit | | 64kb 3' of RP11-79H23.3 |

Query SNP: rs2063244 and variants with $r^2$ >= 0.8

| chr | pos (hg38) | LD ($r^2$) | LD (D') | variant | Ref | Alt | AFR freq | AMR freq | ASN freq | EUR freq | SiPhy cons | Promoter histone marks | Enhancer histone marks | DNAse | Proteins bound | Motifs changed | NHGRI/EBI GWAS hits | GRASP QTL hits | Selected eQTL hits | GENCODE genes | dbSNP func annot |
|---|---|---|---|---|---|---|---|---|---|---|---|---|---|---|---|---|---|---|---|---|---|
| 5 | 90908948 | 0.82 | 0.91 | rs6452914 | G | T | 0.16 | 0.30 | 0.00 | 0.53 | | | 6 tissues | VAS | | 7 altered motifs | | | | GPR98 | intronic |
| 5 | 90931719 | 0.92 | -0.97 | rs3105790 | G | T | 0.57 | 0.70 | 0.99 | 0.46 | | | 8 tissues | | MAFK | 11 altered motifs | | | | GPR98 | intronic |
| 5 | 90932242 | 0.89 | -0.97 | rs3114655 | C | T | 0.51 | 0.69 | 0.99 | 0.46 | | | 10 tissues | 28 tissues | 7 bound proteins | 5 altered motifs | | | | GPR98 | intronic |
| 5 | 90932614 | 0.92 | -0.97 | rs3105789 | A | T | 0.64 | 0.70 | 1.00 | 0.47 | | | 9 tissues | ESC | | DMRT2,Sox | | | | GPR98 | intronic |
| 5 | 90933609 | 0.96 | -0.99 | rs10942618 | T | G | 0.65 | 0.70 | 1.00 | 0.47 | | | | | | SREBP | | | | GPR98 | intronic |
| 5 | 90933713 | 0.97 | 1 | rs11744005 | C | A | 0.34 | 0.30 | 0.00 | 0.53 | | | 11 tissues | | | Ets,GATA | | | | GPR98 | intronic |
| 5 | 90933725 | 0.95 | 0.97 | rs11750515 | A | G | 0.26 | 0.30 | 0.00 | 0.53 | | | 11 tissues | | | Rad21 | | 1 hit | | GPR98 | intronic |
| 5 | 90934955 | 0.97 | 1 | rs4415111 | A | G | 0.34 | 0.30 | 0.00 | 0.53 | | | ESDR, PLCNT, LIV | | | SP1 | | | | GPR98 | intronic |
| 5 | 90935421 | 0.97 | 1 | rs2367182 | T | C | 0.40 | 0.30 | 0.01 | 0.54 | | | | | | 6 altered motifs | | | | GPR98 | intronic |
| 5 | 90935431 | 0.95 | 0.97 | rs6867530 | G | T | 0.26 | 0.30 | 0.00 | 0.53 | | | | | | 6 altered motifs | | | | GPR98 | intronic |
| 5 | 90939480 | 1 | 1 | rs11749470 | C | T | 0.34 | 0.30 | 0.00 | 0.52 | | | 11 tissues | 13 tissues | | AP-4 | | | | GPR98 | intronic |
| 5 | 90940830 | 1 | 1 | rs60301478 | AC | A | 0.34 | 0.30 | 0.00 | 0.52 | | | 18 tissues | 31 tissues | 6 bound proteins | GATA,SP2 | | | | GPR98 | intronic |
| 5 | 90943088 | 0.96 | 1 | rs1967413 | G | A | 0.39 | 0.31 | 0.01 | 0.52 | | | | | | GR,MZF1::1-4 | | | | GPR98 | intronic |
| 5 | 90944211 | 1 | 1 | rs2063244 | T | A | 0.34 | 0.30 | 0.00 | 0.51 | | | BRST, SKIN, LNG | 6 tissues | | 4 altered motifs | | | | GPR98 | intronic |
| 5 | 90948287 | 0.94 | 0.97 | rs13163934 | G | A | 0.36 | 0.30 | 0.00 | 0.51 | | | GI, LNG, LIV | | | GATA,GR,HDAC2 | | | | GPR98 | intronic |
| 5 | 90948711 | 0.94 | 0.97 | rs10942619 | A | T | 0.36 | 0.30 | 0.00 | 0.51 | | | | | | Mrg1::Hoxa9,PRDM1 | | | | GPR98 | intronic |
| 5 | 90949095 | 0.92 | 0.96 | rs7716083 | C | A | 0.37 | 0.30 | 0.00 | 0.51 | | | GI, LNG, LIV | | MAFF,MAFK | Brachyury | | | | GPR98 | intronic |
| 5 | 90949896 | 0.94 | 0.97 | rs10057134 | C | T | 0.36 | 0.30 | 0.00 | 0.51 | | | | 11 tissues | 15 bound proteins | GR,Nkx3,TCF4 | | | | GPR98 | intronic |
| 5 | 90951025 | 0.94 | 0.97 | rs7705201 | T | G | 0.36 | 0.30 | 0.00 | 0.51 | | | LIV | | | PEBP | | | | GPR98 | intronic |

Query SNP: rs1952183 and variants with $r^2$ >= 0.8

| chr | pos (hg38) | LD ($r^2$) | LD (D') | variant | Ref | Alt | AFR freq | AMR freq | ASN freq | EUR freq | SiPhy cons | Promoter histone marks | Enhancer histone marks | DNAse | Proteins bound | Motifs changed | NHGRI/EBI GWAS hits | GRASP QTL hits | Selected eQTL hits | GENCODE genes | dbSNP func annot |
|---|---|---|---|---|---|---|---|---|---|---|---|---|---|---|---|---|---|---|---|---|---|
| 14 | 89370999 | 0.9 | 1 | rs35510295 | ACT | A | 0.03 | 0.57 | 0.38 | 0.51 | | GI | 15 tissues | 10 tissues | FOXA1 | 5 altered motifs | | | 3 hits | FOXN3 | intronic |
| 14 | 89371352 | 0.9 | 1 | rs12895962 | G | A | 0.03 | 0.57 | 0.39 | 0.51 | | | 15 tissues | BLD | | 4 altered motifs | | | 3 hits | FOXN3 | intronic |
| 14 | 89371759 | 0.9 | 1 | rs1958070 | A | G | 0.03 | 0.57 | 0.39 | 0.52 | | MUS, GI | 20 tissues | 6 tissues | | EWSR1-FLI1,STAT,p300 | | | 3 hits | FOXN3 | intronic |
| 14 | 89372748 | 0.92 | 1 | rs764815 | T | C | 0.04 | 0.57 | 0.39 | 0.51 | | | 11 tissues | 4 tissues | | 4 altered motifs | | | 3 hits | FOXN3 | intronic |
| 14 | 89377395 | 0.96 | 1 | rs2160218 | C | T | 0.03 | 0.58 | 0.27 | 0.51 | | | BLD, ADRL | MUS | | GCNF,Pax-6 | | | 3 hits | FOXN3 | intronic |
| 14 | 89377767 | 1 | 1 | rs1952183 | A | G | 0.12 | 0.59 | 0.27 | 0.51 | | | 4 tissues | | | Arnt | | | 4 hits | FOXN3 | intronic |
| 14 | 89378952 | 0.93 | 1 | rs12890227 | C | T | 0.03 | 0.58 | 0.28 | 0.51 | | | 12 tissues | | | 4 altered motifs | | | 3 hits | FOXN3 | intronic |
| 14 | 89380667 | 0.92 | 1 | rs12880900 | C | T | 0.03 | 0.57 | 0.28 | 0.51 | | | 4 tissues | BLD | | 4 altered motifs | | | 3 hits | FOXN3 | intronic |
| 14 | 89381249 | 0.93 | 0.97 | rs10484022 | G | A | 0.17 | 0.59 | 0.27 | 0.51 | | | | GI | | Arid5a,RXRA | | | 3 hits | FOXN3 | intronic |
| 14 | 89384322 | 0.84 | 0.96 | rs200648484 | T | TAGAAA | 0.10 | 0.57 | 0.20 | 0.49 | | BLD | 9 tissues | 9 tissues | 4 bound proteins | 7 altered motifs | | | 3 hits | FOXN3 | intronic |

Query SNP: rs11784756 and variants with $r^2$ >= 0.8

| chr | pos (hg38) | LD ($r^2$) | LD (D') | variant | Ref | Alt | AFR freq | AMR freq | ASN freq | EUR freq | SiPhy cons | Promoter histone marks | Enhancer histone marks | DNAse | Proteins bound | Motifs changed | NHGRI/EBI GWAS hits | GRASP QTL hits | Selected eQTL hits | GENCODE genes | dbSNP func annot |
|---|---|---|---|---|---|---|---|---|---|---|---|---|---|---|---|---|---|---|---|---|---|
| 8 | 72189416 | 0.83 | 0.94 | rs60053573 | A | G | 0.17 | 0.05 | 0.00 | 0.07 | | | | | | DMRT2,Hand1,TAL1 | | | | 6.9kb 5' of RP11-142A23.1 | |
| 8 | 72195504 | 1 | 1 | rs11784756 | C | T | 0.12 | 0.05 | 0.00 | 0.07 | | | | | JUND | | | | | 829bp 5' of RP11-142A23.1 | |

Query SNP: rs2201409 and variants with $r^2$ >= 0.8

| chr | pos (hg38) | LD ($r^2$) | LD (D') | variant | Ref | Alt | AFR freq | AMR freq | ASN freq | EUR freq | SiPhy cons | Promoter histone marks | Enhancer histone marks | DNAse | Proteins bound | Motifs changed | NHGRI/EBI GWAS hits | GRASP QTL hits | Selected eQTL hits | GENCODE genes | dbSNP func annot |
|---|---|---|---|---|---|---|---|---|---|---|---|---|---|---|---|---|---|---|---|---|---|
| 5 | 35798580 | 1 | 1 | rs2201409 | A | G | 0.03 | 0.21 | 0.09 | 0.18 | | | BLD | | | 6 altered motifs | | | | CTD-2113L7.1 | intronic |

Query SNP: rs12499915 and variants with $r^2$ >= 0.8

| chr | pos (hg38) | LD ($r^2$) | LD (D') | variant | Ref | Alt | AFR freq | AMR freq | ASN freq | EUR freq | SiPhy cons | Promoter histone marks | Enhancer histone marks | DNAse | Proteins bound | Motifs changed | NHGRI/EBI GWAS hits | GRASP QTL hits | Selected eQTL hits | GENCODE genes | dbSNP func annot |
|---|---|---|---|---|---|---|---|---|---|---|---|---|---|---|---|---|---|---|---|---|---|

| chr | pos (hg38) | LD (r²) | LD (D') | variant | Ref | Alt | AFR freq | AMR freq | ASN freq | EUR freq | SiPhy cons | Promoter histone marks | Enhancer histone marks | DNAse | Proteins bound | Motifs changed | NHGRI/ EBI GWAS hits | GRASP QTL hits | Selected eQTL hits | GENCODE genes | dbSNP func annot |
|---|---|---|---|---|---|---|---|---|---|---|---|---|---|---|---|---|---|---|---|---|---|
| 4 | 111693635 | 0.86 | -0.95 | rs35288474 | CTG | C | 0.40 | 0.60 | 0.42 | 0.67 | | | GI | | | 5 altered motifs | | | | 45kb 3' of RP11-255I10.2 | |
| 4 | 111706142 | 0.89 | 0.96 | rs13110448 | A | C | 0.55 | 0.40 | 0.58 | 0.33 | | | | | | Cdc5,TATA | | | | 57kb 3' of RP11-255I10.2 | |
| 4 | 111731295 | 1 | 1 | rs12499915 | G | A | 0.48 | 0.38 | 0.58 | 0.32 | | | | | | Cdx,HNF4 | | | 1 hit | 72kb 5' of RP11-269F21.1 | |
| 4 | 111731920 | 0.82 | 1 | rs1450786 | A | G | 0.55 | 0.43 | 0.58 | 0.37 | | | | | | 7 altered motifs | | | | 71kb 5' of RP11-269F21.1 | |
| 4 | 111739429 | 0.94 | 0.98 | rs1450783 | T | A | 0.45 | 0.38 | 0.58 | 0.32 | | | | | | Irf | | | | 63kb 5' of RP11-269F21.1 | |
| 4 | 111755929 | 0.94 | 0.98 | rs1381010 | G | A | 0.45 | 0.38 | 0.58 | 0.32 | | | | | | NF-I,STAT,TLX1::NFIC | | | | 47kb 5' of RP11-269F21.1 | |
| 4 | 111759644 | 0.91 | 0.96 | rs4833976 | C | T | 0.41 | 0.38 | 0.58 | 0.32 | | | | | | 5 altered motifs | | | | 43kb 5' of RP11-269F21.1 | |
| 4 | 111787889 | 0.9 | 0.96 | rs1903403 | T | G | 0.42 | 0.38 | 0.60 | 0.32 | | 4 tissues | 4 tissues | | | 6 altered motifs | | | | 15kb 5' of RP11-269F21.1 | |

Query SNP: rs1896384 and variants with $r^2$ >= 0.8

| chr | pos (hg38) | LD (r²) | LD (D') | variant | Ref | Alt | AFR freq | AMR freq | ASN freq | EUR freq | SiPhy cons | Promoter histone marks | Enhancer histone marks | DNAse | Proteins bound | Motifs changed | NHGRI/ EBI GWAS hits | GRASP QTL hits | Selected eQTL hits | GENCODE genes | dbSNP func annot |
|---|---|---|---|---|---|---|---|---|---|---|---|---|---|---|---|---|---|---|---|---|---|
| 10 | 123711496 | 0.88 | 0.95 | rs12218312 | C | T | 0.24 | 0.54 | 0.66 | 0.47 | | | | | | HNF1,Homez,p300 | | | | CPXM2 | |
| 10 | 123720669 | 0.96 | 0.99 | rs10902799 | C | T | 0.25 | 0.55 | 0.66 | 0.48 | | | | | ZNF263 | HDAC2 | | | | CPXM2 | |
| 10 | 123722561 | 0.9 | 0.97 | rs1896385 | G | A | 0.25 | 0.54 | 0.66 | 0.47 | | | | | | CDP,Myc | | | | CPXM2 | |
| 10 | 123724318 | 0.9 | 0.97 | rs4980152 | T | C | 0.23 | 0.54 | 0.65 | 0.47 | | | | | | 5 altered motifs | | | | CPXM2 | |
| 10 | 123726603 | 1 | 1 | rs1896384 | C | T | 0.22 | 0.55 | 0.65 | 0.48 | | | BRN | | | HDAC2,HNF4 | | | | CPXM2 | |
| 10 | 123728399 | 0.92 | 0.98 | rs12358957 | C | T | 0.24 | 0.54 | 0.65 | 0.47 | | | ESC, ESDR | | | BRCA1,MIF-1 | | | | CPXM2 | |
| 10 | 123732949 | 0.84 | -0.93 | rs1955107 | G | C | 0.62 | 0.44 | 0.35 | 0.53 | | | | | | Cdx | | | | CPXM2 | |
| 10 | 123734084 | 0.96 | 0.99 | rs10902805 | G | A | 0.22 | 0.54 | 0.66 | 0.48 | | | | | | 8 altered motifs | | | | CPXM2 | |
| 10 | 123735341 | 0.97 | 0.99 | rs10902807 | C | G | 0.22 | 0.54 | 0.65 | 0.48 | | | | BRN | | Nkx6-1 | | | | CPXM2 | |
| 10 | 123735859 | 0.86 | 0.97 | rs34870401 | C | T | 0.25 | 0.57 | 0.65 | 0.49 | | | | | | 7 altered motifs | | | | CPXM2 | |
| 10 | 123736493 | 0.98 | 0.99 | rs12218121 | T | C | 0.22 | 0.55 | 0.66 | 0.48 | | | | | | DMRT2,DMRT3 | | | | CPXM2 | |

Query SNP: rs12680681 and variants with $r^2$ >= 0.8

| chr | pos (hg38) | LD (r²) | LD (D') | variant | Ref | Alt | AFR freq | AMR freq | ASN freq | EUR freq | SiPhy cons | Promoter histone marks | Enhancer histone marks | DNAse | Proteins bound | Motifs changed | NHGRI/ EBI GWAS hits | GRASP QTL hits | Selected eQTL hits | GENCODE genes | dbSNP func annot |
|---|---|---|---|---|---|---|---|---|---|---|---|---|---|---|---|---|---|---|---|---|---|
| 8 | 59876083 | 0.88 | 0.94 | rs7815966 | C | T | 0.74 | 0.41 | 0.63 | 0.45 | | | | 4 tissues | GATA2,TAL1 | CHOP::CEBPalpha | | | | 174kb 5' of RP11-27P7.1 | |
| 8 | 59879089 | 0.89 | 0.95 | rs6471839 | A | G | 0.74 | 0.41 | 0.63 | 0.45 | | | | | | Hoxa10,Nanog | | | | 170kb 5' of RP11-27P7.1 | |
| 8 | 59892806 | 0.98 | 0.99 | rs4737550 | G | T | 0.82 | 0.42 | 0.63 | 0.45 | | | | | | Zfp187 | | | | 157kb 5' of RP11-27P7.1 | |
| 8 | 59894128 | 0.97 | 0.99 | rs10094881 | A | T | 0.74 | 0.41 | 0.63 | 0.45 | | | | | | | | | | 155kb 5' of RP11-27P7.1 | |
| 8 | 59894594 | 0.97 | 0.99 | rs1600676 | T | G | 0.74 | 0.41 | 0.63 | 0.45 | | | | | | 5 altered motifs | | | | 155kb 5' of RP11-27P7.1 | |
| 8 | 59905166 | 0.97 | 0.99 | rs6471841 | T | C | 0.74 | 0.41 | 0.63 | 0.45 | | | | | | 6 altered motifs | | | | 144kb 5' of RP11-27P7.1 | |
| 8 | 59906090 | 0.97 | 0.99 | rs6471842 | A | G | 0.82 | 0.41 | 0.63 | 0.45 | | | | | | Mef2,TCF12,TEF | | | | 143kb 5' of RP11-27P7.1 | |
| 8 | 59909197 | 0.99 | 1 | rs1351437 | T | G | 0.74 | 0.41 | 0.63 | 0.45 | | | | | | 4 altered motifs | | | | 140kb 5' of RP11-27P7.1 | |
| 8 | 59911743 | 1 | 1 | rs7012579 | G | C | 0.82 | 0.42 | 0.63 | 0.45 | | | | | | | | | | 138kb 5' of RP11-27P7.1 | |
| 8 | 59912024 | 0.99 | 1 | rs2170278 | A | C | 0.74 | 0.41 | 0.63 | 0.45 | | | | | | GR,NF-kappaB,Pax-5 | | | | 138kb 5' of RP11-27P7.1 | |
| 8 | 59912243 | 1 | 1 | rs2127830 | A | C | 0.82 | 0.42 | 0.63 | 0.45 | | | | | | 7 altered motifs | | | | 137kb 5' of RP11-27P7.1 | |
| 8 | 59914019 | 0.99 | 1 | rs7838569 | T | A | 0.74 | 0.41 | 0.64 | 0.45 | | | | | | Foxp1,Mef2,p300 | | | | 136kb 5' of RP11-27P7.1 | |
| 8 | 59923110 | 0.99 | 1 | rs4737551 | A | G | 0.74 | 0.41 | 0.63 | 0.45 | | | LNG | | | GR,TCF4 | | | | 126kb 5' of RP11-27P7.1 | |
| 8 | 59927525 | 1 | 1 | rs6471843 | T | C | 0.76 | 0.42 | 0.63 | 0.46 | | | | | | 6 altered motifs | | | | 122kb 5' of RP11-27P7.1 | |
| 8 | 59929843 | 1 | 1 | rs1497032 | T | G | 0.77 | 0.42 | 0.63 | 0.46 | | | | | | Maf,Pou6f1 | | | | 120kb 5' of RP11-27P7.1 | |
| 8 | 59932041 | 1 | 1 | rs6471844 | C | T | 0.76 | 0.42 | 0.63 | 0.46 | | | 5 tissues | CRVX | | 7 altered motifs | | | | 118kb 5' of RP11-27P7.1 | |
| 8 | 59935098 | 0.82 | 1 | rs4257994 | G | A | 0.64 | 0.37 | 0.60 | 0.39 | | | | | | | | | | 114kb 5' of RP11-27P7.1 | |
| 8 | 59935454 | 1 | 1 | rs7845735 | C | A | 0.76 | 0.42 | 0.63 | 0.46 | | | | | | 4 altered motifs | | | | 114kb 5' of RP11-27P7.1 | |
| 8 | 59939038 | 1 | 1 | rs7830239 | A | G | 0.79 | 0.42 | 0.63 | 0.46 | | | | | | 17 altered motifs | | | | 111kb 5' of RP11-27P7.1 | |
| 8 | 59943194 | 1 | 1 | rs12680681 | G | A | 0.76 | 0.42 | 0.63 | 0.46 | | | | | | | | | | 106kb 5' of RP11-27P7.1 | |
| 8 | 59943590 | 1 | 1 | rs11777453 | A | G | 0.76 | 0.42 | 0.63 | 0.46 | | | | | | MZF1::1-4 | | | | 106kb 5' of RP11-27P7.1 | |
| 8 | 59946471 | 1 | 1 | rs4256581 | T | C | 0.76 | 0.42 | 0.63 | 0.46 | | | | BRN,OVRY,PANC | | 4 altered motifs | | | | 103kb 5' of RP11-27P7.1 | |
| 8 | 59949854 | 0.98 | 0.99 | rs530844 | T | C | 0.75 | 0.42 | 0.63 | 0.45 | | | | | | BDP1,GR | | | | 100kb 5' of RP11-27P7.1 | |
| 8 | 59951314 | 0.92 | 0.99 | rs653657 | A | T | 0.66 | 0.40 | 0.52 | 0.46 | | | | | | Mef2 | | | | 98kb 5' of RP11-27P7.1 | |
| 8 | 59951759 | 0.9 | 0.99 | rs665987 | T | G | 0.65 | 0.40 | 0.52 | 0.45 | | | | | | DMRT5 | | | | 98kb 5' of RP11-27P7.1 | |
| 8 | 59952365 | 0.82 | 0.99 | rs813630 | A | C | 0.68 | 0.38 | 0.57 | 0.41 | | | | | | 6 altered motifs | | | | 97kb 5' of RP11-27P7.1 | |
| 8 | 59952385 | 0.85 | 0.99 | rs809137 | C | A | 0.58 | 0.38 | 0.49 | 0.42 | | | | | | | | | | 97kb 5' of RP11-27P7.1 | |
| 8 | 59952676 | 0.88 | 1 | rs1084448 | A | G | 0.71 | 0.39 | 0.56 | 0.42 | | | | | | 4 altered motifs | | | | 97kb 5' of RP11-27P7.1 | |
| 8 | 59952869 | 0.84 | 0.98 | rs1084449 | G | A | 0.64 | 0.39 | 0.51 | 0.44 | | | | | | | | | | 97kb 5' of RP11-27P7.1 | |
| 8 | 59956513 | 0.93 | 0.98 | rs782021 | T | C | 0.77 | 0.42 | 0.63 | 0.46 | | | | | | 26 altered motifs | | | | 93kb 5' of RP11-27P7.1 | |
| 8 | 59957960 | 0.87 | 0.93 | rs782022 | G | A | 0.79 | 0.42 | 0.63 | 0.46 | | | | | | 6 altered motifs | | | | 92kb 5' of RP11-27P7.1 | |
| 8 | 59959817 | 0.87 | 0.93 | rs526249 | T | C | 0.79 | 0.42 | 0.63 | 0.46 | | | ESC, IPSC | | | 5 altered motifs | | | | 90kb 5' of RP11-27P7.1 | |
| 8 | 59961997 | 0.82 | 0.93 | rs574368 | G | A | 0.68 | 0.40 | 0.52 | 0.45 | | | BLD, LNG | LNG | | | | | | 88kb 5' of RP11-27P7.1 | |
| 8 | 59962548 | 0.82 | 0.93 | rs668563 | T | C | 0.68 | 0.40 | 0.52 | 0.45 | | | BLD, LNG | | | DMRT7,Foxp1 | | | | 87kb 5' of RP11-27P7.1 | |
| 8 | 59963041 | 0.86 | 0.93 | rs522182 | T | C | 0.76 | 0.41 | 0.63 | 0.46 | | | BLD, HRT | | | 4 altered motifs | | | | 87kb 5' of RP11-27P7.1 | |

| | | | | | | | | | | | | | | | | | | | | |
|---|---|---|---|---|---|---|---|---|---|---|---|---|---|---|---|---|---|---|---|---|
| 8 | 59963265 | 0.83 | 0.93 | rs671655 | A | C | 0.71 | 0.41 | 0.52 | 0.45 | | BLD | BLD, HRT | | | HNF6,Hoxa10,Pou3f2 | | | | 86kb 5' of RP11-27P7.1 |
| 8 | 59964893 | 0.86 | 0.93 | rs2688659 | A | C | 0.76 | 0.41 | 0.63 | 0.46 | | | BLD | | | Foxl1 | | | | 85kb 5' of RP11-27P7.1 |
| 8 | 59965370 | 0.82 | 0.93 | rs594035 | T | C | 0.68 | 0.40 | 0.52 | 0.46 | | | BLD | | | 6 altered motifs | | | | 84kb 5' of RP11-27P7.1 |
| | | 0.86 | 0.93 | rs62742161 | G | A | 0.76 | 0.41 | 0.63 | 0.46 | | | | | | 4 altered motifs | | | | 84kb 5' of RP11-27P7.1 |
| 8 | 59967263 | 0.82 | 0.93 | rs475714 | C | T | 0.68 | 0.40 | 0.52 | 0.46 | | | BLD | PANC | | | | | | 82kb 5' of RP11-27P7.1 |
| 8 | 59967602 | 0.86 | 0.93 | rs472799 | G | A | 0.76 | 0.41 | 0.63 | 0.46 | | | BLD, GI, BRN | 4 tissues | | Foxo,Mef2 | | | | 82kb 5' of RP11-27P7.1 |
| 8 | 59968087 | 0.86 | 0.93 | rs2037561 | C | G | 0.76 | 0.41 | 0.63 | 0.46 | | | BLD, BRN, GI | MUS | | 12 altered motifs | | | | 81kb 5' of RP11-27P7.1 |
| 8 | 59970440 | 0.82 | 0.93 | rs493713 | C | T | 0.68 | 0.40 | 0.52 | 0.46 | | | | ESDR | | | | | | 79kb 5' of RP11-27P7.1 |
| 8 | 59971011 | 0.82 | 0.93 | rs488934 | T | C | 0.68 | 0.40 | 0.52 | 0.45 | | | ESDR, BLD, PLCNT | ESDR,PLCNT | | 4 altered motifs | | | | 79kb 5' of RP11-27P7.1 |
| 8 | 59973661 | 0.83 | 0.93 | rs627156 | G | A | 0.78 | 0.41 | 0.63 | 0.44 | | | | | | AFP1,Mef2 | | | | 76kb 5' of RP11-27P7.1 |
| 8 | 59973973 | 0.81 | 0.92 | rs639026 | A | G | 0.76 | 0.41 | 0.63 | 0.44 | | | | MUS | | CEBPA | | | | 76kb 5' of RP11-27P7.1 |
| 8 | 59974195 | 0.82 | 0.93 | rs485523 | T | A | 0.76 | 0.40 | 0.63 | 0.44 | | | | | | | | | | 75kb 5' of RP11-27P7.1 |
| 8 | 59975855 | 0.82 | 0.93 | rs673973 | T | C | 0.76 | 0.40 | 0.63 | 0.44 | | | | | | 6 altered motifs | | | | 74kb 5' of RP11-27P7.1 |
| 8 | 59976637 | 0.83 | 0.93 | rs497326 | C | A | 0.79 | 0.41 | 0.63 | 0.44 | | | | | | HNF1,Hdx | | | | 73kb 5' of RP11-27P7.1 |
| 8 | 59976692 | 0.83 | 0.93 | rs688365 | A | C | 0.79 | 0.41 | 0.63 | 0.44 | | | | | | 5 altered motifs | | | | 73kb 5' of RP11-27P7.1 |
| 8 | 59977455 | 0.83 | 0.93 | rs594017 | T | A,C | 0.78 | 0.41 | 0.63 | 0.44 | | | | | | | | | | 72kb 5' of RP11-27P7.1 |
| 8 | 59977855 | 0.82 | 0.93 | rs595812 | A | G | 0.76 | 0.40 | 0.63 | 0.44 | | | | | | | | | | 72kb 5' of RP11-27P7.1 |
| 8 | 59980115 | 0.82 | 0.93 | rs491891 | T | G | 0.76 | 0.40 | 0.63 | 0.44 | | | | | | 4 altered motifs | | | | 69kb 5' of RP11-27P7.1 |
| 8 | 59980912 | 0.82 | 0.93 | rs484432 | C | G | 0.75 | 0.40 | 0.63 | 0.44 | | | | | | Zfp128 | | | | 69kb 5' of RP11-27P7.1 |
| 8 | 59985162 | 0.81 | 0.92 | rs619715 | C | G | 0.79 | 0.41 | 0.63 | 0.44 | | | | | | | | | | 64kb 5' of RP11-27P7.1 |
| 8 | 59991100 | 0.81 | 0.92 | rs600437 | A | G | 0.79 | 0.41 | 0.63 | 0.44 | | | | | | SRF | | | | 58kb 5' of RP11-27P7.1 |
| 8 | 59996092 | 0.81 | 0.92 | rs582877 | G | A | 0.79 | 0.41 | 0.63 | 0.44 | | | | | | 20 altered motifs | | | | 53kb 5' of RP11-27P7.1 |
| 8 | 59998149 | 0.81 | 0.92 | rs541081 | A | T | 0.75 | 0.41 | 0.63 | 0.44 | | | | | | 4 altered motifs | | | | 51kb 5' of RP11-27P7.1 |
| 8 | 59998968 | 0.81 | 0.92 | rs533776 | A | T | 0.79 | 0.41 | 0.63 | 0.44 | | | | | | Pax-5,TCF11::MafG | | | | 51kb 5' of RP11-27P7.1 |
| 8 | 59999201 | 0.8 | 0.92 | rs627807 | C | A | 0.71 | 0.40 | 0.63 | 0.44 | | | | | | | | | | 50kb 5' of RP11-27P7.1 |
| 8 | 60000138 | 0.81 | 0.93 | rs642072 | G | A | 0.69 | 0.40 | 0.63 | 0.44 | | | | BRN | | 4 altered motifs | | | | 49kb 5' of RP11-27P7.1 |
| 8 | 60000427 | 0.81 | 0.92 | rs478307 | G | T | 0.75 | 0.41 | 0.63 | 0.44 | | | | | | TEF | | | | 49kb 5' of RP11-27P7.1 |
| 8 | 60000700 | 0.81 | 0.92 | rs475684 | T | C | 0.84 | 0.41 | 0.63 | 0.44 | | | | | | ATF3,CTCF | | | | 49kb 5' of RP11-27P7.1 |
| 8 | 60002224 | 0.81 | 0.93 | rs671985 | G | A | 0.69 | 0.40 | 0.63 | 0.44 | | | | | | 8 altered motifs | | | | 47kb 5' of RP11-27P7.1 |
| 8 | 60002829 | 0.81 | 0.93 | rs674767 | C | T | 0.69 | 0.40 | 0.63 | 0.44 | | | | | | 7 altered motifs | | | | 47kb 5' of RP11-27P7.1 |
| 8 | 60003173 | 0.81 | 0.93 | rs686889 | C | A | 0.69 | 0.40 | 0.63 | 0.44 | | | | | | CACD,INSM1,RREB-1 | | | | 46kb 5' of RP11-27P7.1 |

Query SNP: rs10890879 and variants with $r^2$ >= 0.8

| chr | pos (hg38) | LD ($r^2$) | LD (D') | variant | Ref | Alt | AFR freq | AMR freq | ASN freq | EUR freq | SiPhy cons | Promoter histone marks | Enhancer histone marks | DNAse | Proteins bound | Motifs changed | NHGRI/EBI GWAS hits | GRASP QTL hits | Selected eQTL hits | GENCODE genes | dbSNP func annot |
|---|---|---|---|---|---|---|---|---|---|---|---|---|---|---|---|---|---|---|---|---|---|
| 11 | 108660942 | 0.92 | 0.99 | rs10749923 | C | G | 0.31 | 0.60 | 0.44 | 0.56 | | | | SKIN | | EWSR1-FLI1,Isl2 | | | | 4.1kb 5' of DDX10 | |
| 11 | 108665065 | 0.86 | 1 | rs2306118 | C | T | 0.10 | 0.57 | 0.44 | 0.56 | | 24 tissues | | 53 tissues | 24 bound proteins | 4 altered motifs | | | | DDX10 | |
| 11 | 108665560 | 0.91 | 1 | rs10789675 | A | C | 0.10 | 0.59 | 0.44 | 0.56 | | | | 5 tissues | | | | | | DDX10 | intronic |
| 11 | 108669211 | 0.92 | 0.99 | rs10890877 | A | G | 0.32 | 0.60 | 0.44 | 0.56 | | | | | | | | | | DDX10 | intronic |
| 11 | 108669775 | 0.86 | 1 | rs11212750 | G | A | 0.09 | 0.57 | 0.44 | 0.56 | | BLD | 6 tissues | 4 tissues | | 9 altered motifs | | | | DDX10 | intronic |
| 11 | 108675081 | 1 | 1 | rs10890879 | A | G | 0.31 | 0.61 | 0.44 | 0.56 | | | | ESDR,VAS | CFOS,CJUN | 15 altered motifs | | | | DDX10 | intronic |
| 11 | 108687234 | 0.89 | 0.99 | rs10789676 | A | G | 0.09 | 0.59 | 0.44 | 0.56 | | | | | | 5 altered motifs | | | | DDX10 | intronic |
| 11 | 108688326 | 0.84 | 0.99 | rs10890883 | A | G | 0.09 | 0.57 | 0.44 | 0.56 | | | | | | AIRE,Dobox4 | | | | DDX10 | intronic |
| 11 | 108693228 | 0.89 | 0.99 | rs10789677 | T | C | 0.10 | 0.59 | 0.43 | 0.56 | | | | | | 6 altered motifs | | | | DDX10 | intronic |
| 11 | 108694827 | 0.89 | 0.99 | rs3781867 | G | C | 0.09 | 0.59 | 0.43 | 0.55 | | | 4 tissues | | | ERalpha-a,Esr2 | | | | DDX10 | intronic |
| 11 | 108695791 | 0.87 | 0.96 | rs2169562 | A | G | 0.09 | 0.59 | 0.43 | 0.55 | | | | | | 7 altered motifs | | | | DDX10 | intronic |
| 11 | 108698679 | 0.81 | 0.91 | rs10749926 | T | C | 0.25 | 0.61 | 0.49 | 0.56 | | | | BRST,SKIN | | Evi-1,Smad3 | | | | DDX10 | intronic |
| 11 | 108709220 | 0.87 | 0.98 | rs10890888 | A | C | 0.15 | 0.59 | 0.43 | 0.56 | | | | BLD | | Pax-4 | | | | DDX10 | intronic |
| 11 | 108713020 | 0.95 | 0.98 | rs7116062 | A | G | 0.34 | 0.61 | 0.43 | 0.56 | | BLD | STRM, BRN | ESDR,BLD | | Gfi1 | | | | DDX10 | intronic |
| 11 | 108717498 | 0.82 | 0.98 | rs1824664 | A | T | 0.09 | 0.57 | 0.43 | 0.55 | | | | | | GATA,Lmo2-complex,Mef2 | | | | DDX10 | intronic |
| 11 | 108718587 | 0.82 | 0.98 | rs4027488 | G | A | 0.09 | 0.57 | 0.43 | 0.56 | | | STRM | | | 4 altered motifs | | | | DDX10 | intronic |
| 11 | 108721600 | 0.86 | 0.98 | rs3781866 | C | T | 0.09 | 0.59 | 0.43 | 0.56 | | | | | | 4 altered motifs | | | | DDX10 | intronic |
| 11 | 108721781 | 0.87 | 0.98 | rs3781864 | T | C | 0.09 | 0.59 | 0.43 | 0.56 | | | | | | SRF | | | | DDX10 | intronic |
| 11 | 108736640 | 0.81 | 0.96 | rs11212771 | G | C | 0.09 | 0.58 | 0.43 | 0.56 | | | | | | 4 altered motifs | | | | DDX10 | intronic |
| 11 | 108738884 | 0.94 | 0.98 | rs11212774 | A | C | 0.31 | 0.61 | 0.43 | 0.56 | | | ESDR, BLD | | | 5 altered motifs | | | | DDX10 | intronic |
| 11 | 108739720 | 0.89 | 0.96 | rs7940595 | T | G | 0.32 | 0.60 | 0.43 | 0.56 | | | | | | 8 altered motifs | | | | DDX10 | intronic |
| 11 | 108739954 | 0.81 | 0.96 | rs7103086 | G | A | 0.10 | 0.58 | 0.44 | 0.56 | | | | | | 7 altered motifs | | | | DDX10 | intronic |
| 11 | 108743367 | 0.84 | 0.92 | rs3902843 | G | T | 0.44 | 0.61 | 0.43 | 0.55 | | | | | | AIRE,Cdx2,Hoxa10 | | | | DDX10 | intronic |
| 11 | 108750868 | 0.81 | 0.94 | rs77056910 | CCACTT | C | 0.30 | 0.59 | 0.43 | 0.55 | | | | | | 4 altered motifs | | | | DDX10 | intronic |

Query SNP: rs636926 and variants with $r^2$ >= 0.8

| chr | pos (hg38) | LD ($r^2$) | LD (D') | variant | Ref | Alt | AFR freq | AMR freq | ASN freq | EUR freq | SiPhy cons | Promoter histone marks | Enhancer histone marks | DNAse | Proteins bound | Motifs changed | NHGRI/EBI GWAS hits | GRASP QTL hits | Selected eQTL hits | GENCODE genes | dbSNP func annot |
|---|---|---|---|---|---|---|---|---|---|---|---|---|---|---|---|---|---|---|---|---|---|
| 5 | 73121814 | 0.98 | 1 | rs556535 | T | A | 0.48 | 0.36 | 0.57 | 0.26 | | 6 tissues | 7 tissues | | | Nr2f2,TLX1::NFIC | | | 2 hits | TMEM171 | intronic |
| 5 | 73123629 | 1 | 1 | rs637450 | C | G | 0.51 | 0.35 | 0.57 | 0.26 | | | 13 tissues | ESC,IPSC | | CTCF,HDAC2,Pax-5 | | | 2 hits | TMEM171 | missense |
| 5 | 73123790 | 1 | 1 | rs636926 | C | A | 0.55 | 0.35 | 0.57 | 0.26 | | | 7 tissues | | | LBP-9,p300 | | 3 hits | 2 hits | TMEM171 | missense |
| 5 | 73126191 | 0.96 | 0.99 | rs570888 | G | A | 0.60 | 0.35 | 0.57 | 0.26 | | | 5 tissues | | | 8 altered motifs | | | 1 hit | TMEM171 | intronic |

Query SNP: rs848481 and variants with $r^2$ >= 0.8

| chr | pos (hg38) | LD ($r^2$) | LD (D') | variant | Ref | Alt | AFR freq | AMR freq | ASN freq | EUR freq | SiPhy cons | Promoter histone marks | Enhancer histone marks | DNAse | Proteins bound | Motifs changed | NHGRI/EBI GWAS hits | GRASP QTL hits | Selected eQTL hits | GENCODE genes | dbSNP func annot |
|---|---|---|---|---|---|---|---|---|---|---|---|---|---|---|---|---|---|---|---|---|---|
| 7 | 77776424 | 0.83 | -0.92 | rs2868813 | T | C | 0.88 | 0.43 | 0.31 | 0.50 | | | | | | 5 altered motifs | | 1 hit | 38 hits | RSBN1L | intronic |
| 7 | 77780065 | 0.83 | -0.92 | rs28716648 | T | C | 0.87 | 0.43 | 0.31 | 0.50 | | | BRN | | | 10 altered motifs | | | 34 hits | 373bp 3' of RSBN1L | |
| 7 | 77780997 | 0.8 | -0.92 | rs6954671 | G | C | 0.85 | 0.42 | 0.30 | 0.50 | | | GI | | | 4 altered motifs | | | 38 hits | 1.3kb 3' of RSBN1L | |
| 7 | 77784468 | 0.82 | -0.92 | rs6958015 | A | G | 0.86 | 0.42 | 0.31 | 0.49 | | | LIV, GI | | | 5 altered motifs | | | 33 hits | 4.8kb 3' of RSBN1L | |
| 7 | 77786223 | 0.84 | -0.93 | rs10242184 | C | G | 0.87 | 0.42 | 0.31 | 0.49 | | | | BRN,LNG | | Pax-6 | | | 35 hits | 6.5kb 3' of RSBN1L | |
| 7 | 77787630 | 0.84 | -0.93 | rs6949795 | T | C | 0.87 | 0.42 | 0.30 | 0.48 | | | LNG | | | 5 altered motifs | | | 33 hits | 6.1kb 3' of TMEM60 | |
| 7 | 77788384 | 0.84 | -0.93 | rs1544458 | A | G | 0.87 | 0.42 | 0.31 | 0.49 | | | | | | | | | 35 hits | 5.3kb 3' of TMEM60 | |
| 7 | 77789973 | 0.8 | -0.92 | rs1544457 | G | T | 0.86 | 0.42 | 0.31 | 0.49 | | | BLD | GI | | GZF1 | | 1 hit | 40 hits | 3.8kb 3' of TMEM60 | |
| 7 | 77820544 | 0.81 | -0.9 | rs9769871 | T | G | 0.88 | 0.43 | 0.30 | 0.52 | | | | | | 5 altered motifs | | | 31 hits | PHTF2 | intronic |
| 7 | 77822495 | 0.84 | -0.92 | rs10248334 | C | T | 0.90 | 0.43 | 0.30 | 0.50 | | | | HRT,MUS,BLD | 7 bound proteins | 4 altered motifs | | | 34 hits | PHTF2 | intronic |
| 7 | 77824787 | 0.83 | -0.92 | rs6465848 | T | C | 0.89 | 0.44 | 0.30 | 0.52 | | BLD | BLD | BLD | | | | | 35 hits | PHTF2 | intronic |
| 7 | 77837891 | 0.86 | -0.93 | rs1526748 | G | A | 0.91 | 0.43 | 0.30 | 0.50 | | | 7 tissues | | | Myc | | | 38 hits | PHTF2 | intronic |
| 7 | 77847685 | 0.85 | -0.93 | rs2091402 | A | G | 0.91 | 0.44 | 0.31 | 0.52 | | | | | | | | | 32 hits | PHTF2 | intronic |
| 7 | 77851254 | 0.86 | -0.93 | rs10248871 | G | A | 0.91 | 0.43 | 0.30 | 0.50 | | | | | | Hoxa9,Hoxd10,NF-kappaB | | | 32 hits | PHTF2 | intronic |
| 7 | 77856917 | 0.85 | -0.93 | rs1405420 | T | G | 0.89 | 0.44 | 0.31 | 0.52 | | | MUS | | | | | | 35 hits | PHTF2 | intronic |
| 7 | 77861917 | 0.87 | -0.94 | rs7802691 | T | C | 0.90 | 0.44 | 0.31 | 0.51 | | | | | | 5 altered motifs | | | 31 hits | PHTF2 | intronic |
| 7 | 77868155 | 0.89 | -0.94 | rs6465867 | T | C | 0.90 | 0.43 | 0.31 | 0.50 | | | 7 tissues | LNG | | 11 altered motifs | | | 33 hits | PHTF2 | intronic |
| 7 | 77868403 | 0.89 | -0.94 | rs7803160 | A | G | 0.91 | 0.43 | 0.30 | 0.49 | | | 5 tissues | SKIN | | BDP1,Hic1 | | | 33 hits | PHTF2 | intronic |
| 7 | 77874778 | 0.9 | -0.95 | rs2471584 | G | C | 0.87 | 0.43 | 0.31 | 0.51 | | | MUS | | | | | | 32 hits | PHTF2 | intronic |
| 7 | 77880557 | 0.89 | -0.95 | rs2463008 | T | C | 0.88 | 0.44 | 0.31 | 0.50 | | | 7 tissues | IPSC | | 8 altered motifs | | | 36 hits | PHTF2 | intronic |
| 7 | 77884568 | 0.89 | -0.95 | rs2463010 | T | C | 0.87 | 0.44 | 0.31 | 0.51 | | | | | | HMG-IY,Mef2,Pax-4 | | | 36 hits | PHTF2 | intronic |
| 7 | 77885072 | 0.89 | -0.95 | rs2463011 | A | C | 0.87 | 0.44 | 0.31 | 0.51 | | | | | | 5 altered motifs | | | 32 hits | PHTF2 | intronic |
| 7 | 77894204 | 0.89 | -0.95 | rs10267180 | A | G | 0.90 | 0.44 | 0.31 | 0.51 | | | | | | Pou2f2 | | | 31 hits | PHTF2 | intronic |
| 7 | 77894388 | 0.89 | -0.95 | rs10252383 | C | T | 0.88 | 0.44 | 0.31 | 0.51 | | | | | | | | | 32 hits | PHTF2 | intronic |
| 7 | 77895882 | 0.89 | -0.95 | rs2471603 | T | C | 0.90 | 0.44 | 0.31 | 0.52 | | | BLD | BLD | | Brachyury | | 3 hits | 31 hits | Y_RNA | intronic |
| 7 | 77898139 | 0.9 | 0.95 | rs848449 | A | G | 0.09 | 0.57 | 0.70 | 0.50 | | | | | | 8 altered motifs | | | 36 hits | PHTF2 | intronic |
| 7 | 77903204 | 0.96 | 0.98 | rs848493 | A | C | 0.10 | 0.57 | 0.68 | 0.49 | | | BLD | | | 9 altered motifs | | 1 hit | 37 hits | PHTF2 | intronic |
| 7 | 77910427 | 0.97 | 1 | rs711309 | G | A | 0.10 | 0.56 | 0.69 | 0.50 | | | | | CTCF | 8 altered motifs | | 2 hits | 36 hits | PHTF2 | intronic |
| 7 | 77914702 | 0.99 | 1 | rs848478 | T | G | 0.09 | 0.57 | 0.70 | 0.50 | | | BLD | | | Ik-2,LUN-1 | | | 34 hits | PHTF2 | intronic |
| 7 | 77915787 | 1 | 1 | rs848481 | T | A | 0.13 | 0.57 | 0.70 | 0.50 | | | BLD, GI | | | Ets,GR | | 1 hit | 36 hits | PHTF2 | intronic |
| 7 | 77916209 | 0.98 | 1 | rs848482 | A | C | 0.09 | 0.56 | 0.70 | 0.50 | | | PANC | | | Irf,PRDM1,PU.1 | | | 32 hits | PHTF2 | intronic |
| 7 | 77920004 | 0.98 | 1 | rs848484 | C | A | 0.13 | 0.56 | 0.69 | 0.48 | | | | | | 5 altered motifs | | | 33 hits | PHTF2 | intronic |
| 7 | 77922165 | 0.98 | 1 | rs848485 | G | T | 0.13 | 0.56 | 0.69 | 0.48 | | | | | | 4 altered motifs | | | 33 hits | PHTF2 | intronic |
| 7 | 77925305 | 0.98 | 1 | rs848489 | G | A | 0.13 | 0.56 | 0.69 | 0.48 | | | | | | PLZF | | | 33 hits | PHTF2 | intronic |
| 7 | 77926224 | 0.98 | 1 | rs848491 | C | G | 0.13 | 0.56 | 0.69 | 0.49 | | | | | | 4 altered motifs | | | 31 hits | PHTF2 | intronic |
| 7 | 77932057 | 0.93 | 0.98 | rs1725749 | C | T | 0.10 | 0.56 | 0.69 | 0.49 | | | | ESDR,BLD | | 4 altered motifs | | | 32 hits | PHTF2 | intronic |
| 7 | 77932585 | 0.91 | 0.97 | rs6150174 | 17-mer | G | 0.12 | 0.56 | 0.67 | 0.48 | | | BLD, CRVX | | | HDAC2,PRDM1 | | | 33 hits | PHTF2 | intronic |
| | | 0.9 | 0.97 | rs150364025 | CTTAG | C | 0.10 | 0.56 | 0.69 | 0.49 | | | BLD | | | LUN-1 | | | 1 hit | PHTF2 | intronic |
| 7 | 77948501 | 0.91 | 0.95 | rs34160426 | A | C | 0.09 | 0.57 | 0.69 | 0.51 | | | | | | 5 altered motifs | | | 36 hits | PHTF2 | intronic |
| 7 | 77952948 | 0.91 | 0.95 | rs848461 | T | C | 0.08 | 0.57 | 0.69 | 0.50 | | | | IPSC | | Irf,PU.1 | | 1 hit | 36 hits | PHTF2 | intronic |
| 7 | 77953346 | 0.91 | 0.95 | rs848462 | A | G | 0.10 | 0.57 | 0.69 | 0.50 | | | | | | 52 altered motifs | | | 33 hits | PHTF2 | intronic |
| 7 | 77962605 | 0.9 | 0.95 | rs861049 | G | C | 0.16 | 0.57 | 0.67 | 0.49 | | | | | | CAC-binding-protein,TATA | | | 33 hits | 5.1kb 3' of PHTF2 | |

Query SNP: rs7232775 and variants with $r^2$ >= 0.8

| chr | pos (hg38) | LD ($r^2$) | LD (D') | variant | Ref | Alt | AFR freq | AMR freq | ASN freq | EUR freq | SiPhy cons | Promoter histone marks | Enhancer histone marks | DNAse | Proteins bound | Motifs changed | NHGRI/EBI GWAS hits | GRASP QTL hits | Selected eQTL hits | GENCODE genes | dbSNP func annot |
|---|---|---|---|---|---|---|---|---|---|---|---|---|---|---|---|---|---|---|---|---|---|
| 18 | 45610826 | 0.87 | -0.94 | rs17669582 | A | G | 0.05 | 0.25 | 0.18 | 0.30 | | | 4 tissues | THYM | | 5 altered motifs | | | | 4kb 5' of SLC14A2 | intronic |
| 18 | 45612481 | 0.93 | 0.99 | rs1905673 | T | C | 0.86 | 0.74 | 0.82 | 0.70 | | | 4 tissues | | | AIRE,Pou5f1 | | 1 hit | | 2.3kb 5' of SLC14A2 | intronic |
| 18 | 45622439 | 1 | 1 | rs7232775 | C | T | 0.92 | 0.75 | 0.82 | 0.72 | | | | ESDR | | Nkx2,Nkx3,Pax-6 | 1 hit | 1 hit | | SLC14A2 | intronic |

Query SNP: rs4917774 and variants with $r^2$ >= 0.8

| chr | pos (hg38) | LD ($r^2$) | LD (D') | variant | Ref | Alt | AFR freq | AMR freq | ASN freq | EUR freq | SiPhy cons | Promoter histone marks | Enhancer histone marks | DNAse | Proteins bound | Motifs changed | NHGRI/EBI GWAS hits | GRASP QTL hits | Selected eQTL hits | GENCODE genes | dbSNP func annot |
|---|---|---|---|---|---|---|---|---|---|---|---|---|---|---|---|---|---|---|---|---|---|
| 10 | 97520476 | 0.98 | 1 | rs4919105 | A | G | 0.51 | 0.51 | 0.61 | 0.45 | | | 6 tissues | | | SZF1-1 | | 1 hit | 9 hits | UBTD1 | intronic |
| 10 | 97520808 | 0.93 | 0.98 | rs7075656 | G | C | 0.61 | 0.52 | 0.61 | 0.45 | | | 7 tissues | ESC,IPSC | | CTCF,Rad21,SMC3 | | | 11 hits | UBTD1 | intronic |
| 10 | 97522036 | 0.89 | 1 | rs7089060 | A | G | 0.44 | 0.48 | 0.54 | 0.44 | | | 9 tissues | ESC | | | | 1 hit | 8 hits | UBTD1 | intronic |
| 10 | 97522115 | 1 | 1 | rs4917774 | A | G | 0.51 | 0.51 | 0.61 | 0.45 | | | 10 tissues | | | | | 9 hits | 9 hits | UBTD1 | intronic |

Query SNP: rs17776811 and variants with $r^2$ >= 0.8

| chr | pos (hg38) | LD ($r^2$) | LD (D') | variant | Ref | Alt | AFR freq | AMR freq | ASN freq | EUR freq | SiPhy cons | Promoter histone marks | Enhancer histone marks | DNAse | Proteins bound | Motifs changed | NHGRI/EBI GWAS hits | GRASP QTL hits | Selected eQTL hits | GENCODE genes | dbSNP func annot |
|---|---|---|---|---|---|---|---|---|---|---|---|---|---|---|---|---|---|---|---|---|---|
| 14 | 89589124 | 0.84 | 0.99 | rs35889227 | G | T | 0.12 | 0.34 | 0.13 | 0.62 | | BLD | 6 tissues | 4 tissues | 6 bound proteins | 5 altered motifs | | | | FOXN3 | intronic |
| 14 | 89591899 | 0.98 | 1 | rs34513589 | G | A | 0.13 | 0.37 | 0.15 | 0.61 | | | IPSC, BLD, LIV | | | CTCF,SZF1-1 | | | | FOXN3 | intronic |
| 14 | 89599983 | 0.92 | 0.99 | rs11622292 | C | T | 0.22 | 0.36 | 0.15 | 0.60 | | | | BLD | | HNF4,RAR,RXRA | | | | FOXN3 | intronic |
| 14 | 89600107 | 1 | 1 | rs17776811 | A | C | 0.16 | 0.38 | 0.15 | 0.61 | | BLD | BLD | | | PTF1-beta | | | | FOXN3 | intronic |
| 14 | 89600290 | 0.93 | 0.98 | rs35802157 | T | C | 0.14 | 0.37 | 0.15 | 0.62 | | BLD | BLD | BLD,GI | | 7 altered motifs | | | | FOXN3 | intronic |
| 14 | 89603418 | 0.83 | 0.94 | rs11159913 | T | C | 0.18 | 0.39 | 0.18 | 0.65 | | | 5 tissues | GI | | | | 2 hits | | FOXN3 | intronic |

Query SNP: rs2483702 and variants with $r^2$ >= 0.8

| chr | pos (hg38) | LD ($r^2$) | LD (D') | variant | Ref | Alt | AFR freq | AMR freq | ASN freq | EUR freq | SiPhy cons | Promoter histone marks | Enhancer histone marks | DNAse | Proteins bound | Motifs changed | NHGRI/EBI GWAS hits | GRASP QTL hits | Selected eQTL hits | GENCODE genes | dbSNP func annot |
|---|---|---|---|---|---|---|---|---|---|---|---|---|---|---|---|---|---|---|---|---|---|
| 10 | 25354596 | 1 | 1 | rs2483702 | A | G | 0.09 | 0.07 | 0.11 | 0.09 | | | | | | 5 altered motifs | | | | GPR158 | intronic |
| 10 | 25364888 | 0.96 | 1 | rs66484783 | T | G | 0.08 | 0.07 | 0.11 | 0.09 | | | | | | Irf,PLZF | | | | GPR158 | intronic |
| 10 | 25366562 | 0.8 | 1 | rs67814683 | T | A | 0.08 | 0.06 | 0.11 | 0.07 | | | | | | | | | | GPR158 | intronic |
| 10 | 25372558 | 0.8 | 0.95 | rs193165201 | G | A | 0.08 | 0.07 | 0.10 | 0.09 | | | | | | GR | | | | GPR158 | intronic |

| | | | | | | | | | | | | | | | | | | | | | |
|---|---|---|---|---|---|---|---|---|---|---|---|---|---|---|---|---|---|---|---|---|---|
| 10 | 25383388 | 0.92 | 0.96 | rs72798044 | A | T | 0.08 | 0.07 | 0.11 | 0.09 | | | 4 tissues | ESDR | | 4 altered motifs | | | | GPR158 | intronic |
| 10 | 25388278 | 0.92 | 0.96 | rs2887084 | C | T | 0.08 | 0.07 | 0.11 | 0.09 | | | | | | NRSF | | | | GPR158 | intronic |
| 10 | 25392661 | 0.92 | 0.96 | rs2009148 | A | C | 0.09 | 0.07 | 0.11 | 0.09 | | | | | | 8 altered motifs | | | | GPR158 | intronic |

Query SNP: rs7379705 and variants with $r^2$ >= 0.8

| chr | pos (hg38) | LD (r²) | LD (D') | variant | Ref | Alt | AFR freq | AMR freq | ASN freq | EUR freq | SiPhy cons | Promoter histone marks | Enhancer histone marks | DNAse | Proteins bound | Motifs changed | NHGRI/EBI GWAS hits | GRASP QTL hits | Selected eQTL hits | GENCODE genes | dbSNP func annot |
|---|---|---|---|---|---|---|---|---|---|---|---|---|---|---|---|---|---|---|---|---|---|
| 5 | 2141442 | 1 | 1 | rs7379705 | C | T | 0.68 | 0.41 | 0.23 | 0.42 | | | | | | E2F,Lmo2-complex | | | | 43kb 5' of Y_RNA | |

Query SNP: rs1453012 and variants with $r^2$ >= 0.8

| chr | pos (hg38) | LD (r²) | LD (D') | variant | Ref | Alt | AFR freq | AMR freq | ASN freq | EUR freq | SiPhy cons | Promoter histone marks | Enhancer histone marks | DNAse | Proteins bound | Motifs changed | NHGRI/EBI GWAS hits | GRASP QTL hits | Selected eQTL hits | GENCODE genes | dbSNP func annot |
|---|---|---|---|---|---|---|---|---|---|---|---|---|---|---|---|---|---|---|---|---|---|
| 5 | 114874581 | 1 | 1 | rs1453012 | G | C | 0.84 | 0.65 | 0.60 | 0.51 | | | | | | RXRA,Sox | | | | 157kb 3' of RP11-438C19.2 | |
| 5 | 114877069 | 0.88 | 0.98 | rs4577683 | A | T | 0.73 | 0.64 | 0.60 | 0.51 | | | | | | Myc | | | | 154kb 3' of RP11-438C19.2 | |
| 5 | 114878960 | 0.93 | 1 | rs10063141 | A | T | 0.73 | 0.64 | 0.60 | 0.51 | | | | | | 4 altered motifs | | | | 152kb 3' of RP11-438C19.2 | |
| 5 | 114879186 | 0.93 | 1 | rs13355555 | T | C | 0.73 | 0.64 | 0.60 | 0.51 | | | | | | 13 altered motifs | | | | 152kb 3' of RP11-438C19.2 | |
| 5 | 114882338 | 0.8 | 1 | rs4582267 | C | T | 0.71 | 0.60 | 0.56 | 0.46 | | | | | | Sox | | | | 149kb 3' of RP11-438C19.2 | |
| 5 | 114886713 | 0.93 | 1 | rs1119461 | A | T | 0.75 | 0.64 | 0.60 | 0.51 | | | | | | Sin3Ak-20,Sox | | | | 145kb 3' of RP11-438C19.2 | |
| 5 | 114888764 | 0.8 | 1 | rs1947383 | T | C | 0.71 | 0.60 | 0.56 | 0.46 | | | | | | 7 altered motifs | | | | 143kb 3' of RP11-438C19.2 | |
| 5 | 114890748 | 0.96 | 1 | rs9326934 | G | A | 0.85 | 0.64 | 0.60 | 0.51 | | | | | | 6 altered motifs | | | | 141kb 3' of RP11-438C19.2 | |
| 5 | 114892767 | 0.88 | 1 | rs10055168 | C | T | 0.72 | 0.62 | 0.58 | 0.50 | | | | | | 6 altered motifs | | | | 139kb 3' of RP11-438C19.2 | |
| 5 | 114902588 | 0.84 | 1 | rs12521180 | T | C | 0.70 | 0.61 | 0.56 | 0.51 | | | | | | AhR::Arnt,Arnt | | | | 129kb 3' of RP11-438C19.2 | |
| 5 | 114902794 | 0.83 | 1 | rs10073239 | T | G | 0.71 | 0.61 | 0.56 | 0.51 | | | | | | 4 altered motifs | | | | 128kb 3' of RP11-438C19.2 | |
| 5 | 114905113 | 0.83 | 1 | rs6594855 | G | C | 0.71 | 0.61 | 0.56 | 0.51 | | | | | | HNF4,Irf | | | | 126kb 3' of RP11-438C19.2 | |
| 5 | 114906049 | 0.92 | 1 | rs2416384 | C | A | 0.75 | 0.63 | 0.60 | 0.51 | | | | | | Pou2f2 | | | | 125kb 3' of RP11-438C19.2 | |
| 5 | 114906830 | 0.83 | 1 | rs62379442 | T | C | 0.71 | 0.61 | 0.56 | 0.51 | | | | | | 5 altered motifs | | | | 124kb 3' of RP11-438C19.2 | |
| 5 | 114908753 | 0.93 | 1 | rs7716212 | C | T | 0.75 | 0.64 | 0.60 | 0.51 | | | | | | Mxi1 | | | | 123kb 3' of RP11-438C19.2 | |
| 5 | 114910558 | 0.93 | 1 | rs4318770 | C | T | 0.75 | 0.64 | 0.60 | 0.51 | | | | | | | | | | 121kb 3' of RP11-438C19.2 | |
| 5 | 114910999 | 0.84 | 1 | rs6594856 | T | G | 0.71 | 0.61 | 0.56 | 0.51 | | | | | MAFF,MAFK | CTCF,NF-I,TATA | | | | 120kb 3' of RP11-438C19.2 | |
| 5 | 114911487 | 0.84 | 1 | rs6862719 | C | T | 0.69 | 0.61 | 0.55 | 0.49 | | | | | | Foxp1 | | | | 120kb 3' of RP11-438C19.2 | |
| 5 | 114912530 | 0.93 | 1 | rs4374751 | C | T | 0.75 | 0.64 | 0.60 | 0.51 | | | | | | AP-4,GR | | | | 119kb 3' of RP11-438C19.2 | |
| 5 | 114912610 | 0.93 | 1 | rs6889905 | T | C | 0.75 | 0.64 | 0.60 | 0.51 | | | | | | | | | | 119kb 3' of RP11-438C19.2 | |
| 5 | 114916590 | 0.88 | 0.99 | rs2416388 | G | A | 0.70 | 0.63 | 0.59 | 0.51 | | | | | | Fox,Hdx,Nanog | | | | 115kb 3' of RP11-438C19.2 | |
| 5 | 114922494 | 0.93 | 1 | rs12517743 | T | C | 0.85 | 0.64 | 0.60 | 0.51 | | | | | | Pax-6,RFX5 | | | | 109kb 3' of RP11-438C19.2 | |
| 5 | 114923359 | 0.8 | 1 | rs10057114 | G | C | 0.66 | 0.60 | 0.60 | 0.48 | | | | | | Maf | | | | 108kb 3' of RP11-438C19.2 | |
| 5 | 114928471 | 0.93 | 1 | rs11241296 | C | G | 0.82 | 0.64 | 0.60 | 0.51 | | | | | | 8 altered motifs | | | | 103kb 3' of RP11-438C19.2 | |
| 5 | 114936062 | 0.82 | 1 | rs4645325 | C | G | 0.70 | 0.60 | 0.42 | 0.48 | | | | | | CTCF,Nanog,VDR | | | | 95kb 3' of RP11-438C19.2 | |
| 5 | 114941993 | 0.93 | -0.99 | rs4276395 | C | T | 0.16 | 0.36 | 0.53 | 0.52 | | | | | | Mef2,Sox | | | | 89kb 3' of RP11-438C19.2 | |
| 5 | 114943082 | 0.88 | -0.98 | rs921596 | T | C | 0.26 | 0.36 | 0.53 | 0.51 | | | | | | Irf,PRDM1 | | | | 88kb 3' of RP11-438C19.2 | |
| 5 | 114945459 | 0.83 | -0.96 | rs10059249 | T | C | 0.26 | 0.37 | 0.53 | 0.52 | | | | | | p300 | | | | 86kb 3' of RP11-438C19.2 | |

Query SNP: rs3750164 and variants with $r^2$ >= 0.8

| chr | pos (hg38) | LD (r²) | LD (D') | variant | Ref | Alt | AFR freq | AMR freq | ASN freq | EUR freq | SiPhy cons | Promoter histone marks | Enhancer histone marks | DNAse | Proteins bound | Motifs changed | NHGRI/EBI GWAS hits | GRASP QTL hits | Selected eQTL hits | GENCODE genes | dbSNP func annot |
|---|---|---|---|---|---|---|---|---|---|---|---|---|---|---|---|---|---|---|---|---|---|
| 7 | 47258475 | 0.84 | 0.97 | rs7794357 | T | C | 0.78 | 0.77 | 0.71 | 0.67 | | | | | | Mef2,ZBTB33 | | | 2 hits | 17kb 3' of TNS3 | |
| 7 | 47260357 | 0.83 | 0.95 | rs4720587 | T | C | 0.64 | 0.77 | 0.67 | 0.66 | | ESC | | LNG | | Hoxa5,NRSF,PPAR | | | | 15kb 3' of TNS3 | |
| 7 | 47261943 | 0.87 | 0.98 | rs4724555 | C | G | 0.66 | 0.77 | 0.73 | 0.68 | | | | | | SREBP | | | | 13kb 3' of TNS3 | |
| 7 | 47262526 | 0.89 | 1 | rs12540734 | G | A | 0.80 | 0.77 | 0.73 | 0.67 | | | | | | LBP-1,Zbtb3 | | | | 13kb 3' of TNS3 | |
| 7 | 47264124 | 0.87 | 0.98 | rs10951899 | A | G | 0.79 | 0.77 | 0.73 | 0.68 | | | | ESDR,BRN | | ERalpha-a,Osf2,SF1 | | | | 11kb 3' of TNS3 | |
| 7 | 47264511 | 0.89 | 1 | rs11765461 | T | C | 0.80 | 0.77 | 0.73 | 0.68 | | | STRM | ESC,LNG | | NRSF | | | | 11kb 3' of TNS3 | |
| 7 | 47266371 | 0.99 | 1 | rs4724556 | A | C | 0.79 | 0.75 | 0.71 | 0.67 | | | 14 tissues | 37 tissues | 5 bound proteins | 4 altered motifs | | 3 hits | | 8.8kb 3' of TNS3 | |
| 7 | 47268643 | 0.99 | 1 | rs10951901 | C | T | 0.79 | 0.75 | 0.71 | 0.68 | | | | | | 4 altered motifs | | 3 hits | | 6.5kb 3' of TNS3 | |
| 7 | 47271185 | 0.99 | 1 | rs4720589 | C | A | 0.79 | 0.75 | 0.71 | 0.68 | | | | | | Ascl2,GR,Myf | | | | 4kb 3' of TNS3 | |
| 7 | 47272455 | 0.99 | 1 | rs7794685 | G | C | 0.79 | 0.75 | 0.71 | 0.68 | | | | | | Evi-1,Irx,Pbx-1 | | | | 2.7kb 3' of TNS3 | |
| 7 | 47272572 | 0.99 | 1 | rs4724559 | G | C | 0.79 | 0.75 | 0.71 | 0.68 | | | | | | Sox | | | | 2.6kb 3' of TNS3 | |
| 7 | 47277448 | 0.99 | 1 | rs3750161 | C | T | 0.69 | 0.75 | 0.71 | 0.68 | | | | | | Dobox4,XBP-1 | | | | TNS3 | 3'-UTR |
| 7 | 47280125 | 0.99 | 1 | rs4720590 | C | T | 0.80 | 0.75 | 0.65 | 0.66 | | | | | | | | | | TNS3 | intronic |
| 7 | 47281110 | 0.99 | 1 | rs6959292 | T | C | 0.70 | 0.75 | 0.70 | 0.68 | | | ESDR, PLCNT | ESDR,PLCNT | | Myf,NRSF | | | | TNS3 | intronic |
| 7 | 47281452 | 0.9 | 1 | rs12702333 | A | G | 0.43 | 0.73 | 0.71 | 0.67 | | | ESDR, PLCNT | PLCNT,MUS,BRN | | HNF4 | | 3 hits | | TNS3 | intronic |

| | | | | | | | | | | | | | | | | | | | | | |
|---|---|---|---|---|---|---|---|---|---|---|---|---|---|---|---|---|---|---|---|---|---|
| 7 | 47282686 | 0.89 | 1 | rs4724560 | C | T | 0.42 | 0.73 | 0.65 | 0.66 | | | 6 tissues | BRN | ERALPHA_A | 5 altered motifs | | | | TNS3 | intronic |
| 7 | 47283556 | 0.99 | 1 | rs3829000 | A | G | 0.70 | 0.75 | 0.71 | 0.68 | | | 7 tissues | 14 tissues | FOSL2,JUND | Bcl6b,HMG-IY,Mrg1::Hoxa9 | | | | TNS3 | intronic |
| 7 | 47283858 | 1 | 1 | rs3750164 | A | G | 0.70 | 0.75 | 0.71 | 0.67 | | | 6 tissues | 7 tissues | | | | | | TNS3 | synonymous |
| 7 | 47285226 | 0.99 | 1 | rs6942872 | T | G | 0.81 | 0.75 | 0.71 | 0.68 | | | FAT, HRT, LNG | | | 6 altered motifs | | | | TNS3 | intronic |
| 7 | 47288786 | 0.97 | 1 | rs2347785 | G | T | 0.68 | 0.75 | 0.65 | 0.66 | | | 4 tissues | MUS,MUS | | | | | | TNS3 | intronic |
| 7 | 47293223 | 0.99 | 1 | rs3750165 | C | T | 0.66 | 0.75 | 0.65 | 0.66 | | | 17 tissues | 12 tissues | FOXA2,TCF4 | Ets,Mef2 | | | | TNS3 | intronic |

Query SNP: rs10464982 and variants with $r^2$ >= 0.8

| chr | pos (hg38) | LD (r²) | LD (D') | variant | Ref | Alt | AFR freq | AMR freq | ASN freq | EUR freq | SiPhy cons | Promoter histone marks | Enhancer histone marks | DNAse | Proteins bound | Motifs changed | NHGRI/EBI GWAS hits | GRASP QTL hits | Selected eQTL hits | GENCODE genes | dbSNP func annot |
|---|---|---|---|---|---|---|---|---|---|---|---|---|---|---|---|---|---|---|---|---|---|
| 8 | 140249888 | 0.84 | 0.94 | rs4736157 | G | A | 0.40 | 0.44 | 0.74 | 0.37 | | | | | | AP-1 | | | | TRAPPC9 | intronic |
| 8 | 140253239 | 0.82 | 0.95 | rs12334515 | C | A | 0.42 | 0.45 | 0.73 | 0.37 | | | ESDR, THYM, BLD | | | Mef2,TEF | | | | TRAPPC9 | intronic |
| 8 | 140270314 | 1 | 1 | rs10464982 | C | T | 0.41 | 0.42 | 0.73 | 0.35 | | | | | | | | | | TRAPPC9 | intronic |

Query SNP: rs6981562 and variants with $r^2$ >= 0.8

| chr | pos (hg38) | LD (r²) | LD (D') | variant | Ref | Alt | AFR freq | AMR freq | ASN freq | EUR freq | SiPhy cons | Promoter histone marks | Enhancer histone marks | DNAse | Proteins bound | Motifs changed | NHGRI/EBI GWAS hits | GRASP QTL hits | Selected eQTL hits | GENCODE genes | dbSNP func annot |
|---|---|---|---|---|---|---|---|---|---|---|---|---|---|---|---|---|---|---|---|---|---|
| 8 | 29887002 | 1 | 1 | rs6981562 | T | C | 0.61 | 0.18 | 0.63 | 0.27 | | | 12 tissues | | | | | | | 15kb 3' of RP11-94H18.2 | |

Query SNP: rs10201163 and variants with $r^2$ >= 0.8

| chr | pos (hg38) | LD (r²) | LD (D') | variant | Ref | Alt | AFR freq | AMR freq | ASN freq | EUR freq | SiPhy cons | Promoter histone marks | Enhancer histone marks | DNAse | Proteins bound | Motifs changed | NHGRI/EBI GWAS hits | GRASP QTL hits | Selected eQTL hits | GENCODE genes | dbSNP func annot |
|---|---|---|---|---|---|---|---|---|---|---|---|---|---|---|---|---|---|---|---|---|---|
| 2 | 204301769 | 0.97 | 1 | rs13397660 | C | T | 0.22 | 0.10 | 0.00 | 0.10 | | | | | | | | | | 67kb 5' of AC009498.1 | |
| 2 | 204304512 | 1 | 1 | rs201303382 | A | AAC | 0.22 | 0.09 | 0.00 | 0.10 | | | BLD, BRN | GI | | 10 altered motifs | | | | 70kb 5' of AC009498.1 | |
| 2 | 204304513 | 1 | 1 | rs200465392 | TTG | T | 0.22 | 0.09 | 0.00 | 0.10 | | | BLD, BRN | GI | | 7 altered motifs | | | | 70kb 5' of AC009498.1 | |
| 2 | 204304514 | 1 | 1 | rs67231893 | TGTCC | T | 0.20 | 0.09 | 0.00 | 0.10 | | | BLD, BRN | GI | | 7 altered motifs | | | | 70kb 5' of AC009498.1 | |
| 2 | 204304616 | 1 | 1 | rs10201163 | T | C | 0.22 | 0.09 | 0.00 | 0.10 | | | | | | Nkx2 | | 3 hits | | 70kb 5' of AC009498.1 | |
| 2 | 204312320 | 1 | 1 | rs57830749 | C | T | 0.24 | 0.09 | 0.00 | 0.09 | | | 5 tissues | 11 tissues | | | | | | 77kb 5' of AC009498.1 | |
| 2 | 204312975 | 1 | 1 | rs7603542 | C | T | 0.25 | 0.09 | 0.00 | 0.10 | | | 6 tissues | ADRL | | NRSF | | | | 78kb 5' of AC009498.1 | |
| 2 | 204316470 | 0.94 | 1 | rs6435229 | C | G | 0.24 | 0.09 | 0.00 | 0.10 | | | BRN | | | Foxq1,Hoxa13,PLZF | | 1 hit | | 81kb 5' of AC009498.1 | |
| 2 | 204321806 | 0.91 | 1 | rs11904155 | C | T | 0.30 | 0.10 | 0.00 | 0.09 | | | 10 tissues | ESDR,MUS,VAS | | VDR | | 1 hit | | 87kb 5' of AC009498.1 | |

Query SNP: rs324884 and variants with $r^2$ >= 0.8

| chr | pos (hg38) | LD (r²) | LD (D') | variant | Ref | Alt | AFR freq | AMR freq | ASN freq | EUR freq | SiPhy cons | Promoter histone marks | Enhancer histone marks | DNAse | Proteins bound | Motifs changed | NHGRI/EBI GWAS hits | GRASP QTL hits | Selected eQTL hits | GENCODE genes | dbSNP func annot |
|---|---|---|---|---|---|---|---|---|---|---|---|---|---|---|---|---|---|---|---|---|---|
| 5 | 88560042 | 0.94 | 0.98 | rs7710776 | C | T | 0.92 | 0.82 | 0.75 | 0.76 | | | | | | 4 altered motifs | | | | LINC00461 | intronic |
| 5 | 88560241 | 0.88 | 0.96 | rs10071077 | A | G | 0.92 | 0.81 | 0.74 | 0.76 | | | | | | AIRE,Bcl6b,Sp100 | | | | LINC00461 | intronic |
| 5 | 88565282 | 1 | 1 | rs324884 | G | A | 0.92 | 0.82 | 0.75 | 0.76 | | | | | | CCNT2,Cart1 | | | | LINC00461 | intronic |
| 5 | 88574368 | 0.82 | -1 | rs13168144 | T | C | 0.00 | 0.15 | 0.25 | 0.21 | | | | | | 5 altered motifs | | | 1 hit | LINC00461 | intronic |
| 5 | 88583650 | 0.96 | 0.98 | rs2438466 | A | C | 0.92 | 0.82 | 0.75 | 0.76 | | | | MUS,MUS,MUS | | ERalpha-a,Esr2,RAR | | | | LINC00461 | intronic |
| 5 | 88598828 | 0.83 | 0.98 | rs2112424 | A | G | 0.88 | 0.80 | 0.28 | 0.74 | | | | | | 5 altered motifs | | | | LINC00461 | intronic |
| 5 | 88600660 | 0.82 | 0.98 | rs167789 | A | G | 0.89 | 0.80 | 0.74 | 0.75 | | | | | SETDB1 | 5 altered motifs | | | | LINC00461 | intronic |
| 5 | 88618709 | 0.8 | 0.96 | rs324898 | C | T | 0.85 | 0.80 | 0.28 | 0.74 | | | | | | RXRA,STAT | | | | LINC00461 | intronic |

Query SNP: rs10733392 and variants with $r^2$ >= 0.8

| chr | pos (hg38) | LD (r²) | LD (D') | variant | Ref | Alt | AFR freq | AMR freq | ASN freq | EUR freq | SiPhy cons | Promoter histone marks | Enhancer histone marks | DNAse | Proteins bound | Motifs changed | NHGRI/EBI GWAS hits | GRASP QTL hits | Selected eQTL hits | GENCODE genes | dbSNP func annot |
|---|---|---|---|---|---|---|---|---|---|---|---|---|---|---|---|---|---|---|---|---|---|
| 9 | 23694190 | 1 | 1 | rs10733392 | C | T | 0.92 | 0.69 | 0.48 | 0.74 | | | | | | Ets,Hoxa3,Pou2f2 | | | | RP11-315I14.3 | intronic |

Query SNP: rs17366040 and variants with $r^2$ >= 0.8

| chr | pos (hg38) | LD (r²) | LD (D') | variant | Ref | Alt | AFR freq | AMR freq | ASN freq | EUR freq | SiPhy cons | Promoter histone marks | Enhancer histone marks | DNAse | Proteins bound | Motifs changed | NHGRI/EBI GWAS hits | GRASP QTL hits | Selected eQTL hits | GENCODE genes | dbSNP func annot |
|---|---|---|---|---|---|---|---|---|---|---|---|---|---|---|---|---|---|---|---|---|---|
| 8 | 27158809 | 1 | 1 | rs17366040 | C | T | 0.13 | 0.24 | 0.10 | 0.33 | | | | | | | | | | 11kb 3' of AC090150.1 | |
| 8 | 27160516 | 0.98 | 1 | rs62503689 | C | T | 0.13 | 0.23 | 0.10 | 0.33 | | | | | | Dobox4 | | | | 13kb 3' of AC090150.1 | |
| 8 | 27162187 | 0.98 | 1 | rs73237489 | T | C | 0.13 | 0.23 | 0.10 | 0.33 | | | | | | 8 altered motifs | | | | 14kb 3' of AC090150.1 | |
| 8 | 27163490 | 0.98 | 1 | rs17438203 | G | A | 0.11 | 0.23 | 0.10 | 0.33 | | | | | | Ets,Tel2 | | | | 16kb 3' of AC090150.1 | |
| 8 | 27169912 | 0.97 | 1 | rs11990708 | A | G | 0.12 | 0.23 | 0.10 | 0.33 | | | FAT, STRM | | | 5 altered motifs | | | | 14kb 5' of RP11-521M14.1 | |

Query SNP: rs6089240 and variants with $r^2$ >= 0.8

| chr | pos (hg38) | LD (r²) | LD (D') | variant | Ref | Alt | AFR freq | AMR freq | ASN freq | EUR freq | SiPhy cons | Promoter histone marks | Enhancer histone marks | DNAse | Proteins bound | Motifs changed | NHGRI/EBI GWAS hits | GRASP QTL hits | Selected eQTL hits | GENCODE genes | dbSNP func annot |
|---|---|---|---|---|---|---|---|---|---|---|---|---|---|---|---|---|---|---|---|---|---|
| 20 | 61577204 | 1 | 1 | rs6089240 | A | G | 0.08 | 0.52 | 0.47 | 0.55 | | | | | | 7 altered motifs | | | | CDH4 | intronic |

Query SNP: rs2059843 and variants with $r^2$ >= 0.8

| chr | pos (hg38) | LD (r²) | LD (D') | variant | Ref | Alt | AFR freq | AMR freq | ASN freq | EUR freq | SiPhy cons | Promoter histone marks | Enhancer histone marks | DNAse | Proteins bound | Motifs changed | NHGRI/EBI GWAS hits | GRASP QTL hits | Selected eQTL hits | GENCODE genes | dbSNP func annot |
|---|---|---|---|---|---|---|---|---|---|---|---|---|---|---|---|---|---|---|---|---|---|
| 5 | 7423821 | 0.97 | 0.99 | rs7732703 | C | A | 0.91 | 0.70 | 0.49 | 0.81 | | | 4 tissues | | | 8 altered motifs | | | | ADCY2 | intronic |
| 5 | 7424928 | 0.97 | 0.99 | rs1490783 | T | C | 0.90 | 0.70 | 0.49 | 0.81 | | MUS | 5 tissues | | | 5 altered motifs | | | | ADCY2 | intronic |
| 5 | 7427176 | 0.94 | 0.99 | rs7710510 | T | C | 0.71 | 0.69 | 0.49 | 0.81 | | | ESC, IPSC, MUS | | | 4 altered motifs | | | | ADCY2 | intronic |
| 5 | 7428440 | 0.93 | 0.99 | rs7712168 | A | G | 0.91 | 0.71 | 0.49 | 0.82 | | | 4 tissues | | | 8 altered motifs | | | | ADCY2 | intronic |
| 5 | 7431081 | 0.92 | 0.99 | rs2397354 | A | T | 0.62 | 0.69 | 0.49 | 0.81 | | | | | | 8 altered motifs | | | 1 hit | ADCY2 | intronic |
| 5 | 7432274 | 0.97 | 0.99 | rs1490784 | T | C | 0.91 | 0.70 | 0.49 | 0.81 | | | IPSC, OVRY, MUS | | | Ets,GATA | | | | ADCY2 | intronic |
| 5 | 7432550 | 0.93 | 0.99 | rs7737816 | T | C | 0.91 | 0.71 | 0.49 | 0.82 | | | 4 tissues | | | Gm397 | | | | ADCY2 | intronic |
| 5 | 7433405 | 0.93 | 0.97 | rs6555464 | A | G | 0.88 | 0.70 | 0.48 | 0.81 | | | BRN, MUS | | | SRF | | | | ADCY2 | intronic |
| 5 | 7434504 | 0.96 | 1 | rs6555465 | A | G | 0.91 | 0.71 | 0.49 | 0.82 | | | ESDR, BRN, MUS | | | | | | | ADCY2 | intronic |
| 5 | 7436628 | 0.96 | 1 | rs1864072 | C | G | 0.91 | 0.71 | 0.49 | 0.82 | | MUS | MUS | PLCNT,MUS | USF1,CTCF | | | | | ADCY2 | intronic |
| 5 | 7436799 | 0.85 | 0.96 | rs1864071 | T | C | 0.68 | 0.69 | 0.49 | 0.82 | | MUS | BRST, SKIN, MUS | 4 tissues | USF1 | | | 1 hit | | ADCY2 | intronic |
| 5 | 7437390 | 0.96 | 1 | rs7712495 | C | G | 0.89 | 0.71 | 0.49 | 0.82 | | MUS | BRST, MUS, PLCNT | | | Arnt | | | | ADCY2 | intronic |
| 5 | 7438542 | 0.96 | 1 | rs4702466 | A | C | 0.91 | 0.71 | 0.49 | 0.82 | | | MUS | | | 5 altered motifs | | | | ADCY2 | intronic |
| 5 | 7439913 | 0.91 | 1 | rs35755966 | TC | T | 0.68 | 0.68 | 0.48 | 0.81 | | | | | | VDR | | | | ADCY2 | intronic |
| 5 | 7439971 | 0.91 | 1 | rs6883259 | G | A | 0.68 | 0.68 | 0.49 | 0.81 | | | | | | Fox,PLZF | | | | ADCY2 | intronic |
| 5 | 7441216 | 1 | 1 | rs6893414 | G | A | 0.90 | 0.70 | 0.49 | 0.81 | | | MUS | | | Myb,Myf,NRSF | | | | ADCY2 | intronic |
| 5 | 7441808 | 1 | 1 | rs6555466 | A | G | 0.90 | 0.70 | 0.49 | 0.82 | | | MUS | | | SREBP | | | | ADCY2 | intronic |
| 5 | 7442932 | 0.97 | 1 | rs6859782 | G | T | 0.92 | 0.71 | 0.49 | 0.82 | | | MUS | | | | | | | ADCY2 | intronic |
| 5 | 7443797 | 1 | 1 | rs2059843 | A | C | 0.90 | 0.70 | 0.49 | 0.82 | | | | | | | | | | ADCY2 | intronic |
| 5 | 7444394 | 0.92 | 1 | rs2130751 | T | C | 0.84 | 0.69 | 0.47 | 0.79 | | | | | | 4 altered motifs | | | | ADCY2 | intronic |
| 5 | 7444401 | 0.83 | 1 | rs6871528 | C | T | 0.65 | 0.66 | 0.47 | 0.79 | | | | | | E2A,Pax-4 | | | 1 hit | ADCY2 | intronic |
| 5 | 7444410 | 0.91 | 1 | rs1963032 | G | C | 0.87 | 0.68 | 0.48 | 0.79 | | | | | | 11 altered motifs | | | | ADCY2 | intronic |
| 5 | 7444737 | 1 | 1 | rs1025292 | T | G | 0.90 | 0.70 | 0.49 | 0.82 | | | | | | HDAC2 | | | | ADCY2 | intronic |
| 5 | 7445253 | 0.97 | 1 | rs7715966 | G | A | 0.92 | 0.71 | 0.49 | 0.82 | | | 4 tissues | MUS | | 5 altered motifs | | | | ADCY2 | intronic |
| 5 | 7445423 | 1 | 1 | rs7703114 | T | C | 0.89 | 0.70 | 0.49 | 0.82 | | | ESDR, MUS | | | GR,Zic | | | | ADCY2 | intronic |
| 5 | 7445704 | 1 | 1 | rs10057329 | C | A | 0.90 | 0.70 | 0.49 | 0.81 | | | | | | CHOP::CEBPalpha,Maf,NF-E2 | | | | ADCY2 | intronic |
| 5 | 7446776 | 1 | 1 | rs766643 | C | T | 0.90 | 0.70 | 0.49 | 0.81 | | | BRN | | | 6 altered motifs | | | | ADCY2 | intronic |

Query SNP: rs6912831 and variants with $r^2$ >= 0.8

| chr | pos (hg38) | LD (r²) | LD (D') | variant | Ref | Alt | AFR freq | AMR freq | ASN freq | EUR freq | SiPhy cons | Promoter histone marks | Enhancer histone marks | DNAse | Proteins bound | Motifs changed | NHGRI/EBI GWAS hits | GRASP QTL hits | Selected eQTL hits | GENCODE genes | dbSNP func annot |
|---|---|---|---|---|---|---|---|---|---|---|---|---|---|---|---|---|---|---|---|---|---|
| 6 | 139242777 | 0.81 | 0.9 | rs9495391 | C | G | 0.83 | 0.36 | 0.23 | 0.35 | | | 5 tissues | | | | | | 3 hits | TXLNB | missense |
| 6 | 139246271 | 0.94 | 1 | rs7772534 | C | A | 0.83 | 0.34 | 0.15 | 0.36 | | | | | | 9 altered motifs | | | 3 hits | TXLNB | intronic |
| 6 | 139246531 | 0.95 | 1 | rs6570320 | A | G | 0.83 | 0.34 | 0.15 | 0.37 | | | | | | 5 altered motifs | | | 3 hits | TXLNB | intronic |
| 6 | 139249159 | 1 | 1 | rs6912831 | A | C | 0.90 | 0.35 | 0.17 | 0.37 | | | FAT, BLD, GI | 5 tissues | | 4 altered motifs | | | 3 hits | TXLNB | intronic |
| 6 | 139250003 | 1 | 1 | rs9495394 | G | A | 0.87 | 0.35 | 0.17 | 0.36 | | | 7 tissues | | | 11 altered motifs | | | 3 hits | TXLNB | intronic |
| 6 | 139254715 | 0.99 | 1 | rs4534002 | C | T | 0.87 | 0.35 | 0.17 | 0.36 | | | BLD, GI | LNG,OVRY | | 5 altered motifs | | | 3 hits | TXLNB | intronic |
| 6 | 139254878 | 0.99 | 1 | rs4598096 | A | G | 0.88 | 0.35 | 0.17 | 0.36 | | | BLD, MUS, GI | | | 5 altered motifs | | | 3 hits | TXLNB | intronic |
| 6 | 139258622 | 0.94 | 0.98 | rs6940670 | C | T | 0.85 | 0.35 | 0.16 | 0.36 | | | BLD | | | 5 altered motifs | | | 3 hits | TXLNB | intronic |
| 6 | 139262853 | 0.94 | 0.98 | rs6922150 | A | T | 0.86 | 0.35 | 0.17 | 0.37 | | BLD | 7 tissues | 11 tissues | MEF2A,MEF2C,PU1 | 5 altered motifs | | | 4 hits | TXLNB | intronic |
| 6 | 139263681 | 0.94 | 0.98 | rs5880422 | G | 7-mer | 0.88 | 0.35 | 0.17 | 0.37 | | | BLD, BRN | BRN,BRN | | 5 altered motifs | | | 4 hits | TXLNB | intronic |

Query SNP: rs6903827 and variants with $r^2$ >= 0.8

| chr | pos (hg38) | LD (r²) | LD (D') | variant | Ref | Alt | AFR freq | AMR freq | ASN freq | EUR freq | SiPhy cons | Promoter histone marks | Enhancer histone marks | DNAse | Proteins bound | Motifs changed | NHGRI/EBI GWAS hits | GRASP QTL hits | Selected eQTL hits | GENCODE genes | dbSNP func annot |
|---|---|---|---|---|---|---|---|---|---|---|---|---|---|---|---|---|---|---|---|---|---|
| 6 | 149064375 | 0.82 | 0.92 | rs6932386 | G | A | 0.18 | 0.27 | 0.24 | 0.26 | | | 13 tissues | BLD,MUS,MUS | | 4 altered motifs | | | | UST | intronic |
| 6 | 149065820 | 0.82 | 0.92 | rs10872630 | G | A | 0.18 | 0.27 | 0.24 | 0.26 | | | | 4 tissues | | RXRA | | | | UST | intronic |
| 6 | 149067559 | 0.82 | 0.92 | rs10872631 | G | A | 0.27 | 0.27 | 0.24 | 0.26 | | | | BLD | | CAC-binding-protein,Zfx | | | 1 hit | UST | intronic |
| 6 | 149068617 | 0.82 | 0.92 | rs2879862 | G | A | 0.27 | 0.27 | 0.24 | 0.26 | | | | | | 12 altered motifs | | | 1 hit | UST | intronic |
| 6 | 149070055 | 0.82 | 0.92 | rs10872632 | C | T | 0.18 | 0.27 | 0.33 | 0.27 | | | | | | Foxa | | | | UST | intronic |
| 6 | 149071336 | 0.82 | 0.92 | rs17732007 | A | G | 0.18 | 0.27 | 0.24 | 0.26 | | | | | | Ik-1,Mef2,STAT | | 1 hit | | UST | intronic |
| 6 | 149071697 | 0.82 | 0.92 | rs62426156 | G | A | 0.18 | 0.27 | 0.24 | 0.26 | | | | | | Foxp1,HDAC2 | | | | UST | intronic |
| 6 | 149072530 | 0.84 | 0.92 | rs4897077 | G | A | 0.18 | 0.28 | 0.24 | 0.26 | | | | | | Crx,GATA | | | | UST | intronic |
| 6 | 149073231 | 0.83 | 0.93 | rs58644381 | G | A | 0.18 | 0.27 | 0.24 | 0.25 | | | | | | | | | | UST | intronic |
| 6 | 149076607 | 0.9 | 0.97 | rs11330840 | AT | A | 0.31 | 0.29 | 0.23 | 0.27 | | | 13 tissues | 6 tissues | 4 bound proteins | Hoxa9 | | | | UST | 3'-UTR |
| 6 | 149077113 | 0.91 | 0.97 | rs6939103 | T | G | 0.29 | 0.29 | 0.24 | 0.26 | | | 8 tissues | | | 8 altered motifs | | | | 122bp 3' of UST | |
| 6 | 149082333 | 1 | 1 | rs6903827 | A | C | 0.33 | 0.28 | 0.24 | 0.27 | | | 5 tissues | 27 tissues | 9 bound proteins | 9 altered motifs | | | | 5.3kb 3' of UST | |
| 6 | 149083891 | 0.86 | 0.93 | rs7772696 | T | C | 0.32 | 0.28 | 0.24 | 0.26 | | | BRST, BLD | BLD | | Elf3,Nanog,Pou5f1 | | | | 6.9kb 3' of UST | |
| 6 | 149084346 | 0.84 | 0.92 | rs4897078 | C | G | 0.32 | 0.28 | 0.24 | 0.26 | | | BRST, BLD, PANC | ESDR,PLCNT,BLD | | | | | | 7.4kb 3' of UST | |
| 6 | 149090029 | 0.8 | 0.92 | rs11755147 | G | A | 0.34 | 0.27 | 0.24 | 0.24 | | | BLD | | | Irf,STAT,TATA | | | | 13kb 3' of UST | |

Query SNP: rs10230715 and variants with $r^2$ >= 0.8

| chr | pos (hg38) | LD (r²) | LD (D') | variant | Ref | Alt | AFR freq | AMR freq | ASN freq | EUR freq | SiPhy cons | Promoter histone marks | Enhancer histone marks | DNAse | Proteins bound | Motifs changed | NHGRI/EBI GWAS hits | GRASP QTL hits | Selected eQTL hits | GENCODE genes | dbSNP func annot |
|---|---|---|---|---|---|---|---|---|---|---|---|---|---|---|---|---|---|---|---|---|---|
| 7 | 42145719 | 0.92 | 0.97 | rs3823727 | G | A,C,T | 0.48 | 0.47 | 0.29 | 0.53 | | SKIN | 7 tissues | THYM | | | | | | GLI3 | intronic |
| 7 | 42149257 | 1 | 1 | rs10230715 | G | A | 0.50 | 0.47 | 0.29 | 0.53 | | | | BRN,PLCNT | | Nanog,Pax-4 | | | | GLI3 | intronic |

Query SNP: rs4054037 and variants with $r^2$ >= 0.8

| chr | pos (hg38) | LD (r²) | LD (D') | variant | Ref | Alt | AFR freq | AMR freq | ASN freq | EUR freq | SiPhy cons | Promoter histone marks | Enhancer histone marks | DNAse | Proteins bound | Motifs changed | NHGRI/EBI GWAS hits | GRASP QTL hits | Selected eQTL hits | GENCODE genes | dbSNP func annot |
|---|---|---|---|---|---|---|---|---|---|---|---|---|---|---|---|---|---|---|---|---|---|

| | | | | | | | | | | | | | | | | | | | | |
|---|---|---|---|---|---|---|---|---|---|---|---|---|---|---|---|---|---|---|---|---|
| 4 | 24359871 | 0.81 | 1 | rs4547785 | T | A | 0.35 | 0.55 | 0.61 | 0.43 | | | | | | | | | | 61kb 3' of AC092846.1 |
| 4 | 24373700 | 1 | 1 | rs4054037 | T | C | 0.31 | 0.50 | 0.61 | 0.43 | | | LIV, OVRY, MUS | | | 7 altered motifs | | | | 47kb 3' of AC092846.1 |
| 4 | 24375265 | 0.98 | 1 | rs34520738 | TA | T | 0.28 | 0.49 | 0.60 | 0.43 | | | LIV | | | 6 altered motifs | | | | 46kb 3' of AC092846.1 |
| 4 | 24375267 | 0.98 | 1 | rs201455240 | A | AGG | 0.27 | 0.49 | 0.60 | 0.43 | | | LIV | | | 5 altered motifs | | | | 46kb 3' of AC092846.1 |
| 4 | 24375268 | 0.98 | 1 | rs202136623 | AT | A | 0.28 | 0.49 | 0.60 | 0.43 | | | LIV | | | 5 altered motifs | | | | 46kb 3' of AC092846.1 |
| 4 | 24375991 | 0.99 | 1 | rs6840491 | C | T | 0.27 | 0.49 | 0.60 | 0.43 | | | | | | YY1 | | | | 45kb 3' of AC092846.1 |

Query SNP: rs11675322 and variants with $r^2$ >= 0.8

| chr | pos (hg38) | LD ($r^2$) | LD (D') | variant | Ref | Alt | AFR freq | AMR freq | ASN freq | EUR freq | SiPhy cons | Promoter histone marks | Enhancer histone marks | DNAse | Proteins bound | Motifs changed | NHGRI/ EBI GWAS hits | GRASP QTL hits | Selected eQTL hits | GENCODE genes | dbSNP func annot |
|---|---|---|---|---|---|---|---|---|---|---|---|---|---|---|---|---|---|---|---|---|---|
| 2 | 12586889 | 1 | 1 | rs13035320 | G | A | 0.07 | 0.07 | 0.10 | 0.08 | | | BLD, THYM, GI | | | 5 altered motifs | | | | 8.5kb 3' of AC096559.1 | |
| 2 | 12586965 | 1 | 1 | rs11684629 | G | C | 0.07 | 0.07 | 0.10 | 0.09 | | | BLD, THYM, GI | THYM,GI,GI | | | | 1 hit | | 8.6kb 3' of AC096559.1 | |
| 2 | 12590252 | 1 | 1 | rs11689528 | G | A | 0.07 | 0.07 | 0.12 | 0.09 | | | 4 tissues | | NFKB | Gfi1,Nr2f2,SEF-1 | | | | 8.7kb 5' of AC096559.2 | |
| 2 | 12592695 | 1 | 1 | rs13023811 | C | T | 0.07 | 0.07 | 0.12 | 0.09 | | | | | | Nkx2 | | | | 6.3kb 5' of AC096559.2 | |
| 2 | 12595488 | 1 | 1 | rs11692451 | A | G | 0.10 | 0.07 | 0.12 | 0.09 | | | | | | Nrf-2 | | | | 3.5kb 5' of AC096559.2 | |
| 2 | 12595601 | 1 | 1 | rs11675322 | G | A | 0.11 | 0.07 | 0.12 | 0.09 | | | | | | | | 1 hit | | 3.4kb 5' of AC096559.2 | |
| 2 | 12597486 | 1 | 1 | rs34778879 | C | T | 0.10 | 0.07 | 0.12 | 0.09 | | | 8 tissues | | | | | | | 1.5kb 5' of AC096559.2 | |

Query SNP: rs9375084 and variants with $r^2$ >= 0.8

| chr | pos (hg38) | LD ($r^2$) | LD (D') | variant | Ref | Alt | AFR freq | AMR freq | ASN freq | EUR freq | SiPhy cons | Promoter histone marks | Enhancer histone marks | DNAse | Proteins bound | Motifs changed | NHGRI/ EBI GWAS hits | GRASP QTL hits | Selected eQTL hits | GENCODE genes | dbSNP func annot |
|---|---|---|---|---|---|---|---|---|---|---|---|---|---|---|---|---|---|---|---|---|---|
| 6 | 121975720 | 0.88 | -0.98 | rs2260075 | C | T | 0.60 | 0.78 | 0.58 | 0.78 | | | | | | Foxk1,Maf,TEF-1 | | | | 236kb 5' of RNU1-18P | |
| 6 | 121976036 | 0.9 | 1 | rs9320846 | C | T | 0.31 | 0.22 | 0.42 | 0.22 | | | | | | Foxl1,Foxq1 | | | | 236kb 5' of RNU1-18P | |
| 6 | 121983088 | 0.9 | 1 | rs9388030 | T | C | 0.32 | 0.22 | 0.42 | 0.22 | | | ESDR | | | AIRE,Mef2,Zfp410 | | | | 229kb 5' of RNU1-18P | |
| 6 | 121983466 | 0.98 | -1 | rs2684221 | C | T | 0.42 | 0.76 | 0.54 | 0.75 | | | | | | 9 altered motifs | | | | 228kb 5' of RNU1-18P | |
| 6 | 121983644 | 1 | 1 | rs9482205 | G | A | 0.49 | 0.23 | 0.47 | 0.25 | | | | | | CIZ,Foxj2,HDAC2 | | | | 228kb 5' of RNU1-18P | |
| 6 | 121983714 | 1 | 1 | rs9490373 | G | A | 0.49 | 0.23 | 0.46 | 0.25 | | | | | | | | | | 228kb 5' of RNU1-18P | |
| 6 | 121983765 | 0.98 | -1 | rs2684224 | C | A | 0.41 | 0.76 | 0.54 | 0.75 | | | | | | PPAR | | | | 228kb 5' of RNU1-18P | |
| 6 | 121984180 | 0.9 | 1 | rs9490374 | G | C | 0.32 | 0.22 | 0.42 | 0.22 | | | | | | MIZF,Pax-2,TCF4 | | | | 227kb 5' of RNU1-18P | |
| 6 | 121984262 | 0.98 | -1 | rs2679684 | G | A | 0.41 | 0.76 | 0.54 | 0.74 | | | | | | Ets,NERF1a | | 1 hit | | 227kb 5' of RNU1-18P | |
| 6 | 121984776 | 1 | 1 | rs7755057 | G | A | 0.49 | 0.23 | 0.46 | 0.25 | | | ESDR | | | MZF1::1-4 | | | | 227kb 5' of RNU1-18P | |
| 6 | 121985149 | 1 | 1 | rs6918418 | A | G | 0.49 | 0.23 | 0.46 | 0.25 | | | ESDR, ESC, IPSC | ESDR,ESDR,IPSC | | 6 altered motifs | | | | 226kb 5' of RNU1-18P | |
| 6 | 121986027 | 1 | 1 | rs5879624 | TA | T | 0.49 | 0.23 | 0.46 | 0.26 | | | | | | 14 altered motifs | | | | 226kb 5' of RNU1-18P | |
| 6 | 121986626 | 1 | 1 | rs9320848 | A | C | 0.49 | 0.23 | 0.46 | 0.25 | | | | | | 8 altered motifs | | | | 225kb 5' of RNU1-18P | |
| 6 | 121986812 | 0.98 | -1 | rs2679683 | T | C | 0.41 | 0.76 | 0.54 | 0.75 | | | | | | | | | | 225kb 5' of RNU1-18P | |
| 6 | 121986850 | 0.98 | -1 | rs2684226 | C | T | 0.41 | 0.76 | 0.54 | 0.75 | | | | | | En-1,Sox | | | | 225kb 5' of RNU1-18P | |
| 6 | 121987046 | 0.97 | 1 | rs9320849 | C | T | 0.50 | 0.23 | 0.46 | 0.25 | | | | | | CDP,Obox6,Pitx2 | | | | 225kb 5' of RNU1-18P | |
| 6 | 121987364 | 0.9 | -1 | rs2945809 | G | A | 0.41 | 0.75 | 0.54 | 0.75 | | | | | | ATF3,Zbtb3 | | | | 224kb 5' of RNU1-18P | |
| 6 | 121987842 | 0.97 | -1 | rs1607935 | G | A | 0.42 | 0.76 | 0.54 | 0.75 | | | | | | HDAC2,PPAR,STAT | | | | 224kb 5' of RNU1-18P | |
| 6 | 121987856 | 0.98 | 1 | rs6936739 | G | A | 0.49 | 0.24 | 0.46 | 0.25 | | | | | | 6 altered motifs | | | | 224kb 5' of RNU1-18P | |
| 6 | 121987948 | 0.98 | 1 | rs145276825 | T | 7-mer | 0.50 | 0.24 | 0.46 | 0.24 | | | | | | | | | | 224kb 5' of RNU1-18P | |
| 6 | 121987949 | 0.97 | 1 | rs35062282 | G | GAA | 0.50 | 0.24 | 0.45 | 0.24 | | | | | | 20 altered motifs | | | | 224kb 5' of RNU1-18P | |
| 6 | 121989297 | 0.98 | -1 | rs2684273 | C | T | 0.42 | 0.76 | 0.54 | 0.75 | | | | | | 4 altered motifs | | | | 222kb 5' of RNU1-18P | |
| 6 | 121989342 | 1 | 1 | rs11330732 | AT | A | 0.50 | 0.23 | 0.46 | 0.26 | | | | | | 17 altered motifs | | | | 222kb 5' of RNU1-18P | |
| 6 | 121989522 | 1 | 1 | rs6927573 | T | C | 0.50 | 0.23 | 0.46 | 0.25 | | | | | | Nkx2,PEBP,TATA | | | | 222kb 5' of RNU1-18P | |
| 6 | 121989640 | 1 | 1 | rs9388031 | T | G | 0.49 | 0.23 | 0.46 | 0.25 | | | | | | GATA,Pou1f1 | | | | 222kb 5' of RNU1-18P | |
| 6 | 121989869 | 1 | 1 | rs2019484 | A | G | 0.49 | 0.23 | 0.46 | 0.25 | | | | | | NF-kappaB | | | | 222kb 5' of RNU1-18P | |
| 6 | 121990024 | 0.9 | 1 | rs2019486 | G | A | 0.32 | 0.22 | 0.42 | 0.22 | | | | | | GATA | | | | 222kb 5' of RNU1-18P | |
| 6 | 121990556 | 0.98 | -1 | rs2679677 | A | G | 0.41 | 0.76 | 0.54 | 0.75 | | | | | | 10 altered motifs | | | | 221kb 5' of RNU1-18P | |
| 6 | 121990905 | 0.98 | 1 | rs9320850 | A | G | 0.49 | 0.24 | 0.47 | 0.25 | | | | | | 5 altered motifs | | | | 221kb 5' of RNU1-18P | |
| 6 | 121991056 | 1 | 1 | rs78213043 | TA | T | 0.50 | 0.23 | 0.46 | 0.25 | | | | | | 36 altered motifs | | | | 221kb 5' of RNU1-18P | |
| 6 | 121991288 | 1 | 1 | rs9320851 | A | C | 0.49 | 0.23 | 0.46 | 0.25 | | | | | | PRDM1,STAT | | | | 220kb 5' of RNU1-18P | |
| 6 | 121991611 | 1 | 1 | rs9490375 | C | T | 0.49 | 0.23 | 0.46 | 0.25 | | | | | | 7 altered motifs | | | | 220kb 5' of RNU1-18P | |
| 6 | 121991626 | 0.9 | 1 | rs9490376 | C | T | 0.32 | 0.22 | 0.42 | 0.22 | | | | | | Gfi1,NF-E2,STAT | | | | 220kb 5' of RNU1-18P | |
| 6 | 121991664 | 0.98 | -1 | rs2679674 | G | A | 0.41 | 0.76 | 0.54 | 0.75 | | | | | | GZF1,RREB-1 | | | | 220kb 5' of RNU1-18P | |
| 6 | 121991689 | 1 | 1 | rs9490377 | T | C | 0.49 | 0.23 | 0.46 | 0.25 | | | | | | 5 altered motifs | | | | 220kb 5' of RNU1-18P | |
| 6 | 121992093 | 0.85 | -1 | rs2679673 | G | T | 0.42 | 0.73 | 0.47 | 0.73 | | | BLD | | | 6 altered motifs | | | 1 hit | 220kb 5' of RNU1-18P | |
| 6 | 121992120 | 1 | 1 | rs9490378 | A | G | 0.50 | 0.23 | 0.46 | 0.25 | | | BLD | SKIN | | 4 altered motifs | | | | 220kb 5' of RNU1-18P | |

| | | | | | | | | | | | | | | | | | | | | |
|---|---|---|---|---|---|---|---|---|---|---|---|---|---|---|---|---|---|---|---|---|
| 6 | 121992412 | 0.83 | -1 | rs2679672 | G | A | 0.20 | 0.73 | 0.54 | 0.75 | | | SKIN, BLD, BRST | | | 6 altered motifs | | | | 219kb 5' of RNU1-18P |
| 6 | 121992473 | 0.85 | 1 | rs201758111 | T | TA | 0.30 | 0.21 | 0.39 | 0.19 | | | | | | 27 altered motifs | | | | 219kb 5' of RNU1-18P |
| 6 | 121992475 | 0.85 | 1 | rs12196252 | T | A | 0.30 | 0.21 | 0.39 | 0.19 | | | | | | Arid5a,CEBPG | | | | 219kb 5' of RNU1-18P |
| 6 | 121992476 | 0.88 | 0.98 | rs199686254 | TA | T | 0.34 | 0.22 | 0.39 | 0.20 | | | | | | Arid5a,CEBPG,Pou2f2 | | | | 219kb 5' of RNU1-18P |
| 6 | 121992477 | 0.95 | 1 | rs9490379 | A | T | 0.48 | 0.23 | 0.43 | 0.23 | | | | | | 4 altered motifs | | | | 219kb 5' of RNU1-18P |
| 6 | 121992555 | 1 | 1 | rs9375083 | G | A | 0.49 | 0.23 | 0.46 | 0.25 | | | | | | | | | | 219kb 5' of RNU1-18P |
| 6 | 121992698 | 1 | 1 | rs9375084 | C | A | 0.50 | 0.23 | 0.46 | 0.25 | | | | | | CDP,Hmx | | | | 219kb 5' of RNU1-18P |
| 6 | 121993003 | 0.9 | 1 | rs1851210 | G | A | 0.32 | 0.22 | 0.42 | 0.22 | | | | | | 4 altered motifs | | | | 219kb 5' of RNU1-18P |
| 6 | 121993111 | 0.93 | 0.98 | rs1851212 | G | A | 0.46 | 0.24 | 0.45 | 0.25 | | | | | | EBF | | | | 219kb 5' of RNU1-18P |
| 6 | 121993396 | 0.98 | -1 | rs1474227 | C | T | 0.41 | 0.76 | 0.54 | 0.75 | | | | | | 5 altered motifs | | | | 218kb 5' of RNU1-18P |
| 6 | 121993477 | 1 | 1 | rs7764548 | C | T | 0.49 | 0.23 | 0.46 | 0.25 | | | 4 tissues | | | 7 altered motifs | | | | 218kb 5' of RNU1-18P |
| 6 | 121993795 | 1 | 1 | rs9320852 | G | A | 0.49 | 0.23 | 0.46 | 0.25 | | | 4 tissues | | | 5 altered motifs | | | | 218kb 5' of RNU1-18P |
| 6 | 121994894 | 1 | 1 | rs9490381 | A | C | 0.50 | 0.23 | 0.46 | 0.25 | | | ESDR, IPSC, BRST | ESDR,SKIN | | 5 altered motifs | | | | 217kb 5' of RNU1-18P |
| 6 | 121994909 | 1 | 1 | rs9375086 | A | T | 0.49 | 0.23 | 0.46 | 0.25 | | | ESDR, IPSC, BRST | ESDR,SKIN | | 4 altered motifs | | | | 217kb 5' of RNU1-18P |
| 6 | 121995189 | 1 | 1 | rs9388033 | C | T | 0.50 | 0.23 | 0.46 | 0.25 | | | ESDR, IPSC, SKIN | | | 6 altered motifs | | | | 216kb 5' of RNU1-18P |
| 6 | 121995205 | 1 | 1 | rs9388034 | G | C | 0.49 | 0.23 | 0.46 | 0.25 | | | ESDR, IPSC, SKIN | | | Mef2,Pou2f2 | | | | 216kb 5' of RNU1-18P |
| 6 | 121995446 | 0.97 | 0.98 | rs9385226 | G | C | 0.50 | 0.23 | 0.46 | 0.25 | | | | | | 12 altered motifs | | | | 216kb 5' of RNU1-18P |
| 6 | 121995493 | 1 | 1 | rs9388035 | G | A | 0.50 | 0.23 | 0.46 | 0.25 | | | | | | 4 altered motifs | | | | 216kb 5' of RNU1-18P |
| 6 | 121995862 | 0.82 | -0.98 | rs2684278 | C | A | 0.42 | 0.79 | 0.54 | 0.82 | | | | | | CIZ,Fox,Nkx2 | | | | 216kb 5' of RNU1-18P |
| 6 | 121996122 | 0.84 | 1 | rs9375088 | T | A | 0.49 | 0.20 | 0.46 | 0.18 | | | | | | Mrg | | | | 216kb 5' of RNU1-18P |
| 6 | 121997371 | 0.81 | 1 | rs9401480 | T | C | 0.49 | 0.20 | 0.45 | 0.18 | | | | | | 4 altered motifs | | | | 214kb 5' of RNU1-18P |
| 6 | 121997592 | 0.84 | 1 | rs7758158 | G | A | 0.48 | 0.20 | 0.46 | 0.18 | | | | | | 4 altered motifs | | | | 214kb 5' of RNU1-18P |
| 6 | 121998932 | 0.98 | -1 | rs2684280 | G | A | 0.42 | 0.76 | 0.54 | 0.75 | | | 7 tissues | SKIN | | Arnt,Mxi1,Myc | | | | 213kb 5' of RNU1-18P |
| 6 | 122000561 | 0.84 | -0.98 | rs2679670 | G | C | 0.63 | 0.79 | 0.58 | 0.78 | | | ESC, IPSC, BRST | | | | | | | 211kb 5' of RNU1-18P |
| 6 | 122002712 | 0.98 | -1 | rs1521241 | T | C | 0.42 | 0.76 | 0.54 | 0.75 | | | 4 tissues | | | Rad21,Znf143 | | | | 209kb 5' of RNU1-18P |
| 6 | 122002869 | 0.98 | -1 | rs2684282 | A | C | 0.42 | 0.76 | 0.54 | 0.75 | | | BLD, CRVX | | | HNF1 | | | | 209kb 5' of RNU1-18P |
| 6 | 122003716 | 0.98 | -1 | rs2684283 | T | C | 0.42 | 0.76 | 0.54 | 0.75 | | | | | | | | | | 208kb 5' of RNU1-18P |
| 6 | 122004019 | 0.89 | -1 | rs2461006 | A | G | 0.42 | 0.74 | 0.53 | 0.73 | | | | | | 7 altered motifs | | | | 208kb 5' of RNU1-18P |
| 6 | 122004540 | 0.94 | -0.98 | rs2679668 | T | C | 0.45 | 0.77 | 0.55 | 0.76 | | | HRT | | | | | | | 207kb 5' of RNU1-18P |
| 6 | 122004630 | 0.88 | -0.98 | rs2684284 | G | C | 0.60 | 0.78 | 0.58 | 0.78 | | | HRT | | | HNF4 | | | | 207kb 5' of RNU1-18P |
| 6 | 122005572 | 0.94 | -0.98 | rs2679667 | C | A | 0.45 | 0.77 | 0.55 | 0.76 | | | STRM, MUS | | | Foxa | | | | 206kb 5' of RNU1-18P |
| 6 | 122007111 | 0.9 | -0.95 | rs2679666 | G | T | 0.35 | 0.76 | 0.55 | 0.76 | | IPSC | 6 tissues | MUS,MUS | | CIZ,Homez,NF-AT | | | | 205kb 5' of RNU1-18P |
| 6 | 122008121 | 0.94 | -0.98 | rs2679665 | A | G | 0.47 | 0.77 | 0.55 | 0.76 | | IPSC | 10 tissues | 5 tissues | | PPAR,RREB-1 | | | | 204kb 5' of RNU1-18P |

Query SNP: rs9576907 and variants with $r^2$ >= 0.8

| chr | pos (hg38) | LD ($r^2$) | LD (D') | variant | Ref | Alt | AFR freq | AMR freq | ASN freq | EUR freq | SiPhy cons | Promoter histone marks | Enhancer histone marks | DNAse | Proteins bound | Motifs changed | NHGRI/EBI GWAS hits | GRASP QTL hits | Selected eQTL hits | GENCODE genes | dbSNP func annot |
|---|---|---|---|---|---|---|---|---|---|---|---|---|---|---|---|---|---|---|---|---|---|
| 13 | 39839788 | 0.88 | 0.94 | rs9548944 | C | T | 0.19 | 0.59 | 0.72 | 0.56 | | | STRM, BONE | | | | | 1 hit | | 17kb 3' of SNORD116 | |
| 13 | 39846533 | 0.87 | -0.98 | rs34556254 | G | GT | 0.66 | 0.39 | 0.27 | 0.45 | | | ESDR | | | 31 altered motifs | | | | 11kb 3' of SNORD116 | |
| 13 | 39846535 | 0.87 | -0.98 | rs2985239 | A | T | 0.66 | 0.39 | 0.27 | 0.44 | | | ESDR | | | 16 altered motifs | | | | 11kb 3' of SNORD116 | |
| 13 | 39846536 | 0.87 | -0.98 | rs199791261 | AT | A | 0.66 | 0.39 | 0.27 | 0.44 | | | ESDR | | | 18 altered motifs | | | | 11kb 3' of SNORD116 | |
| | | 0.87 | -0.98 | rs66487077 | T | A | 0.66 | 0.39 | 0.27 | 0.44 | | | ESDR | | | 17 altered motifs | | | | 11kb 3' of SNORD116 | |
| 13 | 39846625 | 0.96 | -1 | rs9603632 | A | G | 0.66 | 0.40 | 0.29 | 0.44 | | | ESDR | | | 4 altered motifs | | | | 11kb 3' of SNORD116 | |
| 13 | 39846962 | 0.96 | -1 | rs3012139 | C | T | 0.66 | 0.40 | 0.29 | 0.45 | | | ESDR | | | Hbp1 | | | | 10kb 3' of SNORD116 | |
| 13 | 39847335 | 1 | 1 | rs2209234 | C | T | 0.20 | 0.59 | 0.71 | 0.55 | | | ESDR | | | 5 altered motifs | | | | 9.8kb 3' of SNORD116 | |
| 13 | 39847572 | 1 | 1 | rs9576907 | G | A | 0.20 | 0.59 | 0.71 | 0.55 | | | ESDR | MUS,MUS | | 8 altered motifs | | | | 9.6kb 3' of SNORD116 | |
| 13 | 39848676 | 0.9 | 1 | rs3928762 | T | C | 0.20 | 0.57 | 0.69 | 0.53 | | | | | | | | | | 8.5kb 3' of SNORD116 | |
| 13 | 39849316 | 1 | 1 | rs4351932 | C | T | 0.20 | 0.59 | 0.71 | 0.55 | | | | | | 4 altered motifs | | | | 7.8kb 3' of SNORD116 | |
| 13 | 39849413 | 0.9 | 0.97 | rs2324454 | A | G | 0.20 | 0.58 | 0.68 | 0.52 | | | | | | E2F | | | | 7.7kb 3' of SNORD116 | |
| 13 | 39849424 | 0.88 | -1 | rs4480665 | T | C | 0.63 | 0.38 | 0.28 | 0.43 | | | | | | Obox3,Obox6,Pbx3 | | | | 7.7kb 3' of SNORD116 | |
| 13 | 39849791 | 0.93 | 1 | rs2324455 | T | G | 0.20 | 0.57 | 0.67 | 0.53 | | | | | | Foxo,Ik-2,NF-AT | | | | 7.3kb 3' of SNORD116 | |
| 13 | 39851265 | 0.88 | 0.98 | rs79646311 | C | G | 0.20 | 0.57 | 0.70 | 0.54 | | | | | | 4 altered motifs | | | | 5.9kb 3' of SNORD116 | |
| 13 | 39851681 | 0.89 | -0.96 | rs7987851 | T | G | 0.66 | 0.40 | 0.29 | 0.45 | | | | | | Nkx2 | | | | 5.5kb 3' of SNORD116 | |
| 13 | 39852445 | 0.9 | 0.97 | rs9548952 | C | T | 0.20 | 0.58 | 0.71 | 0.55 | | | | | | | | | | 4.7kb 3' of SNORD116 | |
| 13 | 39854252 | 0.93 | 0.97 | rs2875347 | C | T | 0.20 | 0.59 | 0.71 | 0.55 | | | | | | Pax-5 | | | | 2.9kb 3' of SNORD116 | |
| 13 | 39855824 | 0.89 | -0.96 | rs912418 | T | C | 0.66 | 0.40 | 0.29 | 0.45 | | | 5 tissues | ESDR,ESDR | | GR | | | | 1.3kb 3' of SNORD116 | |
| 13 | 39856826 | 0.89 | -0.96 | rs2985248 | A | G | 0.63 | 0.40 | 0.29 | 0.45 | | | ESDR, IPSC, BLD | | | 5 altered motifs | | | | 308bp 3' of SNORD116 | |

| | | | | | | | | | | | | | | | | | | | | |
|---|---|---|---|---|---|---|---|---|---|---|---|---|---|---|---|---|---|---|---|---|
| 13 | 39858567 | 0.93 | 0.97 | rs7139604 | G | A | 0.21 | 0.59 | 0.71 | 0.55 | | | | | | HDAC2,Hoxa7 | | | | 1.3kb 5' of SNORD116 |
| 13 | 39862179 | 0.93 | 0.97 | rs7327167 | C | T | 0.21 | 0.59 | 0.71 | 0.55 | | | | SKIN | | GR,Pax-4,p300 | | | | 5kb 5' of SNORD116 |
| 13 | 39862557 | 0.89 | 0.96 | rs9566480 | T | C | 0.32 | 0.60 | 0.71 | 0.55 | | | VAS | | | HDAC2,p300 | | | | 5.3kb 5' of SNORD116 |
| 13 | 39863519 | 0.93 | 0.97 | rs968130 | G | A | 0.21 | 0.59 | 0.71 | 0.55 | | | 5 tissues | 4 tissues | | Hoxb8,Pou2f2,Pou3f2 | | | | 6.3kb 5' of SNORD116 |
| 13 | 39866173 | 0.89 | 0.96 | rs996546 | A | G | 0.33 | 0.60 | 0.71 | 0.55 | | | HRT | | | 18 altered motifs | | | | 8.9kb 5' of SNORD116 |
| 13 | 39867278 | 0.89 | 0.96 | rs4941945 | C | G | 0.33 | 0.60 | 0.71 | 0.55 | | | | | | NERF1a | | | | 10kb 5' of SNORD116 |

Query SNP: rs2029818 and variants with $r^2$ >= 0.8

| chr | pos (hg38) | LD ($r^2$) | LD (D') | variant | Ref | Alt | AFR freq | AMR freq | ASN freq | EUR freq | SiPhy cons | Promoter histone marks | Enhancer histone marks | DNAse | Proteins bound | Motifs changed | NHGRI/ EBI GWAS hits | GRASP QTL hits | Selected eQTL hits | GENCODE genes | dbSNP func annot |
|---|---|---|---|---|---|---|---|---|---|---|---|---|---|---|---|---|---|---|---|---|---|
| 8 | 79616738 | 0.82 | 0.94 | rs7013510 | G | A | 0.39 | 0.40 | 0.17 | 0.49 | | BRN | FAT, BRN | | | CTCF,LUN-1,NF-kappaB | | | 1 hit | STMN2 | intronic |
| 8 | 79639207 | 0.99 | 1 | rs1159737 | A | G | 0.32 | 0.39 | 0.15 | 0.50 | | | ESC, IPSC | | | 7 altered motifs | | | 1 hit | STMN2 | intronic |
| 8 | 79640833 | 1 | 1 | rs2029818 | T | C | 0.32 | 0.38 | 0.15 | 0.50 | | | BRN | BRN,BRN | | p300 | | | 1 hit | STMN2 | intronic |
| 8 | 79640845 | 0.94 | 0.98 | rs2029819 | A | T | 0.37 | 0.39 | 0.15 | 0.50 | | | BRN | BRN,BRN | | | | | 1 hit | STMN2 | intronic |

Query SNP: rs3768480 and variants with $r^2$ >= 0.8

| chr | pos (hg38) | LD ($r^2$) | LD (D') | variant | Ref | Alt | AFR freq | AMR freq | ASN freq | EUR freq | SiPhy cons | Promoter histone marks | Enhancer histone marks | DNAse | Proteins bound | Motifs changed | NHGRI/ EBI GWAS hits | GRASP QTL hits | Selected eQTL hits | GENCODE genes | dbSNP func annot |
|---|---|---|---|---|---|---|---|---|---|---|---|---|---|---|---|---|---|---|---|---|---|
| 1 | 109968639 | 0.88 | 0.95 | rs74904912 | TAA | T | 0.06 | 0.33 | 0.37 | 0.48 | | | SKIN | | | 4 altered motifs | | | 8 hits | 16kb 5' of AHCYL1 | |
| 1 | 109988373 | 1 | 1 | rs3768480 | G | C | 0.06 | 0.32 | 0.20 | 0.46 | | 4 tissues | 10 tissues | | | | | 1 hit | 9 hits | AHCYL1 | intronic |
| 1 | 110008588 | 0.89 | -0.99 | rs10715355 | AT | A | 0.78 | 0.66 | 0.79 | 0.52 | | | | BRN | | Nanog,PPAR | | | | AHCYL1 | intronic |
| 1 | 110034040 | 0.93 | 0.97 | rs34762282 | G | A | 0.07 | 0.33 | 0.20 | 0.46 | | 22 tissues | 5 tissues | BLD | | | | | 9 hits | FAM40A | |
| 1 | 110037593 | 0.86 | -0.97 | rs10745307 | G | A | 0.77 | 0.65 | 0.80 | 0.54 | | | | | | Pou3f3 | | | 11 hits | FAM40A | intronic |
| 1 | 110042515 | 0.89 | -0.95 | rs6537651 | A | G | 0.92 | 0.68 | 0.78 | 0.59 | | | ESDR, FAT | | | | | 1 hit | 16 hits | FAM40A | intronic |
| 1 | 110046957 | 0.9 | 0.95 | rs1965990 | G | A | 0.06 | 0.32 | 0.21 | 0.42 | | | | | | 5 altered motifs | | | 15 hits | FAM40A | intronic |
| 1 | 110048194 | 0.9 | 0.95 | rs12042401 | C | T | 0.06 | 0.32 | 0.20 | 0.42 | | | 9 tissues | | | Dobox4,Gfi1,Smad4 | | 1 hit | 16 hits | FAM40A | intronic |
| 1 | 110055830 | 0.9 | -0.95 | rs666842 | T | C | 0.94 | 0.68 | 0.80 | 0.58 | | | 12 tissues | SKIN,GI,MUS | | NF-kappaB | | | 14 hits | RP4-773N10.5 | |
| 1 | 110055955 | 0.83 | -0.95 | rs10745308 | A | C | 0.77 | 0.66 | 0.80 | 0.58 | | | | GI | | | | 2 hits | 17 hits | RP4-773N10.5 | |
| 1 | 110063984 | 0.82 | -0.94 | rs751387 | T | G | 0.75 | 0.66 | 0.79 | 0.60 | | | SKIN, MUS | | | NF-kappaB,Pax-2 | | | 15 hits | RP4-773N10.5 | intronic |
| 1 | 110064324 | 0.82 | -0.94 | rs1077581 | C | G | 0.75 | 0.66 | 0.79 | 0.60 | | | | | | CTCF,Pax-2 | | | 16 hits | RP4-773N10.5 | intronic |
| 1 | 110064397 | 0.82 | -0.94 | rs1077580 | A | G | 0.75 | 0.66 | 0.79 | 0.60 | | | | | | AP-3 | | 1 hit | 16 hits | RP4-773N10.5 | intronic |
| 1 | 110066139 | 0.89 | -0.95 | rs481420 | C | G | 0.94 | 0.68 | 0.79 | 0.60 | | ESDR | FAT, SKIN, MUS | | NRSF | | | | 10 hits | RP4-773N10.5 | intronic |
| 1 | 110066666 | 0.82 | -0.94 | rs3754443 | T | G | 0.75 | 0.66 | 0.79 | 0.60 | | MUS | SKIN | | | | | | 15 hits | RP4-773N10.5 | intronic |

Query SNP: rs1039948 and variants with $r^2$ >= 0.8

| chr | pos (hg38) | LD ($r^2$) | LD (D') | variant | Ref | Alt | AFR freq | AMR freq | ASN freq | EUR freq | SiPhy cons | Promoter histone marks | Enhancer histone marks | DNAse | Proteins bound | Motifs changed | NHGRI/ EBI GWAS hits | GRASP QTL hits | Selected eQTL hits | GENCODE genes | dbSNP func annot |
|---|---|---|---|---|---|---|---|---|---|---|---|---|---|---|---|---|---|---|---|---|---|
| 4 | 139717809 | 0.91 | -0.99 | rs345983 | C | T | 0.47 | 0.58 | 0.66 | 0.68 | | | ESDR, OVRY | | | | | | 1 hit | MGST2 | 3'-UTR |
| 4 | 139724119 | 1 | 1 | rs1039948 | G | C | 0.65 | 0.43 | 0.37 | 0.31 | | | | MUS | | 4 altered motifs | | | 1 hit | MGST2 | intronic |

Query SNP: rs454266 and variants with $r^2$ >= 0.8

| chr | pos (hg38) | LD ($r^2$) | LD (D') | variant | Ref | Alt | AFR freq | AMR freq | ASN freq | EUR freq | SiPhy cons | Promoter histone marks | Enhancer histone marks | DNAse | Proteins bound | Motifs changed | NHGRI/ EBI GWAS hits | GRASP QTL hits | Selected eQTL hits | GENCODE genes | dbSNP func annot |
|---|---|---|---|---|---|---|---|---|---|---|---|---|---|---|---|---|---|---|---|---|---|
| 10 | 117569549 | 0.83 | 0.93 | rs2532661 | A | C | 0.26 | 0.55 | 0.55 | 0.66 | | | FAT, MUS | BLD | | | | | | CTA-109P11.1 | |
| 10 | 117570090 | 0.87 | 0.95 | rs386923 | A | G | 0.27 | 0.55 | 0.55 | 0.67 | | | 4 tissues | KID,MUS | | | | | | CTA-109P11.1 | |
| 10 | 117570411 | 0.88 | 0.95 | rs35584605 | AG | A | 0.26 | 0.55 | 0.55 | 0.66 | | | ESDR, FAT, MUS | MUS | | 9 altered motifs | | | | CTA-109P11.1 | |
| 10 | 117570574 | 0.88 | 0.95 | rs399037 | T | C | 0.26 | 0.55 | 0.55 | 0.66 | | | ESDR, FAT, MUS | BLD | | | | | | CTA-109P11.1 | |
| 10 | 117570609 | 0.88 | 0.95 | rs390211 | C | T | 0.26 | 0.55 | 0.55 | 0.66 | | | ESDR, FAT, MUS | BLD,MUS,SKIN | | 5 altered motifs | | | | CTA-109P11.1 | |
| 10 | 117571215 | 0.88 | 0.95 | rs421528 | C | T | 0.26 | 0.55 | 0.55 | 0.66 | | | | | EBF1 | NF-I,STAT | | | | CTA-109P11.1 | |
| 10 | 117571473 | 0.88 | 0.95 | rs396011 | T | C | 0.26 | 0.55 | 0.55 | 0.66 | | | | | | Gfi1 | | | | CTA-109P11.1 | |
| 10 | 117572086 | 0.88 | 0.95 | rs430152 | A | G | 0.26 | 0.55 | 0.55 | 0.66 | | | | | | 4 altered motifs | | | | CTA-109P11.1 | |
| 10 | 117572990 | 0.92 | 0.97 | rs375973 | T | G | 0.25 | 0.54 | 0.55 | 0.64 | | | | | | | | | | 532bp 5' of CTA-109P11.1 | |
| 10 | 117573045 | 0.92 | 0.97 | rs422221 | C | T | 0.25 | 0.54 | 0.55 | 0.64 | | | | | | | | | | 587bp 5' of CTA-109P11.1 | |
| 10 | 117573495 | 0.92 | 0.97 | rs2768311 | A | G | 0.25 | 0.54 | 0.55 | 0.65 | | | | | | 4 altered motifs | | | | 1kb 5' of CTA-109P11.1 | |
| 10 | 117574044 | 0.9 | 0.97 | rs740738 | G | T | 0.25 | 0.53 | 0.55 | 0.64 | | | | | | AP-1 | | 1 hit | | 1.6kb 5' of CTA-109P11.1 | |
| 10 | 117575692 | 0.93 | 0.98 | rs424221 | A | G | 0.31 | 0.53 | 0.55 | 0.64 | | | | | | 7 altered motifs | | | | 3.2kb 5' of CTA-109P11.1 | |
| 10 | 117576271 | 0.97 | 1 | rs374658 | A | C | 0.32 | 0.55 | 0.55 | 0.65 | | | | | | ERalpha-a,Pax-5,Zfx | | | | 3.8kb 5' of CTA-109P11.1 | |
| 10 | 117576509 | 0.98 | 1 | rs728558 | A | G | 0.30 | 0.54 | 0.55 | 0.65 | | | | | | CTCF,Gfi1 | | | | 4.1kb 5' of CTA-109P11.1 | |
| 10 | 117576638 | 0.91 | 1 | rs10510031 | T | C | 0.22 | 0.51 | 0.54 | 0.60 | | | | | | 5 altered motifs | | | | 4.2kb 5' of CTA-109P11.1 | |
| 10 | 117577112 | 1 | 1 | rs454266 | G | A | 0.28 | 0.54 | 0.55 | 0.65 | | | | | | 7 altered motifs | | 1 hit | | 4.7kb 5' of CTA-109P11.1 | |
| 10 | 117577481 | 0.89 | 1 | rs7088735 | C | A | 0.14 | 0.51 | 0.54 | 0.60 | | | | | ZNF263 | 5 altered motifs | | | | 5kb 5' of CTA-109P11.1 | |
| 10 | 117577946 | 1 | 1 | rs2786200 | C | G | 0.29 | 0.54 | 0.55 | 0.65 | | | | | | ERalpha-a,Nanog | | 1 hit | | 5.5kb 5' of CTA-109P11.1 | |
| 10 | 117578229 | 0.91 | 1 | rs4751630 | A | G | 0.22 | 0.51 | 0.55 | 0.60 | | | | | | CDP,COMP1,Pbx-1 | | | | 5.8kb 5' of CTA-109P11.1 | |
| 10 | 117578765 | 1 | -1 | rs2786201 | G | T | 0.72 | 0.46 | 0.45 | 0.36 | | | | | | Dobox4 | | | | 6.3kb 5' of CTA-109P11.1 | |
| 10 | 117578988 | 1 | -1 | rs2786202 | G | A | 0.72 | 0.46 | 0.45 | 0.36 | | | | | | Ets,ZBTB33 | | | | 6.5kb 5' of CTA-109P11.1 | |
| 10 | 117579713 | 1 | -1 | rs2531679 | C | T | 0.74 | 0.46 | 0.45 | 0.35 | | | | | | 17 altered motifs | | | | 7.3kb 5' of CTA-109P11.1 | |
| 10 | 117579887 | 1 | -1 | rs2531680 | T | G | 0.74 | 0.46 | 0.45 | 0.35 | | | | | | GATA | | | | 7.4kb 5' of CTA-109P11.1 | |

| | | | | | | | | | | | | | | | | | | | | |
|---|---|---|---|---|---|---|---|---|---|---|---|---|---|---|---|---|---|---|---|---|
| 10 | 117580404 | 0.88 | -0.97 | rs2531681 | A | T | 0.71 | 0.45 | 0.45 | 0.36 | | | | | | 9 altered motifs | | | | 7.9kb 5' of CTA-109P11.1 |
| 10 | 117580938 | 0.96 | -0.98 | rs2531682 | C | T | 0.73 | 0.46 | 0.46 | 0.35 | | | | | | CHOP::CEBPalpha | | 1 hit | | 8.5kb 5' of CTA-109P11.1 |
| 10 | 117581319 | 0.96 | -0.98 | rs2768313 | T | C | 0.74 | 0.46 | 0.45 | 0.35 | | | FAT | | | 4 altered motifs | | 1 hit | | 8.9kb 5' of CTA-109P11.1 |
| 10 | 117581432 | 0.95 | -0.98 | rs2531669 | A | T | 0.72 | 0.46 | 0.45 | 0.36 | | | FAT | | | 13 altered motifs | | | | 9kb 5' of CTA-109P11.1 |
| 10 | 117581770 | 0.96 | -0.98 | rs2531670 | A | T | 0.73 | 0.46 | 0.45 | 0.35 | | | FAT, MUS, HRT | MUS,SKIN | | Ets,NERF1a,TCF12 | | | | 9.3kb 5' of CTA-109P11.1 |
| 10 | 117582093 | 0.96 | 0.98 | rs4457665 | C | T | 0.27 | 0.54 | 0.55 | 0.65 | | | FAT | MUS | | HP1-site-factor | | 1 hit | | 9.6kb 5' of CTA-109P11.1 |
| 10 | 117583180 | 0.96 | 0.98 | rs6585442 | T | C | 0.27 | 0.54 | 0.55 | 0.65 | | | | | | 7 altered motifs | | 1 hit | | 11kb 5' of CTA-109P11.1 |
| 10 | 117583429 | 0.93 | 0.98 | rs10787778 | T | A | 0.27 | 0.53 | 0.55 | 0.65 | | | | | | | | | | 11kb 5' of CTA-109P11.1 |
| 10 | 117584924 | 0.84 | 0.98 | rs10749244 | T | C | 0.21 | 0.51 | 0.53 | 0.60 | | | | | | GR,Irf | | | | 12kb 5' of CTA-109P11.1 |
| 10 | 117585905 | 0.95 | 0.98 | rs10787779 | A | G | 0.27 | 0.54 | 0.54 | 0.65 | | | | | | Nkx3 | | | | 13kb 5' of CTA-109P11.1 |
| 10 | 117586047 | 0.93 | 0.98 | rs1415424 | G | C | 0.22 | 0.53 | 0.53 | 0.65 | | | MUS | MUS,GI | | NRSF,PPAR,Sin3Ak-20 | | 1 hit | | 14kb 5' of CTA-109P11.1 |
| 10 | 117586415 | 0.91 | 0.97 | rs1415425 | A | C | 0.35 | 0.54 | 0.54 | 0.65 | | | | MUS | | | | | | 14kb 5' of CTA-109P11.1 |

Query SNP: rs4591370 and variants with $r^2$ >= 0.8

| chr | pos (hg38) | LD ($r^2$) | LD (D') | variant | Ref | Alt | AFR freq | AMR freq | ASN freq | EUR freq | SiPhy cons | Promoter histone marks | Enhancer histone marks | DNAse | Proteins bound | Motifs changed | NHGRI/EBI GWAS hits | GRASP QTL hits | Selected eQTL hits | GENCODE genes | dbSNP func annot |
|---|---|---|---|---|---|---|---|---|---|---|---|---|---|---|---|---|---|---|---|---|---|
| 2 | 21074277 | 0.81 | 0.94 | rs478588 | A | G | 0.38 | 0.81 | 0.99 | 0.76 | | | | | | Foxm1 | | | 1 hit | 30kb 5' of APOB | |
| 2 | 21080744 | 0.84 | 0.94 | rs568938 | C | T | 0.40 | 0.82 | 0.99 | 0.76 | | | | | | 4 altered motifs | | | 1 hit | 37kb 5' of APOB | |
| 2 | 21090705 | 0.84 | 0.94 | rs614303 | G | A | 0.41 | 0.82 | 0.99 | 0.76 | | | | | | | | | 1 hit | 33kb 5' of AC010872.2 | |
| 2 | 21102316 | 0.92 | 0.98 | rs312944 | A | T | 0.63 | 0.83 | 0.99 | 0.75 | | | | | | AP-2,PEBP | | | 1 hit | 22kb 5' of AC010872.2 | |
| 2 | 21117059 | 0.89 | 0.98 | rs200202030 | AG | A | 0.62 | 0.81 | 0.99 | 0.75 | | | | | | 9 altered motifs | | | 1 hit | 6.9kb 5' of AC010872.2 | |
| 2 | 21117060 | 0.94 | 0.98 | rs312953 | G | A | 0.64 | 0.82 | 0.99 | 0.75 | | | | | | 7 altered motifs | | | 1 hit | 6.9kb 5' of AC010872.2 | |
| 2 | 21146943 | 1 | 1 | rs312970 | A | T | 0.64 | 0.83 | 0.99 | 0.75 | | | | | | | | 1 hit | 1 hit | 3.7kb 3' of AC010872.2 | |
| 2 | 21148323 | 0.94 | 1 | rs67545613 | CTCT | C | 0.64 | 0.82 | 0.99 | 0.75 | | | | | | HDAC2 | | | 1 hit | 5.1kb 3' of AC010872.2 | |
| 2 | 21148327 | 0.85 | 1 | rs55857688 | TC | T,TCTT | 0.59 | 0.80 | 0.94 | 0.70 | | | | | | | | | 1 hit | 5.1kb 3' of AC010872.2 | |
| 2 | 21154833 | 1 | 1 | rs312979 | A | T | 0.65 | 0.83 | 0.99 | 0.75 | | | LIV | | | AP-1,GATA,PRDM1 | | | 1 hit | 12kb 3' of AC010872.2 | |
| 2 | 21155056 | 1 | 1 | rs72336878 | AATT | A | 0.63 | 0.83 | 0.99 | 0.75 | | | LIV | | | 5 altered motifs | | | 1 hit | 12kb 3' of AC010872.2 | |
| 2 | 21155141 | 1 | 1 | rs35425016 | A | AA,AC | 0.65 | 0.83 | 0.99 | 0.75 | | | LIV | | | | | | 1 hit | 12kb 3' of AC010872.2 | |
| 2 | 21155279 | 1 | 1 | rs312981 | G | A | 0.64 | 0.83 | 0.99 | 0.75 | | | LIV | | | 4 altered motifs | | | 1 hit | 12kb 3' of AC010872.2 | |
| 2 | 21155354 | 1 | 1 | rs312982 | G | C | 0.64 | 0.83 | 0.99 | 0.75 | | | LIV | | | Hlx1 | | | 1 hit | 12kb 3' of AC010872.2 | |
| 2 | 21155559 | 0.98 | 1 | rs34908258 | CT | C | 0.64 | 0.82 | 0.99 | 0.75 | | LIV | LIV | KID,LNG,PLCNT | | 4 altered motifs | | | | 12kb 3' of AC010872.2 | |
| 2 | 21155561 | 0.88 | 1 | rs200200214 | TG | T | 0.60 | 0.81 | 0.97 | 0.73 | | LIV | LIV | KID,LNG,PLCNT | | AP-2,CCNT2,INSM1 | | | | 12kb 3' of AC010872.2 | |
| 2 | 21155708 | 1 | 1 | rs312983 | A | C | 0.64 | 0.83 | 0.99 | 0.75 | | LIV | LIV | | CEBPB | 5 altered motifs | | | 1 hit | 12kb 3' of AC010872.2 | |
| 2 | 21155906 | 1 | 1 | rs312984 | C | T | 0.64 | 0.83 | 0.99 | 0.75 | | LIV | SKIN, LIV | LIV | 11 bound proteins | Hmx,Nkx2,Nkx3 | | | 1 hit | 13kb 3' of AC010872.2 | |
| 2 | 21155933 | 1 | 1 | rs312985 | A | G | 0.64 | 0.83 | 0.99 | 0.75 | | LIV | LIV | LIV | 11 bound proteins | | 1 hit | | 1 hit | 13kb 3' of AC010872.2 | |
| 2 | 21156272 | 1 | 1 | rs151224593 | CAACTA | C | 0.64 | 0.83 | 0.99 | 0.75 | | LIV | | | HEY1 | 4 altered motifs | | | | 13kb 3' of AC010872.2 | |
| 2 | 21158396 | 1 | 1 | rs529396 | G | C,T | 0.64 | 0.83 | 0.99 | 0.75 | | | | | | | | | 1 hit | 15kb 3' of AC010872.2 | |
| 2 | 21158563 | 1 | 1 | rs530474 | G | A | 0.64 | 0.83 | 0.99 | 0.75 | | | | | | 5 altered motifs | | | 1 hit | 15kb 3' of AC010872.2 | |
| 2 | 21158618 | 1 | 1 | rs559318 | C | T | 0.64 | 0.83 | 0.99 | 0.75 | | | | | | E2A,Mef2,ZEB1 | | | 1 hit | 15kb 3' of AC010872.2 | |
| 2 | 21158736 | 1 | 1 | rs532300 | C | A | 0.64 | 0.83 | 0.99 | 0.75 | | | | | | CEBPG,Mef2 | | | 1 hit | 15kb 3' of AC010872.2 | |
| 2 | 21158817 | 0.98 | 1 | rs558130 | T | G | 0.64 | 0.82 | 0.99 | 0.75 | | | | | | INSM1,SRF,ZBTB7A | | | 1 hit | 16kb 3' of AC010872.2 | |
| 2 | 21158830 | 1 | 1 | rs533211 | G | A | 0.64 | 0.83 | 0.99 | 0.75 | | | | | | Pou2f2,SRF | | | 1 hit | 16kb 3' of AC010872.2 | |
| 2 | 21158909 | 1 | 1 | rs557197 | T | G | 0.64 | 0.83 | 0.99 | 0.75 | | | | | | 5 altered motifs | | | 1 hit | 16kb 3' of AC010872.2 | |
| 2 | 21159491 | 1 | 1 | rs560522 | A | C | 0.64 | 0.83 | 0.99 | 0.75 | | | | | | ATF3 | | | 1 hit | 16kb 3' of AC010872.2 | |
| 2 | 21159914 | 1 | 1 | rs527034 | C | A | 0.64 | 0.83 | 0.99 | 0.75 | | | LIV | | | 4 altered motifs | | 1 hit | 1 hit | 17kb 3' of AC010872.2 | |
| 2 | 21160104 | 1 | 1 | rs525172 | T | G | 0.64 | 0.83 | 0.99 | 0.75 | | | LIV | | | Foxj2,SIX5 | | | 1 hit | 17kb 3' of AC010872.2 | |
| 2 | 21160407 | 0.85 | 1 | rs479413 | T | G | 0.62 | 0.80 | 0.96 | 0.73 | | | | | | Irf | | | 1 hit | 17kb 3' of AC010872.2 | |
| 2 | 21160562 | 0.86 | 1 | rs480488 | A | G | 0.63 | 0.80 | 0.98 | 0.73 | | | | | | Ets,ZBTB33 | | | 1 hit | 17kb 3' of AC010872.2 | |
| 2 | 21160642 | 0.86 | 1 | rs1712246 | G | A | 0.61 | 0.80 | 0.97 | 0.74 | | | | | | Crx,Pitx2,STAT | | | | 17kb 3' of AC010872.2 | |
| 2 | 21160652 | 0.96 | 1 | rs1652423 | G | A | 0.64 | 0.82 | 0.98 | 0.74 | | | | | | AP-1 | | | 1 hit | 17kb 3' of AC010872.2 | |
| 2 | 21160845 | 1 | 1 | rs4560142 | C | T | 0.64 | 0.83 | 0.99 | 0.75 | | | | | | HNF1 | | 1 hit | 1 hit | 18kb 3' of AC010872.2 | |
| 2 | 21160870 | 1 | 1 | rs4591370 | A | G | 0.64 | 0.83 | 0.99 | 0.75 | | | | | | ERalpha-a,Esr2,RXRA | | 1 hit | 1 hit | 18kb 3' of AC010872.2 | |
| 2 | 21160969 | 1 | 1 | rs1652422 | A | G | 0.64 | 0.83 | 0.99 | 0.75 | | | | | | | | | 1 hit | 18kb 3' of AC010872.2 | |
| 2 | 21160976 | 1 | 1 | rs1652421 | A | G | 0.64 | 0.83 | 0.99 | 0.75 | | | | | | | | | 1 hit | 18kb 3' of AC010872.2 | |
| 2 | 21161009 | 1 | 1 | rs1652420 | A | T | 0.64 | 0.83 | 0.99 | 0.75 | | | | | | CTCF | | | 1 hit | 18kb 3' of AC010872.2 | |
| 2 | 21161079 | 1 | 1 | rs1712247 | C | T | 0.64 | 0.83 | 0.99 | 0.75 | | | | | | Zfp187 | | 1 hit | 1 hit | 18kb 3' of AC010872.2 | |
| 2 | 21161110 | 0.98 | 1 | rs540897 | A | G | 0.64 | 0.82 | 0.99 | 0.75 | | | | | | Irf,SIX5 | | | 1 hit | 18kb 3' of AC010872.2 | |

| | | | | | | | | | | | | | | | | | |
|---|---|---|---|---|---|---|---|---|---|---|---|---|---|---|---|---|---|
| 2 | 21161400 | 1 | 1 | rs492255 | C | T | 0.64 | 0.83 | 0.99 | 0.75 | | | | 4 altered motifs | | 1 hit | 18kb 3' of AC010872.2 |
| 2 | 21161486 | 1 | 1 | rs544450 | T | C | 0.64 | 0.83 | 0.99 | 0.75 | | | | BDP1,Pax-6 | | 1 hit | 18kb 3' of AC010872.2 |
| 2 | 21161569 | 0.82 | 1 | rs544655 | G | T | 0.62 | 0.80 | 0.98 | 0.74 | | | | GR,Hoxa9 | | 1 hit | 18kb 3' of AC010872.2 |
| 2 | 21161790 | 0.96 | 1 | rs547179 | A | G | 0.63 | 0.82 | 0.99 | 0.75 | | | | | | 1 hit | 19kb 3' of AC010872.2 |
| 2 | 21161810 | 0.96 | 1 | rs547235 | A | G | 0.64 | 0.82 | 0.99 | 0.75 | | | | HDAC2,NRSF | | 1 hit | 19kb 3' of AC010872.2 |
| 2 | 21161814 | 0.94 | 1 | rs547239 | T | A | 0.63 | 0.82 | 0.99 | 0.75 | | | | HDAC2,NRSF | | 1 hit | 19kb 3' of AC010872.2 |
| 2 | 21162277 | 0.98 | 1 | rs572246 | T | C | 0.64 | 0.82 | 0.99 | 0.74 | | | | 11 altered motifs | | 1 hit | 19kb 3' of AC010872.2 |
| 2 | 21162289 | 1 | 1 | rs573314 | C | G | 0.64 | 0.83 | 0.99 | 0.75 | | | | 6 altered motifs | | 1 hit | 19kb 3' of AC010872.2 |
| 2 | 21162669 | 1 | 1 | rs548506 | A | G | 0.64 | 0.83 | 0.99 | 0.75 | SKIN | | | 4 altered motifs | | 1 hit | 19kb 3' of AC010872.2 |
| 2 | 21162777 | 1 | 1 | rs1652419 | C | T | 0.64 | 0.83 | 0.99 | 0.75 | | | | p300 | | 1 hit | 20kb 3' of AC010872.2 |
| 2 | 21162906 | 0.94 | 1 | rs1712248 | C | G | 0.52 | 0.82 | 0.99 | 0.74 | | | | YY1 | | 1 hit | 20kb 3' of AC010872.2 |
| 2 | 21163102 | 0.83 | 0.98 | rs1712249 | T | C | 0.68 | 0.80 | 0.99 | 0.71 | | | | Cart1,Hsf,RBP-Jkappa | | 2 hits | 20kb 3' of AC010872.2 |
| 2 | 21163186 | 0.98 | 1 | rs1712250 | C | T | 0.68 | 0.83 | 0.99 | 0.75 | ESDR | | | | | 1 hit | 20kb 3' of AC010872.2 |
| 2 | 21163432 | 0.98 | 1 | rs1367120 | T | C | 0.66 | 0.83 | 0.99 | 0.75 | ESDR | | | HDAC2 | | 1 hit | 20kb 3' of AC010872.2 |
| 2 | 21163503 | 0.98 | 1 | rs1367119 | A | C,G | 0.67 | 0.83 | 0.99 | 0.75 | ESDR | | | | | 1 hit | 20kb 3' of AC010872.2 |
| 2 | 21164085 | 0.96 | 0.98 | rs522963 | T | C | 0.67 | 0.83 | 0.99 | 0.75 | | | | 18 altered motifs | | 1 hit | 21kb 3' of AC010872.2 |
| 2 | 21164241 | 0.98 | 1 | rs522250 | T | C | 0.68 | 0.83 | 0.99 | 0.75 | | | | 6 altered motifs | | 1 hit | 21kb 3' of AC010872.2 |
| 2 | 21165076 | 0.98 | 1 | rs529697 | G | T | 0.68 | 0.83 | 0.99 | 0.75 | | | MAFK | 4 altered motifs | | 1 hit | 22kb 3' of AC010872.2 |
| 2 | 21165352 | 0.94 | 0.98 | rs490757 | C | T | 0.68 | 0.82 | 0.99 | 0.75 | LIV | | | 14 altered motifs | | 1 hit | 22kb 3' of AC010872.2 |
| 2 | 21165584 | 0.98 | 1 | rs1652418 | T | C | 0.68 | 0.83 | 0.99 | 0.75 | | 4 tissues | CTCF,RAD21 | Pax-1 | | 1 hit | 22kb 3' of AC010872.2 |
| 2 | 21166147 | 0.98 | 1 | rs538928 | A | G | 0.68 | 0.83 | 0.99 | 0.75 | | | | BRCA1 | 1 hit | 1 hit | 23kb 3' of AC010872.2 |
| 2 | 21166236 | 0.96 | 0.98 | rs560844 | A | G | 0.67 | 0.83 | 0.99 | 0.75 | | | | DMRT2,RXRA | | 1 hit | 23kb 3' of AC010872.2 |
| 2 | 21166558 | 0.98 | 1 | rs563696 | T | A | 0.68 | 0.83 | 0.99 | 0.75 | | | | HNF1,Nkx2,Nkx3 | | 1 hit | 23kb 3' of AC010872.2 |
| 2 | 21166613 | 0.98 | 1 | rs475887 | T | G | 0.68 | 0.83 | 0.99 | 0.75 | | | | 6 altered motifs | | 1 hit | 23kb 3' of AC010872.2 |
| 2 | 21167025 | 0.98 | 1 | rs479545 | T | C | 0.68 | 0.83 | 0.99 | 0.75 | | | | | | 1 hit | 24kb 3' of AC010872.2 |
| 2 | 21167128 | 0.98 | 1 | rs501863 | G | A | 0.68 | 0.83 | 0.99 | 0.75 | | | | Pou2f2 | | 1 hit | 24kb 3' of AC010872.2 |
| 2 | 21167277 | 0.98 | 1 | rs480732 | A | T | 0.68 | 0.83 | 0.99 | 0.75 | | | | PLZF,Pax-5,Zbtb12 | | 1 hit | 24kb 3' of AC010872.2 |
| 2 | 21167297 | 0.98 | 1 | rs480787 | A | G | 0.68 | 0.83 | 0.99 | 0.75 | | | | 4 altered motifs | | 1 hit | 24kb 3' of AC010872.2 |
| 2 | 21167437 | 0.96 | 1 | rs36058849 | TA | T | 0.69 | 0.83 | 0.99 | 0.75 | | | | 12 altered motifs | | 1 hit | 24kb 3' of AC010872.2 |
| 2 | 21167535 | 0.98 | 1 | rs483436 | G | A | 0.68 | 0.83 | 0.99 | 0.75 | | | | | | 1 hit | 24kb 3' of AC010872.2 |
| 2 | 21167802 | 0.98 | 1 | rs486139 | G | A | 0.68 | 0.83 | 0.99 | 0.75 | | | | 7 altered motifs | | 1 hit | 25kb 3' of AC010872.2 |
| 2 | 21168120 | 0.96 | 0.98 | rs489010 | G | A | 0.67 | 0.83 | 0.99 | 0.75 | | | | 4 altered motifs | | 1 hit | 25kb 3' of AC010872.2 |
| 2 | 21168659 | 0.98 | 1 | rs514757 | G | A | 0.68 | 0.83 | 0.99 | 0.75 | | | | Myb,RREB-1 | | 1 hit | 25kb 3' of AC010872.2 |
| 2 | 21168868 | 0.98 | 1 | rs538528 | T | C | 0.68 | 0.83 | 0.99 | 0.75 | | | | 9 altered motifs | | 1 hit | 26kb 3' of AC010872.2 |
| 2 | 21169020 | 0.98 | 1 | rs518280 | G | A | 0.68 | 0.83 | 0.99 | 0.75 | | | | E4BP4,Pax-3 | | 1 hit | 26kb 3' of AC010872.2 |
| 2 | 21169106 | 0.98 | 1 | rs540439 | C | T | 0.67 | 0.83 | 0.99 | 0.75 | | | | 4 altered motifs | | 1 hit | 26kb 3' of AC010872.2 |
| 2 | 21169281 | 0.98 | 1 | rs563280 | T | G | 0.68 | 0.83 | 0.99 | 0.75 | | | | 8 altered motifs | | 1 hit | 26kb 3' of AC010872.2 |
| 2 | 21169341 | 0.98 | 1 | rs564073 | A | T | 0.68 | 0.83 | 0.99 | 0.75 | | | | | | 1 hit | 26kb 3' of AC010872.2 |
| 2 | 21169357 | 0.98 | 1 | rs542261 | G | A | 0.69 | 0.83 | 0.99 | 0.75 | | | | Cdx | | 1 hit | 26kb 3' of AC010872.2 |
| 2 | 21169408 | 0.96 | 0.98 | rs34878249 | CT | C | 0.68 | 0.83 | 0.99 | 0.75 | | | | 10 altered motifs | | 1 hit | 26kb 3' of AC010872.2 |
| 2 | 21169554 | 0.98 | 1 | rs565894 | T | C | 0.67 | 0.83 | 0.99 | 0.75 | | | | Zec | | 1 hit | 26kb 3' of AC010872.2 |
| 2 | 21169698 | 0.98 | 1 | rs566913 | C | T | 0.68 | 0.83 | 0.99 | 0.75 | | | | 5 altered motifs | | 1 hit | 26kb 3' of AC010872.2 |
| 2 | 21169872 | 0.98 | 1 | rs568740 | C | T | 0.68 | 0.83 | 0.99 | 0.75 | | | | 5 altered motifs | | 1 hit | 27kb 3' of AC010872.2 |
| 2 | 21170046 | 0.98 | 1 | rs548594 | T | C | 0.68 | 0.83 | 0.99 | 0.75 | | | | 9 altered motifs | | 1 hit | 27kb 3' of AC010872.2 |
| 2 | 21170449 | 0.98 | 1 | rs484802 | C | T | 0.68 | 0.83 | 0.99 | 0.75 | | | | Gfi1,Gfi1b,RXRA | | 1 hit | 27kb 3' of AC010872.2 |
| 2 | 21170476 | 0.98 | 1 | rs484906 | G | T | 0.67 | 0.83 | 0.99 | 0.75 | | | | LBP-1 | | 1 hit | 27kb 3' of AC010872.2 |
| 2 | 21170751 | 0.98 | 1 | rs576203 | A | G | 0.68 | 0.83 | 0.99 | 0.75 | | | | GATA | 1 hit | 1 hit | 27kb 3' of AC010872.2 |
| 2 | 21170817 | 0.98 | 1 | rs488507 | G | T | 0.68 | 0.83 | 0.99 | 0.75 | | | | RFX5,TAL1 | 2 hits | 1 hit | 28kb 3' of AC010872.2 |
| 2 | 21170994 | 0.98 | 1 | rs578095 | A | G | 0.68 | 0.83 | 0.99 | 0.75 | | | | 5 altered motifs | | 1 hit | 28kb 3' of AC010872.2 |
| 2 | 21171065 | 0.98 | 1 | rs578864 | C | G | 0.68 | 0.83 | 0.99 | 0.75 | | | | Zfp187 | | 1 hit | 28kb 3' of AC010872.2 |
| 2 | 21171327 | 0.96 | 0.98 | rs492364 | C | T | 0.65 | 0.83 | 0.99 | 0.75 | | | | NF-I,Nr2e3 | | 1 hit | 28kb 3' of AC010872.2 |
| 2 | 21171329 | 0.96 | 0.98 | rs492365 | A | G | 0.67 | 0.83 | 0.99 | 0.75 | | | | 4 altered motifs | | 1 hit | 28kb 3' of AC010872.2 |
| 2 | 21171376 | 0.98 | 1 | rs492494 | C | T | 0.68 | 0.83 | 0.99 | 0.75 | | | | Pou2f2 | | 1 hit | 28kb 3' of AC010872.2 |
| 2 | 21171468 | 0.96 | 0.98 | rs493404 | G | A | 0.68 | 0.83 | 0.99 | 0.75 | | | | 7 altered motifs | | 1 hit | 28kb 3' of AC010872.2 |
| 2 | 21171586 | 0.98 | 1 | rs549959 | T | C | 0.68 | 0.83 | 0.99 | 0.75 | | | | 4 altered motifs | | 1 hit | 28kb 3' of AC010872.2 |
| 2 | 21171742 | 0.98 | 1 | rs496100 | A | G | 0.66 | 0.83 | 0.99 | 0.75 | | | | 4 altered motifs | | 1 hit | 28kb 3' of AC010872.2 |
| 2 | 21171926 | 0.98 | 1 | rs553523 | T | G | 0.68 | 0.83 | 0.99 | 0.75 | | | | FXR,Irf,Pou5f1 | | 1 hit | 29kb 3' of AC010872.2 |

| | | | | | | | | | | | | | | | | | | | |
|---|---|---|---|---|---|---|---|---|---|---|---|---|---|---|---|---|---|---|---|
| 2 | 21172020 | 0.98 | 1 | rs554414 | T | C | 0.67 | 0.83 | 0.99 | 0.75 | | | | | 8 altered motifs | | | 1 hit | 29kb 3' of AC010872.2 |
| 2 | 21172389 | 0.98 | 1 | rs556504 | C | G | 0.68 | 0.83 | 0.99 | 0.75 | | | | | EWSR1-FLI1 | | | 1 hit | 29kb 3' of AC010872.2 |
| 2 | 21172465 | 0.98 | 1 | rs557316 | A | G | 0.68 | 0.83 | 0.99 | 0.75 | | | | | Foxl1,Pou3f3 | | | 1 hit | 29kb 3' of AC010872.2 |
| 2 | 21172597 | 0.98 | 1 | rs558342 | G | A | 0.68 | 0.83 | 0.99 | 0.75 | | ESDR | | | | | | 1 hit | 29kb 3' of AC010872.2 |
| 2 | 21172726 | 0.98 | 1 | rs559967 | A | G | 0.68 | 0.83 | 0.99 | 0.75 | | ESDR | | | MZF1::1-4 | | | 1 hit | 29kb 3' of AC010872.2 |
| 2 | 21172933 | 0.98 | 1 | rs561850 | A | G | 0.68 | 0.83 | 0.99 | 0.75 | | | | | HNF4,PPAR,RXRA | | | 1 hit | 30kb 3' of AC010872.2 |
| 2 | 21172958 | 0.98 | 1 | rs473269 | T | C | 0.68 | 0.83 | 0.99 | 0.75 | | | | | | | | 1 hit | 30kb 3' of AC010872.2 |
| 2 | 21173129 | 0.98 | 1 | rs563719 | C | A | 0.68 | 0.83 | 0.99 | 0.75 | | | | | AP-1,Nkx2,RXRA | | | 1 hit | 30kb 3' of AC010872.2 |
| 2 | 21173141 | 0.98 | 1 | rs563752 | A | G | 0.68 | 0.83 | 0.99 | 0.75 | | | | | AP-1,BDP1,CEBPG | | | 1 hit | 30kb 3' of AC010872.2 |
| 2 | 21173462 | 0.98 | 1 | rs477146 | A | G | 0.68 | 0.83 | 0.99 | 0.75 | | | ESDR | | Pbx-1 | | | 1 hit | 30kb 3' of AC010872.2 |
| 2 | 21173616 | 0.98 | 1 | rs13411597 | G | A | 0.68 | 0.83 | 0.99 | 0.75 | | | | | HNF1,Obox3 | | | 1 hit | 30kb 3' of AC010872.2 |
| 2 | 21173645 | 0.98 | 1 | rs1652417 | T | C | 0.68 | 0.83 | 0.99 | 0.75 | | | | | 9 altered motifs | | | 1 hit | 30kb 3' of AC010872.2 |
| 2 | 21173679 | 0.98 | 1 | rs1712251 | G | A | 0.68 | 0.83 | 0.99 | 0.75 | | | GI | | 5 altered motifs | | | 1 hit | 30kb 3' of AC010872.2 |
| 2 | 21173737 | 0.98 | 1 | rs1652416 | A | G | 0.68 | 0.83 | 0.99 | 0.75 | | | GI | | Hoxa5,Sox | | | 1 hit | 30kb 3' of AC010872.2 |
| 2 | 21174163 | 0.98 | 1 | rs483621 | T | A | 0.68 | 0.83 | 0.99 | 0.75 | | | | | NRSF | | | 1 hit | 31kb 3' of AC010872.2 |
| 2 | 21174310 | 0.98 | 1 | rs506585 | G | A | 0.68 | 0.83 | 0.99 | 0.75 | | | | | | 3 hits | 1 hit | 1 hit | 31kb 3' of AC010872.2 |
| 2 | 21174449 | 0.98 | 1 | rs507616 | G | C | 0.67 | 0.83 | 0.99 | 0.75 | | | | | EBF | | | 1 hit | 31kb 3' of AC010872.2 |
| 2 | 21174689 | 0.98 | 1 | rs531380 | C | G | 0.68 | 0.83 | 0.99 | 0.75 | | | | | Rad21 | | | 1 hit | 31kb 3' of AC010872.2 |
| 2 | 21174765 | 0.98 | 1 | rs532225 | T | C | 0.68 | 0.83 | 0.99 | 0.75 | | | | | 5 altered motifs | | | 1 hit | 31kb 3' of AC010872.2 |
| 2 | 21175507 | 0.98 | 1 | rs560408 | T | C | 0.68 | 0.83 | 0.99 | 0.75 | | | | | DMRT1,DMRT7 | | | 1 hit | 32kb 3' of AC010872.2 |
| 2 | 21175896 | 0.98 | 1 | rs541569 | G | A | 0.68 | 0.83 | 0.99 | 0.75 | | BRN | BRN | CTCF | Pou1f1,Pou2f2,Pou3f3 | | | 1 hit | 33kb 3' of AC010872.2 |
| 2 | 21175985 | 0.93 | 0.98 | rs11321159 | GA | G | 0.67 | 0.82 | 0.99 | 0.75 | | ESDR, BRN | 14 tissues | CTCF,RAD21,SMC3 | 4 altered motifs | | | | 33kb 3' of AC010872.2 |
| 2 | 21176113 | 0.98 | 1 | rs544039 | C | A | 0.68 | 0.83 | 0.99 | 0.75 | | ESDR, BRN | 4 tissues | CTCF,RAD21,SMC3 | HDAC2,Hoxa3,Pou2f2 | | | 1 hit | 33kb 3' of AC010872.2 |
| 2 | 21176344 | 0.98 | 1 | rs478442 | G | T | 0.68 | 0.83 | 0.99 | 0.75 | | 7 tissues | ESC,LNG,OVRY | | Mef2 | | 1 hit | 1 hit | 33kb 3' of AC010872.2 |
| 2 | 21176695 | 0.98 | 1 | rs570033 | C | G | 0.68 | 0.83 | 0.99 | 0.75 | | 5 tissues | | | 4 altered motifs | | | 1 hit | 33kb 3' of AC010872.2 |
| 2 | 21177101 | 0.98 | 1 | rs66702176 | TC | T | 0.67 | 0.83 | 0.99 | 0.75 | | BRN, OVRY | | | 4 altered motifs | | | 1 hit | 34kb 3' of AC010872.2 |
| 2 | 21177141 | 0.98 | 1 | rs574461 | A | G | 0.68 | 0.83 | 0.99 | 0.75 | | ESDR, BRN, OVRY | | | | | | 1 hit | 34kb 3' of AC010872.2 |
| 2 | 21177433 | 0.98 | 1 | rs487858 | G | A | 0.68 | 0.83 | 0.99 | 0.75 | | BRN | | | 10 altered motifs | | | 1 hit | 34kb 3' of AC010872.2 |
| 2 | 21178078 | 0.94 | 0.98 | rs494315 | C | T | 0.68 | 0.83 | 0.99 | 0.75 | | SKIN, BRN | | | Arid5b,GATA,Hoxa7 | | | 1 hit | 35kb 3' of AC010872.2 |
| 2 | 21178138 | 0.94 | 0.98 | rs494465 | C | T | 0.67 | 0.83 | 0.99 | 0.75 | | SKIN | | | 4 altered motifs | | | 1 hit | 35kb 3' of AC010872.2 |
| 2 | 21178979 | 0.94 | 0.98 | rs522822 | A | G | 0.68 | 0.83 | 0.99 | 0.75 | | | | | 5 altered motifs | | | 1 hit | 36kb 3' of AC010872.2 |
| 2 | 21180425 | 0.94 | 0.98 | rs502323 | A | C | 0.68 | 0.83 | 0.99 | 0.75 | | ESDR | | | Foxp1,Ik-1,Pou3f2 | | | 1 hit | 37kb 3' of AC010872.2 |
| 2 | 21180609 | 0.94 | 0.98 | rs504091 | T | G | 0.69 | 0.83 | 0.99 | 0.75 | | ESDR | | | 4 altered motifs | | | 1 hit | 37kb 3' of AC010872.2 |
| 2 | 21183879 | 0.94 | 0.98 | rs486246 | G | A | 0.68 | 0.83 | 0.99 | 0.75 | | | PLCNT | | Hltf | | | 1 hit | 37kb 5' of AC067959.1 |
| 2 | 21185775 | 0.94 | 0.98 | rs312936 | T | A | 0.68 | 0.83 | 0.99 | 0.75 | | LNG | | | | | | 1 hit | 35kb 5' of AC067959.1 |
| 2 | 21190306 | 0.94 | 0.98 | rs142786027 | G | C | 0.68 | 0.83 | 0.99 | 0.75 | | | | | AP-1,p300 | | | 1 hit | 31kb 5' of AC067959.1 |
| 2 | 21190346 | 0.89 | 0.96 | rs141479646 | A | G | 0.67 | 0.82 | 0.99 | 0.75 | | | | | LBP-9 | | | 1 hit | 31kb 5' of AC067959.1 |
| 2 | 21191148 | 0.87 | 0.96 | rs504616 | C | G | 0.66 | 0.82 | 0.99 | 0.74 | | | | | 5 altered motifs | | | 1 hit | 30kb 5' of AC067959.1 |

Query SNP: rs10892984 and variants with $r^2$ >= 0.8

| chr | pos (hg38) | LD ($r^2$) | LD (D') | variant | Ref | Alt | AFR freq | AMR freq | ASN freq | EUR freq | SiPhy cons | Promoter histone marks | Enhancer histone marks | DNAse | Proteins bound | Motifs changed | NHGRI/EBI GWAS hits | GRASP QTL hits | Selected eQTL hits | GENCODE genes | dbSNP func annot |
|---|---|---|---|---|---|---|---|---|---|---|---|---|---|---|---|---|---|---|---|---|---|
| 11 | 123222490 | 0.87 | 0.95 | rs12272976 | C | T | 0.64 | 0.14 | 0.00 | 0.18 | | | FAT | | | Foxj1,Foxo,Sox | | | | CTD-2216M2.1 | |
| 11 | 123223442 | 1 | 1 | rs10892984 | A | T | 0.53 | 0.14 | 0.00 | 0.18 | | | | | | GATA,LBP-9 | | 1 hit | | CTD-2216M2.1 | |

Query SNP: rs10802219 and variants with $r^2$ >= 0.8

| chr | pos (hg38) | LD ($r^2$) | LD (D') | variant | Ref | Alt | AFR freq | AMR freq | ASN freq | EUR freq | SiPhy cons | Promoter histone marks | Enhancer histone marks | DNAse | Proteins bound | Motifs changed | NHGRI/EBI GWAS hits | GRASP QTL hits | Selected eQTL hits | GENCODE genes | dbSNP func annot |
|---|---|---|---|---|---|---|---|---|---|---|---|---|---|---|---|---|---|---|---|---|---|
| 1 | 245533008 | 0.92 | 0.98 | rs6677480 | G | A | 0.71 | 0.41 | 0.00 | 0.47 | | | | | | | | | | KIF26B | intronic |
| 1 | 245535783 | 1 | 1 | rs10802219 | G | A | 0.73 | 0.41 | 0.00 | 0.48 | | | | | | 5 altered motifs | | 2 hits | | KIF26B | intronic |
| 1 | 245535932 | 0.93 | 0.99 | rs7518092 | A | G | 0.67 | 0.42 | 0.00 | 0.49 | | | | | | Ets,Nrf-2,SRF | | | | KIF26B | intronic |

Query SNP: rs7698051 and variants with $r^2$ >= 0.8

| chr | pos (hg38) | LD ($r^2$) | LD (D') | variant | Ref | Alt | AFR freq | AMR freq | ASN freq | EUR freq | SiPhy cons | Promoter histone marks | Enhancer histone marks | DNAse | Proteins bound | Motifs changed | NHGRI/EBI GWAS hits | GRASP QTL hits | Selected eQTL hits | GENCODE genes | dbSNP func annot |
|---|---|---|---|---|---|---|---|---|---|---|---|---|---|---|---|---|---|---|---|---|---|
| 4 | 59406962 | 0.94 | 0.99 | rs2048464 | A | G | 0.38 | 0.67 | 0.37 | 0.63 | | | | | | Foxa,HNF1,STAT | | | | 127kb 5' of Y_RNA | |
| 4 | 59408325 | 0.96 | 0.99 | rs62305735 | C | T | 0.38 | 0.66 | 0.36 | 0.63 | | | | | | BDP1,ELF1,ERalpha-a | | | | 125kb 5' of Y_RNA | |
| 4 | 59408409 | 0.96 | 0.99 | rs62305736 | G | T | 0.39 | 0.66 | 0.36 | 0.63 | | | | | | DMRT1,DMRT5,DMRT7 | | | | 125kb 5' of Y_RNA | |
| 4 | 59408660 | 0.96 | 0.99 | rs6551786 | A | G | 0.38 | 0.66 | 0.36 | 0.63 | | | | | | DMRT2,DMRT4,YY1 | | | | 125kb 5' of Y_RNA | |

| | | | | | | | | | | | | | | | | | | | | |
|---|---|---|---|---|---|---|---|---|---|---|---|---|---|---|---|---|---|---|---|---|
| 4 | 59408988 | 0.98 | 1 | rs6817349 | A | T | 0.39 | 0.67 | 0.36 | 0.63 | | | | | | Ets,Pbx-1 | | | | 125kb 5' of Y_RNA |
| 4 | 59409351 | 0.98 | 1 | rs1079463 | T | C | 0.39 | 0.67 | 0.36 | 0.64 | | | | | | Foxj1,Foxo | | | | 124kb 5' of Y_RNA |
| 4 | 59410236 | 0.99 | 1 | rs6849275 | G | T | 0.39 | 0.66 | 0.36 | 0.64 | | | | | | 15 altered motifs | | | | 124kb 5' of Y_RNA |
| 4 | 59412766 | 1 | 1 | rs7698051 | A | G | 0.38 | 0.66 | 0.37 | 0.64 | | SKIN | | | | ERalpha-a,Gfi1 | | | | 121kb 5' of Y_RNA |
| 4 | 59414887 | 0.99 | 1 | rs953447 | C | T | 0.36 | 0.66 | 0.37 | 0.63 | | | | | | 5 altered motifs | | | | 119kb 5' of Y_RNA |
| 4 | 59420869 | 0.95 | 1 | rs10035014 | G | A | 0.38 | 0.65 | 0.37 | 0.63 | | | | | | Cphx,Sox | | | | 113kb 5' of Y_RNA |
| 4 | 59421593 | 0.99 | 1 | rs114684146 | C | A | 0.36 | 0.66 | 0.37 | 0.63 | | | | | | AP-1 | | | | 112kb 5' of Y_RNA |
| 4 | 59422893 | 0.99 | 1 | rs6811854 | G | C | 0.37 | 0.66 | 0.37 | 0.62 | | | | | | Cdx | | | | 111kb 5' of Y_RNA |
| 4 | 59423152 | 0.88 | 0.99 | rs114938864 | A | G | 0.38 | 0.68 | 0.38 | 0.62 | | | | | | 6 altered motifs | | | | 111kb 5' of Y_RNA |
| 4 | 59425783 | 0.82 | 0.97 | rs140767734 | 10-mer | T | 0.41 | 0.69 | 0.38 | 0.63 | | | | | | 11 altered motifs | | | | 108kb 5' of Y_RNA |
| 4 | 59429698 | 0.8 | 0.97 | rs2175440 | G | T | 0.40 | 0.70 | 0.38 | 0.64 | | | | | | | | | | 104kb 5' of Y_RNA |
| 4 | 59431205 | 0.95 | 0.98 | rs7356474 | G | T | 0.38 | 0.66 | 0.36 | 0.64 | | | | | | 4 altered motifs | | | | 103kb 5' of Y_RNA |
| 4 | 59431625 | 0.93 | 0.97 | rs116671113 | T | G | 0.37 | 0.67 | 0.37 | 0.63 | | | | | | 4 altered motifs | | | | 102kb 5' of Y_RNA |
| 4 | 59431836 | 0.86 | 0.97 | rs4449440 | G | A | 0.37 | 0.64 | 0.35 | 0.62 | | | | | | Pbx-1 | | | | 102kb 5' of Y_RNA |
| 4 | 59432688 | 0.82 | 0.96 | rs4370162 | C | T | 0.35 | 0.63 | 0.37 | 0.61 | | | | | | Pax-4,Pou5f1,TBX5 | | | | 101kb 5' of Y_RNA |
| 4 | 59432850 | 0.93 | 0.96 | rs6551799 | T | C | 0.37 | 0.66 | 0.37 | 0.64 | | | | | | | | | | 101kb 5' of Y_RNA |
| 4 | 59433942 | 0.92 | 0.96 | rs6817253 | G | A | 0.36 | 0.66 | 0.37 | 0.63 | | | | | | GATA,Hsf,Irf | | | | 100kb 5' of Y_RNA |
| 4 | 59436280 | 0.92 | 0.96 | rs10007393 | T | C | 0.38 | 0.66 | 0.37 | 0.64 | | | | | | 16 altered motifs | | | | 98kb 5' of Y_RNA |
| 4 | 59440286 | 0.8 | 0.95 | rs2341563 | C | T | 0.37 | 0.63 | 0.36 | 0.63 | | | | | | CIZ,Ik-2,Nkx3 | | | | 93kb 5' of Y_RNA |
| 4 | 59440471 | 0.9 | 0.95 | rs1355446 | A | G | 0.38 | 0.66 | 0.37 | 0.64 | | | | | | 5 altered motifs | | | | 93kb 5' of Y_RNA |
| 4 | 59440782 | 0.9 | 0.95 | rs2048462 | T | C | 0.38 | 0.66 | 0.36 | 0.64 | | | | | | 5 altered motifs | | 1 hit | | 93kb 5' of Y_RNA |
| 4 | 59441671 | 0.9 | 0.95 | rs6835115 | G | T | 0.38 | 0.66 | 0.37 | 0.64 | | | | | | STAT | | | | 92kb 5' of Y_RNA |
| 4 | 59445628 | 0.85 | 0.94 | rs1397808 | A | T | 0.36 | 0.65 | 0.37 | 0.64 | | | | | | 32 altered motifs | | | | 88kb 5' of Y_RNA |
| 4 | 59448053 | 0.8 | 0.91 | rs4860566 | T | G | 0.40 | 0.67 | 0.37 | 0.64 | | | | | | E2F | | | | 86kb 5' of Y_RNA |
| 4 | 59448520 | 0.8 | 0.91 | rs4860567 | G | A | 0.40 | 0.67 | 0.37 | 0.64 | | | | | | AP-1,PRDM1,Pax-4 | | | | 85kb 5' of Y_RNA |
| 4 | 59448610 | 0.82 | 0.91 | rs11735526 | T | A | 0.40 | 0.66 | 0.36 | 0.64 | | | | | | | | | | 85kb 5' of Y_RNA |
| 4 | 59448949 | 0.8 | 0.91 | rs35821878 | CAT | C | 0.39 | 0.65 | 0.36 | 0.63 | | | | | | Foxj1,Foxp1,HNF1 | | | | 85kb 5' of Y_RNA |
| 4 | 59449187 | 0.83 | 0.92 | rs6826593 | T | C | 0.40 | 0.67 | 0.37 | 0.64 | | | | | | 4 altered motifs | | 1 hit | | 85kb 5' of Y_RNA |
| 4 | 59449407 | 0.82 | 0.91 | rs6827039 | T | C | 0.39 | 0.66 | 0.37 | 0.64 | | | ESC | | | 4 altered motifs | | | | 84kb 5' of Y_RNA |
| 4 | 59449647 | 0.82 | 0.91 | rs6827654 | T | A | 0.40 | 0.66 | 0.37 | 0.64 | | | ESC | | | 6 altered motifs | | | | 84kb 5' of Y_RNA |
| 4 | 59449817 | 0.82 | 0.91 | rs1018263 | T | G | 0.42 | 0.66 | 0.37 | 0.64 | | | | | | | | | | 84kb 5' of Y_RNA |
| 4 | 59450819 | 0.85 | 0.93 | rs1018264 | C | T | 0.40 | 0.66 | 0.37 | 0.64 | | | | | | | | | | 83kb 5' of Y_RNA |
| 4 | 59451743 | 0.82 | 0.91 | rs1355447 | G | A | 0.39 | 0.66 | 0.37 | 0.63 | | | | | | Sin3Ak-20,TCF12,Znf143 | | | | 82kb 5' of Y_RNA |
| 4 | 59453203 | 0.84 | 0.91 | rs4241647 | C | T | 0.40 | 0.66 | 0.37 | 0.64 | | | | | | | | | | 81kb 5' of Y_RNA |
| 4 | 59456021 | 0.86 | 0.94 | rs7665683 | C | T | 0.36 | 0.65 | 0.37 | 0.63 | | | | | | CTCF,Evi-1,Foxj2 | | | | 78kb 5' of Y_RNA |
| 4 | 59456912 | 0.84 | 0.91 | rs7676524 | A | C | 0.40 | 0.66 | 0.37 | 0.63 | | | | | | 7 altered motifs | | | | 77kb 5' of Y_RNA |

Query SNP: rs188384 and variants with $r^2$ >= 0.8

| chr | pos (hg38) | LD ($r^2$) | LD (D') | variant | Ref | Alt | AFR freq | AMR freq | ASN freq | EUR freq | SiPhy cons | Promoter histone marks | Enhancer histone marks | DNAse | Proteins bound | Motifs changed | NHGRI/ EBI GWAS hits | GRASP QTL hits | Selected eQTL hits | GENCODE genes | dbSNP func annot |
|---|---|---|---|---|---|---|---|---|---|---|---|---|---|---|---|---|---|---|---|---|---|
| 3 | 191357084 | 1 | 1 | rs188384 | C | G | 0.90 | 0.67 | 0.92 | 0.54 | | 7 tissues | 17 tissues | 21 tissues | CFOS,AP2GAMMA,RAD21 | 4 altered motifs | | 2 hits | 15 hits | CCDC50 | intronic |
| 3 | 191363417 | 0.99 | 1 | rs293802 | G | T | 0.83 | 0.67 | 0.92 | 0.55 | | | 7 tissues | 7 tissues | STAT3 | 5 altered motifs | | 1 hit | 14 hits | CCDC50 | intronic |
| 3 | 191365046 | 0.99 | 1 | rs293800 | C | A | 0.85 | 0.67 | 0.91 | 0.55 | | | ESC, IPSC, VAS | | | 8 altered motifs | | 1 hit | 14 hits | CCDC50 | intronic |
| 3 | 191367835 | 1 | 1 | rs437244 | G | A | 0.90 | 0.67 | 0.91 | 0.55 | | | 4 tissues | | | 12 altered motifs | | | 13 hits | CCDC50 | intronic |
| 3 | 191369762 | 0.99 | 1 | rs697905 | G | A | 0.85 | 0.67 | 0.91 | 0.55 | | ESC | 7 tissues | 14 tissues | CEBPB,P300 | CEBPB,HNF4,Hand1 | | | 14 hits | CCDC50 | intronic |
| 3 | 191372984 | 0.99 | 1 | rs293804 | G | A | 0.90 | 0.67 | 0.91 | 0.55 | | | MUS, SKIN | | | Pou2f2 | | | 13 hits | CCDC50 | intronic |
| 3 | 191373570 | 0.99 | 1 | rs293805 | G | A | 0.85 | 0.67 | 0.91 | 0.55 | | | 7 tissues | 4 tissues | | 4 altered motifs | | | 14 hits | CCDC50 | intronic |
| 3 | 191374467 | 1 | 1 | rs186063 | T | C | 0.90 | 0.67 | 0.91 | 0.54 | | | 5 tissues | | | Bbx,Pax-4 | | 1 hit | 14 hits | CCDC50 | intronic |
| 3 | 191375645 | 0.99 | 1 | rs293806 | A | C,G,T | 0.85 | 0.67 | 0.91 | 0.55 | | | 5 tissues | | | | | | 14 hits | CCDC50 | intronic |
| 3 | 191376870 | 0.95 | 0.99 | rs176820 | A | C | 0.83 | 0.67 | 0.93 | 0.55 | | 8 tissues | 14 tissues | 20 tissues | CFOS,AP2GAMMA,KAP1 | 5 altered motifs | | | 13 hits | CCDC50 | intronic |
| 3 | 191377834 | 0.99 | 1 | rs367747 | A | G | 0.85 | 0.67 | 0.91 | 0.55 | | | 15 tissues | | | 8 altered motifs | | | 14 hits | CCDC50 | intronic |
| 3 | 191378827 | 1 | 1 | rs186066 | T | A | 0.90 | 0.67 | 0.91 | 0.55 | | HRT | 13 tissues | | | | | | 14 hits | CCDC50 | intronic |
| 3 | 191379593 | 0.99 | 1 | rs293808 | T | A | 0.85 | 0.67 | 0.91 | 0.55 | | | 12 tissues | BLD,THYM | | AIRE,NF-kappaB,PU.1 | | | 14 hits | CCDC50 | intronic |
| 3 | 191379626 | 0.99 | 1 | rs390744 | G | C | 0.85 | 0.67 | 0.91 | 0.55 | | | 12 tissues | BLD,BLD,THYM | | NF-AT,Pax-5,SPIB | | | 14 hits | CCDC50 | intronic |
| 3 | 191381679 | 0.98 | 0.99 | rs432085 | T | C | 0.90 | 0.67 | 0.91 | 0.55 | | | 5 tissues | | | 4 altered motifs | | 1 hit | 14 hits | CCDC50 | intronic |
| 3 | 191385125 | 0.98 | -0.99 | rs368665 | C | T | 0.10 | 0.33 | 0.09 | 0.45 | | | MUS, SKIN | | | | | | 13 hits | CCDC50 | intronic |
| 3 | 191385958 | 0.96 | -0.99 | rs369174 | A | T | 0.15 | 0.33 | 0.09 | 0.45 | | | SKIN | | | 4 altered motifs | | | 15 hits | CCDC50 | intronic |
| 3 | 191386078 | 0.96 | -0.99 | rs434556 | G | A | 0.15 | 0.33 | 0.09 | 0.45 | | | SKIN | | | Foxp1,Sox | | | 14 hits | CCDC50 | intronic |
| 3 | 191386514 | 0.96 | -0.99 | rs383075 | C | T | 0.15 | 0.33 | 0.09 | 0.45 | | | | | | | | | 14 hits | CCDC50 | intronic |
| 3 | 191388786 | 0.94 | -0.99 | rs566548 | T | C | 0.15 | 0.34 | 0.09 | 0.45 | | | | | | | | | 14 hits | CCDC50 | intronic |
| 3 | 191392531 | 0.96 | -0.99 | rs529523 | G | A | 0.15 | 0.33 | 0.09 | 0.45 | | | 6 tissues | ESDR,ESC,SKIN | | 5 altered motifs | | | 13 hits | CCDC50 | 3'-UTR |
| 3 | 191392755 | 0.96 | -0.99 | rs150504 | T | G | 0.15 | 0.33 | 0.09 | 0.45 | | | 5 tissues | SKIN | | 7 altered motifs | | | 13 hits | CCDC50 | 3'-UTR |
| 3 | 191398344 | 0.95 | -0.99 | rs6953 | C | T | 0.16 | 0.33 | 0.09 | 0.45 | | | | | | Fox,Foxp1 | | 1 hit | 14 hits | CCDC50 | 3'-UTR |
| 3 | 191399496 | 0.82 | -0.97 | rs188385 | C | T | 0.10 | 0.30 | 0.09 | 0.42 | | | | | | 4 altered motifs | | | 12 hits | 825bp 3' of CCDC50 | |
| 3 | 191400736 | 0.98 | -0.99 | rs709142 | T | C | 0.10 | 0.33 | 0.09 | 0.45 | | | | | | CDP,HNF6,Hoxa9 | | 1 hit | 14 hits | 2.1kb 3' of CCDC50 | |
| 3 | 191401904 | 0.96 | -0.99 | rs381090 | G | A | 0.10 | 0.33 | 0.09 | 0.46 | | | BLD | | ZNF263 | GLI,RREB-1,RXRA | | | 13 hits | 3.2kb 3' of CCDC50 | |
| 3 | 191405638 | 0.94 | -0.97 | rs293851 | T | C | 0.10 | 0.33 | 0.09 | 0.45 | | | | | | | | | 14 hits | 7kb 3' of CCDC50 | |

| | | | | | | | | | | | | | | | | | | | | |
|---|---|---|---|---|---|---|---|---|---|---|---|---|---|---|---|---|---|---|---|---|
| 3 | 191409527 | 0.95 | -0.97 | rs150506 | G | T | 0.10 | 0.33 | 0.09 | 0.45 | | | | | | Zbtb12 | | | 12 hits | 11kb 3' of CCDC50 |
| 3 | 191409994 | 0.95 | -0.97 | rs373035 | A | G | 0.10 | 0.33 | 0.09 | 0.45 | | | | | KAP1,SETDB1 | MOVO-B,Spz1 | | 3 hits | 13 hits | 11kb 3' of CCDC50 |
| 3 | 191411826 | 0.81 | -0.97 | rs426466 | G | C | 0.28 | 0.36 | 0.09 | 0.46 | | | | | | RXRA | | | 12 hits | 13kb 3' of CCDC50 |
| 3 | 191414027 | 0.95 | -0.97 | rs159489 | C | T | 0.11 | 0.33 | 0.09 | 0.45 | | | 4 tissues | | | 6 altered motifs | | | 12 hits | 15kb 3' of CCDC50 |
| 3 | 191414157 | 0.95 | -0.97 | rs293844 | C | T | 0.10 | 0.33 | 0.09 | 0.45 | | | 4 tissues | | | 4 altered motifs | | 3 hits | 13 hits | 15kb 3' of CCDC50 |
| 3 | 191414557 | 0.95 | -0.97 | rs200157727 | GAGT | G | 0.10 | 0.33 | 0.09 | 0.45 | | | 6 tissues | SKIN,SKIN | | 34 altered motifs | | | 13 hits | 16kb 3' of CCDC50 |
| 3 | 191416524 | 0.94 | -0.97 | rs293846 | C | A | 0.10 | 0.33 | 0.09 | 0.45 | | | | | | CEBPA | | 2 hits | 14 hits | 18kb 3' of CCDC50 |
| 3 | 191416719 | 0.95 | -0.97 | rs526785 | T | C | 0.10 | 0.33 | 0.09 | 0.45 | | | | | | 5 altered motifs | | 2 hits | 14 hits | 18kb 3' of CCDC50 |
| 3 | 191416844 | 0.95 | -0.97 | rs525774 | G | A | 0.10 | 0.33 | 0.09 | 0.45 | | | ESDR, HRT | SKIN | | Pax-4,Pax-5 | | | 13 hits | 18kb 3' of CCDC50 |

Query SNP: rs9973340 and variants with $r^2$ >= 0.8

| chr | pos (hg38) | LD ($r^2$) | LD (D') | variant | Ref | Alt | AFR freq | AMR freq | ASN freq | EUR freq | SiPhy cons | Promoter histone marks | Enhancer histone marks | DNAse | Proteins bound | Motifs changed | NHGRI/EBI GWAS hits | GRASP QTL hits | Selected eQTL hits | GENCODE genes | dbSNP func annot |
|---|---|---|---|---|---|---|---|---|---|---|---|---|---|---|---|---|---|---|---|---|---|
| 2 | 4418948 | 0.99 | -1 | rs688917 | C | G | 0.83 | 0.45 | 0.78 | 0.46 | | | | | | LUN-1 | | | | 190kb 5' of AC068292.1 | |
| 2 | 4421542 | 1 | 1 | rs9973340 | C | T | 0.12 | 0.55 | 0.22 | 0.54 | | | ESDR, VAS | VAS | CFOS | BDP1 | | | | 193kb 5' of AC068292.1 | |
| 2 | 4423279 | 0.97 | -1 | rs639159 | A | T | 0.74 | 0.44 | 0.78 | 0.46 | | | | | | GR | | | | 195kb 5' of AC068292.1 | |
| 2 | 4423548 | 0.9 | -1 | rs2677912 | A | G | 0.54 | 0.43 | 0.64 | 0.44 | | | | | | ERalpha-a,GCNF | | | | 195kb 5' of AC068292.1 | |
| 2 | 4424366 | 0.9 | -1 | rs669557 | A | G | 0.74 | 0.43 | 0.78 | 0.47 | | | | | MAFF,MAFK | 7 altered motifs | | | | 196kb 5' of AC068292.1 | |
| 2 | 4425455 | 0.86 | -0.98 | rs664699 | A | G | 0.80 | 0.43 | 0.77 | 0.44 | | | | | | Pbx3,VDR | | | | 197kb 5' of AC068292.1 | |

Query SNP: rs9561487 and variants with $r^2$ >= 0.8

| chr | pos (hg38) | LD ($r^2$) | LD (D') | variant | Ref | Alt | AFR freq | AMR freq | ASN freq | EUR freq | SiPhy cons | Promoter histone marks | Enhancer histone marks | DNAse | Proteins bound | Motifs changed | NHGRI/EBI GWAS hits | GRASP QTL hits | Selected eQTL hits | GENCODE genes | dbSNP func annot |
|---|---|---|---|---|---|---|---|---|---|---|---|---|---|---|---|---|---|---|---|---|---|
| 13 | 94000068 | 1 | 1 | rs9561487 | T | C | 0.17 | 0.53 | 0.43 | 0.54 | | | | | | TATA | | | | GPC6 | intronic |
| 13 | 94001264 | 0.95 | 1 | rs9556346 | C | T | 0.14 | 0.52 | 0.43 | 0.54 | | | | | | CTCF,GATA,TAL1 | | | | GPC6 | intronic |
| 13 | 94001622 | 0.98 | 1 | rs4773770 | G | A | 0.17 | 0.52 | 0.43 | 0.54 | | | | | | Hic1 | | | | GPC6 | intronic |
| 13 | 94014060 | 0.92 | 0.99 | rs9556347 | C | T | 0.17 | 0.52 | 0.44 | 0.54 | | | 4 tissues | | | 5 altered motifs | | | | GPC6 | intronic |
| 13 | 94014905 | 0.86 | -0.97 | rs9301925 | C | T | 0.78 | 0.49 | 0.56 | 0.49 | | | | | | Dobox4,Mef2 | | | | GPC6 | intronic |
| 13 | 94014969 | 0.81 | 0.98 | rs9561489 | G | A | 0.21 | 0.49 | 0.36 | 0.49 | | | | | | Pbx3 | | | | GPC6 | intronic |

Query SNP: rs17127435 and variants with $r^2$ >= 0.8

| chr | pos (hg38) | LD ($r^2$) | LD (D') | variant | Ref | Alt | AFR freq | AMR freq | ASN freq | EUR freq | SiPhy cons | Promoter histone marks | Enhancer histone marks | DNAse | Proteins bound | Motifs changed | NHGRI/EBI GWAS hits | GRASP QTL hits | Selected eQTL hits | GENCODE genes | dbSNP func annot |
|---|---|---|---|---|---|---|---|---|---|---|---|---|---|---|---|---|---|---|---|---|---|
| 11 | 123523557 | 0.86 | 0.97 | rs4936820 | C | T | 0.20 | 0.13 | 0.45 | 0.15 | | | ADRL | | | 6 altered motifs | | | | 2.1kb 5' of GRAMD1B | |
| 11 | 123524640 | 1 | 1 | rs17127435 | A | G | 0.10 | 0.12 | 0.44 | 0.14 | | | 6 tissues | | | Pax-3,Pax-5,p300 | | 1 hit | | 995bp 5' of GRAMD1B | |

Query SNP: rs2019917 and variants with $r^2$ >= 0.8

| chr | pos (hg38) | LD ($r^2$) | LD (D') | variant | Ref | Alt | AFR freq | AMR freq | ASN freq | EUR freq | SiPhy cons | Promoter histone marks | Enhancer histone marks | DNAse | Proteins bound | Motifs changed | NHGRI/EBI GWAS hits | GRASP QTL hits | Selected eQTL hits | GENCODE genes | dbSNP func annot |
|---|---|---|---|---|---|---|---|---|---|---|---|---|---|---|---|---|---|---|---|---|---|
| 18 | 25634698 | 0.92 | 0.97 | rs2592061 | T | C | 0.28 | 0.54 | 0.60 | 0.67 | | | | BLD | | 4 altered motifs | | | | 184kb 3' of Metazoa_SRP | |
| 18 | 25638032 | 0.93 | 0.97 | rs2604480 | T | C | 0.27 | 0.53 | 0.60 | 0.67 | | | ESDR | | | GATA,TAL1 | | | | 180kb 3' of Metazoa_SRP | |
| 18 | 25638120 | 0.93 | 0.97 | rs2604481 | T | C | 0.28 | 0.53 | 0.60 | 0.67 | | | | | | DMRT1,DMRT3,Irf | | | | 180kb 3' of Metazoa_SRP | |
| 18 | 25640108 | 0.93 | 0.97 | rs2728505 | C | A | 0.27 | 0.53 | 0.60 | 0.67 | | | LNG, MUS, SKIN | | | Myf,Sin3Ak-20 | | | | 178kb 3' of Metazoa_SRP | |
| 18 | 25646896 | 1 | 1 | rs2019917 | T | C | 0.27 | 0.53 | 0.60 | 0.67 | | | 6 tissues | BRN | | 5 altered motifs | | | | 171kb 3' of Metazoa_SRP | |
| 18 | 25650522 | 0.89 | 1 | rs1840437 | G | C | 0.70 | 0.56 | 0.61 | 0.67 | | 6 tissues | 8 tissues | 16 tissues | | | | | | 168kb 3' of Metazoa_SRP | |
| 18 | 25651500 | 1 | 1 | rs1992726 | A | G | 0.28 | 0.53 | 0.59 | 0.67 | | ESDR | 9 tissues | MUS | | GR,Myc | | | | 167kb 3' of Metazoa_SRP | |
| 18 | 25657408 | 0.98 | 1 | rs1455188 | C | T | 0.19 | 0.53 | 0.59 | 0.67 | | | | | | | | | | 161kb 3' of Metazoa_SRP | |
| 18 | 25660690 | 0.97 | 1 | rs12456784 | C | T | 0.19 | 0.52 | 0.59 | 0.66 | | | | | | | | | | 158kb 3' of Metazoa_SRP | |
| 18 | 25668147 | 0.88 | 1 | rs11083145 | T | C | 0.20 | 0.50 | 0.47 | 0.65 | | | | | | GR,Irx,NR4A | | 1 hit | | 150kb 3' of Metazoa_SRP | |
| 18 | 25677062 | 0.81 | 0.93 | rs1840441 | G | A | 0.45 | 0.52 | 0.49 | 0.65 | | | ESC | | | Pou2f2,Pou3f3 | | | | 141kb 3' of Metazoa_SRP | |
| 18 | 25677222 | 0.88 | 1 | rs1840440 | T | C | 0.20 | 0.50 | 0.47 | 0.65 | | | ESC | | | ZEB1 | 1 hit | 1 hit | | 141kb 3' of Metazoa_SRP | |
| 18 | 25681256 | 0.88 | 1 | rs965178 | T | C | 0.22 | 0.50 | 0.47 | 0.65 | | | ESDR | | | AP-4,E2A,LBP-1 | | | | 137kb 3' of Metazoa_SRP | |
| | | 0.8 | 0.92 | rs79002705 | T | A | 0.50 | 0.52 | 0.49 | 0.65 | | | ESC | | | | | | | 133kb 3' of Metazoa_SRP | |
| 18 | 25691322 | 0.86 | 0.99 | rs12955389 | T | C | 0.22 | 0.50 | 0.47 | 0.66 | | | | | | Pou2f2,Sox,Zfp105 | | | | 127kb 3' of Metazoa_SRP | |

Query SNP: rs810083 and variants with $r^2$ >= 0.8

| chr | pos (hg38) | LD ($r^2$) | LD (D') | variant | Ref | Alt | AFR freq | AMR freq | ASN freq | EUR freq | SiPhy cons | Promoter histone marks | Enhancer histone marks | DNAse | Proteins bound | Motifs changed | NHGRI/EBI GWAS hits | GRASP QTL hits | Selected eQTL hits | GENCODE genes | dbSNP func annot |
|---|---|---|---|---|---|---|---|---|---|---|---|---|---|---|---|---|---|---|---|---|---|
| 10 | 78899394 | 1 | 1 | rs810083 | T | C | 0.59 | 0.32 | 0.66 | 0.44 | | | 4 tissues | | | 7 altered motifs | | | | 44kb 3' of RP11-202P11.1 | |
| 10 | 78900209 | 0.85 | 0.97 | rs2813479 | C | T | 0.87 | 0.35 | 0.66 | 0.42 | | | 10 tissues | 20 tissues | | ELF1,NERF1a,STAT | | | 1 hit | 43kb 3' of RP11-202P11.1 | |

| | | | | | | | | | | | | | | | | | | | | |
|---|---|---|---|---|---|---|---|---|---|---|---|---|---|---|---|---|---|---|---|---|
| 10 | 78900849 | 1 | 1 | rs2559797 | A | T | 0.61 | 0.32 | 0.66 | 0.44 | | | OVRY | | | 14 altered motifs | | | | 42kb 3' of RP11-202P11.1 |
| 10 | 78903430 | 0.95 | 0.99 | rs2067528 | G | A | 0.59 | 0.31 | 0.66 | 0.42 | | | | | | ATF3,XBP-1 | | | | 40kb 3' of RP11-202P11.1 |
| 10 | 78906933 | 0.84 | 0.96 | rs2559794 | C | A | 0.80 | 0.34 | 0.66 | 0.41 | | | 4 tissues | | | 5 altered motifs | | | 2 hits | 36kb 3' of RP11-202P11.1 |

Query SNP: rs1192439 and variants with $r^2$ >= 0.8

| chr | pos (hg38) | LD ($r^2$) | LD (D') | variant | Ref | Alt | AFR freq | AMR freq | ASN freq | EUR freq | SiPhy cons | Promoter histone marks | Enhancer histone marks | DNAse | Proteins bound | Motifs changed | NHGRI/EBI GWAS hits | GRASP QTL hits | Selected eQTL hits | GENCODE genes | dbSNP func annot |
|---|---|---|---|---|---|---|---|---|---|---|---|---|---|---|---|---|---|---|---|---|---|
| 6 | 61972862 | 1 | 1 | rs1192439 | T | A | 0.10 | 0.28 | 0.24 | 0.37 | | | | | | Pax-5 | | | 1 hit | KHDRBS2 | intronic |
| 6 | 61979974 | 0.99 | 1 | rs1192427 | G | A | 0.09 | 0.28 | 0.24 | 0.37 | | | | | | 7 altered motifs | | | 1 hit | KHDRBS2 | intronic |

Query SNP: rs17559708 and variants with $r^2$ >= 0.8

| chr | pos (hg38) | LD ($r^2$) | LD (D') | variant | Ref | Alt | AFR freq | AMR freq | ASN freq | EUR freq | SiPhy cons | Promoter histone marks | Enhancer histone marks | DNAse | Proteins bound | Motifs changed | NHGRI/EBI GWAS hits | GRASP QTL hits | Selected eQTL hits | GENCODE genes | dbSNP func annot |
|---|---|---|---|---|---|---|---|---|---|---|---|---|---|---|---|---|---|---|---|---|---|
| 5 | 169788665 | 1 | 1 | rs17559708 | T | C | 0.03 | 0.19 | 0.07 | 0.32 | | | 6 tissues | MUS | | SIX5,Zfp410 | | | | DOCK2 | intronic |

Query SNP: rs10058089 and variants with $r^2$ >= 0.8

| chr | pos (hg38) | LD ($r^2$) | LD (D') | variant | Ref | Alt | AFR freq | AMR freq | ASN freq | EUR freq | SiPhy cons | Promoter histone marks | Enhancer histone marks | DNAse | Proteins bound | Motifs changed | NHGRI/EBI GWAS hits | GRASP QTL hits | Selected eQTL hits | GENCODE genes | dbSNP func annot |
|---|---|---|---|---|---|---|---|---|---|---|---|---|---|---|---|---|---|---|---|---|---|
| 5 | 111644925 | 0.91 | 0.99 | rs10073336 | A | G | 0.26 | 0.28 | 0.01 | 0.29 | | | | | | Pax-4,Pou5f1 | | | | CTC-426B10.1 | intronic |
| 5 | 111645412 | 0.9 | 0.97 | rs13177489 | A | G | 0.27 | 0.28 | 0.01 | 0.29 | | | LIV, MUS | | | GATA | | | | CTC-426B10.1 | intronic |
| 5 | 111648933 | 0.96 | 1 | rs13354235 | A | G | 0.38 | 0.28 | 0.01 | 0.29 | | | | | | FAC1,Mef2,SIX5 | | | | CTC-426B10.1 | intronic |
| 5 | 111649201 | 0.99 | 1 | rs13157350 | G | C | 0.43 | 0.29 | 0.07 | 0.29 | | | STRM | | | Hoxa5,SP1 | | | | CTC-426B10.1 | intronic |
| 5 | 111651657 | 0.93 | 1 | rs7725162 | G | A | 0.27 | 0.28 | 0.01 | 0.28 | | | LIV | | | NRSF,PLAG1,Zfx | | | | CTC-426B10.1 | intronic |
| 5 | 111653950 | 0.97 | 0.99 | rs1840979 | G | C | 0.42 | 0.29 | 0.07 | 0.29 | | | LIV | | | | | | | CTC-426B10.1 | intronic |
| 5 | 111654561 | 0.97 | 0.99 | rs1379551 | T | G | 0.43 | 0.29 | 0.07 | 0.30 | | | LIV | | | 9 altered motifs | | | | CTC-426B10.1 | intronic |
| 5 | 111655104 | 1 | 1 | rs10058089 | G | C | 0.43 | 0.29 | 0.07 | 0.29 | | | | 4 tissues | | 4 altered motifs | | | | CTC-426B10.1 | intronic |
| 5 | 111656499 | 0.92 | 0.99 | rs17549632 | G | A | 0.26 | 0.28 | 0.01 | 0.29 | | | LIV | | | 29 altered motifs | | | | CTC-426B10.1 | intronic |
| 5 | 111656828 | 0.96 | 0.99 | rs13182708 | G | A | 0.42 | 0.29 | 0.07 | 0.29 | | | LIV | GI | | Cdx2,Hoxd10,Sox | | | | CTC-426B10.1 | intronic |

Query SNP: rs11642880 and variants with $r^2$ >= 0.8

| chr | pos (hg38) | LD ($r^2$) | LD (D') | variant | Ref | Alt | AFR freq | AMR freq | ASN freq | EUR freq | SiPhy cons | Promoter histone marks | Enhancer histone marks | DNAse | Proteins bound | Motifs changed | NHGRI/EBI GWAS hits | GRASP QTL hits | Selected eQTL hits | GENCODE genes | dbSNP func annot |
|---|---|---|---|---|---|---|---|---|---|---|---|---|---|---|---|---|---|---|---|---|---|
| 16 | 11673199 | 0.83 | 0.95 | rs8191298 | C | A | 0.05 | 0.22 | 0.27 | 0.47 | | 6 tissues | 20 tissues | 9 tissues | | Glis2,RXRA,Smad | | | 6 hits | SNN | intronic |
| 16 | 11676092 | 0.83 | 0.95 | rs1050068 | C | T | 0.04 | 0.22 | 0.27 | 0.48 | | | | ESDR,OVRY,MUS | POL2 | CACD,Klf7 | | | 6 hits | SNN | synonymous |
| 16 | 11676107 | 0.82 | 0.95 | rs1050069 | C | T | 0.07 | 0.22 | 0.27 | 0.48 | | | | 4 tissues | POL2 | Nanog,Sox | | | 7 hits | SNN | synonymous |
| 16 | 11685462 | 0.83 | 0.95 | rs34340800 | G | A | 0.04 | 0.22 | 0.27 | 0.47 | | | BLD, MUS | | | | | | 6 hits | TXNDC11 | intronic |
| 16 | 11687726 | 0.82 | 0.95 | rs12920377 | A | C | 0.07 | 0.22 | 0.28 | 0.48 | | | 10 tissues | 5 tissues | | 7 altered motifs | | 4 hits | 7 hits | TXNDC11 | intronic |
| 16 | 11701723 | 0.9 | 0.95 | rs11645506 | C | A | 0.10 | 0.21 | 0.18 | 0.43 | | | BLD, GI, LIV | SKIN | | 4 altered motifs | | 2 hits | 4 hits | TXNDC11 | intronic |
| 16 | 11712156 | 0.87 | 0.96 | rs12149831 | A | G | 0.02 | 0.20 | 0.19 | 0.39 | | | 4 tissues | | | AP-1,Mef2 | | | 3 hits | TXNDC11 | intronic |
| 16 | 11717311 | 0.95 | 0.98 | rs11861532 | G | A | 0.04 | 0.20 | 0.19 | 0.42 | | | GI, BLD, LIV | | | 6 altered motifs | | | 3 hits | TXNDC11 | intronic |
| 16 | 11717933 | 0.98 | 1 | rs11644248 | C | T | 0.04 | 0.21 | 0.19 | 0.42 | | | 6 tissues | IPSC,BLD | | 4 altered motifs | | | 3 hits | TXNDC11 | intronic |
| 16 | 11727598 | 0.84 | 1 | rs10492850 | G | A | 0.03 | 0.18 | 0.19 | 0.37 | | | | | | Foxm1 | | | 4 hits | TXNDC11 | intronic |
| 16 | 11727652 | 0.95 | 1 | rs12919035 | A | G | 0.07 | 0.22 | 0.20 | 0.43 | | | BLD, LIV | BLD | | HMG-IY,Pax-4,SETDB1 | | | 3 hits | TXNDC11 | intronic |
| 16 | 11727852 | 1 | 1 | rs10492852 | G | A | 0.06 | 0.21 | 0.19 | 0.42 | | | | | | TCF4 | | 1 hit | 4 hits | TXNDC11 | intronic |
| 16 | 11732292 | 1 | 1 | rs11642880 | T | C | 0.06 | 0.21 | 0.19 | 0.43 | | | 5 tissues | | | PLZF,Pdx1 | | 2 hits | 3 hits | TXNDC11 | intronic |
| 16 | 11734425 | 0.87 | 1 | rs35955905 | T | A | 0.09 | 0.19 | 0.19 | 0.40 | | | 10 tissues | 4 tissues | 5 bound proteins | Mef2,TATA,Zfp105 | | | 2 hits | TXNDC11 | intronic |
| 16 | 11738353 | 0.86 | 1 | rs11642852 | A | G | 0.36 | 0.23 | 0.20 | 0.44 | | BRN, PANC | 11 tissues | BRN,SKIN | | 7 altered motifs | | | 1 hit | TXNDC11 | intronic |
| 16 | 11739796 | 0.94 | 1 | rs11644466 | A | G | 0.06 | 0.22 | 0.19 | 0.44 | | BLD, GI | 11 tissues | BLD | | | | | 2 hits | TXNDC11 | intronic |
| 16 | 11742090 | 0.85 | 0.93 | rs3743586 | C | A | 0.03 | 0.20 | 0.19 | 0.40 | | 24 tissues | HRT | 28 tissues | POL2 | Pax-5,ZBTB33,Zbtb12 | | | 2 hits | RP11-490O6.2 | intronic |

Query SNP: rs1559439 and variants with $r^2$ >= 0.8

| chr | pos (hg38) | LD ($r^2$) | LD (D') | variant | Ref | Alt | AFR freq | AMR freq | ASN freq | EUR freq | SiPhy cons | Promoter histone marks | Enhancer histone marks | DNAse | Proteins bound | Motifs changed | NHGRI/EBI GWAS hits | GRASP QTL hits | Selected eQTL hits | GENCODE genes | dbSNP func annot |
|---|---|---|---|---|---|---|---|---|---|---|---|---|---|---|---|---|---|---|---|---|---|
| 16 | 83277251 | 0.84 | 1 | rs12921363 | A | C | 0.37 | 0.73 | 0.80 | 0.72 | | | | BRN | | Pbx3 | | | | CDH13 | intronic |
| 16 | 83277318 | 1 | 1 | rs1559439 | G | C | 0.34 | 0.70 | 0.78 | 0.72 | | | | | | GR | | | | CDH13 | intronic |

Query SNP: rs4717540 and variants with $r^2$ >= 0.8

| chr | pos (hg38) | LD ($r^2$) | LD (D') | variant | Ref | Alt | AFR freq | AMR freq | ASN freq | EUR freq | SiPhy cons | Promoter histone marks | Enhancer histone marks | DNAse | Proteins bound | Motifs changed | NHGRI/EBI GWAS hits | GRASP QTL hits | Selected eQTL hits | GENCODE genes | dbSNP func annot |
|---|---|---|---|---|---|---|---|---|---|---|---|---|---|---|---|---|---|---|---|---|---|
| 7 | 70631915 | 0.83 | 0.92 | rs4718972 | G | A | 0.47 | 0.48 | 0.86 | 0.44 | | | 7 tissues | 6 tissues | NANOG | PTF1-beta | | | | AUTS2 | intronic |
| 7 | 70639990 | 0.9 | 1 | rs35749841 | GA | G | 0.40 | 0.46 | 0.85 | 0.43 | | | 8 tissues | ESDR | | 5 altered motifs | | | | AUTS2 | intronic |
| 7 | 70640311 | 0.97 | 1 | rs9638590 | G | A | 0.46 | 0.48 | 0.86 | 0.44 | | | 7 tissues | ESDR | | 9 altered motifs | | | | AUTS2 | intronic |
| 7 | 70641583 | 1 | 1 | rs4717540 | A | G | 0.53 | 0.49 | 0.86 | 0.44 | | | | | | Mrg1::Hoxa9,Nr2f2 | | | | AUTS2 | intronic |

Query SNP: rs3924222 and variants with $r^2$ >= 0.8

| chr | pos (hg38) | LD ($r^2$) | LD (D') | variant | Ref | Alt | AFR freq | AMR freq | ASN freq | EUR freq | SiPhy cons | Promoter histone marks | Enhancer histone marks | DNAse | Proteins bound | Motifs changed | NHGRI/EBI GWAS hits | GRASP QTL hits | Selected eQTL hits | GENCODE genes | dbSNP func annot |
|---|---|---|---|---|---|---|---|---|---|---|---|---|---|---|---|---|---|---|---|---|---|
| 14 | 65805929 | 0.82 | 0.94 | rs6573632 | A | G | 0.46 | 0.48 | 0.57 | 0.56 | | | | | | ELF1 | | 3 hits | | 62kb 3' of FUT8 | |
| 14 | 65805946 | 0.82 | 0.94 | rs6573633 | G | A | 0.45 | 0.48 | 0.57 | 0.56 | | | | | | Dmbx1 | | | | 62kb 3' of FUT8 | |
| 14 | 65807020 | 1 | 1 | rs3924222 | C | T | 0.42 | 0.50 | 0.58 | 0.59 | | | | | | 6 altered motifs | | 8 hits | | 63kb 3' of FUT8 | |

| | | | | | | | | | | | | | | | | | | | | |
|---|---|---|---|---|---|---|---|---|---|---|---|---|---|---|---|---|---|---|---|---|
| 14 | 65810559 | 0.92 | 0.98 | rs10149050 | C | T | 0.36 | 0.49 | 0.58 | 0.59 | | | | 5 tissues | | 13 altered motifs | | | | 66kb 3' of FUT8 |
| 14 | 65810567 | 0.92 | 0.98 | rs10138193 | T | G | 0.35 | 0.49 | 0.58 | 0.59 | | | | 5 tissues | | Foxo,MZF1::1-4,TATA | | | | 66kb 3' of FUT8 |
| 14 | 65810649 | 0.92 | 0.98 | rs10149325 | G | A | 0.36 | 0.49 | 0.58 | 0.59 | | | | KID,VAS | | | | 9 hits | | 67kb 3' of FUT8 |

Query SNP: rs10887087 and variants with $r^2$ >= 0.8

| chr | pos (hg38) | LD ($r^2$) | LD (D') | variant | Ref | Alt | AFR freq | AMR freq | ASN freq | EUR freq | SiPhy cons | Promoter histone marks | Enhancer histone marks | DNAse | Proteins bound | Motifs changed | NHGRI/EBI GWAS hits | GRASP QTL hits | Selected eQTL hits | GENCODE genes | dbSNP func annot |
|---|---|---|---|---|---|---|---|---|---|---|---|---|---|---|---|---|---|---|---|---|---|
| 10 | 122119164 | 0.8 | 0.97 | rs2421005 | T | C | 0.22 | 0.67 | 0.58 | 0.67 | | | 6 tissues | ESDR | | | | | 1 hit | TACC2 | intronic |
| 10 | 122120465 | 0.91 | 1 | rs2421004 | G | A | 0.22 | 0.72 | 0.80 | 0.80 | | | 9 tissues | HRT | | Hic1,Pax-5 | | | | TACC2 | intronic |
| 10 | 122120539 | 0.91 | 1 | rs2421002 | T | C | 0.26 | 0.72 | 0.80 | 0.79 | | | 8 tissues | ADRL,HRT,LNG | | BDP1,Maf | | | | TACC2 | intronic |
| 10 | 122120716 | 0.92 | 1 | rs4457688 | A | C | 0.25 | 0.72 | 0.80 | 0.80 | | | 7 tissues | | | 6 altered motifs | | | | TACC2 | intronic |
| 10 | 122120921 | 0.81 | 0.91 | rs4639871 | T | C | 0.26 | 0.70 | 0.80 | 0.80 | | | 6 tissues | MUS | | 5 altered motifs | | | | TACC2 | intronic |
| 10 | 122121308 | 0.91 | 1 | rs4751873 | A | G | 0.23 | 0.72 | 0.80 | 0.80 | | | 7 tissues | 10 tissues | GR,STAT3 | AhR::Arnt,Arnt | | | | TACC2 | intronic |
| 10 | 122121586 | 0.91 | 1 | rs4752649 | C | T | 0.23 | 0.72 | 0.80 | 0.80 | | | 7 tissues | | | BATF,Smad3 | | | | TACC2 | intronic |
| 10 | 122121663 | 0.91 | 1 | rs4751874 | G | A | 0.23 | 0.72 | 0.80 | 0.80 | | | 7 tissues | IPSC | | ATF6,HNF4,RXRA | | | | TACC2 | intronic |
| 10 | 122121788 | 0.92 | 1 | rs6585787 | G | A | 0.23 | 0.72 | 0.80 | 0.80 | | | 7 tissues | | | | | | | TACC2 | intronic |
| 10 | 122121921 | 0.91 | 1 | rs6585788 | A | G | 0.23 | 0.72 | 0.80 | 0.80 | | | 5 tissues | | | CTCF,p53 | | | | TACC2 | intronic |
| 10 | 122121945 | 0.91 | 1 | rs6585789 | C | T | 0.23 | 0.72 | 0.80 | 0.80 | | | 5 tissues | HRT | | 10 altered motifs | | | | TACC2 | intronic |
| 10 | 122122087 | 0.91 | 1 | rs6585790 | G | A | 0.23 | 0.72 | 0.80 | 0.80 | | | ESDR, IPSC, HRT | | | | | | | TACC2 | intronic |
| 10 | 122122329 | 0.82 | 0.91 | rs7073365 | G | T | 0.23 | 0.70 | 0.79 | 0.78 | | | 4 tissues | | | BCL,CTCF | | | | TACC2 | intronic |
| 10 | 122122692 | 0.91 | 1 | rs4751875 | T | C | 0.21 | 0.72 | 0.80 | 0.80 | | | 6 tissues | 12 tissues | | | | | | TACC2 | intronic |
| 10 | 122122765 | 0.91 | 1 | rs4752652 | T | C | 0.21 | 0.72 | 0.80 | 0.80 | | | 6 tissues | 10 tissues | | AhR::Arnt,Arnt,Pax-5 | | | | TACC2 | intronic |
| 10 | 122122935 | 0.82 | 0.91 | rs11200414 | A | G | 0.21 | 0.70 | 0.80 | 0.80 | | | 6 tissues | HRT | | Pou1f1 | | | | TACC2 | intronic |
| 10 | 122123015 | 0.91 | 1 | rs11200415 | G | A | 0.21 | 0.72 | 0.80 | 0.80 | | | 6 tissues | HRT | | 4 altered motifs | | | | TACC2 | intronic |
| 10 | 122123019 | 0.88 | 1 | rs11200416 | A | G | 0.21 | 0.73 | 0.80 | 0.80 | | | 6 tissues | HRT | | Foxd3,HMG-IY,Homez | | | | TACC2 | intronic |
| 10 | 122123294 | 0.92 | 1 | rs11200417 | C | T | 0.20 | 0.72 | 0.80 | 0.80 | | | 4 tissues | LNG | | Sin3Ak-20 | | | | TACC2 | intronic |
| 10 | 122123460 | 0.92 | 1 | rs10887083 | T | C | 0.20 | 0.72 | 0.80 | 0.80 | | | | | | 4 altered motifs | | | | TACC2 | intronic |
| 10 | 122123638 | 0.91 | 1 | rs10788246 | C | T | 0.21 | 0.72 | 0.80 | 0.80 | | | | HRT | | Maf,Pou2f2,Pou3f3 | | | | TACC2 | intronic |
| 10 | 122123963 | 0.91 | 1 | rs10887085 | G | A | 0.21 | 0.72 | 0.80 | 0.80 | | | | | | Myc,ZBTB7A,Zfp161 | | | | TACC2 | intronic |
| 10 | 122124560 | 0.99 | 1 | rs4405248 | A | G | 0.14 | 0.71 | 0.66 | 0.75 | | | 5 tissues | ESDR,LIV | GR | 7 altered motifs | | | | TACC2 | intronic |
| 10 | 122124790 | 0.99 | 1 | rs10788247 | C | T | 0.14 | 0.71 | 0.66 | 0.75 | | | | IPSC,HRT,KID | | | | | | TACC2 | intronic |
| 10 | 122125497 | 1 | 1 | rs10887086 | T | G | 0.13 | 0.70 | 0.69 | 0.74 | | | | | | RXRA | | | | TACC2 | intronic |
| 10 | 122125516 | 1 | 1 | rs10887087 | A | G | 0.13 | 0.70 | 0.69 | 0.75 | | | | | | BCL,Ets,Sin3Ak-20 | | | | TACC2 | intronic |
| 10 | 122125684 | 1 | 1 | rs10887088 | G | A | 0.13 | 0.70 | 0.69 | 0.74 | | | | | | Mrg1::Hoxa9,YY1 | | | | TACC2 | intronic |
| 10 | 122126097 | 0.8 | 1 | rs202127041 | TC | T | 0.13 | 0.65 | 0.68 | 0.71 | | | 5 tissues | | | 7 altered motifs | | | | TACC2 | intronic |
| 10 | 122126263 | 1 | 1 | rs4373860 | G | A | 0.13 | 0.70 | 0.69 | 0.75 | | | 5 tissues | HRT | | 4 altered motifs | | | | TACC2 | intronic |
| 10 | 122126274 | 1 | 1 | rs2421001 | T | C | 0.13 | 0.70 | 0.69 | 0.75 | | | 5 tissues | HRT | | AIRE,CDP,Foxm1 | | | | TACC2 | intronic |
| 10 | 122126374 | 1 | 1 | rs2421000 | A | G | 0.13 | 0.70 | 0.69 | 0.75 | | | 8 tissues | HRT | | Ets | | | | TACC2 | intronic |
| 10 | 122126381 | 1 | 1 | rs2901302 | C | T | 0.13 | 0.70 | 0.69 | 0.75 | | | 8 tissues | HRT | | | | | | TACC2 | intronic |
| 10 | 122126415 | 1 | 1 | rs11200419 | G | A | 0.13 | 0.70 | 0.69 | 0.75 | | | 8 tissues | HRT,LNG | | HNF4 | | | | TACC2 | intronic |
| 10 | 122126443 | 0.97 | 1 | rs11200420 | G | A | 0.13 | 0.70 | 0.69 | 0.74 | | | 8 tissues | SKIN,HRT,LNG | | Gm397,Irx | | | | TACC2 | intronic |
| 10 | 122127074 | 1 | 1 | rs10788248 | A | T | 0.13 | 0.70 | 0.69 | 0.74 | | | | 9 tissues | 4 bound proteins | RXRA | | | | TACC2 | intronic |
| 10 | 122127268 | 0.96 | 1 | rs10749447 | T | C | 0.13 | 0.71 | 0.69 | 0.76 | | | | 21 tissues | POL2 | | | | | TACC2 | intronic |
| 10 | 122127286 | 1 | 1 | rs10749448 | T | G | 0.13 | 0.70 | 0.69 | 0.75 | | | | 22 tissues | GTF2F1,POL2 | 8 altered motifs | | | | TACC2 | intronic |
| 10 | 122127408 | 1 | 1 | rs10788249 | T | C | 0.13 | 0.70 | 0.69 | 0.75 | | | | 33 tissues | 10 bound proteins | 4 altered motifs | | | | TACC2 | intronic |
| 10 | 122127744 | 1 | 1 | rs10887089 | C | G | 0.13 | 0.70 | 0.69 | 0.75 | | | | 6 tissues | | ZBRK1 | | | | TACC2 | intronic |
| 10 | 122133871 | 0.92 | 1 | rs4237537 | G | A | 0.13 | 0.69 | 0.65 | 0.73 | | | 5 tissues | | | Egr-1,YY1 | | 1 hit | | TACC2 | intronic |

Query SNP: rs2238090 and variants with $r^2$ >= 0.8

| chr | pos (hg38) | LD ($r^2$) | LD (D') | variant | Ref | Alt | AFR freq | AMR freq | ASN freq | EUR freq | SiPhy cons | Promoter histone marks | Enhancer histone marks | DNAse | Proteins bound | Motifs changed | NHGRI/EBI GWAS hits | GRASP QTL hits | Selected eQTL hits | GENCODE genes | dbSNP func annot |
|---|---|---|---|---|---|---|---|---|---|---|---|---|---|---|---|---|---|---|---|---|---|
| 12 | 2574166 | 1 | 1 | rs2238090 | A | G | 0.22 | 0.64 | 0.74 | 0.69 | | | | | | | | | | CACNA1C | intronic |
| 12 | 2581215 | 0.8 | 0.93 | rs7310458 | G | A | 0.27 | 0.65 | 0.72 | 0.72 | | | 4 tissues | | | 4 altered motifs | | | | CACNA1C | intronic |
| 12 | 2581536 | 0.81 | 0.93 | rs11832738 | G | A | 0.25 | 0.65 | 0.71 | 0.72 | | | LNG, BRN, GI | ESC,IPSC,IPSC | | NRSF,Nanog,PLAG1 | | | | CACNA1C | intronic |
| 12 | 2582094 | 0.82 | 0.92 | rs2071641 | C | G | 0.22 | 0.64 | 0.71 | 0.72 | | | | | | CEBPB,CEBPD,Zbtb3 | | | | CACNA1C | intronic |

Query SNP: rs11530859 and variants with $r^2$ >= 0.8

| chr | pos (hg38) | LD ($r^2$) | LD (D') | variant | Ref | Alt | AFR freq | AMR freq | ASN freq | EUR freq | SiPhy cons | Promoter histone marks | Enhancer histone marks | DNAse | Proteins bound | Motifs changed | NHGRI/EBI GWAS hits | GRASP QTL hits | Selected eQTL hits | GENCODE genes | dbSNP func annot |
|---|---|---|---|---|---|---|---|---|---|---|---|---|---|---|---|---|---|---|---|---|---|
| 11 | 83217776 | 1 | 1 | rs11530859 | C | G | 0.08 | 0.05 | 0.04 | 0.06 | | | | ESDR,GI | | Sox | | | 3 hits | ANKRD42 | intronic |

Query SNP: rs12709295 and variants with $r^2$ >= 0.8

| chr | pos (hg38) | LD ($r^2$) | LD (D') | variant | Ref | Alt | AFR freq | AMR freq | ASN freq | EUR freq | SiPhy cons | Promoter histone marks | Enhancer histone marks | DNAse | Proteins bound | Motifs changed | NHGRI/EBI GWAS hits | GRASP QTL hits | Selected eQTL hits | GENCODE genes | dbSNP func annot |
|---|---|---|---|---|---|---|---|---|---|---|---|---|---|---|---|---|---|---|---|---|---|
| 17 | 12740883 | 0.84 | -0.92 | rs62060373 | T | A | 0.14 | 0.21 | 0.28 | 0.13 | | | | | | Gm397,Mtf1 | | | | MYOCD | intronic |
| 17 | 12744173 | 1 | 1 | rs12709295 | T | A,C | 0.67 | 0.79 | 0.72 | 0.87 | | HRT | 9 tissues | SKIN,HRT | | | | | | MYOCD | intronic |

Query SNP: rs12438724 and variants with $r^2$ >= 0.8

| chr | pos (hg38) | LD ($r^2$) | LD (D') | variant | Ref | Alt | AFR freq | AMR freq | ASN freq | EUR freq | SiPhy cons | Promoter histone marks | Enhancer histone marks | DNAse | Proteins bound | Motifs changed | NHGRI/EBI GWAS hits | GRASP QTL hits | Selected eQTL hits | GENCODE genes | dbSNP func annot |
|---|---|---|---|---|---|---|---|---|---|---|---|---|---|---|---|---|---|---|---|---|---|
| 15 | 85279318 | 0.91 | 1 | rs1995952 | C | T | 0.46 | 0.49 | 0.62 | 0.37 | | | | | | CEBPD,Foxp3,ZEB1 | | | 3 hits | 35kb 3' of RP11-561C5.6 | |
| 15 | 85279370 | 0.86 | 0.97 | rs1995953 | G | A | 0.47 | 0.48 | 0.62 | 0.37 | | | | | | AP-2,Zfp410 | | | 4 hits | 35kb 3' of RP11-561C5.6 | |
| 15 | 85282062 | 1 | 1 | rs12438724 | C | T | 0.40 | 0.46 | 0.62 | 0.36 | | | LNG, GI | | | 7 altered motifs | | | 4 hits | 33kb 3' of RP11-561C5.6 | |
| 15 | 85283655 | 1 | 1 | rs12437932 | G | A | 0.39 | 0.46 | 0.62 | 0.36 | | | 6 tissues | THYM | | GR | | | 4 hits | 31kb 3' of RP11-561C5.6 | |
| 15 | 85283918 | 0.99 | 1 | rs6496935 | C | A | 0.39 | 0.46 | 0.62 | 0.36 | | | 5 tissues | | CTCF,BATF | 5 altered motifs | | | 3 hits | 31kb 3' of RP11-561C5.6 | |
| 15 | 85284412 | 0.97 | 1 | rs2002641 | C | T | 0.39 | 0.45 | 0.62 | 0.34 | | | BLD | | | IRC900814 | | | 2 hits | 30kb 3' of RP11-561C5.6 | |

| | | | | | | | | | | | | | | | | | | | | |
|---|---|---|---|---|---|---|---|---|---|---|---|---|---|---|---|---|---|---|---|---|
| 15 | 85286436 | 0.97 | 1 | rs6496940 | C | T | 0.39 | 0.45 | 0.62 | 0.34 | | | | | | Mrg,Tgif1 | | | 2 hits | 28kb 3' of RP11-561C5.6 |
| 15 | 85291233 | 0.9 | 0.97 | rs28456180 | A | C | 0.46 | 0.45 | 0.57 | 0.36 | | | | | | 7 altered motifs | | | 4 hits | 23kb 3' of RP11-561C5.6 |
| 15 | 85291252 | 0.9 | 1 | rs28671066 | A | G | 0.45 | 0.44 | 0.57 | 0.36 | | | | | | SIX5,TEF | | | 3 hits | 23kb 3' of RP11-561C5.6 |
| 15 | 85293583 | 0.94 | 1 | rs11074075 | G | T | 0.44 | 0.44 | 0.55 | 0.36 | | | | CRVX | GATA2 | 5 altered motifs | | | 2 hits | 21kb 3' of RP11-561C5.6 |
| 15 | 85294166 | 0.94 | 1 | rs7169412 | A | G | 0.44 | 0.44 | 0.55 | 0.36 | | | | BRST | | 4 altered motifs | | | 2 hits | 21kb 3' of RP11-561C5.6 |
| 15 | 85295159 | 0.9 | 0.97 | rs8035975 | A | G | 0.53 | 0.45 | 0.55 | 0.36 | | | | | | CEBPA,Hic1 | | | 2 hits | 20kb 3' of RP11-561C5.6 |
| 15 | 85295671 | 0.91 | 0.99 | rs10520583 | C | A | 0.44 | 0.44 | 0.56 | 0.36 | | | | BLD | | 5 altered motifs | | | 3 hits | 19kb 3' of RP11-561C5.6 |
| 15 | 85296936 | 0.9 | 0.99 | rs13379676 | T | C | 0.45 | 0.44 | 0.56 | 0.36 | | 19 tissues | 15 tissues | 38 tissues | CFOS,CTCF,ZNF263 | ATF6,XBP-1 | | | 2 hits | 18kb 3' of RP11-561C5.6 |
| 15 | 85297098 | 0.9 | 0.99 | rs16941991 | T | C | 0.45 | 0.44 | 0.56 | 0.36 | | 19 tissues | 13 tissues | 15 tissues | CFOS | CTCF,Cart1,IRC900814 | | | 2 hits | 18kb 3' of RP11-561C5.6 |
| 15 | 85297681 | 0.87 | 0.95 | rs1828481 | A | C | 0.52 | 0.45 | 0.56 | 0.36 | | LIV | 5 tissues | 4 tissues | | 6 altered motifs | | | 2 hits | 17kb 3' of RP11-561C5.6 |
| 15 | 85298572 | 0.9 | 0.99 | rs8037546 | A | G | 0.45 | 0.44 | 0.56 | 0.36 | | | LIV | | | 4 altered motifs | | | 2 hits | 16kb 3' of RP11-561C5.6 |
| 15 | 85298850 | 0.9 | 0.99 | rs2048439 | T | C | 0.44 | 0.44 | 0.56 | 0.36 | | | LIV | | | | | | 3 hits | 16kb 3' of RP11-561C5.6 |
| 15 | 85299561 | 0.9 | 0.99 | rs2342194 | T | G | 0.45 | 0.44 | 0.56 | 0.36 | | | ESDR | | | HDAC2,Ik-2,TATA | | | 2 hits | 15kb 3' of RP11-561C5.6 |
| 15 | 85300286 | 0.88 | 0.98 | rs7172789 | T | A,C,G | 0.45 | 0.44 | 0.56 | 0.36 | | | | | | | | | 2 hits | 14kb 3' of RP11-561C5.6 |
| 15 | 85301356 | 0.88 | 0.98 | rs28425568 | A | C | 0.45 | 0.44 | 0.55 | 0.36 | | | SKIN, BLD | | | CHOP::CEBPalpha,LRH1 | | | 2 hits | 13kb 3' of RP11-561C5.6 |
| 15 | 85302617 | 0.81 | 0.9 | rs7165242 | G | T | 0.61 | 0.46 | 0.56 | 0.37 | | | BLD | | | YY1 | | | 3 hits | 12kb 3' of RP11-561C5.6 |
| 15 | 85303185 | 0.81 | 0.9 | rs28595486 | A | G | 0.53 | 0.46 | 0.55 | 0.37 | | | BLD | | | KAP1 | | | 3 hits | 11kb 3' of RP11-561C5.6 |
| 15 | 85303538 | 0.81 | 0.9 | rs11857046 | G | A | 0.53 | 0.46 | 0.55 | 0.37 | | | | | | | | | 3 hits | 11kb 3' of RP11-561C5.6 |
| 15 | 85303952 | 0.81 | 0.9 | rs8032871 | G | A | 0.53 | 0.46 | 0.55 | 0.37 | | | | | | Brachyury,TBX5 | | | 3 hits | 11kb 3' of RP11-561C5.6 |
| 15 | 85304102 | 0.88 | 0.98 | rs200455667 | AAAG | A | 0.43 | 0.44 | 0.55 | 0.36 | | | | | | 8 altered motifs | | | 3 hits | 11kb 3' of RP11-561C5.6 |
| 15 | 85304103 | 0.88 | 0.98 | rs201187121 | AAG | A | 0.44 | 0.44 | 0.55 | 0.36 | | | | | | 7 altered motifs | | | 2 hits | 11kb 3' of RP11-561C5.6 |
| 15 | 85308832 | 0.86 | 0.97 | rs7176564 | C | T | 0.40 | 0.44 | 0.55 | 0.36 | | | BLD | | | CTCF,Zbtb3 | | | 4 hits | 5.8kb 3' of RP11-561C5.6 |
| 15 | 85317886 | 0.8 | 0.93 | rs28673870 | G | A | 0.39 | 0.44 | 0.55 | 0.36 | | | 11 tissues | KID,CRVX | | 5 altered motifs | | | 4 hits | 2.4kb 5' of RP11-561C5.6 |
| 15 | 85325822 | 0.8 | 0.93 | rs8032899 | C | T | 0.39 | 0.44 | 0.55 | 0.36 | | | BLD, SKIN | BLD,BLD,MUS | | 4 altered motifs | | | 4 hits | 10kb 5' of RP11-561C5.6 |

Query SNP: rs4516649 and variants with $r^2 >= 0.8$

| chr | pos (hg38) | LD ($r^2$) | LD (D') | variant | Ref | Alt | AFR freq | AMR freq | ASN freq | EUR freq | SiPhy cons | Promoter histone marks | Enhancer histone marks | DNAse | Proteins bound | Motifs changed | NHGRI/EBI GWAS hits | GRASP QTL hits | Selected eQTL hits | GENCODE genes | dbSNP func annot |
|---|---|---|---|---|---|---|---|---|---|---|---|---|---|---|---|---|---|---|---|---|---|
| 3 | 131851106 | 0.85 | 0.94 | rs143774255 | TATG | T | 0.61 | 0.73 | 0.55 | 0.63 | | | | | | 4 altered motifs | | | | CPNE4 | intronic |
| 3 | 131860608 | 0.91 | 0.99 | rs1355779 | G | A | 0.70 | 0.74 | 0.56 | 0.63 | | | | SKIN | | HNF4,RAR,RXRA | | | 3 hits | CPNE4 | intronic |
| 3 | 131862783 | 0.87 | -1 | rs9682807 | G | A | 0.19 | 0.25 | 0.45 | 0.37 | | | | | | 5 altered motifs | | | 2 hits | CPNE4 | intronic |
| 3 | 131863428 | 0.92 | 1 | rs144915838 | A | G | 0.47 | 0.71 | 0.55 | 0.63 | | | | | | Evi-1,Irf,Osf2 | | | 2 hits | CPNE4 | intronic |
| 3 | 131866348 | 0.92 | 1 | rs7621543 | C | T | 0.71 | 0.74 | 0.55 | 0.63 | | | IPSC, BRN | | | 4 altered motifs | | | 3 hits | CPNE4 | intronic |
| 3 | 131870649 | 0.95 | 0.97 | rs4854844 | A | T | 0.51 | 0.72 | 0.55 | 0.64 | | | ESDR | | | Pou1f1,Znf143 | | | 1 hit | CPNE4 | intronic |
| 3 | 131870732 | 0.95 | 0.97 | rs6767374 | A | C | 0.51 | 0.72 | 0.55 | 0.64 | | | ESDR | | | Ets,STAT | | 1 hit | 1 hit | CPNE4 | intronic |
| 3 | 131872503 | 1 | 1 | rs9852025 | A | C | 0.60 | 0.72 | 0.55 | 0.64 | | | ESDR | | | TBX5 | | | 2 hits | CPNE4 | intronic |
| 3 | 131872716 | 1 | 1 | rs9834748 | T | C | 0.59 | 0.72 | 0.55 | 0.64 | | | ESDR | | | GATA,p300 | | | 2 hits | CPNE4 | intronic |
| 3 | 131872832 | 0.97 | 1 | rs9834926 | T | G | 0.61 | 0.73 | 0.55 | 0.64 | | | ESDR | | | Mrg1::Hoxa9 | | | 3 hits | CPNE4 | intronic |
| 3 | 131874044 | 0.97 | 1 | rs3896910 | G | A | 0.62 | 0.73 | 0.55 | 0.64 | | | | | | 4 altered motifs | | | 3 hits | CPNE4 | intronic |
| 3 | 131874763 | 0.96 | 0.99 | rs6787523 | G | T | 0.67 | 0.72 | 0.55 | 0.64 | | | | | | 14 altered motifs | | | 3 hits | CPNE4 | intronic |
| 3 | 131875799 | 0.85 | 1 | rs34294494 | T | C | 0.58 | 0.69 | 0.50 | 0.60 | | | | | | CTCF,FAC1,Gm397 | | | 2 hits | CPNE4 | intronic |
| 3 | 131876882 | 1 | 1 | rs4444755 | C | A | 0.60 | 0.72 | 0.56 | 0.64 | | | | | | GR,Pax-6,Sin3Ak-20 | | | 3 hits | CPNE4 | intronic |
| 3 | 131877200 | 0.86 | 1 | rs4605590 | T | C | 0.59 | 0.69 | 0.53 | 0.62 | | | | | | 5 altered motifs | | | 3 hits | CPNE4 | intronic |
| 3 | 131877213 | 1 | 1 | rs4566581 | C | T | 0.59 | 0.72 | 0.55 | 0.64 | | | | | | 4 altered motifs | | | 2 hits | CPNE4 | intronic |
| 3 | 131877247 | 1 | 1 | rs4516649 | A | G | 0.60 | 0.72 | 0.56 | 0.64 | | | | | | DMRT1 | | 1 hit | 3 hits | CPNE4 | intronic |
| 3 | 131877268 | 1 | 1 | rs4339144 | C | T | 0.60 | 0.72 | 0.56 | 0.64 | | | | | | 5 altered motifs | | | 3 hits | CPNE4 | intronic |
| 3 | 131877465 | 1 | 1 | rs12629564 | C | G | 0.60 | 0.72 | 0.56 | 0.64 | | | | | | | | 1 hit | 3 hits | CPNE4 | intronic |
| 3 | 131877504 | 0.99 | 1 | rs938240 | G | A | 0.60 | 0.73 | 0.56 | 0.64 | | | | | | Foxa | | 1 hit | 2 hits | CPNE4 | intronic |
| 3 | 131877676 | 0.97 | 1 | rs9850694 | A | G | 0.61 | 0.73 | 0.56 | 0.64 | | | | | | | | 1 hit | 3 hits | CPNE4 | intronic |
| 3 | 131877805 | 1 | 1 | rs6797925 | G | C | 0.60 | 0.72 | 0.56 | 0.64 | | | | | | | | | 3 hits | CPNE4 | intronic |
| 3 | 131878017 | 1 | 1 | rs6797963 | C | T | 0.60 | 0.72 | 0.56 | 0.64 | | | | | | | | | 3 hits | CPNE4 | intronic |
| 3 | 131878102 | 1 | 1 | rs6798225 | G | A | 0.60 | 0.72 | 0.56 | 0.64 | | | | | | | | | 3 hits | CPNE4 | intronic |
| 3 | 131878511 | 0.99 | 1 | rs6798492 | C | T | 0.60 | 0.73 | 0.56 | 0.64 | | | | | | GR,NRSF | | | 2 hits | CPNE4 | intronic |
| 3 | 131878775 | 1 | 1 | rs9289401 | G | T | 0.60 | 0.72 | 0.56 | 0.64 | | | | | | GATA | | | 3 hits | CPNE4 | intronic |
| 3 | 131878882 | 1 | 1 | rs9289404 | C | T | 0.60 | 0.72 | 0.56 | 0.64 | | | | | | 4 altered motifs | | | 3 hits | CPNE4 | intronic |
| 3 | 131878974 | 1 | 1 | rs9289405 | A | G | 0.60 | 0.72 | 0.56 | 0.64 | | | | | | 4 altered motifs | | | 3 hits | CPNE4 | intronic |
| 3 | 131879066 | 1 | 1 | rs9289406 | T | C | 0.60 | 0.72 | 0.56 | 0.64 | | | | | | CEBPB | | | 3 hits | CPNE4 | intronic |
| 3 | 131879362 | 1 | 1 | rs9860875 | A | G | 0.60 | 0.72 | 0.56 | 0.64 | | | | | | GATA,STAT | | | 3 hits | CPNE4 | intronic |
| 3 | 131879526 | 1 | 1 | rs9843570 | T | G | 0.60 | 0.72 | 0.56 | 0.64 | | | | | | 5 altered motifs | | | 3 hits | CPNE4 | intronic |
| 3 | 131879597 | 1 | 1 | rs9843606 | T | G | 0.60 | 0.72 | 0.56 | 0.64 | | | BRN | | | DMRT1,Pax-4,Znf143 | | | 3 hits | CPNE4 | intronic |
| 3 | 131879794 | 1 | 1 | rs1461148 | A | G | 0.60 | 0.72 | 0.56 | 0.64 | | | | | | | | | 3 hits | CPNE4 | intronic |
| 3 | 131880120 | 0.8 | -0.97 | rs34508095 | A | G | 0.17 | 0.25 | 0.44 | 0.36 | | | | | | 8 altered motifs | | | 2 hits | CPNE4 | intronic |
| 3 | 131880332 | 0.82 | 0.96 | rs9866113 | A | G | 0.59 | 0.70 | 0.54 | 0.62 | | | | | | | | | 2 hits | CPNE4 | intronic |
| 3 | 131880384 | 0.82 | 0.96 | rs9828857 | G | A | 0.44 | 0.70 | 0.53 | 0.60 | | | | | | Pitx2,STAT | | | 2 hits | CPNE4 | intronic |
| 3 | 131880665 | 0.96 | 0.99 | rs4854847 | A | C | 0.60 | 0.72 | 0.55 | 0.64 | | | | | | DMRT2,Pax-4,Pou2f2 | | | 3 hits | CPNE4 | intronic |
| 3 | 131880749 | 0.89 | 0.97 | rs4854848 | G | A | 0.70 | 0.73 | 0.56 | 0.64 | | | | | | 7 altered motifs | | | 3 hits | CPNE4 | intronic |
| 3 | 131880808 | 0.95 | 0.97 | rs4854849 | A | G | 0.60 | 0.72 | 0.56 | 0.64 | | | BRN | | | 4 altered motifs | | | 3 hits | CPNE4 | intronic |
| 3 | 131880850 | 0.95 | 0.97 | rs4854852 | C | T | 0.60 | 0.72 | 0.56 | 0.64 | | | BRN | | | p300 | | | 3 hits | CPNE4 | intronic |
| 3 | 131881611 | 0.92 | 0.97 | rs1355781 | T | C | 0.62 | 0.73 | 0.56 | 0.64 | | | | | | Brachyury,HNF1 | | 1 hit | 3 hits | CPNE4 | intronic |
| 3 | 131881739 | 0.95 | 0.97 | rs1355780 | G | C | 0.60 | 0.72 | 0.56 | 0.64 | | | | | | Homez,SRF | | | 3 hits | CPNE4 | intronic |
| 3 | 131882963 | 0.85 | 0.93 | rs2369224 | T | C | 0.62 | 0.73 | 0.56 | 0.63 | | | SKIN | | | Smad3 | | | 2 hits | CPNE4 | intronic |
| 3 | 131883410 | 0.89 | 0.97 | rs2369228 | G | A | 0.64 | 0.73 | 0.56 | 0.64 | | | 4 tissues | | | GATA,RXRA,TAL1 | | | 3 hits | CPNE4 | intronic |
| 3 | 131884267 | 0.88 | 0.97 | rs1381075 | C | T | 0.64 | 0.74 | 0.56 | 0.65 | | | MUS, SKIN | SKIN,SKIN | | Gfi1,Gfi1b,Myc | | | 2 hits | CPNE4 | intronic |
| 3 | 131884378 | 0.92 | 0.97 | rs6439322 | G | A | 0.63 | 0.73 | 0.56 | 0.64 | | | SKIN | | | 7 altered motifs | | | 3 hits | CPNE4 | intronic |
| 3 | 131885326 | 0.87 | 0.97 | rs2369229 | G | A | 0.71 | 0.74 | 0.56 | 0.63 | | | | | | Pax-4 | | | 2 hits | CPNE4 | intronic |
| 3 | 131885688 | 0.92 | 0.97 | rs6777513 | T | C | 0.59 | 0.73 | 0.55 | 0.62 | | | | | | 19 altered motifs | | | 2 hits | CPNE4 | intronic |
| 3 | 131886063 | 0.87 | 0.97 | rs6789443 | A | G | 0.72 | 0.74 | 0.56 | 0.64 | | | | | | Rad21 | | | 3 hits | CPNE4 | intronic |
| 3 | 131886184 | 0.89 | 0.97 | rs1903437 | C | G | 0.64 | 0.73 | 0.56 | 0.64 | | | | | | ERalpha-a,Esr2,GR | | | 3 hits | CPNE4 | intronic |
| 3 | 131887579 | 0.89 | 0.97 | rs7615970 | C | A | 0.64 | 0.73 | 0.56 | 0.64 | | | | | | Foxp1,HDAC2,p300 | | | 3 hits | CPNE4 | intronic |
| 3 | 131891118 | 0.91 | 0.96 | rs12695562 | C | T | 0.63 | 0.73 | 0.56 | 0.64 | | | | | | 8 altered motifs | | | 3 hits | CPNE4 | intronic |
| 3 | 131895903 | 0.89 | 0.97 | rs1461144 | C | A | 0.64 | 0.73 | 0.56 | 0.64 | | | | ESDR | | 5 altered motifs | | | 3 hits | CPNE4 | intronic |

| | | | | | | | | | | | | | | | | | | | | | |
|---|---|---|---|---|---|---|---|---|---|---|---|---|---|---|---|---|---|---|---|---|---|
| 3 | 131898383 | 0.87 | 0.97 | rs6788559 | C | G | 0.72 | 0.74 | 0.56 | 0.64 | | | FAT, SKIN | SKIN | | CACD,STAT | | | 3 hits | CPNE4 | intronic |
| 3 | 131898583 | 0.92 | 0.97 | rs4854629 | C | T | 0.63 | 0.73 | 0.56 | 0.64 | | | FAT, BRN, SKIN | SKIN | | p300 | | | 3 hits | CPNE4 | intronic |
| 3 | 131899476 | 0.89 | 0.97 | rs9809611 | A | G | 0.64 | 0.73 | 0.56 | 0.64 | | | 4 tissues | | | CCNT2,Nanog,RFX5 | | | 3 hits | CPNE4 | intronic |
| 3 | 131903478 | 0.87 | 0.94 | rs4854856 | A | C | 0.64 | 0.73 | 0.61 | 0.63 | | | 9 tissues | | | 18 altered motifs | | | 3 hits | CPNE4 | intronic |

Query SNP: rs272709 and variants with $r^2$ >= 0.8

| chr | pos (hg38) | LD ($r^2$) | LD (D') | variant | Ref | Alt | AFR freq | AMR freq | ASN freq | EUR freq | SiPhy cons | Promoter histone marks | Enhancer histone marks | DNAse | Proteins bound | Motifs changed | NHGRI/ EBI GWAS hits | GRASP QTL hits | Selected eQTL hits | GENCODE genes | dbSNP func annot |
|---|---|---|---|---|---|---|---|---|---|---|---|---|---|---|---|---|---|---|---|---|---|
| 7 | 24108271 | 0.9 | 0.98 | rs272653 | T | C | 0.55 | 0.50 | 0.51 | 0.56 | | | | | | Foxa,Foxp1,NF-I | | | | 24kb 5' of RN5S228 | |
| 7 | 24108358 | 0.89 | 0.97 | rs111620459 | 7-mer | A | 0.54 | 0.49 | 0.51 | 0.55 | | | | | | Pou1f1 | | | | 24kb 5' of RN5S228 | |
| 7 | 24109673 | 0.92 | 0.98 | rs2813838 | C | G | 0.54 | 0.49 | 0.51 | 0.56 | | | GI, LIV | | | | | | | 23kb 5' of RN5S228 | |
| 7 | 24110244 | 0.96 | 0.98 | rs272712 | G | T | 0.41 | 0.48 | 0.51 | 0.56 | | | GI, LIV | GI,GI,GI | | KAP1 | | | | 22kb 5' of RN5S228 | |
| 7 | 24110323 | 0.92 | 0.98 | rs272711 | G | A | 0.54 | 0.49 | 0.51 | 0.56 | | | GI, LIV | GI,GI,GI | | PU.1,TFIIA | | | | 22kb 5' of RN5S228 | |
| 7 | 24111129 | 0.96 | 0.99 | rs272710 | A | T | 0.54 | 0.49 | 0.65 | 0.55 | | | | | | AP-2 | | | | 22kb 5' of RN5S228 | |
| 7 | 24111478 | 1 | 1 | rs272709 | T | A | 0.41 | 0.48 | 0.65 | 0.55 | | | | | | 4 altered motifs | | | | 21kb 5' of RN5S228 | |

Query SNP: rs2020009 and variants with $r^2$ >= 0.8

| chr | pos (hg38) | LD ($r^2$) | LD (D') | variant | Ref | Alt | AFR freq | AMR freq | ASN freq | EUR freq | SiPhy cons | Promoter histone marks | Enhancer histone marks | DNAse | Proteins bound | Motifs changed | NHGRI/ EBI GWAS hits | GRASP QTL hits | Selected eQTL hits | GENCODE genes | dbSNP func annot |
|---|---|---|---|---|---|---|---|---|---|---|---|---|---|---|---|---|---|---|---|---|---|
| 4 | 151892080 | 0.94 | 0.99 | rs518213 | C | A | 0.12 | 0.35 | 0.10 | 0.48 | | | ADRL | | | 4 altered motifs | | | 2 hits | 816bp 5' of RP11-503L23.1 | |
| 4 | 151894661 | 0.92 | 0.98 | rs361131 | C | T | 0.10 | 0.35 | 0.10 | 0.47 | | | PLCNT | | | 10 altered motifs | | | 1 hit | 3.4kb 5' of RP11-503L23.1 | |
| 4 | 151902434 | 0.94 | 0.99 | rs523554 | G | A | 0.11 | 0.35 | 0.10 | 0.48 | | | | | | 4 altered motifs | | | 1 hit | 2.5kb 3' of RP11-73G16.3 | |
| 4 | 151903231 | 0.94 | 0.99 | rs495213 | C | T | 0.11 | 0.35 | 0.10 | 0.48 | | | | | | 5 altered motifs | | | 1 hit | 1.7kb 3' of RP11-73G16.3 | |
| 4 | 151904880 | 0.8 | 0.99 | rs499797 | A | C | 0.12 | 0.38 | 0.10 | 0.54 | | | BLD | | | CEBPB,DEC,LXR | | | 2 hits | 51bp 3' of RP11-73G16.3 | |
| 4 | 151905597 | 0.92 | 0.99 | rs511880 | T | A | 0.11 | 0.35 | 0.11 | 0.47 | | | ESC, BLD, THYM | BLD | | Dbx1,PLZF,Zfp105 | | | 1 hit | RP11-73G16.3 | |
| 4 | 151913244 | 0.83 | 1 | rs360918 | C | T | 0.16 | 0.38 | 0.08 | 0.54 | | THYM | THYM | | | CEBPB | | | 1 hit | RP11-73G16.3 | |
| 4 | 151916302 | 1 | 1 | rs2020009 | C | T | 0.12 | 0.34 | 0.08 | 0.48 | | | | | | Pou3f3 | | | 2 hits | RP11-73G16.3 | |
| 4 | 151925597 | 0.96 | 1 | rs360936 | C | A | 0.15 | 0.35 | 0.07 | 0.48 | | | | | | 23 altered motifs | | | 1 hit | RP11-73G16.2 | |
| 4 | 151927168 | 0.95 | 0.98 | rs360937 | C | T | 0.05 | 0.34 | 0.02 | 0.47 | | | | | | 4 altered motifs | | 1 hit | 1 hit | RP11-73G16.2 | |
| 4 | 151932638 | 0.93 | 0.96 | rs530315 | C | T | 0.14 | 0.34 | 0.06 | 0.48 | | | | | | HDAC2,TCF4 | | | 1 hit | RP11-73G16.2 | |

Query SNP: rs6916224 and variants with $r^2$ >= 0.8

| chr | pos (hg38) | LD ($r^2$) | LD (D') | variant | Ref | Alt | AFR freq | AMR freq | ASN freq | EUR freq | SiPhy cons | Promoter histone marks | Enhancer histone marks | DNAse | Proteins bound | Motifs changed | NHGRI/ EBI GWAS hits | GRASP QTL hits | Selected eQTL hits | GENCODE genes | dbSNP func annot |
|---|---|---|---|---|---|---|---|---|---|---|---|---|---|---|---|---|---|---|---|---|---|
| 6 | 85035286 | 1 | 1 | rs6916224 | A | G | 0.34 | 0.23 | 0.43 | 0.19 | | | | | | Nr2e3 | | 1 hit | | 271kb 5' of TBX18 | |
| 6 | 85042211 | 0.88 | 0.98 | rs9444287 | T | C | 0.34 | 0.25 | 0.43 | 0.20 | | | VAS, BLD | | | DMRT1,Irf | | | | 278kb 5' of TBX18 | |
| 6 | 85050917 | 0.88 | 0.98 | rs7760395 | T | C | 0.34 | 0.25 | 0.43 | 0.20 | | | FAT, STRM | | | | | | | 286kb 5' of TBX18 | |

Query SNP: rs4732439 and variants with $r^2$ >= 0.8

| chr | pos (hg38) | LD ($r^2$) | LD (D') | variant | Ref | Alt | AFR freq | AMR freq | ASN freq | EUR freq | SiPhy cons | Promoter histone marks | Enhancer histone marks | DNAse | Proteins bound | Motifs changed | NHGRI/ EBI GWAS hits | GRASP QTL hits | Selected eQTL hits | GENCODE genes | dbSNP func annot |
|---|---|---|---|---|---|---|---|---|---|---|---|---|---|---|---|---|---|---|---|---|---|
| 7 | 75840909 | 0.91 | 0.95 | rs6971988 | A | G | 0.12 | 0.36 | 0.70 | 0.24 | | | | | | Pax-5 | | | 3 hits | 1.7kb 5' of RHBDD2 | |
| 7 | 75841084 | 0.91 | 0.95 | rs6964835 | C | T | 0.12 | 0.36 | 0.70 | 0.24 | | | | | | Lmo2-complex,TCF12,ZEB1 | | | 3 hits | 1.5kb 5' of RHBDD2 | |
| 7 | 75843049 | 0.91 | 0.95 | rs10954667 | A | T | 0.12 | 0.36 | 0.70 | 0.24 | | | | 37 tissues | 24 bound proteins | 6 altered motifs | | | 6 hits | RHBDD2 | |
| 7 | 75843131 | 0.91 | 0.95 | rs12533520 | A | G | 0.12 | 0.36 | 0.70 | 0.24 | | 5 tissues | 19 tissues | 39 tissues | 30 bound proteins | Nkx3 | | | 3 hits | RHBDD2 | |
| 7 | 75845419 | 0.99 | 1 | rs6952762 | C | T | 0.12 | 0.35 | 0.69 | 0.24 | | | | | | HNF4,Maf,SREBP | | | 3 hits | RHBDD2 | |
| 7 | 75845979 | 1 | 1 | rs4732439 | G | A | 0.12 | 0.36 | 0.69 | 0.24 | | | | | | Rad21,SIX5,p300 | | | 6 hits | RHBDD2 | |
| 7 | 75847795 | 0.99 | 1 | rs6958613 | G | T | 0.12 | 0.35 | 0.70 | 0.21 | | | BLD | BLD,MUS,BLD | | | | | 3 hits | RHBDD2 | |
| 7 | 75848099 | 0.87 | 0.99 | rs2868173 | T | C | 0.16 | 0.38 | 0.79 | 0.30 | | | BLD | OVRY | | 11 altered motifs | | | 6 hits | RHBDD2 | |
| 7 | 75851084 | 0.96 | 1 | rs58556529 | G | A | 0.12 | 0.35 | 0.64 | 0.23 | | | BLD | | | E2A,Myf | | | 3 hits | RHBDD2 | |
| 7 | 75854641 | 0.88 | 0.97 | rs58160288 | C | T | 0.12 | 0.34 | 0.64 | 0.23 | | | FAT, BLD | | | 4 altered motifs | | | 3 hits | RHBDD2 | |
| 7 | 75858148 | 0.91 | 0.96 | rs60791363 | C | T | 0.12 | 0.35 | 0.65 | 0.23 | | | | | | HP1-site-factor,Zbtb3 | | | 2 hits | RHBDD2 | |
| 7 | 75859720 | 0.93 | 0.99 | rs12535057 | G | A | 0.10 | 0.35 | 0.63 | 0.23 | | | | | | SP1 | | 3 hits | 6 hits | RHBDD2 | |

Query SNP: rs9882701 and variants with $r^2$ >= 0.8

| chr | pos (hg38) | LD ($r^2$) | LD (D') | variant | Ref | Alt | AFR freq | AMR freq | ASN freq | EUR freq | SiPhy cons | Promoter histone marks | Enhancer histone marks | DNAse | Proteins bound | Motifs changed | NHGRI/ EBI GWAS hits | GRASP QTL hits | Selected eQTL hits | GENCODE genes | dbSNP func annot |
|---|---|---|---|---|---|---|---|---|---|---|---|---|---|---|---|---|---|---|---|---|---|
| 3 | 141698203 | 1 | 1 | rs9882701 | A | G | 0.15 | 0.15 | 0.00 | 0.15 | | | BLD | | | 4 altered motifs | | | | RP11-340E6.1 | intronic |

Query SNP: rs17655474 and variants with $r^2$ >= 0.8

| chr | pos (hg38) | LD ($r^2$) | LD (D') | variant | Ref | Alt | AFR freq | AMR freq | ASN freq | EUR freq | SiPhy cons | Promoter histone marks | Enhancer histone marks | DNAse | Proteins bound | Motifs changed | NHGRI/ EBI GWAS hits | GRASP QTL hits | Selected eQTL hits | GENCODE genes | dbSNP func annot |
|---|---|---|---|---|---|---|---|---|---|---|---|---|---|---|---|---|---|---|---|---|---|
| 17 | 78497571 | 0.91 | 1 | rs62073549 | C | T | 0.06 | 0.11 | 0.02 | 0.16 | | BRN | 12 tissues | ESC,LIV | | Irf,Pax-5,Sin3Ak-20 | | | | DNAH17 | intronic |
| 17 | 78499404 | 1 | 1 | rs17655474 | C | G | 0.05 | 0.10 | 0.02 | 0.16 | | | 6 tissues | ESDR | | HNF4 | | | | DNAH17 | intronic |
| 17 | 78500101 | 0.94 | 1 | rs2028733 | G | A | 0.12 | 0.10 | 0.02 | 0.16 | | | | | | NRSF | | | | DNAH17 | intronic |

Query SNP: rs1501142 and variants with $r^2$ >= 0.8

| chr | pos (hg38) | LD (r²) | LD (D') | variant | Ref | Alt | AFR freq | AMR freq | ASN freq | EUR freq | SiPhy cons | Promoter histone marks | Enhancer histone marks | DNAse | Proteins bound | Motifs changed | NHGRI/EBI GWAS hits | GRASP QTL hits | Selected eQTL hits | GENCODE genes | dbSNP func annot |
|---|---|---|---|---|---|---|---|---|---|---|---|---|---|---|---|---|---|---|---|---|---|
| 4 | 16493086 | 0.82 | 0.96 | rs6822723 | A | C | 0.31 | 0.26 | 0.08 | 0.22 | | | | 4 tissues | | CEBPB,EWSR1-FLI1,STAT | | | | RP11-446J8.1 | |
| 4 | 16493817 | 0.86 | 1 | rs10029427 | G | A | 0.22 | 0.25 | 0.08 | 0.22 | | | | | | AP-1 | | | | RP11-446J8.1 | |
| 4 | 16495233 | 1 | 1 | rs4333183 | T | A | 0.22 | 0.28 | 0.13 | 0.22 | | | | | | Dlx3 | | | | RP11-446J8.1 | |
| 4 | 16495505 | 1 | 1 | rs1501142 | A | G | 0.22 | 0.28 | 0.13 | 0.22 | | | | | | GCNF,HMG-IY,Ik-2 | | | | RP11-446J8.1 | |
| 4 | 16498210 | 1 | 1 | rs7698804 | A | G | 0.22 | 0.28 | 0.13 | 0.22 | | | | | | | | 2 hits | | RP11-446J8.1 | |
| 4 | 16502561 | 0.97 | 0.99 | rs872478 | G | C | 0.22 | 0.28 | 0.13 | 0.22 | | | BRN | | | 5 altered motifs | | | | RP11-446J8.1 | 3'-UTR |
| 4 | 16503896 | 0.91 | 0.99 | rs3792678 | T | C | 0.25 | 0.30 | 0.13 | 0.24 | | | BRN | BRN,BRN,MUS | | Pou1f1 | | | | RP11-446J8.1 | intronic |

Query SNP: rs299289 and variants with $r^2$ >= 0.8

| chr | pos (hg38) | LD (r²) | LD (D') | variant | Ref | Alt | AFR freq | AMR freq | ASN freq | EUR freq | SiPhy cons | Promoter histone marks | Enhancer histone marks | DNAse | Proteins bound | Motifs changed | NHGRI/EBI GWAS hits | GRASP QTL hits | Selected eQTL hits | GENCODE genes | dbSNP func annot |
|---|---|---|---|---|---|---|---|---|---|---|---|---|---|---|---|---|---|---|---|---|---|
| 5 | 163483703 | 0.86 | 0.99 | rs299310 | G | A | 0.52 | 0.27 | 0.11 | 0.30 | | | BLD | | | Osf2 | | | 4 hits | RP11-80G7.1 | intronic |
| 5 | 163495817 | 1 | 1 | rs299289 | C | A | 0.57 | 0.30 | 0.11 | 0.33 | | | ESC | | | | | | 6 hits | 1.8kb 5' of RP11-80G7.1 | |
| 5 | 163506803 | 0.8 | -0.94 | rs67375711 | AAAG | A | 0.32 | 0.73 | 0.88 | 0.69 | | 23 tissues | ESDR, ESC, HRT | BLD,CRVX,BRST | | 7 altered motifs | | | 2 hits | MAT2B | intronic |

Query SNP: rs9319501 and variants with $r^2$ >= 0.8

| chr | pos (hg38) | LD (r²) | LD (D') | variant | Ref | Alt | AFR freq | AMR freq | ASN freq | EUR freq | SiPhy cons | Promoter histone marks | Enhancer histone marks | DNAse | Proteins bound | Motifs changed | NHGRI/EBI GWAS hits | GRASP QTL hits | Selected eQTL hits | GENCODE genes | dbSNP func annot |
|---|---|---|---|---|---|---|---|---|---|---|---|---|---|---|---|---|---|---|---|---|---|
| 16 | 77006262 | 0.91 | 1 | rs11640444 | T | G | 0.35 | 0.36 | 0.41 | 0.40 | | | | | | | | | 1 hit | 78kb 3' of CTD-2336H13.2 | |
| 16 | 77009440 | 1 | 1 | rs9319501 | T | A | 0.54 | 0.38 | 0.48 | 0.41 | | | ESDR | | | | | | 1 hit | 81kb 3' of CTD-2336H13.2 | |
| 16 | 77010660 | 0.93 | 1 | rs12103143 | T | A | 0.59 | 0.40 | 0.49 | 0.41 | | | | IPSC | | 5 altered motifs | | | 1 hit | 82kb 3' of CTD-2336H13.2 | |
| 16 | 77010802 | 0.92 | 1 | rs8052754 | T | G | 0.66 | 0.40 | 0.49 | 0.41 | | | | | | 6 altered motifs | | | 1 hit | 83kb 3' of CTD-2336H13.2 | |
| 16 | 77013519 | 0.98 | 0.99 | rs7206200 | G | A | 0.64 | 0.38 | 0.49 | 0.41 | | | | BRN | | Alx4,HNF1 | | | 1 hit | 85kb 3' of CTD-2336H13.2 | |

Query SNP: rs7180563 and variants with $r^2$ >= 0.8

| chr | pos (hg38) | LD (r²) | LD (D') | variant | Ref | Alt | AFR freq | AMR freq | ASN freq | EUR freq | SiPhy cons | Promoter histone marks | Enhancer histone marks | DNAse | Proteins bound | Motifs changed | NHGRI/EBI GWAS hits | GRASP QTL hits | Selected eQTL hits | GENCODE genes | dbSNP func annot |
|---|---|---|---|---|---|---|---|---|---|---|---|---|---|---|---|---|---|---|---|---|---|
| 15 | 29574843 | 1 | 1 | rs7180563 | A | C | 0.92 | 0.88 | 0.99 | 0.84 | | | | | | | | 1 hit | | 4.1kb 5' of FAM189A1 | |
| 15 | 29574990 | 1 | 1 | rs7180782 | A | G | 0.92 | 0.88 | 1.00 | 0.84 | | | | | | | | 1 hit | | 4.3kb 5' of FAM189A1 | |
| 15 | 29582493 | 1 | 1 | rs4779665 | A | G | 0.92 | 0.88 | 1.00 | 0.84 | | | | | | 4 altered motifs | | | | 12kb 5' of FAM189A1 | |
| 15 | 29584396 | 1 | 1 | rs1073545 | T | C | 0.92 | 0.88 | 1.00 | 0.84 | | | | | | 4 altered motifs | | | | 14kb 5' of FAM189A1 | |
| 15 | 29587120 | 1 | 1 | rs10640883 | G | GCTGT | 0.92 | 0.88 | 1.00 | 0.83 | | | | | | Smad | | | | 16kb 5' of FAM189A1 | |
| 15 | 29589145 | 0.86 | -1 | rs62014337 | G | C | 0.26 | 0.14 | 0.00 | 0.16 | | | | | | Znf143 | | | | 18kb 5' of FAM189A1 | |

Query SNP: rs11255291 and variants with $r^2$ >= 0.8

| chr | pos (hg38) | LD (r²) | LD (D') | variant | Ref | Alt | AFR freq | AMR freq | ASN freq | EUR freq | SiPhy cons | Promoter histone marks | Enhancer histone marks | DNAse | Proteins bound | Motifs changed | NHGRI/EBI GWAS hits | GRASP QTL hits | Selected eQTL hits | GENCODE genes | dbSNP func annot |
|---|---|---|---|---|---|---|---|---|---|---|---|---|---|---|---|---|---|---|---|---|---|
| 10 | 7687641 | 0.95 | 0.98 | rs4749162 | A | G | 0.32 | 0.54 | 0.34 | 0.44 | | | BLD, LIV | 10 tissues | TCF4 | Ik-1,Ik-2 | | | | 16kb 5' of ITIH2 | |
| 10 | 7688147 | 0.89 | 0.97 | rs1972220 | A | T | 0.35 | 0.56 | 0.34 | 0.45 | | | | | | 4 altered motifs | | | | 15kb 5' of ITIH2 | |
| 10 | 7690360 | 0.92 | 0.97 | rs7917456 | A | G | 0.27 | 0.55 | 0.34 | 0.45 | | | | BLD | | | | | | 13kb 5' of ITIH2 | |
| 10 | 7690456 | 0.92 | 0.98 | rs7901428 | C | T | 0.17 | 0.54 | 0.34 | 0.45 | | | | | | RFX5 | | | | 13kb 5' of ITIH2 | |
| 10 | 7691641 | 0.92 | 0.97 | rs10905216 | C | T | 0.27 | 0.55 | 0.34 | 0.45 | | | | | | GR,Klf7 | | | | 12kb 5' of ITIH2 | |
| 10 | 7691653 | 0.92 | 0.97 | rs10905217 | C | A | 0.27 | 0.55 | 0.34 | 0.45 | | | | | | GATA,Pax-4 | | | | 12kb 5' of ITIH2 | |
| 10 | 7691807 | 0.95 | 0.98 | rs7080828 | G | A | 0.26 | 0.54 | 0.34 | 0.45 | | | | | | 4 altered motifs | | | | 11kb 5' of ITIH2 | |
| 10 | 7691935 | 0.91 | 0.98 | rs10905218 | C | T | 0.16 | 0.54 | 0.34 | 0.45 | | | | | | Ets,Nanog,Pou2f2 | | | | 11kb 5' of ITIH2 | |
| 10 | 7692378 | 0.92 | 0.98 | rs7069624 | A | G | 0.21 | 0.54 | 0.34 | 0.44 | | | | | SETDB1 | Nkx2,Pbx-1 | | | | 11kb 5' of ITIH2 | |
| 10 | 7693385 | 0.92 | 0.97 | rs6602266 | T | C | 0.27 | 0.55 | 0.34 | 0.45 | | | | | | TLX1::NFIC | | | | 9.9kb 5' of ITIH2 | |
| 10 | 7693913 | 0.92 | 0.97 | rs2182652 | A | G | 0.27 | 0.55 | 0.34 | 0.45 | | | | | | 4 altered motifs | | | | 9.4kb 5' of ITIH2 | |
| 10 | 7694941 | 1 | 1 | rs11255291 | C | T | 0.26 | 0.55 | 0.34 | 0.44 | | | | | | Pou2f2 | 1 hit | | | 8.3kb 5' of ITIH2 | |

Query SNP: rs1240259 and variants with $r^2$ >= 0.8

| chr | pos (hg38) | LD (r²) | LD (D') | variant | Ref | Alt | AFR freq | AMR freq | ASN freq | EUR freq | SiPhy cons | Promoter histone marks | Enhancer histone marks | DNAse | Proteins bound | Motifs changed | NHGRI/EBI GWAS hits | GRASP QTL hits | Selected eQTL hits | GENCODE genes | dbSNP func annot |
|---|---|---|---|---|---|---|---|---|---|---|---|---|---|---|---|---|---|---|---|---|---|
| 12 | 69840154 | 1 | 1 | rs1240259 | T | C | 0.13 | 0.23 | 0.23 | 0.32 | | | GI | BLD,GI | | | | 6 hits | 10 hits | AC025263.3 | |
| 12 | 69841415 | 1 | 1 | rs1240261 | A | G | 0.14 | 0.23 | 0.22 | 0.32 | | | 4 tissues | PANC,GI | | 8 altered motifs | | | 6 hits | AC025263.3 | |
| 12 | 69852759 | 0.94 | 0.98 | rs1240246 | G | C | 0.11 | 0.22 | 0.24 | 0.32 | | | | | | 5 altered motifs | | | 5 hits | AC025263.3 | |
| 12 | 69853198 | 0.88 | 0.95 | rs1240247 | G | C | 0.14 | 0.23 | 0.23 | 0.32 | | | | | | 19 altered motifs | | | 4 hits | AC025263.3 | |
| 12 | 69853538 | 0.88 | 0.97 | rs1240248 | C | T | 0.11 | 0.22 | 0.22 | 0.31 | | | | | | RBP-Jkappa | | | 5 hits | AC025263.3 | |
| 12 | 69858206 | 1 | 1 | rs2446481 | G | C | 0.11 | 0.23 | 0.22 | 0.32 | | | | | | 4 altered motifs | | | 5 hits | C12orf28 | |
| 12 | 69859472 | 0.95 | 1 | rs1240213 | C | A | 0.11 | 0.23 | 0.22 | 0.32 | | | | | | Pax-4,RXRA | | 5 hits | 8 hits | C12orf28 | |
| 12 | 69860175 | 1 | 1 | rs1261657 | C | A | 0.13 | 0.23 | 0.22 | 0.32 | | | | | | Irf | | | 5 hits | C12orf28 | |
| 12 | 69863262 | 0.97 | 1 | rs1240232 | C | A | 0.11 | 0.22 | 0.22 | 0.32 | | | | | | 5 altered motifs | | | 6 hits | C12orf28 | |

| | | | | | | | | | | | | | | | | | | | | |
|---|---|---|---|---|---|---|---|---|---|---|---|---|---|---|---|---|---|---|---|---|
| 12 | 69863416 | 0.98 | 1 | rs1240231 | C | T | 0.11 | 0.23 | 0.22 | 0.32 | | | | | | 4 altered motifs | | | 5 hits | C12orf28 |
| 12 | 69863681 | 1 | 1 | rs1240230 | A | C | 0.11 | 0.23 | 0.22 | 0.32 | | | | | | Foxa,PU.1,STAT | | | 5 hits | C12orf28 |
| 12 | 69868006 | 1 | 1 | rs1272951 | T | C | 0.11 | 0.23 | 0.23 | 0.32 | | | | | | AIRE | | 5 hits | 8 hits | C12orf28 |
| 12 | 69868908 | 1 | 1 | rs1240228 | G | A | 0.11 | 0.23 | 0.23 | 0.32 | | | GI | ESDR | CMYC | 4 altered motifs | | | 5 hits | C12orf28 |
| 12 | 69869155 | 1 | 1 | rs1240227 | A | G | 0.11 | 0.23 | 0.23 | 0.32 | | | | IPSC | | FAC1,Sox | | | 5 hits | C12orf28 |
| 12 | 69869298 | 1 | 1 | rs1240226 | G | T | 0.11 | 0.23 | 0.22 | 0.32 | | | | | | Foxk1,Foxo,Sox | | 5 hits | 8 hits | C12orf28 |
| 12 | 69869313 | 0.97 | 1 | rs1240225 | A | G | 0.11 | 0.22 | 0.23 | 0.32 | | | | | | HP1-site-factor | | | 5 hits | C12orf28 |
| 12 | 69870472 | 1 | 1 | rs1240223 | T | C | 0.11 | 0.23 | 0.23 | 0.32 | | | | | | E2F,Nr2e3 | | | 5 hits | C12orf28 |
| 12 | 69871766 | 0.98 | 1 | rs1261658 | T | C | 0.11 | 0.22 | 0.23 | 0.32 | | | | | | 12 altered motifs | | 5 hits | 8 hits | C12orf28 |

Query SNP: rs7432667 and variants with $r^2 >= 0.8$

| chr | pos (hg38) | LD ($r^2$) | LD (D') | variant | Ref | Alt | AFR freq | AMR freq | ASN freq | EUR freq | SiPhy cons | Promoter histone marks | Enhancer histone marks | DNAse | Proteins bound | Motifs changed | NHGRI/ EBI GWAS hits | GRASP QTL hits | Selected eQTL hits | GENCODE genes | dbSNP func annot |
|---|---|---|---|---|---|---|---|---|---|---|---|---|---|---|---|---|---|---|---|---|---|
| 3 | 74450183 | 0.94 | 0.99 | rs7645458 | A | T | 0.49 | 0.57 | 0.28 | 0.52 | | | | | | 5 altered motifs | | | | CNTN3 | intronic |
| 3 | 74451655 | 0.81 | 0.94 | rs190873176 | G | A | 0.41 | 0.54 | 0.27 | 0.46 | | | | | | | | | | CNTN3 | intronic |
| 3 | 74453819 | 0.94 | 0.99 | rs7621294 | G | A | 0.49 | 0.57 | 0.28 | 0.52 | | | | | | | | | | CNTN3 | intronic |
| 3 | 74454904 | 0.82 | 0.96 | rs7616738 | T | A | 0.41 | 0.53 | 0.27 | 0.50 | | | | | | ERalpha-a,PEBP | | | | CNTN3 | intronic |
| 3 | 74455810 | 0.96 | 1 | rs9681000 | T | C | 0.49 | 0.57 | 0.28 | 0.53 | | | LNG | | | Pax-6,Pax-8 | | | | CNTN3 | intronic |
| 3 | 74456014 | 0.97 | 1 | rs9680894 | A | G | 0.49 | 0.57 | 0.28 | 0.52 | | | LNG | | | 4 altered motifs | | | | CNTN3 | intronic |
| 3 | 74456226 | 0.94 | 0.98 | rs66623295 | TGAA | T | 0.49 | 0.56 | 0.28 | 0.51 | | | LNG, GI | | | 4 altered motifs | | | | CNTN3 | intronic |
| 3 | 74456252 | 0.94 | 0.98 | rs13063976 | G | T | 0.48 | 0.56 | 0.28 | 0.52 | | | LNG, GI | | | 10 altered motifs | | | | CNTN3 | intronic |
| 3 | 74456365 | 0.99 | 1 | rs13065506 | C | A | 0.45 | 0.56 | 0.28 | 0.52 | | | LNG, GI | LNG,MUS,MUS | | | | | | CNTN3 | intronic |
| 3 | 74456523 | 0.98 | 1 | rs9815102 | C | T | 0.48 | 0.57 | 0.28 | 0.52 | | | LNG, GI | MUS | | 4 altered motifs | | | | CNTN3 | intronic |
| 3 | 74456908 | 0.97 | 0.99 | rs9815683 | C | T | 0.48 | 0.56 | 0.28 | 0.51 | | | | | | 5 altered motifs | | | | CNTN3 | intronic |
| 3 | 74457615 | 0.98 | 1 | rs35003866 | GA | G | 0.48 | 0.57 | 0.28 | 0.52 | | | | LNG | | 5 altered motifs | | | | CNTN3 | intronic |
| 3 | 74458520 | 0.96 | 1 | rs7628916 | T | C | 0.49 | 0.57 | 0.28 | 0.53 | | | | | | Evi-1,SETDB1 | | | | CNTN3 | intronic |
| 3 | 74458615 | 0.96 | 1 | rs7615326 | C | T | 0.49 | 0.57 | 0.28 | 0.53 | | | | | | 13 altered motifs | | | | CNTN3 | intronic |
| 3 | 74459275 | 0.95 | 1 | rs7428834 | G | A | 0.57 | 0.57 | 0.28 | 0.53 | | | | | | HNF6,Maf,Pbx-1 | | | | CNTN3 | intronic |
| 3 | 74459505 | 0.92 | 0.98 | rs9681632 | C | A | 0.57 | 0.57 | 0.29 | 0.52 | | | | | | FXR | | | | CNTN3 | intronic |
| 3 | 74460224 | 0.95 | 1 | rs4677398 | C | T | 0.57 | 0.57 | 0.28 | 0.52 | | | | | | Sox | | | | CNTN3 | intronic |
| 3 | 74460443 | 0.98 | 1 | rs4676970 | G | A | 0.48 | 0.57 | 0.28 | 0.51 | | | | | | 5 altered motifs | | | | CNTN3 | intronic |
| 3 | 74461462 | 0.98 | 1 | rs7629343 | G | A | 0.45 | 0.57 | 0.28 | 0.53 | | | | | | 8 altered motifs | | | | CNTN3 | intronic |
| 3 | 74462547 | 0.93 | 1 | rs34223336 | T | C | 0.59 | 0.58 | 0.28 | 0.53 | | | | | | Evi-1,Hoxa7,NF-I | | | | CNTN3 | intronic |
| 3 | 74463682 | 0.9 | 1 | rs9852645 | C | G | 0.73 | 0.59 | 0.28 | 0.53 | | | ESC, IPSC | 6 tissues | | DBP,HNF1 | | | | CNTN3 | intronic |
| 3 | 74464246 | 0.9 | 1 | rs7429824 | A | G | 0.74 | 0.59 | 0.28 | 0.53 | | | | | | 14 altered motifs | | | | CNTN3 | intronic |
| 3 | 74465249 | 0.95 | 1 | rs9862571 | C | T | 0.57 | 0.57 | 0.28 | 0.53 | | | | | | DMRT2,Nanog | | | | CNTN3 | intronic |
| 3 | 74465927 | 0.96 | 1 | rs7632222 | A | T | 0.58 | 0.57 | 0.28 | 0.52 | | VAS | 10 tissues | BRN,BRN | | GATA,HDAC2,Maf | | | | CNTN3 | intronic |
| 3 | 74466062 | 1 | 1 | rs7432667 | G | C | 0.45 | 0.56 | 0.28 | 0.52 | | VAS | 5 tissues | ESDR,BRST,BRN | | Irf,Pou5f1 | | | | CNTN3 | intronic |
| 3 | 74466101 | 0.98 | 1 | rs7432669 | G | A | 0.45 | 0.56 | 0.28 | 0.52 | | VAS | 5 tissues | LNG,BRST,SKIN | | DMRT7,Nkx2 | | | | CNTN3 | intronic |
| 3 | 74467622 | 0.93 | 1 | rs71277517 | T | TAG | 0.59 | 0.58 | 0.28 | 0.53 | | | BRN | | | Foxp1,GATA,HDAC2 | | | | CNTN3 | intronic |
| 3 | 74468262 | 0.98 | 0.99 | rs7433810 | G | T | 0.42 | 0.56 | 0.28 | 0.53 | | | HRT | | | Pou2f2,Sox | | | | CNTN3 | intronic |
| 3 | 74468299 | 0.93 | 1 | rs7430007 | C | T | 0.59 | 0.58 | 0.28 | 0.53 | | | HRT | | | RP58 | | | | CNTN3 | intronic |
| 3 | 74468377 | 0.87 | 0.94 | rs7432452 | T | A | 0.57 | 0.57 | 0.28 | 0.53 | | | HRT | | | 5 altered motifs | | | | CNTN3 | intronic |
| 3 | 74469317 | 0.91 | 0.99 | rs11719441 | A | T | 0.59 | 0.58 | 0.28 | 0.53 | | | SKIN | 4 tissues | | Nrf1 | | | | CNTN3 | intronic |
| 3 | 74469446 | 0.88 | 0.95 | rs11720391 | T | C | 0.59 | 0.57 | 0.28 | 0.53 | | | SKIN | 4 tissues | | 4 altered motifs | | | | CNTN3 | intronic |
| 3 | 74476520 | 0.93 | 0.98 | rs9826396 | G | A | 0.46 | 0.56 | 0.28 | 0.51 | | | | | | Pbx-1 | | | | CNTN3 | intronic |
| 3 | 74476699 | 0.92 | 0.98 | rs7623053 | C | T | 0.41 | 0.55 | 0.28 | 0.51 | | | | | | 15 altered motifs | | | | CNTN3 | intronic |

Query SNP: rs7302554 and variants with $r^2 >= 0.8$

| chr | pos (hg38) | LD ($r^2$) | LD (D') | variant | Ref | Alt | AFR freq | AMR freq | ASN freq | EUR freq | SiPhy cons | Promoter histone marks | Enhancer histone marks | DNAse | Proteins bound | Motifs changed | NHGRI/ EBI GWAS hits | GRASP QTL hits | Selected eQTL hits | GENCODE genes | dbSNP func annot |
|---|---|---|---|---|---|---|---|---|---|---|---|---|---|---|---|---|---|---|---|---|---|
| 12 | 119092463 | 1 | 1 | rs35463038 | T | TG | 0.80 | 0.76 | 0.47 | 0.65 | | | ESDR | | | 7 altered motifs | | | | SRRM4 | intronic |
| 12 | 119092619 | 1 | 1 | rs7302554 | A | G | 0.79 | 0.76 | 0.47 | 0.65 | | | | | | KAP1,Pax-4,Smad4 | | | | SRRM4 | intronic |
| 12 | 119092916 | 0.96 | 1 | rs7306264 | T | C | 0.70 | 0.75 | 0.47 | 0.65 | | | | | | 6 altered motifs | | | | SRRM4 | intronic |
| 12 | 119094861 | 0.97 | 0.98 | rs7314733 | T | C | 0.79 | 0.76 | 0.47 | 0.65 | | | ESDR, IPSC, ESC | | | Nr2f2,STAT | | | | SRRM4 | intronic |
| 12 | 119101583 | 0.9 | 0.97 | rs12829158 | G | A | 0.80 | 0.75 | 0.47 | 0.65 | | | ESDR | | | | | | | SRRM4 | intronic |

Query SNP: rs6464781 and variants with $r^2 >= 0.8$

| chr | pos (hg38) | LD ($r^2$) | LD (D') | variant | Ref | Alt | AFR freq | AMR freq | ASN freq | EUR freq | SiPhy cons | Promoter histone marks | Enhancer histone marks | DNAse | Proteins bound | Motifs changed | NHGRI/ EBI GWAS hits | GRASP QTL hits | Selected eQTL hits | GENCODE genes | dbSNP func annot |
|---|---|---|---|---|---|---|---|---|---|---|---|---|---|---|---|---|---|---|---|---|---|
| 7 | 147068646 | 1 | 1 | rs10270384 | T | C | 0.74 | 0.71 | 0.77 | 0.72 | | | | BRN | | AP-1,Pax-4 | | | | CNTNAP2 | intronic |
| 7 | 147069636 | 0.99 | 1 | rs6956700 | A | G | 0.74 | 0.71 | 0.77 | 0.72 | | | | | | Foxm1,Ik-2 | | | | CNTNAP2 | intronic |
| 7 | 147069644 | 1 | 1 | rs6960831 | T | C | 0.75 | 0.71 | 0.77 | 0.72 | | | | | | CEBPB,Foxm1 | | | | CNTNAP2 | intronic |
| 7 | 147069722 | 0.96 | 1 | rs6975159 | C | T | 0.71 | 0.70 | 0.77 | 0.72 | | | | | | 14 altered motifs | | | | CNTNAP2 | intronic |
| 7 | 147070312 | 1 | 1 | rs740809 | A | G | 0.74 | 0.71 | 0.76 | 0.72 | | | LNG | | | 9 altered motifs | | | | CNTNAP2 | intronic |
| 7 | 147070484 | 1 | 1 | rs740810 | T | C | 0.74 | 0.71 | 0.76 | 0.72 | | | LNG | | | Myb | | | | CNTNAP2 | intronic |
| 7 | 147070737 | 1 | 1 | rs10260427 | C | G | 0.74 | 0.71 | 0.76 | 0.72 | | | | LNG | | STAT | | | | CNTNAP2 | intronic |
| 7 | 147070968 | 0.97 | 1 | rs740811 | T | C | 0.81 | 0.72 | 0.76 | 0.72 | | | KID | LNG | | CEBPD,SREBP,YY1 | | | | CNTNAP2 | intronic |
| 7 | 147071735 | 1 | 1 | rs6464781 | C | G | 0.74 | 0.71 | 0.76 | 0.72 | | | GI, KID, PANC | GI,GI,GI | | 4 altered motifs | | | | CNTNAP2 | intronic |
| 7 | 147072346 | 1 | 1 | rs6464782 | A | G | 0.74 | 0.71 | 0.76 | 0.72 | | | KID | | | Irf,Pou5f1 | | | | CNTNAP2 | intronic |
| 7 | 147072383 | 1 | 1 | rs6464783 | G | A | 0.74 | 0.71 | 0.76 | 0.72 | | | KID | | | Foxa,Foxd3 | | | | CNTNAP2 | intronic |
| 7 | 147073836 | 0.97 | 1 | rs2191272 | G | T | 0.75 | 0.71 | 0.76 | 0.72 | | | | | | AIRE,Foxj2,Nkx2 | | | | CNTNAP2 | intronic |
| 7 | 147078241 | 0.93 | 0.97 | rs1014686 | G | A | 0.76 | 0.72 | 0.76 | 0.72 | | | | GI,KID | | Gfi1 | | | | CNTNAP2 | intronic |
| 7 | 147085279 | 0.83 | 0.97 | rs10256502 | A | G | 0.73 | 0.69 | 0.74 | 0.72 | | | | | | 6 altered motifs | | | | AC006004.1 | intronic |
| 7 | 147086540 | 0.85 | 1 | rs7802708 | G | A | 0.73 | 0.68 | 0.74 | 0.72 | | | | | | 5 altered motifs | | | | AC006004.1 | intronic |
| 7 | 147087973 | 0.8 | 0.96 | rs9640233 | C | T | 0.76 | 0.69 | 0.74 | 0.72 | | | | | | 8 altered motifs | | | | AC006004.1 | intronic |

Query SNP: rs17046334 and variants with $r^2 >= 0.8$

| chr | pos (hg38) | LD ($r^2$) | LD (D') | variant | Ref | Alt | AFR freq | AMR freq | ASN freq | EUR freq | SiPhy cons | Promoter histone marks | Enhancer histone marks | DNAse | Proteins bound | Motifs changed | NHGRI/ EBI GWAS hits | GRASP QTL hits | Selected eQTL hits | GENCODE genes | dbSNP func annot |
|---|---|---|---|---|---|---|---|---|---|---|---|---|---|---|---|---|---|---|---|---|---|
| 12 | 79072024 | 1 | 1 | rs56198149 | A | T | 0.11 | 0.15 | 0.03 | 0.11 | | | | | | 14 altered motifs | | | | SYT1 | intronic |
| 12 | 79073717 | 1 | 1 | rs17046334 | G | C | 0.10 | 0.15 | 0.03 | 0.11 | | | ESC | | MAFF,MAFF | FAC1,Foxj2,Msx-1 | | | | SYT1 | intronic |
| 12 | 79075262 | 0.85 | 1 | rs11609700 | C | T | 0.07 | 0.13 | 0.03 | 0.11 | | | | | | 5 altered motifs | | | | SYT1 | intronic |
| 12 | 79081015 | 0.96 | 1 | rs17041558 | A | C | 0.08 | 0.15 | 0.03 | 0.11 | | | | | | 4 altered motifs | | | | SYT1 | intronic |
| 12 | 79081478 | 1 | 1 | rs12319380 | A | G | 0.11 | 0.15 | 0.03 | 0.11 | | | | | | AP-1,HNF1,Obox6 | | | | SYT1 | intronic |

Query SNP: rs660586 and variants with $r^2 >= 0.8$

| chr | pos | LD | LD | variant | Ref | Alt | AFR | AMR | ASN | EUR | SiPhy | Promoter | Enhancer | DNAse | Proteins | Motifs | NHGRI/ | GRASP | Selected | GENCODE | dbSNP |
|---|---|---|---|---|---|---|---|---|---|---|---|---|---|---|---|---|---|---|---|---|---|

| chr | pos (hg38) | LD (r²) | LD (D') | variant | Ref | Alt | AFR freq | AMR freq | ASN freq | EUR freq | SiPhy cons | Promoter histone marks | Enhancer histone marks | DNAse | Proteins bound | Motifs changed | NHGRI/EBI GWAS hits | GRASP QTL hits | Selected eQTL hits | GENCODE genes | dbSNP func annot |
|---|---|---|---|---|---|---|---|---|---|---|---|---|---|---|---|---|---|---|---|---|---|
| 6 | 16743148 | 0.91 | 0.99 | rs1143886 | C | T | 0.66 | 0.64 | 0.56 | 0.57 | | | 10 tissues | | | Foxp1 | | | 1 hit | AL137003.1 | intronic |
| 6 | 16743822 | 1 | 1 | rs660586 | G | A | 0.64 | 0.62 | 0.50 | 0.54 | | | 12 tissues | BLD,BLD | PU1 | 5 altered motifs | | | 1 hit | AL137003.1 | intronic |
| 6 | 16743841 | 1 | 1 | rs492412 | A | G | 0.64 | 0.62 | 0.50 | 0.54 | | | 12 tissues | BLD,BLD | PU1 | Nkx2 | | | 1 hit | AL137003.1 | intronic |
| 6 | 16743938 | 0.98 | 1 | rs493352 | T | C | 0.64 | 0.62 | 0.50 | 0.54 | | | 12 tissues | | MAFF,MAFK,PU1 | | | | 1 hit | AL137003.1 | intronic |
| 6 | 16744034 | 0.97 | 1 | rs639621 | T | G | 0.64 | 0.61 | 0.50 | 0.54 | | | 12 tissues | | MAFF,MAFK | Foxd3,Myf,NF-I | | | 1 hit | AL137003.1 | intronic |
| 6 | 16744269 | 0.94 | 0.99 | rs648411 | G | C | 0.64 | 0.61 | 0.50 | 0.54 | | LNG | 13 tissues | | | 5 altered motifs | | | 1 hit | AL137003.1 | intronic |

Query SNP: rs7943132 and variants with $r^2$ >= 0.8

| chr | pos (hg38) | LD (r²) | LD (D') | variant | Ref | Alt | AFR freq | AMR freq | ASN freq | EUR freq | SiPhy cons | Promoter histone marks | Enhancer histone marks | DNAse | Proteins bound | Motifs changed | NHGRI/EBI GWAS hits | GRASP QTL hits | Selected eQTL hits | GENCODE genes | dbSNP func annot |
|---|---|---|---|---|---|---|---|---|---|---|---|---|---|---|---|---|---|---|---|---|---|
| 11 | 71182132 | 1 | 1 | rs7943132 | T | A | 0.63 | 0.52 | 0.54 | 0.39 | | | ESC, BRN | | | | | | | SHANK2 | intronic |

Query SNP: rs9567431 and variants with $r^2$ >= 0.8

| chr | pos (hg38) | LD (r²) | LD (D') | variant | Ref | Alt | AFR freq | AMR freq | ASN freq | EUR freq | SiPhy cons | Promoter histone marks | Enhancer histone marks | DNAse | Proteins bound | Motifs changed | NHGRI/EBI GWAS hits | GRASP QTL hits | Selected eQTL hits | GENCODE genes | dbSNP func annot |
|---|---|---|---|---|---|---|---|---|---|---|---|---|---|---|---|---|---|---|---|---|---|
| 13 | 44424194 | 1 | 1 | rs9567431 | A | G | 0.59 | 0.55 | 0.62 | 0.46 | | | BLD, PLCNT, THYM | PLCNT,SKIN | | En-1 | | | | 9.3kb 3' of TSC22D1 | |
| 13 | 44427868 | 0.93 | 0.99 | rs9567432 | C | T | 0.42 | 0.54 | 0.62 | 0.46 | | | HRT | BLD | | 4 altered motifs | | | | 5.7kb 3' of TSC22D1 | |
| 13 | 44456043 | 0.8 | 0.92 | rs7335836 | T | C | 0.49 | 0.57 | 0.63 | 0.46 | | | GI | GI,GI,GI | | CEBPB,DEC | | | | TSC22D1 | intronic |
| 13 | 44456200 | 0.8 | 0.92 | rs7336215 | T | C | 0.49 | 0.57 | 0.63 | 0.46 | | GI | GI | | | CDP | | | | TSC22D1 | intronic |

Query SNP: rs1105460 and variants with $r^2$ >= 0.8

| chr | pos (hg38) | LD (r²) | LD (D') | variant | Ref | Alt | AFR freq | AMR freq | ASN freq | EUR freq | SiPhy cons | Promoter histone marks | Enhancer histone marks | DNAse | Proteins bound | Motifs changed | NHGRI/EBI GWAS hits | GRASP QTL hits | Selected eQTL hits | GENCODE genes | dbSNP func annot |
|---|---|---|---|---|---|---|---|---|---|---|---|---|---|---|---|---|---|---|---|---|---|
| 18 | 36543779 | 0.93 | 0.97 | rs12604401 | T | C | 0.42 | 0.34 | 0.48 | 0.46 | | | 8 tissues | BRN,BRN,OVRY | | Maf | | | | FHOD3 | intronic |
| 18 | 36544017 | 0.96 | 0.99 | rs12958886 | G | A | 0.35 | 0.34 | 0.48 | 0.46 | | | 8 tissues | BRN | | Cdc5,Hdx | | | | FHOD3 | intronic |
| 18 | 36544074 | 0.96 | 0.99 | rs12958613 | A | G | 0.34 | 0.34 | 0.48 | 0.46 | | SKIN, GI | 11 tissues | BRN | | 10 altered motifs | | | | FHOD3 | intronic |
| 18 | 36544820 | 1 | 1 | rs1105460 | A | G | 0.35 | 0.33 | 0.48 | 0.46 | | SKIN | 10 tissues | 4 tissues | | Roaz | | | | FHOD3 | intronic |
| 18 | 36544987 | 1 | 1 | rs1105459 | A | G | 0.35 | 0.33 | 0.48 | 0.46 | | | 5 tissues | | | PLZF,RXRA,SREBP | | | | FHOD3 | intronic |
| 18 | 36545606 | 0.99 | 1 | rs12964583 | A | C | 0.35 | 0.33 | 0.48 | 0.46 | | | 4 tissues | | | Gfi1,RREB-1 | | | | FHOD3 | intronic |
| 18 | 36545649 | 1 | 1 | rs12964847 | C | T | 0.35 | 0.33 | 0.48 | 0.46 | | | 4 tissues | BRN | | | | | | FHOD3 | intronic |
| 18 | 36545951 | 1 | 1 | rs12968765 | A | G | 0.35 | 0.33 | 0.48 | 0.46 | | | 4 tissues | BRN,BRN,LNG | | 6 altered motifs | | | | FHOD3 | intronic |
| 18 | 36546687 | 0.88 | 1 | rs12971018 | T | C | 0.35 | 0.31 | 0.48 | 0.42 | | | 4 tissues | IPSC | | 4 altered motifs | | | | FHOD3 | intronic |
| 18 | 36546785 | 0.88 | 1 | rs11665489 | G | A | 0.35 | 0.31 | 0.48 | 0.42 | | | 4 tissues | | | Egr-1,GR | | | 1 hit | FHOD3 | intronic |
| 18 | 36546973 | 0.88 | 1 | rs12956952 | T | A | 0.35 | 0.31 | 0.48 | 0.42 | | | ESDR, HRT, MUS | | | HNF4,Nr2f2 | | | 1 hit | FHOD3 | intronic |
| 18 | 36547057 | 1 | 1 | rs12957090 | T | C | 0.35 | 0.33 | 0.48 | 0.46 | | | ESDR, HRT, MUS | HRT | | 8 altered motifs | | | | FHOD3 | intronic |
| 18 | 36547229 | 0.87 | 1 | rs11659739 | G | T | 0.36 | 0.30 | 0.48 | 0.42 | | | ESDR, HRT, MUS | ESC | | Irf,TCF12 | | | 1 hit | FHOD3 | intronic |
| 18 | 36547271 | 0.85 | 1 | rs3947413 | C | T | 0.35 | 0.30 | 0.48 | 0.42 | | | HRT | | | 5 altered motifs | | 1 hit | 1 hit | FHOD3 | intronic |

Query SNP: rs12354965 and variants with $r^2$ >= 0.8

| chr | pos (hg38) | LD (r²) | LD (D') | variant | Ref | Alt | AFR freq | AMR freq | ASN freq | EUR freq | SiPhy cons | Promoter histone marks | Enhancer histone marks | DNAse | Proteins bound | Motifs changed | NHGRI/EBI GWAS hits | GRASP QTL hits | Selected eQTL hits | GENCODE genes | dbSNP func annot |
|---|---|---|---|---|---|---|---|---|---|---|---|---|---|---|---|---|---|---|---|---|---|
| 10 | 77043958 | 0.86 | 0.97 | rs12248504 | C | T | 0.10 | 0.10 | 0.00 | 0.16 | | | BRN | | | RXRA | | | | KCNMA1 | intronic |
| 10 | 77086847 | 1 | 1 | rs12354965 | T | C | 0.04 | 0.11 | 0.00 | 0.16 | | | BRST, SKIN | ESDR,IPSC,MUS | | HNF4 | | 1 hit | | KCNMA1 | intronic |
| 10 | 77099051 | 0.97 | 1 | rs35993060 | T | C | 0.02 | 0.11 | 0.00 | 0.16 | | | | | | | | | | KCNMA1 | intronic |
| 10 | 77106415 | 0.95 | 1 | rs10824490 | C | T | 0.14 | 0.12 | 0.00 | 0.16 | | | | 6 tissues | GATA2,P300 | LBP-9,VDR | | | | KCNMA1 | intronic |

Query SNP: rs41365345 and variants with $r^2$ >= 0.8

| chr | pos (hg38) | LD (r²) | LD (D') | variant | Ref | Alt | AFR freq | AMR freq | ASN freq | EUR freq | SiPhy cons | Promoter histone marks | Enhancer histone marks | DNAse | Proteins bound | Motifs changed | NHGRI/EBI GWAS hits | GRASP QTL hits | Selected eQTL hits | GENCODE genes | dbSNP func annot |
|---|---|---|---|---|---|---|---|---|---|---|---|---|---|---|---|---|---|---|---|---|---|
| 3 | 120693644 | 0.82 | 0.95 | rs142261370 | TA | T | 0.10 | 0.21 | 0.13 | 0.33 | | IPSC, LNG | 18 tissues | 25 tissues | 15 bound proteins | 12 altered motifs | | | 2 hits | RABL3 | intronic |
| 3 | 120695938 | 0.91 | 0.95 | rs34319994 | A | G | 0.16 | 0.23 | 0.15 | 0.35 | | | IPSC, LNG, LIV | | | 5 altered motifs | | | 1 hit | RABL3 | intronic |
| 3 | 120700826 | 0.91 | 0.95 | rs111467306 | G | A | 0.16 | 0.23 | 0.15 | 0.35 | | | | | | RFX5 | | | 2 hits | RABL3 | intronic |
| 3 | 120709582 | 0.88 | 0.95 | rs200379238 | T | TAAG | 0.16 | 0.23 | 0.15 | 0.34 | | | | | | 10 altered motifs | | | 1 hit | RABL3 | intronic |
| 3 | 120709583 | 0.91 | 0.95 | rs111603672 | A | 12-mer | 0.16 | 0.23 | 0.15 | 0.34 | | | | | | 6 altered motifs | | | 1 hit | RABL3 | intronic |
| 3 | 120709774 | 0.91 | 0.95 | rs11720353 | T | C | 0.16 | 0.23 | 0.15 | 0.35 | | | | | | Dbx1,Ncx | | | 3 hits | RABL3 | intronic |
| 3 | 120714092 | 0.89 | 0.95 | rs11713594 | A | G | 0.17 | 0.22 | 0.15 | 0.34 | | | | | | ATF3 | | | 2 hits | RABL3 | intronic |
| 3 | 120718051 | 0.91 | 0.95 | rs11708525 | T | C | 0.16 | 0.23 | 0.15 | 0.34 | | | IPSC, BLD | | | 4 altered motifs | | | 4 hits | RABL3 | intronic |
| 3 | 120719017 | 0.91 | 0.95 | rs6779005 | T | A | 0.16 | 0.23 | 0.15 | 0.34 | | 4 tissues | 4 tissues | | | Eomes,PU.1 | | | 2 hits | RABL3 | intronic |
| 3 | 120722930 | 0.8 | 0.96 | rs201180949 | 10-mer | C | 0.10 | 0.20 | 0.13 | 0.32 | | | | | | 4 altered motifs | | | 1 hit | RABL3 | intronic |
| 3 | 120734844 | 0.88 | 0.95 | rs77380805 | A | G | 0.15 | 0.22 | 0.12 | 0.34 | | | | | | 17 altered motifs | | | 1 hit | RABL3 | intronic |
| 3 | 120737485 | 0.97 | 0.98 | rs7637037 | G | A | 0.10 | 0.23 | 0.15 | 0.34 | | | | | | | | | 1 hit | RABL3 | intronic |
| 3 | 120742729 | 0.97 | 0.98 | rs7647895 | T | C | 0.16 | 0.23 | 0.15 | 0.34 | | 24 tissues | | 53 tissues | 28 bound proteins | 6 altered motifs | | | 3 hits | GTF2E1 | 5'-UTR |
| 3 | 120752005 | 0.82 | 1 | rs75862706 | T | C | 0.05 | 0.19 | 0.15 | 0.30 | | | | | | 23 altered motifs | | | 1 hit | GTF2E1 | intronic |
| 3 | 120752248 | 0.8 | 0.98 | rs74800432 | C | T | 0.07 | 0.20 | 0.15 | 0.31 | | | IPSC | IPSC | | | | | | GTF2E1 | intronic |
| 3 | 120754723 | 0.82 | 1 | rs17243218 | C | T | 0.07 | 0.19 | 0.14 | 0.30 | | | 8 tissues | | | NF-kappaB,NRSF,SP1 | | 1 hit | 2 hits | GTF2E1 | intronic |
| 3 | 120754872 | 1 | 1 | rs17183618 | A | T | 0.16 | 0.23 | 0.15 | 0.34 | | | IPSC, STRM, BLD | | | 6 altered motifs | | | 3 hits | GTF2E1 | intronic |
| 3 | 120759205 | 0.82 | 1 | rs111579154 | G | A | 0.07 | 0.19 | 0.15 | 0.30 | | | | | | 5 altered motifs | | | | GTF2E1 | intronic |
| 3 | 120763028 | 0.82 | 1 | rs77756602 | A | T | 0.07 | 0.19 | 0.15 | 0.31 | | | 5 tissues | | | Foxk1,Foxo | | | 1 hit | GTF2E1 | intronic |
| 3 | 120764390 | 1 | 1 | rs41365345 | T | C | 0.16 | 0.23 | 0.15 | 0.34 | | | 10 tissues | 5 tissues | | STAT | | | 2 hits | GTF2E1 | intronic |
| 3 | 120771079 | 1 | 1 | rs2877413 | G | T | 0.16 | 0.23 | 0.15 | 0.34 | | | ESC, IPSC, BLD | | | | | | 3 hits | GTF2E1 | intronic |
| 3 | 120772109 | 0.82 | 1 | rs11719207 | C | T | 0.07 | 0.19 | 0.15 | 0.31 | | | 5 tissues | | | 7 altered motifs | | | 1 hit | GTF2E1 | intronic |
| 3 | 120773153 | 0.8 | 0.98 | rs147468460 | AAAAC | A | 0.05 | 0.20 | 0.15 | 0.31 | | | 4 tissues | | | 6 altered motifs | | | 1 hit | GTF2E1 | intronic |
| 3 | 120773581 | 1 | 1 | rs6769985 | A | G | 0.10 | 0.23 | 0.15 | 0.34 | | | 4 tissues | | | Nrf-2 | | | 1 hit | GTF2E1 | intronic |
| 3 | 120777195 | 1 | 1 | rs2292170 | G | C | 0.16 | 0.23 | 0.15 | 0.34 | | | | | | Pax-5,SRF | | | 3 hits | GTF2E1 | intronic |
| 3 | 120781599 | 1 | 1 | rs2229308 | T | A | 0.16 | 0.23 | 0.15 | 0.34 | | 4 tissues | 4 tissues | | | Pbx-1,Pbx3,Znf143 | | | 3 hits | GTF2E1 | 3'-UTR |

| | | | | | | | | | | | | | | | | | | | | | |
|---|---|---|---|---|---|---|---|---|---|---|---|---|---|---|---|---|---|---|---|---|---|
| 3 | 120781872 | 0.97 | 1 | rs55697401 | T | C | 0.14 | 0.23 | 0.13 | 0.34 | | LNG, MUS | 4 tissues | | | TCF12 | | | 1 hit | GTF2E1 | 3'-UTR |
| 3 | 120784621 | 1 | 1 | rs17184204 | C | G | 0.11 | 0.23 | 0.15 | 0.34 | | | ESDR | | | 5 altered motifs | | | 2 hits | 1.6kb 3' of GTF2E1 | |
| 3 | 120785113 | 0.98 | 1 | rs11719896 | A | G | 0.10 | 0.22 | 0.15 | 0.34 | | | | | | NRSF | | | 1 hit | 2kb 3' of GTF2E1 | |
| 3 | 120785903 | 0.82 | 1 | rs11712360 | G | A | 0.05 | 0.19 | 0.15 | 0.31 | | LNG, GI, BLD | 7 tissues | 5 tissues | | NRSF | | | 1 hit | 2.8kb 3' of GTF2E1 | |
| 3 | 120786180 | 1 | 1 | rs111303747 | G | A | 0.11 | 0.23 | 0.15 | 0.34 | | LNG, BLD | 4 tissues | 4 tissues | 5 bound proteins | 4 altered motifs | | | 1 hit | 3.1kb 3' of GTF2E1 | |
| 3 | 120787969 | 1 | 1 | rs73183728 | G | A | 0.10 | 0.23 | 0.15 | 0.35 | | | | | | 6 altered motifs | | | 1 hit | 4.9kb 3' of GTF2E1 | |
| 3 | 120788258 | 0.8 | 1 | rs145456169 | TTAGA | T | 0.07 | 0.19 | 0.15 | 0.30 | | | | | | AFP1,AP-1,Pou2f2 | | | 1 hit | 5.2kb 3' of GTF2E1 | |
| 3 | 120788565 | 0.82 | 1 | rs17243933 | G | A | 0.07 | 0.19 | 0.15 | 0.31 | | | | | | MZF1::1-4,YY1 | | | 1 hit | 5.5kb 3' of GTF2E1 | |
| 3 | 120789752 | 0.8 | 0.98 | rs111689703 | G | A,T | 0.05 | 0.20 | 0.15 | 0.31 | | | | | | | | | 1 hit | 6.7kb 3' of GTF2E1 | |
| 3 | 120789984 | 1 | 1 | rs73183730 | A | G | 0.10 | 0.23 | 0.15 | 0.35 | | | | | | Sox | | | 1 hit | 6.9kb 3' of GTF2E1 | |
| 3 | 120791343 | 0.94 | 0.97 | rs201863609 | T | TG,TGT | 0.12 | 0.23 | 0.14 | 0.33 | | | 5 tissues | | | | | | 1 hit | 8.3kb 3' of GTF2E1 | |
| 3 | 120791862 | 0.98 | 1 | rs55869122 | A | G | 0.13 | 0.23 | 0.15 | 0.35 | | | 5 tissues | | | | | | 2 hits | 8.8kb 3' of GTF2E1 | |

Query SNP: rs9397199 and variants with $r^2$ >= 0.8

| chr | pos (hg38) | LD ($r^2$) | LD (D') | variant | Ref | Alt | AFR freq | AMR freq | ASN freq | EUR freq | SiPhy cons | Promoter histone marks | Enhancer histone marks | DNAse | Proteins bound | Motifs changed | NHGRI/EBI GWAS hits | GRASP QTL hits | Selected eQTL hits | GENCODE genes | dbSNP func annot |
|---|---|---|---|---|---|---|---|---|---|---|---|---|---|---|---|---|---|---|---|---|---|
| 6 | 154698089 | 1 | 1 | rs9397199 | G | T | 0.16 | 0.32 | 0.56 | 0.25 | | | BLD | BLD | YY1 | ERalpha-a | | | 1 hit | 35kb 5' of SCAF8 | |

Query SNP: rs6430728 and variants with $r^2$ >= 0.8

| chr | pos (hg38) | LD ($r^2$) | LD (D') | variant | Ref | Alt | AFR freq | AMR freq | ASN freq | EUR freq | SiPhy cons | Promoter histone marks | Enhancer histone marks | DNAse | Proteins bound | Motifs changed | NHGRI/EBI GWAS hits | GRASP QTL hits | Selected eQTL hits | GENCODE genes | dbSNP func annot |
|---|---|---|---|---|---|---|---|---|---|---|---|---|---|---|---|---|---|---|---|---|---|
| 2 | 137526181 | 0.81 | 0.95 | rs10196011 | T | C | 0.84 | 0.55 | 0.36 | 0.51 | | | | LNG | | Osr | | | | THSD7B | intronic |
| 2 | 137527813 | 0.81 | 0.95 | rs995287 | T | G | 0.84 | 0.55 | 0.36 | 0.51 | | | | | | Ets,VDR | | | | THSD7B | intronic |
| 2 | 137531186 | 0.89 | 0.96 | rs1375358 | G | C | 0.76 | 0.51 | 0.36 | 0.49 | | | | LNG | | Ets,NERF1a,PU.1 | | | | THSD7B | intronic |
| 2 | 137541308 | 0.91 | 0.96 | rs1584436 | A | G | 0.83 | 0.52 | 0.36 | 0.49 | | | | | | 6 altered motifs | | | | THSD7B | intronic |
| 2 | 137550244 | 0.98 | 1 | rs10177189 | T | C,G | 0.84 | 0.52 | 0.36 | 0.50 | | | | | FOXA1 | | | | | THSD7B | intronic |
| 2 | 137551348 | 1 | 1 | rs6430728 | G | A | 0.84 | 0.52 | 0.35 | 0.49 | | | | | | 6 altered motifs | | | | THSD7B | intronic |
| 2 | 137554615 | 1 | 1 | rs7604024 | A | G | 0.84 | 0.52 | 0.35 | 0.49 | | | | | | AP-1 | | | | THSD7B | intronic |
| 2 | 137555602 | 0.98 | 1 | rs6430729 | G | A | 0.80 | 0.51 | 0.36 | 0.49 | | | | | | 6 altered motifs | | | | THSD7B | intronic |
| 2 | 137557259 | 0.97 | 0.99 | rs7559285 | A | G | 0.82 | 0.52 | 0.35 | 0.49 | | | | | | RREB-1,TBX5,TCF12 | | | | THSD7B | intronic |
| 2 | 137559546 | 0.91 | 0.98 | rs7569823 | A | C | 0.77 | 0.51 | 0.34 | 0.48 | | | | | | Foxj1 | | | | THSD7B | intronic |
| 2 | 137560526 | 0.82 | 0.96 | rs6705253 | C | A | 0.64 | 0.49 | 0.36 | 0.49 | | | | | | GR,SMC3 | | | | THSD7B | intronic |
| 2 | 137560539 | 0.84 | 0.94 | rs6723115 | A | C | 0.64 | 0.51 | 0.34 | 0.48 | | | | | | MZF1::1-4 | | | | THSD7B | intronic |

Query SNP: rs7716581 and variants with $r^2$ >= 0.8

| chr | pos (hg38) | LD ($r^2$) | LD (D') | variant | Ref | Alt | AFR freq | AMR freq | ASN freq | EUR freq | SiPhy cons | Promoter histone marks | Enhancer histone marks | DNAse | Proteins bound | Motifs changed | NHGRI/EBI GWAS hits | GRASP QTL hits | Selected eQTL hits | GENCODE genes | dbSNP func annot |
|---|---|---|---|---|---|---|---|---|---|---|---|---|---|---|---|---|---|---|---|---|---|
| 5 | 117336927 | 0.89 | 0.96 | rs5870753 | CT | C | 0.72 | 0.31 | 0.14 | 0.38 | | | ESDR | | | 11 altered motifs | | | | 79kb 5' of CTC-504A5.1 | |
| 5 | 117337324 | 0.89 | 0.96 | rs6861880 | A | C | 0.74 | 0.31 | 0.14 | 0.37 | | | | | | AIRE,Nr2f2,RXRA | | | | 78kb 5' of CTC-504A5.1 | |
| 5 | 117337344 | 0.89 | 0.96 | rs6883430 | T | C | 0.74 | 0.31 | 0.14 | 0.37 | | | | | | 8 altered motifs | | | | 78kb 5' of CTC-504A5.1 | |
| 5 | 117337371 | 0.89 | 0.96 | rs4266448 | G | T | 0.74 | 0.31 | 0.14 | 0.37 | | | | | | 4 altered motifs | | | | 78kb 5' of CTC-504A5.1 | |
| 5 | 117337574 | 0.89 | 0.96 | rs4481380 | C | A | 0.73 | 0.31 | 0.13 | 0.37 | | | GI | | | GATA,NRSF,Sox | | | | 78kb 5' of CTC-504A5.1 | |
| 5 | 117337598 | 0.89 | 0.96 | rs4547957 | T | C | 0.73 | 0.31 | 0.13 | 0.37 | | | GI | | | CTCF,RFX5,Zbtb12 | | | | 78kb 5' of CTC-504A5.1 | |
| 5 | 117338096 | 0.81 | 0.92 | rs34205737 | AG | A | 0.71 | 0.29 | 0.13 | 0.36 | | | GI | | | 5 altered motifs | | | | 77kb 5' of CTC-504A5.1 | |
| 5 | 117338219 | 0.85 | 0.92 | rs11948331 | T | G | 0.69 | 0.30 | 0.13 | 0.37 | | | | | | 21 altered motifs | | | | 77kb 5' of CTC-504A5.1 | |
| 5 | 117338903 | 0.85 | 0.92 | rs6595035 | C | T | 0.69 | 0.30 | 0.13 | 0.36 | | | | | | 4 altered motifs | | | | 77kb 5' of CTC-504A5.1 | |
| 5 | 117340406 | 0.87 | 0.95 | rs149028599 | C | G | 0.72 | 0.31 | 0.13 | 0.37 | | | | | | 7 altered motifs | | | | 75kb 5' of CTC-504A5.1 | |
| 5 | 117343428 | 0.86 | 0.96 | rs7707308 | G | C | 0.67 | 0.29 | 0.13 | 0.37 | | | | | | RP58,RREB-1,ZEB1 | | | | 72kb 5' of CTC-504A5.1 | |
| 5 | 117343849 | 0.9 | 0.96 | rs115308308 | T | C | 0.68 | 0.30 | 0.13 | 0.37 | | LIV | | 4 tissues | | 22 altered motifs | | | | 72kb 5' of CTC-504A5.1 | |
| 5 | 117343906 | 0.92 | 0.96 | rs139383525 | G | T | 0.73 | 0.30 | 0.13 | 0.37 | | 4 tissues | ESDR | | | Egr-1 | | | | 72kb 5' of CTC-504A5.1 | |
| 5 | 117343987 | 0.9 | 0.96 | rs7708255 | G | A | 0.70 | 0.30 | 0.13 | 0.37 | | 4 tissues | ESDR | | | PTF1-beta,RBP-Jkappa,YY1 | | | | 72kb 5' of CTC-504A5.1 | |
| 5 | 117344018 | 0.85 | 0.96 | rs35130393 | C | G | 0.69 | 0.28 | 0.11 | 0.36 | | 4 tissues | ESDR | | | ERalpha-a,NF-kappaB,RXRA | | | | 71kb 5' of CTC-504A5.1 | |
| 5 | 117344034 | 0.87 | 0.96 | rs35533033 | G | T | 0.69 | 0.29 | 0.11 | 0.36 | | 4 tissues | ESDR | | | E2A,ZEB1 | | | | 71kb 5' of CTC-504A5.1 | |
| 5 | 117344307 | 0.92 | 0.96 | rs7712524 | G | A | 0.73 | 0.30 | 0.13 | 0.37 | | 4 tissues | ESC, ESDR | | | 10 altered motifs | | | | 71kb 5' of CTC-504A5.1 | |
| 5 | 117344460 | 0.92 | 0.96 | rs7732461 | T | G | 0.73 | 0.30 | 0.13 | 0.37 | | 4 tissues | ESC, ESDR | | | HDAC2,Nanog,TCF4 | | | | 71kb 5' of CTC-504A5.1 | |
| 5 | 117344462 | 0.92 | 0.96 | rs7712311 | A | G | 0.73 | 0.30 | 0.13 | 0.37 | | 4 tissues | ESC, ESDR | | | Bcl6b | | | | 71kb 5' of CTC-504A5.1 | |
| 5 | 117344840 | 0.92 | 0.96 | rs11749394 | G | T | 0.73 | 0.30 | 0.13 | 0.37 | | | | | | GATA | | | | 71kb 5' of CTC-504A5.1 | |
| 5 | 117344924 | 0.92 | 0.96 | rs10061651 | A | C | 0.73 | 0.30 | 0.13 | 0.37 | | | | | | Zbtb12 | | | | 71kb 5' of CTC-504A5.1 | |
| 5 | 117344962 | 0.92 | 0.96 | rs4552663 | C | T | 0.73 | 0.30 | 0.13 | 0.37 | | | | | | 5 altered motifs | | | | 71kb 5' of CTC-504A5.1 | |
| 5 | 117345064 | 0.92 | 0.96 | rs4457117 | C | T | 0.73 | 0.30 | 0.13 | 0.37 | | | | | | Nkx2,Nkx3 | | | | 70kb 5' of CTC-504A5.1 | |
| 5 | 117345101 | 0.92 | 0.96 | rs4443456 | T | C | 0.73 | 0.30 | 0.13 | 0.37 | | | | | | CDP,Mrg,Pbx-1 | | | | 70kb 5' of CTC-504A5.1 | |
| 5 | 117345384 | 0.95 | 0.99 | rs6595039 | G | A | 0.73 | 0.30 | 0.13 | 0.37 | | | | | | 5 altered motifs | | | | 70kb 5' of CTC-504A5.1 | |
| 5 | 117345404 | 0.94 | 0.97 | rs6595040 | T | C | 0.73 | 0.30 | 0.13 | 0.37 | | | | | | | | | | 70kb 5' of CTC-504A5.1 | |

| | | | | | | | | | | | | | | |
|---|---|---|---|---|---|---|---|---|---|---|---|---|---|---|
| 5 | 117345447 | 0.95 | 0.99 | rs4623185 | A | G | 0.73 | 0.30 | 0.13 | 0.37 | | | Irf | 70kb 5' of CTC-504A5.1 |
| 5 | 117345544 | 0.96 | 0.99 | rs4448037 | T | C | 0.73 | 0.30 | 0.13 | 0.37 | | | RXRA,TAL1 | 70kb 5' of CTC-504A5.1 |
| 5 | 117345772 | 0.97 | 0.99 | rs4568397 | C | T | 0.73 | 0.30 | 0.13 | 0.37 | | | 4 altered motifs | 70kb 5' of CTC-504A5.1 |
| 5 | 117346098 | 0.97 | 0.99 | rs13187413 | G | A | 0.73 | 0.30 | 0.13 | 0.37 | | | Irf,Pou2f2,YY1 | 69kb 5' of CTC-504A5.1 |
| 5 | 117346212 | 0.96 | 0.99 | rs6879112 | C | T | 0.73 | 0.30 | 0.13 | 0.37 | | | 11 altered motifs | 69kb 5' of CTC-504A5.1 |
| 5 | 117346241 | 0.96 | 0.99 | rs6860005 | T | C | 0.73 | 0.30 | 0.13 | 0.37 | | | | 69kb 5' of CTC-504A5.1 |
| 5 | 117346687 | 0.97 | 0.99 | rs4593300 | C | A | 0.75 | 0.30 | 0.13 | 0.37 | | | Foxp1,Hdx | 69kb 5' of CTC-504A5.1 |
| 5 | 117346791 | 0.97 | 0.99 | rs4464733 | G | A | 0.73 | 0.30 | 0.13 | 0.37 | | | Nkx2 | 69kb 5' of CTC-504A5.1 |
| 5 | 117346805 | 0.97 | 0.99 | rs4464734 | G | A | 0.73 | 0.30 | 0.13 | 0.37 | | | AhR::Arnt,Arnt,FAC1 | 69kb 5' of CTC-504A5.1 |
| 5 | 117347570 | 0.97 | 0.99 | rs4458619 | C | T | 0.73 | 0.30 | 0.13 | 0.37 | 4 tissues | | Crx,Nkx3 | 68kb 5' of CTC-504A5.1 |
| 5 | 117347589 | 0.97 | 0.99 | rs4629633 | A | G | 0.73 | 0.30 | 0.13 | 0.37 | 4 tissues | | | 68kb 5' of CTC-504A5.1 |
| 5 | 117347700 | 0.97 | 0.99 | rs4469226 | G | C | 0.73 | 0.30 | 0.13 | 0.37 | 4 tissues | | 4 altered motifs | 68kb 5' of CTC-504A5.1 |
| 5 | 117347864 | 0.96 | 0.99 | rs7447809 | A | G | 0.73 | 0.30 | 0.13 | 0.37 | ESDR, ESC, VAS | | BDP1 | 68kb 5' of CTC-504A5.1 |
| 5 | 117347970 | 0.97 | 0.99 | rs13159487 | A | G | 0.73 | 0.30 | 0.13 | 0.37 | VAS | | GR,Sin3Ak-20,p300 | 68kb 5' of CTC-504A5.1 |
| 5 | 117348367 | 0.97 | 0.99 | rs10069843 | A | G | 0.75 | 0.30 | 0.13 | 0.37 | | | 5 altered motifs | 67kb 5' of CTC-504A5.1 |
| 5 | 117348929 | 0.97 | 0.99 | rs10039799 | T | C | 0.75 | 0.30 | 0.13 | 0.37 | | | 9 altered motifs | 67kb 5' of CTC-504A5.1 |
| 5 | 117349011 | 0.97 | 0.99 | rs10039823 | T | C | 0.73 | 0.30 | 0.13 | 0.37 | | | 6 altered motifs | 66kb 5' of CTC-504A5.1 |
| 5 | 117349236 | 0.97 | 0.99 | rs10036986 | G | T | 0.73 | 0.30 | 0.13 | 0.37 | | | DMRT2,HNF1,Sox | 66kb 5' of CTC-504A5.1 |
| 5 | 117349566 | 0.97 | 0.99 | rs61332533 | T | C | 0.73 | 0.30 | 0.13 | 0.37 | | | HNF1,RFX5,RORalpha1 | 66kb 5' of CTC-504A5.1 |
| 5 | 117349652 | 0.97 | 0.99 | rs62380367 | T | C | 0.73 | 0.30 | 0.13 | 0.37 | | | DMRT3,DMRT5,Lhx3 | 66kb 5' of CTC-504A5.1 |
| 5 | 117349743 | 0.95 | 0.99 | rs34937570 | A | G | 0.74 | 0.30 | 0.13 | 0.37 | | | | 66kb 5' of CTC-504A5.1 |
| 5 | 117349801 | 0.8 | 0.94 | rs62380369 | T | A | 0.65 | 0.28 | 0.11 | 0.33 | | | 4 altered motifs | 66kb 5' of CTC-504A5.1 |
| 5 | 117349802 | 0.82 | 0.94 | rs62380370 | A | G | 0.65 | 0.28 | 0.11 | 0.33 | | | Hoxa7,Pou2f2 | 66kb 5' of CTC-504A5.1 |
| 5 | 117349837 | 0.97 | 0.99 | rs34205076 | A | T | 0.75 | 0.30 | 0.13 | 0.37 | | | 5 altered motifs | 66kb 5' of CTC-504A5.1 |
| 5 | 117349916 | 0.95 | 0.99 | rs34227028 | G | A | 0.73 | 0.30 | 0.13 | 0.36 | | | Hdx | 66kb 5' of CTC-504A5.1 |
| 5 | 117349998 | 0.92 | 0.99 | rs34173130 | G | A | 0.72 | 0.29 | 0.13 | 0.36 | | | Pax-4 | 66kb 5' of CTC-504A5.1 |
| 5 | 117350048 | 0.85 | 0.99 | rs10073605 | T | G | 0.65 | 0.27 | 0.11 | 0.33 | | | | 65kb 5' of CTC-504A5.1 |
| 5 | 117350325 | 0.97 | 0.99 | rs4259204 | A | G | 0.75 | 0.30 | 0.13 | 0.37 | | | Cdx,HDAC2,RFX5 | 65kb 5' of CTC-504A5.1 |
| 5 | 117350517 | 0.97 | 0.99 | rs4563664 | T | A | 0.75 | 0.30 | 0.13 | 0.37 | | | 39 altered motifs | 65kb 5' of CTC-504A5.1 |
| 5 | 117350601 | 0.96 | 0.99 | rs4566827 | G | A | 0.71 | 0.30 | 0.13 | 0.37 | | | 6 altered motifs | 65kb 5' of CTC-504A5.1 |
| 5 | 117350611 | 0.95 | 0.99 | rs4498285 | T | C | 0.73 | 0.30 | 0.13 | 0.37 | | | 4 altered motifs | 65kb 5' of CTC-504A5.1 |
| 5 | 117350622 | 0.97 | 0.99 | rs5011797 | T | C | 0.75 | 0.30 | 0.13 | 0.37 | | | 16 altered motifs | 65kb 5' of CTC-504A5.1 |
| 5 | 117350642 | 0.97 | 0.99 | rs6869527 | T | C | 0.75 | 0.30 | 0.13 | 0.37 | | | 6 altered motifs | 65kb 5' of CTC-504A5.1 |
| 5 | 117350931 | 0.97 | 0.99 | rs6870045 | T | C | 0.74 | 0.30 | 0.13 | 0.37 | | | 5 altered motifs | 65kb 5' of CTC-504A5.1 |
| 5 | 117350951 | 0.97 | 0.99 | rs6888983 | C | T | 0.74 | 0.30 | 0.13 | 0.37 | | | STAT,TEF-1 | 65kb 5' of CTC-504A5.1 |
| 5 | 117350957 | 0.97 | 0.99 | rs6870063 | T | A | 0.74 | 0.30 | 0.13 | 0.37 | | | STAT | 65kb 5' of CTC-504A5.1 |
| 5 | 117351418 | 0.97 | 0.99 | rs6595041 | T | C | 0.74 | 0.30 | 0.13 | 0.37 | ESDR | ESDR | 7 altered motifs | 64kb 5' of CTC-504A5.1 |
| 5 | 117351641 | 0.94 | 0.99 | rs7733545 | G | T | 0.65 | 0.29 | 0.13 | 0.35 | | | 7 altered motifs | 64kb 5' of CTC-504A5.1 |
| 5 | 117355637 | 1 | 1 | rs7716581 | A | G | 0.74 | 0.30 | 0.13 | 0.37 | | | Gfi1,HNF1,Pax-5 | 60kb 5' of CTC-504A5.1 |
| 5 | 117356511 | 1 | 1 | rs13172380 | T | C | 0.74 | 0.30 | 0.13 | 0.37 | | | SIX5,ZEB1 | 59kb 5' of CTC-504A5.1 |

Query SNP: rs6778643 and variants with $r^2$ >= 0.8

| chr | pos (hg38) | LD ($r^2$) | LD (D') | variant | Ref | Alt | AFR freq | AMR freq | ASN freq | EUR freq | SiPhy cons | Promoter histone marks | Enhancer histone marks | DNAse | Proteins bound | Motifs changed | NHGRI/EBI GWAS hits | GRASP QTL hits | Selected eQTL hits | GENCODE genes | dbSNP func annot |
|---|---|---|---|---|---|---|---|---|---|---|---|---|---|---|---|---|---|---|---|---|---|
| 3 | 124851971 | 0.87 | 1 | rs3772851 | T | G | 0.40 | 0.37 | 0.70 | 0.40 | | | 16 tissues | ESC | | 7 altered motifs | | | 6 hits | ITGB5 | intronic |
| 3 | 124852112 | 0.88 | 1 | rs3772852 | C | G | 0.40 | 0.38 | 0.70 | 0.40 | | | 16 tissues | | | Pax-2,ZBTB33 | | | 6 hits | ITGB5 | intronic |
| 3 | 124853372 | 0.88 | 1 | rs3772853 | G | A | 0.39 | 0.38 | 0.70 | 0.40 | | | 6 tissues | | | Gfi1,Gfi1b | | 1 hit | 6 hits | ITGB5 | intronic |
| 3 | 124853715 | 0.88 | 1 | rs13074444 | C | A | 0.49 | 0.38 | 0.70 | 0.40 | | | 6 tissues | | P300 | 21 altered motifs | | | 7 hits | ITGB5 | intronic |
| 3 | 124854979 | 0.94 | 1 | rs10934693 | G | C | 0.63 | 0.39 | 0.70 | 0.40 | | | | | | 6 altered motifs | | | 7 hits | ITGB5 | intronic |
| 3 | 124856796 | 0.91 | 1 | rs1499961 | C | G | 0.52 | 0.38 | 0.70 | 0.40 | | | 6 tissues | | | AP-1 | | | 7 hits | ITGB5 | intronic |
| 3 | 124858443 | 0.91 | 1 | rs11928547 | T | C | 0.52 | 0.38 | 0.70 | 0.40 | | | 13 tissues | 9 tissues | | Ik-1,STAT | | | 6 hits | ITGB5 | intronic |
| 3 | 124858708 | 0.9 | 0.99 | rs11928651 | T | C | 0.42 | 0.39 | 0.70 | 0.40 | | FAT | 13 tissues | 4 tissues | | | | 1 hit | 6 hits | ITGB5 | intronic |
| 3 | 124858979 | 1 | 1 | rs6778643 | C | T | 0.67 | 0.41 | 0.70 | 0.40 | | | 17 tissues | 4 tissues | | 4 altered motifs | | 1 hit | 7 hits | ITGB5 | intronic |
| 3 | 124869070 | 0.85 | 0.96 | rs9813129 | G | T | 0.52 | 0.38 | 0.70 | 0.40 | | | 4 tissues | PLCNT | | SREBP,TFE | | 1 hit | 7 hits | ITGB5 | intronic |
| 3 | 124870437 | 0.85 | 0.96 | rs3772865 | C | A | 0.42 | 0.38 | 0.70 | 0.40 | | | MUS, LNG, SKIN | | | 6 altered motifs | | | 6 hits | ITGB5 | intronic |
| 3 | 124871487 | 0.84 | 0.95 | rs6802275 | C | G | 0.52 | 0.39 | 0.69 | 0.40 | | | | | | BHLHE40,HEY1,Sin3Ak-20 | | | 6 hits | ITGB5 | intronic |
| 3 | 124879179 | 0.88 | 0.96 | rs9875516 | A | T | 0.63 | 0.39 | 0.70 | 0.41 | | 14 tissues | 20 tissues | 26 tissues | 16 bound proteins | RREB-1,SP1,SRF | | | 6 hits | ITGB5 | intronic |
| 3 | 124879679 | 0.88 | 0.96 | rs10804564 | A | G | 0.63 | 0.39 | 0.70 | 0.41 | | | | ESDR,SKIN,LNG | | | | 1 hit | 7 hits | ITGB5 | intronic |
| 3 | 124883125 | 0.85 | 0.96 | rs9968182 | C | T | 0.52 | 0.38 | 0.70 | 0.41 | | ESC, BRN | 17 tissues | ESDR,SKIN,BRN | | AP-4,Mtf1,Pou5f1 | | | 7 hits | ITGB5 | intronic |
| 3 | 124883737 | 0.82 | 0.96 | rs13323528 | G | A | 0.49 | 0.38 | 0.70 | 0.40 | | ESC, BRN | 18 tissues | ESDR,ESDR | | | | | 6 hits | ITGB5 | intronic |
| 3 | 124884035 | 0.82 | 0.96 | rs4422355 | G | C | 0.49 | 0.38 | 0.70 | 0.40 | | 6 tissues | 18 tissues | ESDR,ESDR,IPSC | | Nkx3,PU.1 | | 1 hit | 7 hits | ITGB5 | intronic |
| 3 | 124884171 | 0.82 | 0.96 | rs2047571 | T | C | 0.49 | 0.38 | 0.70 | 0.40 | | 4 tissues | 18 tissues | ESDR,SKIN | CFOS | 5 altered motifs | | | 6 hits | ITGB5 | intronic |
| 3 | 124884280 | 0.82 | 0.96 | rs2047573 | G | C | 0.49 | 0.38 | 0.69 | 0.40 | | 4 tissues | 18 tissues | 15 tissues | CFOS | Gfi1,Hdx,p300 | | | 6 hits | ITGB5 | intronic |
| 3 | 124884440 | 0.82 | 0.96 | rs13079779 | C | T | 0.49 | 0.38 | 0.70 | 0.40 | | ESC, FAT, MUS | 19 tissues | SKIN,SKIN | CFOS | 4 altered motifs | | | 6 hits | ITGB5 | intronic |
| 3 | 124884471 | 0.81 | 0.96 | rs13079799 | C | A | 0.48 | 0.37 | 0.69 | 0.40 | | ESC, FAT, MUS | 19 tissues | | | 5 altered motifs | | | 6 hits | ITGB5 | intronic |

| | | | | | | | | | | | | | | | | | | | | | |
|---|---|---|---|---|---|---|---|---|---|---|---|---|---|---|---|---|---|---|---|---|---|
| 3 | 124885016 | 0.82 | 0.96 | rs35900531 | A | T | 0.38 | 0.38 | 0.70 | 0.40 | | 7 tissues | 18 tissues | 4 tissues | | 4 altered motifs | | | 5 hits | ITGB5 | intronic |
| 3 | 124886011 | 0.84 | 0.93 | rs1007856 | A | G | 0.56 | 0.40 | 0.70 | 0.40 | | | | 9 tissues | | 4 altered motifs | | 1 hit | 7 hits | ITGB5 | intronic |
| 3 | 124894424 | 0.83 | 0.94 | rs6784520 | C | A | 0.50 | 0.39 | 0.68 | 0.40 | | | | | | 7 altered motifs | | | 5 hits | 6.6kb 5' of ITGB5 | |
| 3 | 124895470 | 0.83 | 0.94 | rs9869147 | C | T | 0.51 | 0.39 | 0.69 | 0.39 | | | BRST, GI | | | 8 altered motifs | | | 6 hits | 7.6kb 5' of ITGB5 | |

Query SNP: rs17405958 and variants with $r^2$ >= 0.8

| chr | pos (hg38) | LD ($r^2$) | LD (D') | variant | Ref | Alt | AFR freq | AMR freq | ASN freq | EUR freq | SiPhy cons | Promoter histone marks | Enhancer histone marks | DNAse | Proteins bound | Motifs changed | NHGRI/ EBI GWAS hits | GRASP QTL hits | Selected eQTL hits | GENCODE genes | dbSNP func annot |
|---|---|---|---|---|---|---|---|---|---|---|---|---|---|---|---|---|---|---|---|---|---|
| 13 | 91502239 | 0.81 | 0.97 | rs149268070 | C | G | 0.04 | 0.10 | 0.00 | 0.13 | | | | | | 5 altered motifs | | | | GPC5 | intronic |
| 13 | 91503190 | 0.82 | 0.94 | rs17320630 | A | C | 0.02 | 0.12 | 0.00 | 0.17 | | | | IPSC,BLD | | | | | | GPC5 | intronic |
| 13 | 91510298 | 0.86 | 0.97 | rs79234127 | C | T | 0.02 | 0.10 | 0.00 | 0.13 | | | | | CEBPB | Pax-8 | | | | GPC5 | intronic |
| 13 | 91518416 | 0.97 | 1 | rs112395478 | T | C | 0.02 | 0.11 | 0.00 | 0.15 | | | | | | 5 altered motifs | | | | GPC5 | intronic |
| 13 | 91519199 | 0.86 | 0.97 | rs17405729 | A | G | 0.02 | 0.10 | 0.00 | 0.13 | | | | BLD | | 4 altered motifs | | | | GPC5 | intronic |
| 13 | 91519580 | 0.86 | 0.97 | rs78989063 | G | A | 0.02 | 0.10 | 0.00 | 0.13 | | | | BLD | | CEBPG,Nanog | | | | GPC5 | intronic |
| 13 | 91525044 | 1 | 1 | rs17405958 | A | G | 0.02 | 0.11 | 0.00 | 0.15 | | | | | | Foxp3 | | | | GPC5 | intronic |
| 13 | 91529841 | 0.86 | 0.97 | rs79827779 | A | G | 0.03 | 0.10 | 0.00 | 0.13 | | | | | | 7 altered motifs | | | | GPC5 | intronic |
| 13 | 91532238 | 0.97 | 1 | rs17321293 | T | C | 0.02 | 0.11 | 0.00 | 0.15 | | | | | | BCL,PU.1 | | | | GPC5 | intronic |
| 13 | 91546378 | 0.87 | 0.97 | rs7325234 | G | T | 0.04 | 0.12 | 0.00 | 0.16 | | | | | | 6 altered motifs | | | | GPC5 | intronic |

Query SNP: rs12494729 and variants with $r^2$ >= 0.8

| chr | pos (hg38) | LD ($r^2$) | LD (D') | variant | Ref | Alt | AFR freq | AMR freq | ASN freq | EUR freq | SiPhy cons | Promoter histone marks | Enhancer histone marks | DNAse | Proteins bound | Motifs changed | NHGRI/ EBI GWAS hits | GRASP QTL hits | Selected eQTL hits | GENCODE genes | dbSNP func annot |
|---|---|---|---|---|---|---|---|---|---|---|---|---|---|---|---|---|---|---|---|---|---|
| 3 | 65255065 | 0.88 | 0.95 | rs3921883 | A | G | 0.09 | 0.31 | 0.17 | 0.19 | | | | | | HEY1,Pou4f3,TATA | | | | 22kb 5' of AC104331.1 | |
| 3 | 65259755 | 0.94 | 0.99 | rs60874658 | A | C | 0.09 | 0.30 | 0.17 | 0.19 | | | LNG | | | Myf,ZBRK1 | | | | 27kb 5' of AC104331.1 | |
| 3 | 65262605 | 0.92 | 0.99 | rs4688551 | C | A | 0.09 | 0.30 | 0.17 | 0.19 | | | | 6 tissues | | 4 altered motifs | | | | 30kb 5' of AC104331.1 | |
| 3 | 65263082 | 0.88 | 0.99 | rs4688554 | C | T | 0.08 | 0.29 | 0.17 | 0.18 | | | | | | 7 altered motifs | | | | 30kb 5' of AC104331.1 | |
| 3 | 65263850 | 0.88 | 0.99 | rs73118040 | G | A | 0.08 | 0.29 | 0.16 | 0.17 | | | | | | 4 altered motifs | | | | 31kb 5' of AC104331.1 | |
| 3 | 65263887 | 0.88 | 0.99 | rs73118041 | G | T | 0.09 | 0.29 | 0.17 | 0.17 | | | | | | GR,ZNF263 | | | | 31kb 5' of AC104331.1 | |
| 3 | 65265899 | 1 | 1 | rs12494729 | A | G | 0.09 | 0.31 | 0.17 | 0.19 | | MUS | 8 tissues | SKIN,MUS | | | | 1 hit | | 33kb 5' of AC104331.1 | |
| 3 | 65273260 | 0.84 | 0.95 | rs12638637 | A | G | 0.08 | 0.30 | 0.16 | 0.17 | | | | | | | | | | 40kb 5' of AC104331.1 | |
| 3 | 65276180 | 0.84 | 0.95 | rs4688555 | A | G | 0.08 | 0.30 | 0.16 | 0.16 | | | BRN | | | 6 altered motifs | | | | 43kb 5' of AC104331.1 | |

Query SNP: rs7105948 and variants with $r^2$ >= 0.8

| chr | pos (hg38) | LD ($r^2$) | LD (D') | variant | Ref | Alt | AFR freq | AMR freq | ASN freq | EUR freq | SiPhy cons | Promoter histone marks | Enhancer histone marks | DNAse | Proteins bound | Motifs changed | NHGRI/EBI GWAS hits | GRASP QTL hits | Selected eQTL hits | GENCODE genes | dbSNP func annot |
|---|---|---|---|---|---|---|---|---|---|---|---|---|---|---|---|---|---|---|---|---|---|
| 11 | 114144937 | 1 | 1 | rs7105948 | G | T | 0.46 | 0.25 | 0.37 | 0.17 | | | 12 tissues | BRN | | En-1,Irf | | | | ZBTB16 | intronic |

Query SNP: rs10940058 and variants with $r^2$ >= 0.8

| chr | pos (hg38) | LD ($r^2$) | LD (D') | variant | Ref | Alt | AFR freq | AMR freq | ASN freq | EUR freq | SiPhy cons | Promoter histone marks | Enhancer histone marks | DNAse | Proteins bound | Motifs changed | NHGRI/ EBI GWAS hits | GRASP QTL hits | Selected eQTL hits | GENCODE genes | dbSNP func annot |
|---|---|---|---|---|---|---|---|---|---|---|---|---|---|---|---|---|---|---|---|---|---|
| 5 | 66298307 | 1 | 1 | rs10940058 | T | C | 0.17 | 0.41 | 0.16 | 0.51 | | | | | | CDP,Irf,STAT | | | | RP11-305P14.1 | intronic |

Query SNP: rs2726050 and variants with $r^2$ >= 0.8

| chr | pos (hg38) | LD ($r^2$) | LD (D') | variant | Ref | Alt | AFR freq | AMR freq | ASN freq | EUR freq | SiPhy cons | Promoter histone marks | Enhancer histone marks | DNAse | Proteins bound | Motifs changed | NHGRI/ EBI GWAS hits | GRASP QTL hits | Selected eQTL hits | GENCODE genes | dbSNP func annot |
|---|---|---|---|---|---|---|---|---|---|---|---|---|---|---|---|---|---|---|---|---|---|
| 7 | 36156680 | 0.85 | 1 | rs2726056 | A | G | 0.14 | 0.41 | 0.12 | 0.38 | | 15 tissues | 12 tissues | | EBF1 | 4 altered motifs | | | | EEPD1 | intronic |
| 7 | 36158938 | 0.94 | 1 | rs2700908 | T | C | 0.28 | 0.39 | 0.11 | 0.38 | | | 8 tissues | PANC,MUS | | RXRA | | | | EEPD1 | intronic |
| 7 | 36159518 | 1 | 1 | rs2700909 | T | C | 0.14 | 0.37 | 0.11 | 0.38 | | | 8 tissues | KID | | AP-3,Nanog | | | 1 hit | EEPD1 | intronic |
| 7 | 36160134 | 1 | 1 | rs2726050 | C | T | 0.14 | 0.37 | 0.11 | 0.38 | | FAT, MUS | 12 tissues | | | 6 altered motifs | | | 1 hit | EEPD1 | intronic |
| 7 | 36162104 | 1 | 1 | rs6462657 | G | A | 0.15 | 0.37 | 0.12 | 0.38 | | | 8 tissues | 10 tissues | STAT3,FOXA1 | 5 altered motifs | | | 1 hit | EEPD1 | intronic |
| 7 | 36162897 | 1 | 1 | rs2726099 | A | G | 0.15 | 0.37 | 0.12 | 0.38 | | | 5 tissues | IPSC | | 5 altered motifs | | | 1 hit | EEPD1 | intronic |
| 7 | 36163588 | 1 | 1 | rs1833178 | C | G | 0.15 | 0.37 | 0.11 | 0.38 | | | BLD, MUS, LIV | GI,GI,GI | | GR | | | 1 hit | EEPD1 | intronic |
| 7 | 36165513 | 0.92 | 1 | rs28488583 | G | A | 0.16 | 0.35 | 0.11 | 0.38 | | | 7 tissues | | | ERalpha-a,HNF4,SP1 | | | 1 hit | EEPD1 | intronic |
| 7 | 36167381 | 1 | 1 | rs2700912 | C | A | 0.15 | 0.37 | 0.11 | 0.38 | | | 4 tissues | | | Irf,RFX5 | | | 1 hit | EEPD1 | intronic |
| 7 | 36167482 | 1 | 1 | rs2700913 | C | G | 0.15 | 0.37 | 0.11 | 0.38 | | | 5 tissues | | | 5 altered motifs | | | 1 hit | EEPD1 | intronic |
| 7 | 36168072 | 1 | 1 | rs2700914 | C | T | 0.15 | 0.37 | 0.11 | 0.38 | | | 4 tissues | | | Mrg | | | 1 hit | EEPD1 | intronic |
| 7 | 36168975 | 0.95 | 0.99 | rs2700916 | C | A | 0.16 | 0.38 | 0.11 | 0.38 | | | | | | 7 altered motifs | | | 1 hit | EEPD1 | intronic |
| 7 | 36170380 | 0.96 | 0.99 | rs2700917 | G | A | 0.15 | 0.38 | 0.11 | 0.38 | | | | | | | | | 1 hit | EEPD1 | intronic |
| 7 | 36171137 | 0.96 | 0.99 | rs2700918 | G | A | 0.15 | 0.38 | 0.11 | 0.38 | | | | THYM | | 4 altered motifs | | | 1 hit | EEPD1 | intronic |

Query SNP: rs928641 and variants with $r^2$ >= 0.8

| chr | pos (hg38) | LD ($r^2$) | LD (D') | variant | Ref | Alt | AFR freq | AMR freq | ASN freq | EUR freq | SiPhy cons | Promoter histone marks | Enhancer histone marks | DNAse | Proteins bound | Motifs changed | NHGRI/ EBI GWAS hits | GRASP QTL hits | Selected eQTL hits | GENCODE genes | dbSNP func annot |
|---|---|---|---|---|---|---|---|---|---|---|---|---|---|---|---|---|---|---|---|---|---|
| 9 | 117165231 | 1 | 1 | rs928641 | C | T | 0.54 | 0.63 | 0.70 | 0.54 | | | ESDR | | | Cdx,TCF4 | | | | ASTN2 | intronic |
| 9 | 117165517 | 0.94 | -0.98 | rs10759905 | G | C | 0.47 | 0.36 | 0.34 | 0.46 | | | | ESDR | | ATF3,Spz1 | | | | ASTN2 | intronic |
| 9 | 117165841 | 0.95 | -1 | rs4292766 | G | A | 0.43 | 0.36 | 0.31 | 0.46 | | | | | | 4 altered motifs | | | | ASTN2 | intronic |
| 9 | 117167299 | 0.94 | 0.98 | rs7039227 | C | T | 0.49 | 0.63 | 0.65 | 0.54 | | | | | | 15 altered motifs | | | | ASTN2 | intronic |
| 9 | 117168675 | 0.9 | 0.98 | rs933082 | T | C | 0.49 | 0.65 | 0.66 | 0.55 | | | IPSC | | | Hoxb3 | | | | ASTN2 | intronic |
| 9 | 117171191 | 0.94 | 0.98 | rs7853534 | C | T | 0.49 | 0.63 | 0.66 | 0.54 | | | | | | 6 altered motifs | | | | ASTN2 | intronic |
| 9 | 117176596 | 0.94 | -0.98 | rs10739484 | T | C | 0.42 | 0.36 | 0.30 | 0.46 | | | | | | Elf3,Foxp1,TAL1 | | | | ASTN2 | intronic |
| 9 | 117180370 | 0.97 | 1 | rs977937 | C | T | 0.49 | 0.62 | 0.64 | 0.54 | | | ESDR | IPSC | | DMRT1,GR | | | | ASTN2 | intronic |

| | | | | | | | | | | | | | | |
|---|---|---|---|---|---|---|---|---|---|---|---|---|---|---|
| 9 | 117184981 | 0.81 | 0.94 | rs4838266 | C | G | 0.40 | 0.61 | 0.65 | 0.52 | BRN | ZEB1 | ASTN2 | intronic |
| 9 | 117185182 | 0.91 | -0.99 | rs6478287 | T | G | 0.44 | 0.35 | 0.32 | 0.45 | BRN | PLZF | ASTN2 | intronic |

**Query SNP enhancer summary:**

NOTE: Background for GWAS SNPs is still based on Haploreg v4.0 and will be updated to reflect v4.1

| Cell | Observed | Expected (all SNPs) | Expected (GWAS SNPs) | Binomial p (all SNPs) | Binomial p (GWAS SNPs) |
|---|---|---|---|---|---|
